# Adiposity and cancer: systematic review and meta-analysis

**DOI:** 10.1101/2024.02.16.24302944

**Authors:** Eleanor L. Watts, Amparo Gonzalez-Feliciano, Marc J. Gunter, Nilanjan Chatterjee, Steven C. Moore

## Abstract

Obesity is a growing global health challenge and a major cancer risk factor. We conducted a systematic review and meta-analysis of prospective cohorts quantifying associations between body mass index (BMI) and 25 common cancers, searching PubMed, EMBASE and Scopus through April 23rd, 2025. Across 226 articles comprising 1.5 million cancer data points, BMI was positively associated with 19 cancer types and inversely associated with three. We identified positive associations for leukaemia, non-Hodgkin lymphoma, bladder, and glioma—novel findings which were not reported by major consensus reports. Regional differences were observed, including stronger associations for postmenopausal breast and ovarian cancers in East Asia, and weaker associations for gallbladder cancer. BMI and waist circumference showed similar associations with cancer. We also reviewed Mendelian randomisation studies and emerging imaging-based evidence; genetic data largely supported causality, while imaging-data were limited. Our findings underscore the broad and growing impact of obesity on cancer risk.

## 1. Introduction

Global economic development, urbanisation, and changes in diet and physical activity have driven a worldwide rise in body mass index (BMI). Once limited to high-income countries, obesity (BMI ≥30 kg/m^2^) is now a global challenge^1^, with low- and middle-income countries accounting for 75% of the world’s adult obese population—including China, which ranks second only to the United States in the number of obese individuals^2^. Among the health consequences of rising BMI is an increased risk of cancer^3^, and may contribute to the rapidly growing cancer burden in low- and middle-income countries^4^.

Between 2016 and 2018, landmark reviews by the World Cancer Research Fund (WCRF)^3^, the International Agency for Research on Cancer (IARC)^5^, and others^6^ established that elevated BMI is associated with increased risk of at least 12 types of cancer. While these reviews marked a major advance in our understanding of obesity and cancer, key questions remain. Chief among them whether, and to what extent, these associations generalise across global regions. To our knowledge, only Renehan et al. (2008) systematically addressed this^7^, but their analysis was limited by sparse data outside Europe and North America. If obesity– cancer associations differ by region, this could substantially affect global estimates of cancer burden attributable to obesity and deepen—or complicate—our understanding of the underlying mechanisms. Other key uncertainties include whether BMI is associated with less common or smoking-related cancers, and whether risk estimates would differ using alternative measures of adiposity, such as waist circumference.

Since these reviews, major new epidemiological data has emerged from large, population-wide administrative healthcare databases, such as those from South Korea, and next-generation cohorts with deep phenotypic and genotypic data, including UK Biobank. These resources offer new opportunities to characterise obesity–cancer associations across diverse global populations and to explore new avenues of inquiry, such as the use of Mendelian randomisation (MR) to strengthen causal inference^8^. Yet despite these advances, BMI–cancer associations have not been comprehensively re-assessed through a systematic review and meta-analysis. A precise, up-to-date understanding of obesity’s impact on cancer risk is especially critical in the current era of glucagon-like peptide-1 receptor agonists (GLP-1RA) and other incretin-based therapies, as healthcare systems weigh their high costs against potential public health benefits^9,10^.

We aimed to examine the current global epidemiological landscape of BMI and cancer by conducting a systematic review and meta-analysis of the prospective associations with 25 common cancers, comparing our findings with previous WCRF reviews and with results from our meta-analyses of MR studies. We also explore the regional distribution of epidemiological studies to date and whether associations vary regionally. Finally, we compare associations with BMI with those for waist circumference and summarise emerging evidence from imaging-derived adiposity measures.

## 2. Results

### 2.1. Characteristics of included studies

We identified 4,993 potentially eligible articles through our database search and 69 additional potentially eligible articles from reference review. We excluded 3,309 duplicate articles, 1,089 ineligible articles based on title and abstract screening, and 279 after full-text review. An additional 159 articles were excluded as they were superseded by a more recent publication from the same population with larger case numbers. In total, 226 articles met our inclusion criteria, yielding 557 BMI-cancer associations (some studies reported on multiple cancer types) (**Supplementary** Figure 1).

The included cohorts originated from 24 countries and six major geographic regions (**Figure 1**). For three countries—Australia, Lithuania, and Israel—we included prospective cohorts with <50,000 participants to improve representation from their respective geographic regions. There were no studies originating from Africa, South or Central America, Eastern Europe, South or Central Asia, the Caribbean, or the Pacific Islands (excluding Hawaii) (**Figure 1**). The countries included represent only 33% of the global population^11^, and include none of the global regions where obesity is most prevalent, particularly the Pacific Islands, the Arabian Peninsula, and neighbouring Gulf states^2^.

**Figure 1:**
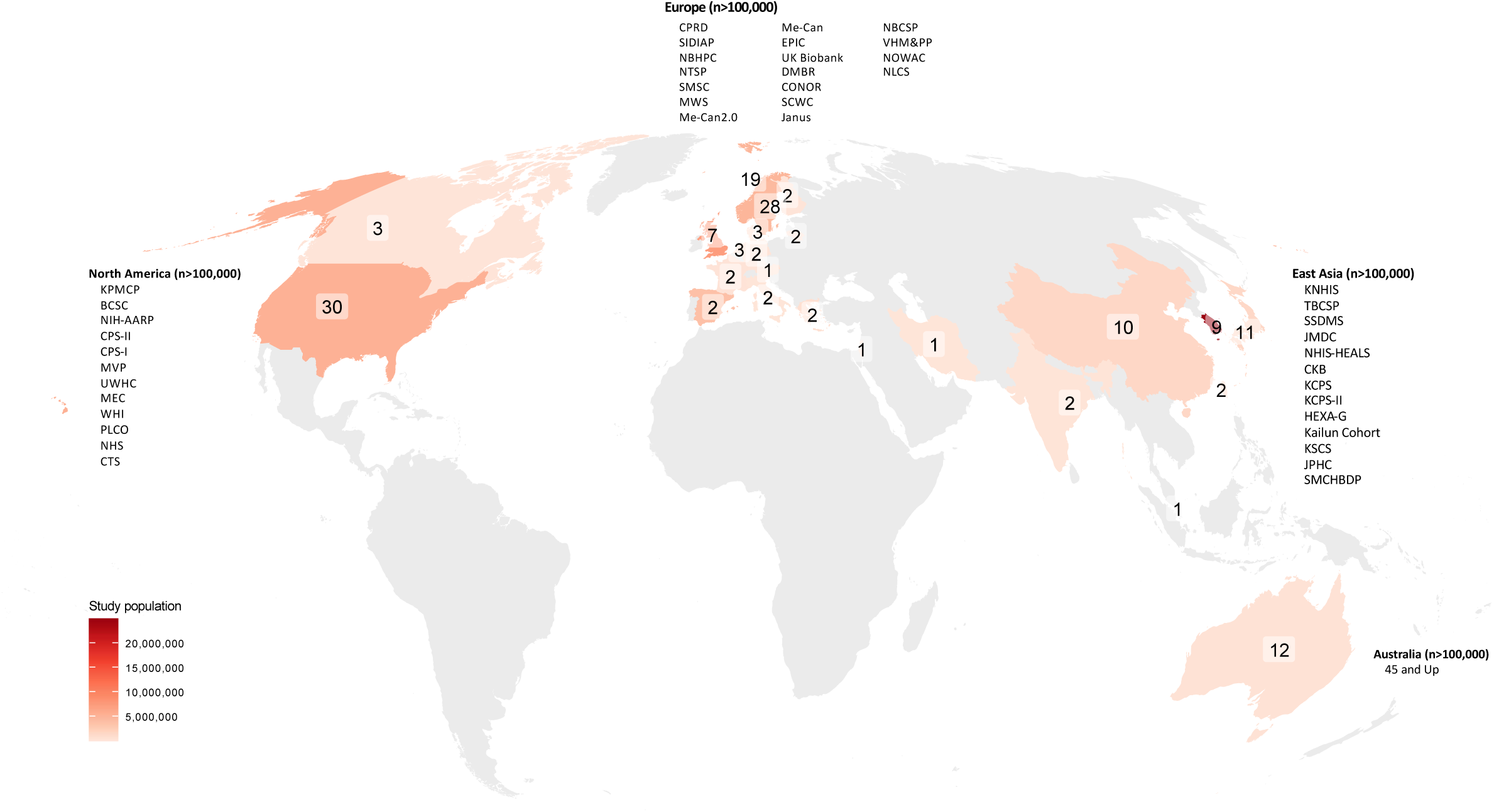
**World map of included cohorts** Numbers show the count of prospective cohorts with cancer data in each country included in this meta-analysis. Study population reflects the total N of participants across cohorts within each country. For cohorts reported in multiple publications with varying analytic sample sizes, the largest sample size was used. Text labels highlight prospective studies with over 100,000 participants in each region. Multi-country cohorts and pooling projects (e.g., EPIC, Adventist Health Study 2) have been disaggregated by country based on participant numbers at each study centre. Where possible, pooling projects consisting of several smaller cohorts were further separated into their individual constituent cohorts. The cohorts in India report on cancer mortality only. Abbreviations: 45 and Up=The Sax Institute’s 45 and Up Study, BCSC=Breast Cancer Surveillance Consortium, CKB=China Kadoorie Biobank, CPRD=UK Clinical Practice Research Datalink, CPS=Cancer Prevention Study, CTS=California Teachers Study, DMBR=Danish Medical Birth Registry, EPIC=European Prospective Study into Cancer and Nutrition, HEXA=Health Examinees, JPHC=Japan Public Health Center, KCPS=Korean Cancer Prevention Study, KNHIS=National Health Insurance Service of Korea, KPNC=Kaiser Permanente Northern California, KSCS=Kangbuk Samsung Cohort Study, MEC=Multiethnic Cohort Study, MIYAGI=Miyagi Cohort Study, MVP=Million Veteran Program, MWS=Million Women Study, NBCSP=Norwegian Breast Cancer Screening Program, NBHPC=Norwegian BMI/Height Prospective Cohort 1963-2001, NHIS-HEALS=National Health Insurance Service-National Health Screening Cohort, NHS=Nurses’ Health Study, NIH-AARP=National Institutes of Health-AARP, NLCS=Netherlands Cohort Study, NOWAC=Norwegian Women and Cancer Study, NTSP=Norwegian Tuberculosis Screening Program, PLCO=Prostate, Lung, Colorectal and Ovarian Cancer Screening Trial, SCWC=Swedish Construction Workers Cohort, SIDIAP=Information System for Research in Primary Care, SMCHBDP=Shandong Multi-Center Healthcare Big Data Platform, SMBR=Swedish Medical Birth Register, SMCR=Swedish Military Conscription Register, SSDMS=Shanghai Standardized Diabetes Management System, TBCSP=Taiwanese Breast Cancer-Screening Program, UWHC=University of Wisconsin Hospital and Clinics Electronic Health Record Study, VHA= Veterans Health Administration, VHM&PP=Vorarlberg Health Monitoring and Prevention Programme, WHI=Women’s Health Initiative

In aggregate, this review includes 1,492,122 incident cancers across the 25 types of cancer, with the most cases for colorectal cancer (N=382,393) and the least for oesophageal squamous cell cancer (SQ, N=998; never-smokers only). This represents at least a twofold increase in the number of cases for each cancer type—and up to 27 times more for certain types—compared to the WCRF reports (**Table 1**). Participant numbers increased similarly; the colorectal cancer analysis, for example, had 44.4 million participants as compared with 4.8 million in the prior WCRF review. This exponential increase in case numbers and participants since 2014 (**Figure 2**), largely stems from large administrative healthcare database studies like that of the National Health Insurance Service of Korea.

**Figure 2:**
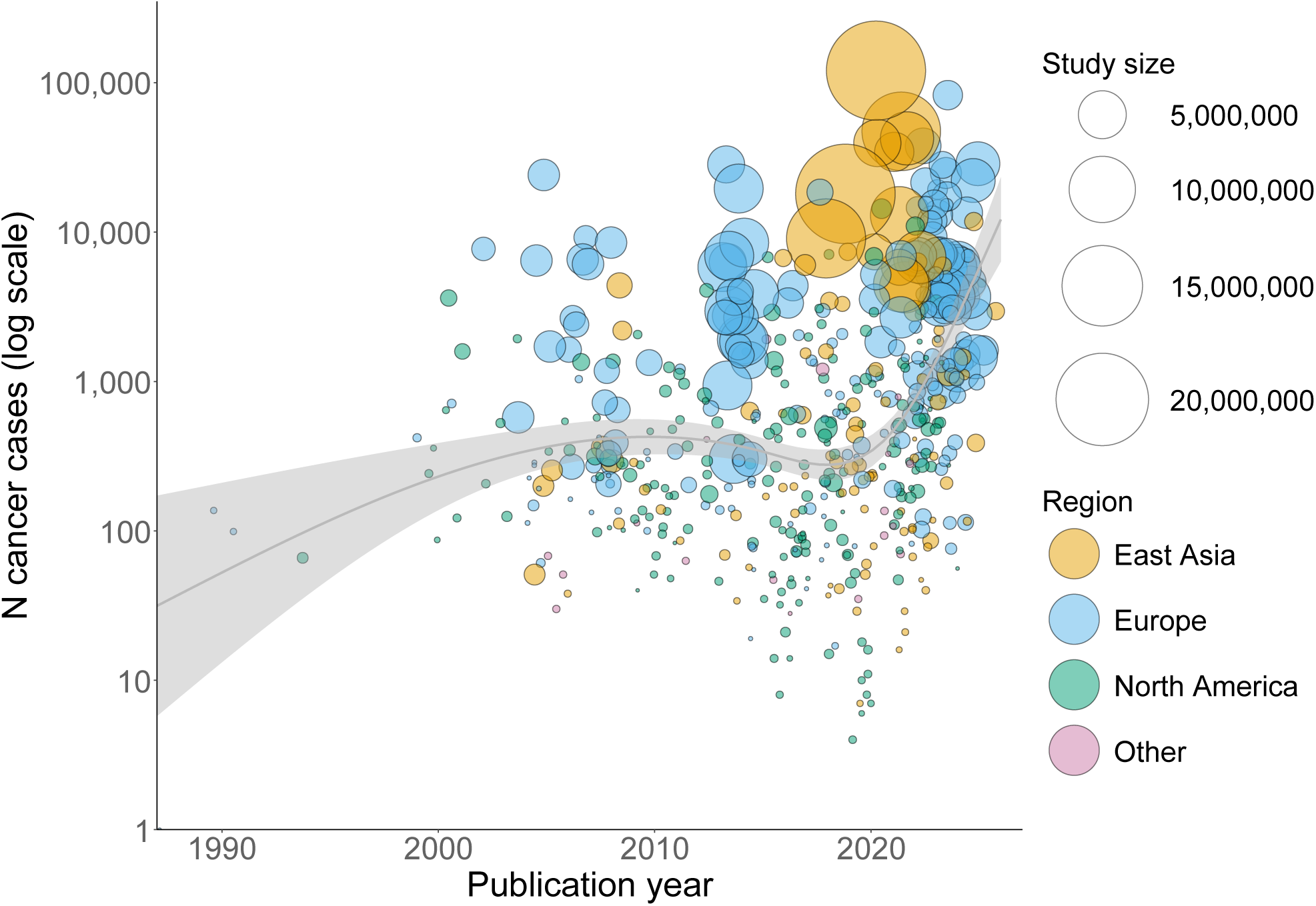
**Prospective cohort cancer studies by the number of incident cancer cases and publication year** Bubble size shows the number of participants for each BMI-cancer study. The trend line shows the average number of cancer cases by publication year (modelled by spline), and the grey band shows the 95% confidence intervals.

**Table 1:** Comparison of current meta-analysis with the WCRF review.

| Cancer type | Current meta-analysis |  |  |  |  | WCRF reviews |  |  |  |  | Meta-analysis vs. WCRF |
| --- | --- | --- | --- | --- | --- | --- | --- | --- | --- | --- | --- |
|  | N cases | Analytic cohort | N cohorts | N countries | N regions | N cases | N cohorts | N countries | N regions | Last study year | Case N ratio |
| Endometrial | 60,478 | 17,079,995 | 56 | 14 | 3 | 18,941 | 25 | 15 | 3 | 2011 | 3:1 |
| Oesophageal (adenocarcinoma) | 6,837 | 10,844,390 | 46 | 12 | 3 | 1,988 | 9 | 7 | 2 | 2013 | 3:1 |
| Kidney | 61,233 | 43,000,793 | 52 | 15 | 3 | 16,076 | 23 | 15 | 3 | 2010 | 4:1 |
| Gallbladder | 11,028 | 18,137,607 | 69 | 20 | 4 | 5,745 | 8 | 13 | 3 | 2013 | 2:1 |
| Gastric (cardia) | 6,153 | 9,295,453 | 49 | 16 | 5 | 2,050 | 7 | 6 | 3 | 2008 | 3:1 |
| Liver | 74,353 | 31,721,583 | 54 | 13 | 3 | 11,664 | 8 | 13 | 2 | 2012 | 6:1 |
| Breast (postmenopausal) | 206,378 | 13,854,162 | 75 | 17 | 4 | 78,847 | 42 | 18 | 4 | 2015 | 3:1 |
| Thyroid | 31,249 | 20,049,521 | 70 | 18 | 4 | - | - | - | - | - | ∞ |
| Meningioma | 8,307 | 8,204,514 | 33 | 12 | 3 | - | - | - | - | - | ∞ |
| Colorectal | 382,393 | 44,378,077 | 90 | 18 | 5 | 46,359 | 29 | 13 | 4 | 2014 | 8:1 |
| Multiple myeloma | 32,630 | 19,857,717 | 45 | 12 | 3 | - | - | - | - | - | ∞ |
| Leukaemia | 42,771 | 20,227,141 | 41 | 14 | 4 | - | - | - | - | - | ∞ |
| Pancreas | 64,271 | 26,791,580 | 68 | 17 | 5 | 9,401 | 23 | 14 | 3 | 2010 | 7:1 |
| Head and neck (NS) | 11,872 | 11,417,335 | 48 | 15 | 4 | 796 | 20 | 13 | 4 | 2015 | 15:1 |
| Ovarian | 46,535 | 14,466,459 | 49 | 15 | 4 | 15,773 | 22 | 16 | 4 | 2012 | 3:1 |
| NHL | 69,209 | 20,596,219 | 49 | 14 | 3 | - | - | - | - | - | ∞ |
| Cervix | 22,979 | 13,589,200 | 37 | 13 | 3 | 5,035 | 9 | 7 | 3 | 2014 | 5:1 |
| Prostate (aggressive)* | 33,746 | 4,794,643 | 60 | 17 | 5 | 9,838 | 23 | 14 | 4 | 2012 | 3:1 |
| Bladder (NS) | 22,221 | 21,015,602 | 34 | 11 | 3 | - | - | - | - | - | ∞ |
| Glioma | 16,846 | 15,383,918 | 37 | 13 | 3 | - | - | - | - | - | ∞ |
| Melanoma | 77,022 | 17,850,030 | 43 | 12 | 4 | 17,689 | 14 | 9 | 4 | 2016 | 4:1 |
| Gastric (non-cardia) | 66,866 | 15,355,406 | 54 | 16 | 5 | 2,462 | 7 | 7 | 4 | 2008 | 27:1 |
| Lung (NS) | 12,873 | 7,609,867 | 53 | 15 | 3 | 4,712 | 12 | 7 | 4 | 2014 | 3:1 |
| Breast (premenopausal) | 123,338 | 14,828,849 | 77 | 17 | 4 | 16,371 | 27 | 14 | 3 | 2015 | 8:1 |
| Oesophageal (SQ, NS) | 988 | 2,024,960 | 38 | 10 | 2 | 114 | 2 | 9 | 1 | 2009 | 9:1 |

\*Aggressive prostate cancer was defined as any of stage 3-4 on the American Joint Committee on Cancer 1992 classification, advanced cancer, advanced or metastatic cancer; metastatic cancer; stage C or D on the Whitmore/Jewett scale; fatal cancer (prostate cancer-specific mortality); high stage or grade; Gleason grade $\geq 7$ ).
Abbreviations: NHL=non-Hodgkin lymphoma, NS=never-smokers, SQ=squamous cell carcinoma, RR=risk ratio, WCRF=World Cancer Research Fund

Cancer cases predominantly originated from Europe (59.0%), followed by East Asia (30.1%), and North America (10.6%). The remainder were from Australia, India, Iran, and Israel (**Figure 3A**). Notably, geographic distribution varied by cancer type; for example, no large studies on oesophageal SQ in never-smokers were available from North America, and for most cancers, no data were available outside Europe, East Asia and North America (**Figure 3B**).

**Figure 3:**
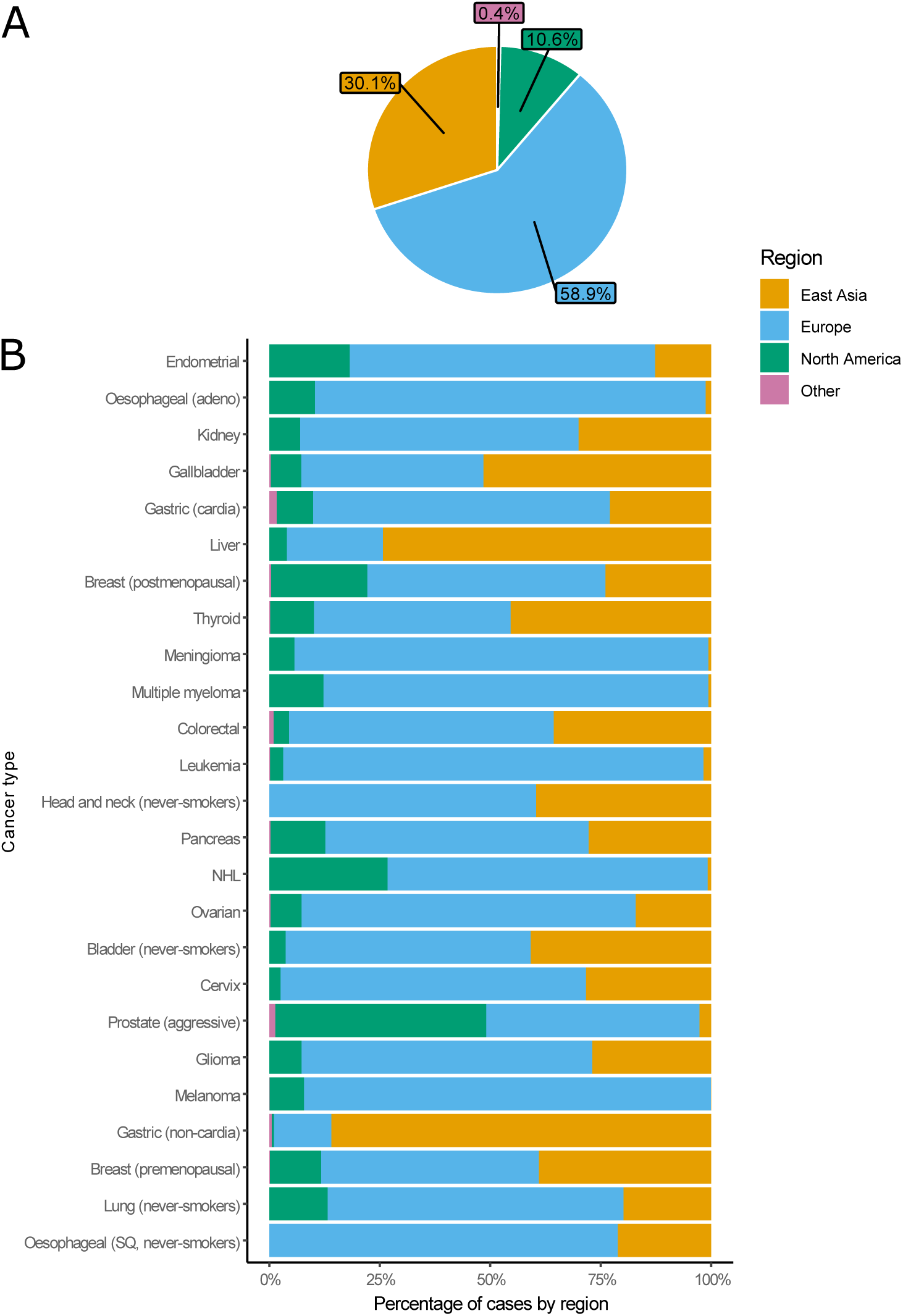
Percentage of cancer cases originating from each geographic region. **A: Sum of the 25 types of incident cancers** **B: Individual cancers** Estimates from pooled studies spanning multiple regions for which country-specific case numbers were not available were excluded from this figure (head and neck). Numbers may not sum due to rounding. Abbreviations: NHL=non-Hodgkin lymphoma, SQ=squamous cell carcinoma

Details for each study including model adjustments, N cases, N analytic population, risk estimates, follow-up duration and originally reported units are available from **Supplementary Data**.

### 2.2. BMI-cancer associations

Higher BMI was associated with varying magnitudes of increased cancer risk, with nearly a twentyfold difference in risks across cancer types. At the upper end, a 5 kg/m^2^ increase in BMI was significantly associated with a 58% increased risk of endometrial cancer (95% confidence interval [CI] 1.51-1.67) and a 47% higher risk of oesophageal adenocarcinoma (1.39-1.56); at the other end, the same increment in BMI was associated with a 3% increased risk of glioma (1.01-1.05). BMI was also positively associated with risk of kidney (1.30, 1.26-1.33), gallbladder (1.27, 1.23-1.32), gastric (cardia) (1.23, 1.11-1.36), liver (1.20, 1.12-1.28), breast (postmenopausal) (1.15, 1.12-1.17), thyroid (1.12, 1.07-1.17), meningioma (1.11, 1.05-1.16), multiple myeloma (1.10, 1.08-1.11), colorectal (1.10, 1.08-1.12), leukaemia (1.09, 1.05-1.13), pancreatic (1.07, 1.05-1.09), ovarian (1.05, 1.02-1.08), non-Hodgkin lymphoma (NHL:1.05, 1.01-1.08), prostate (aggressive) (1.04, 1.00-1.09), and glioma (1.03, 1.01-1.05). BMI was inversely associated with breast (premenopausal) (0.92, 0.89-0.95) (**Figure 4**). For cancers where smoking is the dominant risk factor, we observed increased risks for head and neck (1.07, 1.02-1.13) and bladder cancer (1.04, 1.01-1.07), and decreased risks for oesophageal SQ (0.72, 0.60-0.86) and lung cancer (0.92, 0.86-0.99) for never-smokers. The estimated false discovery rate across the 25 cancers was 5%, indicating a low likelihood that the findings occurred by chance.

**Figure 4:**
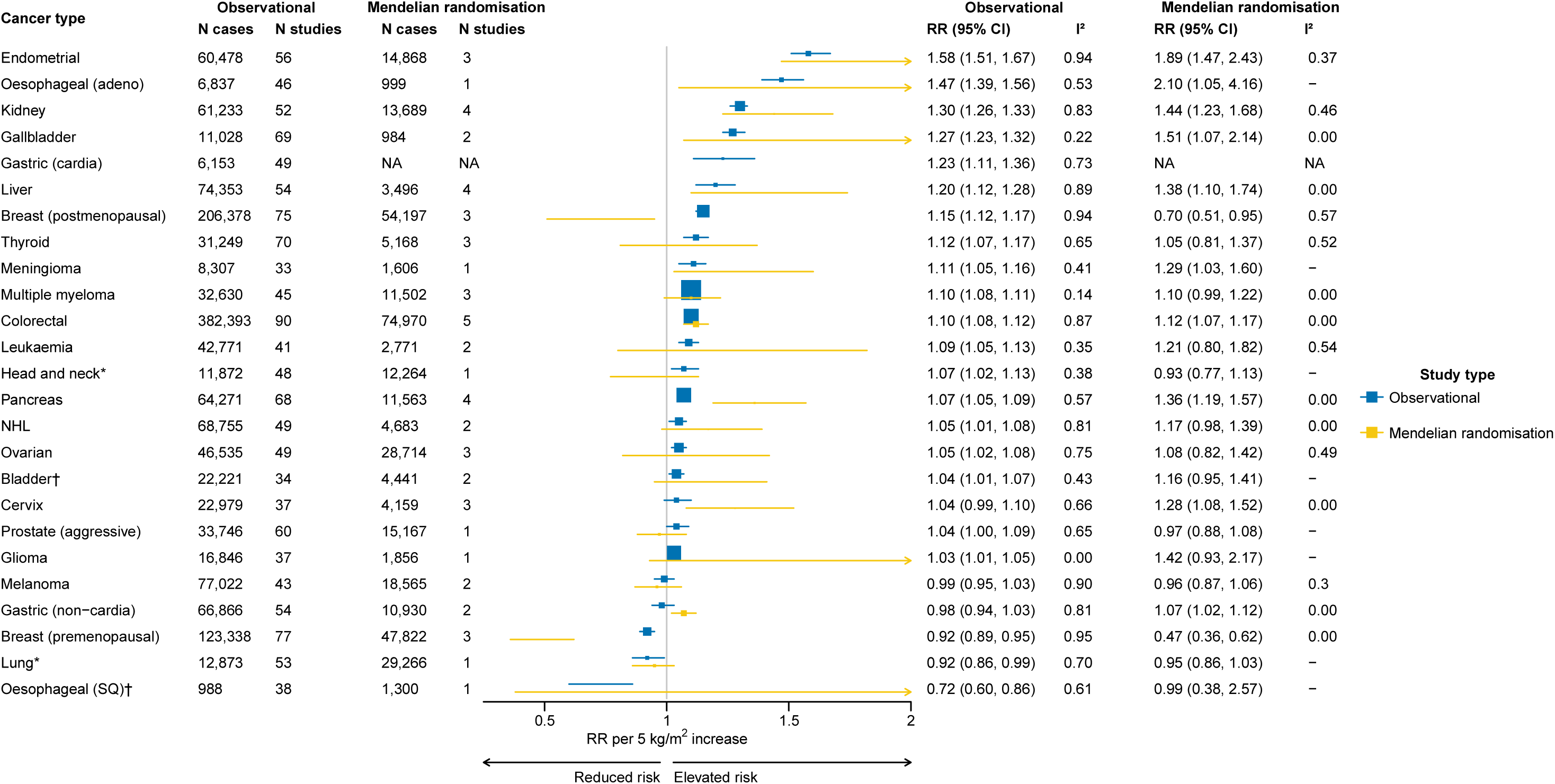
**Associations between BMI with cancer risk, RR per 5 kg/m² increase in BMI** RRs are represented by squares (with their 95% CIs as lines). Observational risk estimates were calculated using random effects meta-analysis. Heterogeneity between studies is quantified using I^2^, an I^2^ value close to 100% indicates substantial heterogeneity but can be affected by the number of studies and the precision of individual study estimates. Further details of model adjustments, follow-up time, analytic population for each study are available from **Supplementary Data**. *Observational estimates are based on never-smokers; MR analyses adjust for genetically predicted smoking. †Observational estimates are based on never-smokers; smoking-adjusted MR analyses were not available, so univariable MR results are shown. Abbreviations: Adeno=adenocarcinoma, BMI=body mass index, CI=confidence interval, MR=Mendelian randomisation, NHL=non-Hodgkin lymphoma, RR=risk ratio, SQ=squamous cell carcinoma

For each cancer type, RRs were broadly consistent in direction across studies, though with some quantitative heterogeneity (i.e., I^2^> 0%; **Supplementary** Figures 2-26). Heterogeneity was greatest for endometrial, breast (pre- and post-menopausal), and melanoma (I^2^ ≥90%). Despite the high I^2^ for endometrial and breast cancers, associations were directionally consistent and statistically significant in most large studies, with broadly similar magnitudes; for endometrial cancer, the interquartile range (IQR) of study-specific RRs was 1.47–1.75. In contrast, for melanoma, some studies reported positive associations and others null (IQR=1.01-1.25).

For most cancer types, there was little evidence of publication bias. Among cancers with >10 studies^12^, Egger’s intercept was statistically significant (p<0.05) for colorectal, premenopausal breast, and endometrial cancers, but the differences between the trim-and-fill RRs and the uncorrected RRs were small (maximum 7% change) (**Supplementary** Figure 27). Overall, study quality was high (mean total score=1.87 out of 2, SD=0.34). Most studies adequately adjusted for confounders (mean confounding control score=0.97, SD=0.17) and had follow-up durations longer than 5 years (mean follow-up score=0.90, SD=0.30). These findings suggest that reverse causation or confounding is unlikely to explain the observed associations. Scores by cancer type are provided in **Supplementary Table 1** and for each study in the **Supplementary Data**.

In sensitivity analyses, restricting to cohorts that adjusted for smoking or were at low risk of bias had minimal effect on associations, with changes in RRs of <5% across all cancer types. Excluding administrative healthcare database studies led to a loss of many cases but yielded similar RRs for most cancers. A notable exception was oesophageal SQ, where the association became more strongly inverse (RR per 5 kg/m²=0.61, 95% CI 0.44–0.85) compared with the primary analysis (RR=0.72). In contrast, for aggressive prostate cancer, the association became more positive (1.16, 0.85–1.58) compared with the primary analysis (1.04; **Supplementary Table 2**).

Risk estimates were broadly consistent with those from the previous WCRF review, though more precise (median absolute difference in RRs=0.03; **Table 2**). The largest difference observed was for oesophageal SQ cell carcinoma (Δ=0.13), which may reflect our study’s substantially greater precision in estimating the risk estimate for this cancer, which is rare in never-smokers.

**Table 2:** Comparison of current meta-analysis with the WCRF review.

| Cancer | Current analysis,<br>RR per 5 kg/m<sup>2</sup> (95% CI) | WCRF,<br>RR per 5 kg/m<sup>2</sup> (95% CI) | Change in RR |
| --- | --- | --- | --- |
| Endometrial | 1.58 (1.51, 1.67) | 1.50 (1.42, 1.59) | 0.08 |
| Oesophageal (adeno) | 1.47 (1.39, 1.56) | 1.48 (1.35, 1.62) | -0.01 |
| Kidney | 1.30 (1.26, 1.33) | 1.30 (1.25, 1.35) | 0.00 |
| Gallbladder | 1.27 (1.23, 1.32) | 1.25 (1.15, 1.37) | 0.02 |
| Gastric (cardia) | 1.23 (1.11, 1.36) | 1.23 (1.07, 1.40) | 0.00 |
| Liver | 1.20 (1.12, 1.28) | 1.30 (1.16, 1.46) | -0.10 |
| Breast (postmenopausal) | 1.15 (1.12, 1.17) | 1.12 (1.09, 1.15) | 0.03 |
| Thyroid | 1.12 (1.07, 1.17) | - | - |
| Meningioma | 1.11 (1.05, 1.16) | - | - |
| Colorectal | 1.10 (1.08, 1.12) | 1.05 (1.03, 1.07) | 0.05 |
| Multiple myeloma | 1.10 (1.08, 1.11) | - | - |
| Leukaemia | 1.09 (1.05, 1.13) | - | - |
| Pancreas | 1.07 (1.05, 1.09) | 1.10 (1.07, 1.14) | -0.03 |
| Head and neck (NS) | 1.07 (1.02, 1.13) | 1.15 (1.06, 1.24) | -0.08 |
| Ovarian | 1.05 (1.02, 1.08) | 1.06 (1.02, 1.11) | -0.01 |
| NHL | 1.04 (1.01, 1.07) | - | - |
| Cervix | 1.04 (0.99, 1.10) | 1.02 (0.97, 1.07) | 0.02 |
| Prostate (aggressive)* | 1.04 (1.00, 1.09) | 1.08 (1.04, 1.12) | -0.04 |
| Bladder (NS) | 1.04 (1.01, 1.07) | - | - |
| Glioma | 1.03 (1.01, 1.05) | - | - |
| Melanoma | 0.99 (0.95, 1.03) | 1.02 (0.98-1.05) | -0.03 |
| Gastric (non-cardia) | 0.98 (0.94, 1.03) | 0.93 (0.85, 1.02) | 0.05 |
| Lung (NS) | 0.92 (0.86, 0.99) | 0.93 (0.84, 1.03) | -0.01 |
| Breast (premenopausal) | 0.92 (0.89, 0.95) | 0.93 (0.90, 0.97) | -0.01 |
| Oesophageal (SQ, NS) | 0.72 (0.60, 0.86) | 0.59 (0.44, 0.79) | 0.13 |

\*Aggressive prostate cancer was defined as any of stage 3-4 on the American Joint Committee on Cancer 1992 classification, advanced cancer, advanced or metastatic cancer; metastatic cancer; stage C or D on the Whitmore/Jewett scale; fatal cancer (prostate cancer-specific mortality); high stage or grade; Gleason grade $\geq 7$ ).
Abbreviations: NHL=non-Hodgkin lymphoma, NS=never-smokers, SQ=squamous cell carcinoma, RR=risk ratio, WCRF=World Cancer Research Fund.

In the MR analysis (see **Supplementary** Figure 28 for included studies), results were generally consistent with those from the observational analysis (**Figure 4**). Of the 22 observational associations, nine showed significant and directionally consistent associations in MR analyses: endometrial (OR per 5 kg/m^2^=1.89, 95% CI 1.47-2.43), oesophageal adenocarcinoma (2.10, 1.05-4.16), kidney (1.44, 1.23-1.68), gallbladder (1.51, 1.07-2.14), liver (1.38, 1.10-1.74), meningioma (1.29, 1.03-1.60), colorectal (1.12, 1.07-1.17), and pancreatic cancers (1.36, 1.19-1.57), as well as an inverse association for pre-menopausal breast cancer (0.47, 0.36-0.62). For thyroid cancer, multiple myeloma, leukaemia, ovarian cancer, non-Hodgkin lymphoma, and glioma, MR estimates were generally slightly stronger than observational estimates; however, reduced precision meant these results were not statistically significant. The larger effect sizes are consistent with MR potentially reflecting the impact of lifelong exposure to elevated BMI. Notably, in contrast to observational findings, MR analyses reported an inverse association with postmenopausal breast cancer (0.70, 0.51–0.95) and a positive association with gastric non-cardia cancer (1.07, 1.02–1.12).

For smoking-related cancers, multivariable MR estimates accounting for genetically predicted smoking were only available for head and neck^13^ and lung cancers^14^. For head and neck cancer, the univariable MR estimate per 5 kg/m^2^ increase in BMI was 1.29 (0.69-2.40), which attenuated to 0.93 (0.77-1.13) after adjustment for smoking. Similarly, the univariable estimate for lung cancer was 1.38 (1.17-1.63), but this was attenuated to 0.95 (0.86-1.03) when smoking was accounted for. For bladder cancer and oesophageal SQ only univariable MR estimates were available. The association for bladder cancer (1.16, 0.95-1.41) was directionally consistent with the observational association, while the estimate for oesophageal SQ was null, though imprecise (0.99, 0.38–2.57). Further study details and risk estimates are available in **Supplementary Data**.

Overall, we had high certainty for eleven cancer types: endometrial, oesophageal (adeno), kidney, gallbladder, gastric (cardia), liver, breast (postmenopausal), thyroid, meningioma, colorectal, multiple myeloma. There was moderate certainty for leukaemia and pancreatic and ovarian cancers, and NHL, and for the inverse associations with breast (pre-menopausal), lung, oesophageal (SQ). There was low certainty for head and neck cancer, prostate cancer (aggressive), bladder cancer, and glioma (**Supplementary Table 3**).

### 2.3. Evaluation of regional heterogeneity

BMI-cancer associations were generally consistent across regions, particularly between Europe and North America (median absolute difference in RRs=0.02). While larger differences were observed between East Asia and Western regions, for example, the difference between East Asia and Europe was 0.12. Comparatively few cases were available from other regions, precluding analysis of obesity-cancer associations for much of the world, underscoring the need for far more data to capture the full breadth of BMI-cancer relationships.

For several cancer types, we observed evidence of heterogeneity, with particularly striking regional differences for postmenopausal breast cancer. The RR per 5 kg/m^2^ increment was 1.12 in North America (95% CI 1.09-1.16) and 1.11 in Europe (1.08-1.15) but substantially higher—1.27—in East Asia (1.22-1.32), a more than doubling of the excess risk. For ovarian cancer, the effect size of the BMI association was also higher in East Asia (1.16, 1.12-1.21) compared to North America (1.02, 0.98-1.06) and Europe (1.04, 1.01-1.07). The associations for thyroid cancer were also stronger in East Asia (1.23, 1.14-1.33) in comparison with North America (1.08, 1.01-1.14) and Europe (1.09, 1.02-1.15). For gallbladder cancer, the pattern was reversed, with a weaker association in East Asia (1.15, 1.09-1.22) compared to Europe (1.33, 1.28-1.38) and North America (1.26, 1.17-1.37). For gastric cardia associations were null in East Asia (0.96, 0.79-1.16), and positive in Europe (1.30, 1.22-1.39) and North America (1.39, 1.20-1.62). For bladder cancer, the BMI association was weaker in Europe (1.02, 1.00-1.04) than in East Asian and North America (RR = 1.10 in both regions) (**Figure 5**).

**Figure 5:**
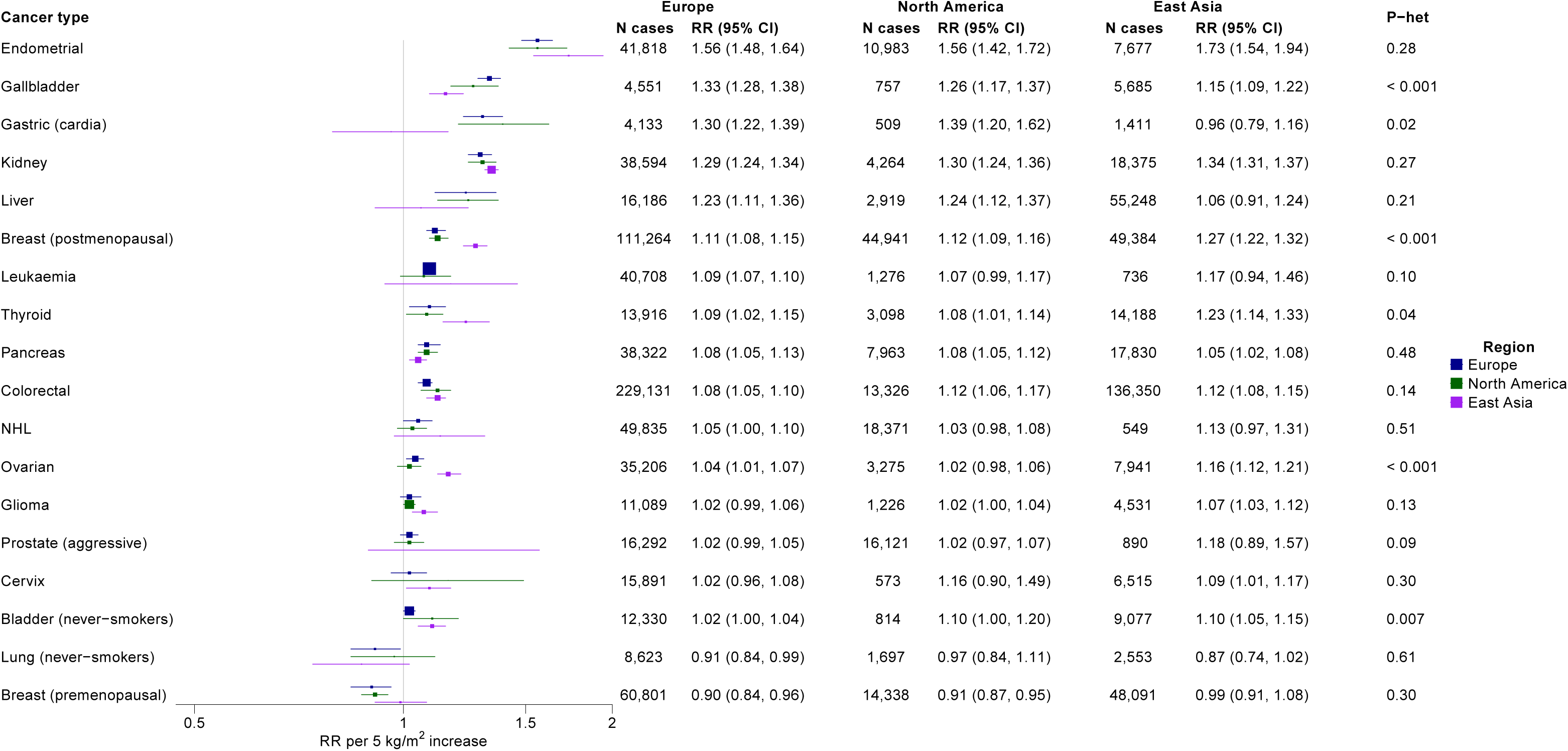
**Associations between BMI and cancer risk by region** Figure restricted to cancer types with >500 cases per region and estimates available for Europe, North America, and East Asia. Risk estimates were calculated using random effects meta-analysis. P-heterogeneity was estimated using the Q-statistic and includes the regions Australia, South Asia, and West Asia (not shown here due to the low number of cancers). Full results for each site and region are available from **Supplementary** Figures 29-53. Abbreviations: CI=confidence interval, NHL=non-Hodgkin lymphoma, RR=risk ratio, SD=standard deviation, SQ=squamous cell carcinoma

Associations by cancer, region and study are available from **Supplementary** Figures 29-53.

### 2.4. Waist circumference vs. BMI and cancer risk

We identified eight studies including up to 2.1 million participants and 122,000 cases across five countries (China, Spain, Sweden, UK, and the USA)^15–22^. Overall, differences in risk estimates between adiposity measures were small (median absolute difference in RRs = 0.02). However, for several smoking-related cancers, we observed evidence of heterogeneity when analyses were not restricted to never-smokers, particularly for lung cancer (RR per 1 SD increase: BMI 0.87 [0.81-0.94] vs. waist circumference 0.96 [0.89-1.05]), bladder cancer (1.00 [0.92-1.08] vs. 0.93 [0.89-0.97], and oesophageal SQ (0.73 [0.59-0.90] vs. 0.84 [0.78-0.90]). For lung cancer, this heterogeneity was attenuated when analyses were restricted to never-smokers. However, similar data were not available for other smoking-related cancers. Statistically significant heterogeneity between adiposity measures was also observed for oesophageal adenocarcinoma (BMI 1.31 [1.08-1.60] vs. waist circumference 1.36 [1.14-1.62]) and colorectal cancer (1.11 [1.04-1.18] vs. 1.15 [1.07-1.24]) (**Figure 6**). Risk associations for each study and type of cancer are available from **Supplementary** Figures 54-79.

**Figure 6:**
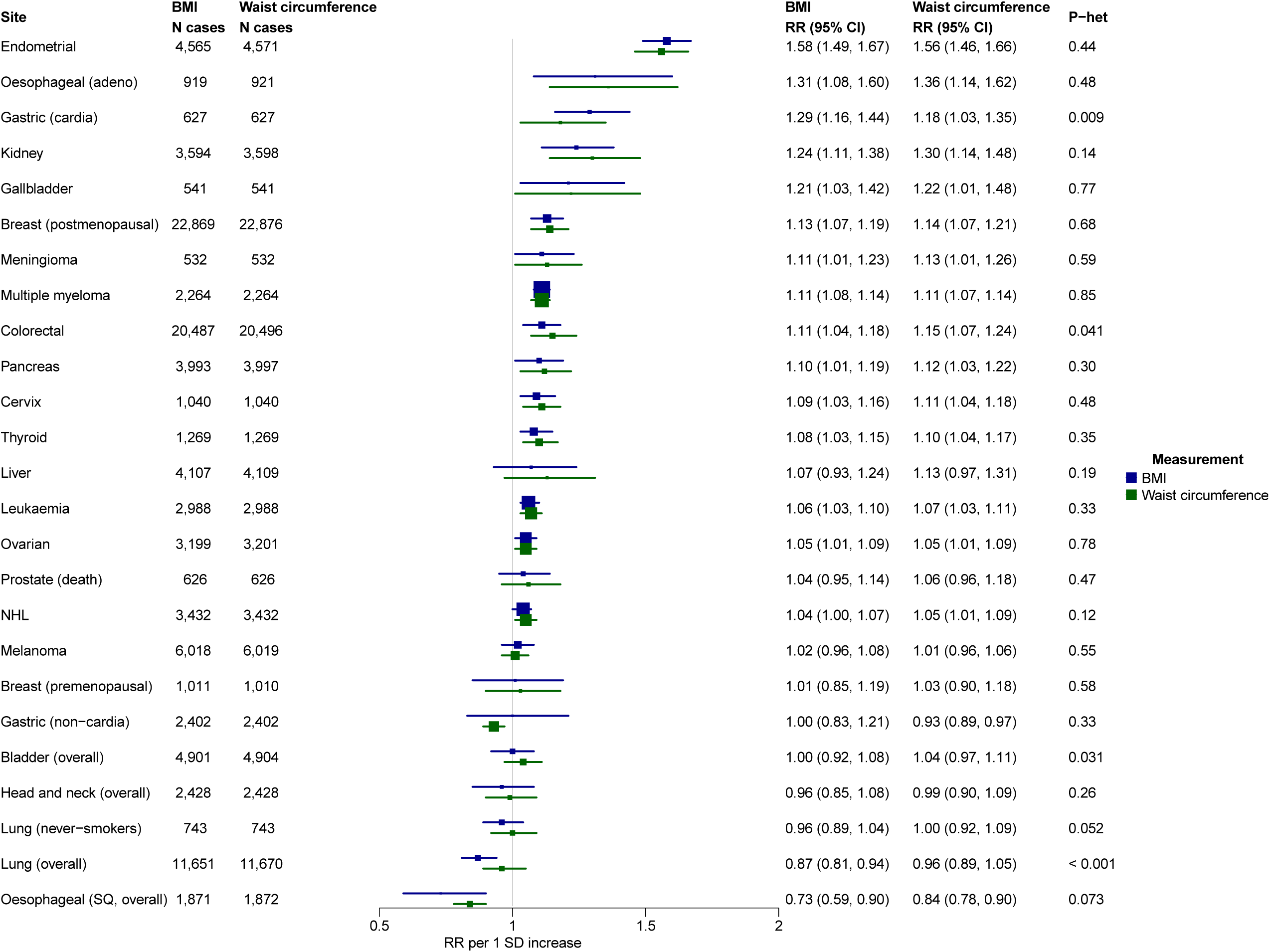
**Associations of BMI, waist circumference and cancer risk, per 1 SD increase** Associations estimated using random effects meta-analysis. Heterogeneity in the associations with each cancer site by adiposity measure was estimated using the Wald statistic. Abbreviations: CI=confidence interval, NHL=non-Hodgkin lymphoma, RR=risk ratio, SD=standard deviation

### 2.5. Adiposity imaging studies

Our database search identified 107 non-duplicate articles, of which 85 were excluded during title and abstract screening and 18 after full-text review, leaving 4 articles (**Supplementary** Figure 80).

Two studies from the Women’s Health Initiative (WHI) evaluated the association between adiposity measured by dual-energy X-ray absorptiometry (DXA) and cancer risk. In one study, which included 639 incident breast cancer cases, higher whole-body fat percentage (RR per 1 SD=1.16, 95% CI 1.07–1.26) and trunk fat mass (1.21, 1.12–1.31) were associated with increased risk, similar to the effect for BMI (1.19, 1.10–1.28)^23^. In contrast, a separate study of 169 colorectal cancer cases found no significant associations between BMI, DXA-derived measures and risk^24^. In the AGES-Reykjavik study, which used computed tomography (CT) imaging, one analysis examined prostate cancer risk, though statistical power was limited (43 aggressive prostate cancers)^25^. A more recent UK Biobank study reported that higher intrapancreatic fat, measured by magnetic resonance imaging (MRI), was associated with increased pancreatic cancer risk (HR per 1 quintile increase=1.55, 1.26-1.92). This association persisted after further adjustment for BMI and central obesity (1.41, 1.10-1.82)^26^. However, the study did not directly compare the magnitude of these associations with those for BMI. Risk estimates are available from **Supplementary Table 4**.

## 3. Discussion

This comprehensive systematic review and meta-analysis of 226 prospective cohort studies, comprising 1.5 million cancers across 25 common types, found that higher BMI was associated with increased risk of 19 cancer types and inversely associated with three. Our findings substantially expand upon prior reviews and offer three key insights: 1) obesity may be associated with a broader range of cancers than previously recognised; 2) while some associations are consistent across regions, risk estimates for breast and ovarian cancers may be higher in East Asia, whereas those for gallbladder cancer may be weaker, and three other cancers show borderline regional heterogeneity; and 3) BMI and waist circumference show similar associations with cancer risk.

### 3.1. BMI-cancer associations

Hundreds of studies have linked obesity to increased cancer risk. In this analysis, we synthesise these data into a comprehensive meta-analysis providing precise risk estimates, including for less common cancer types, and employing multiple sensitivity and genetic analyses to strengthen causal inference. Our primary finding, that BMI was positively associated with the risk of 19 cancer types, suggests that adiposity may be a risk factor for most cancers, emphasising the importance of obesity prevention as a public health priority. By leveraging up to 27 times more cases than earlier reports, we identified novel associations for leukaemia, NHL, bladder cancer and glioma. For other cancers, our findings largely reaffirm associations previously reported in WCRF reviews. Notably, despite the substantially larger sample size, risk estimates for these established associations remained consistent. This stability suggests that BMI–cancer associations may have reached a point of convergence for many cancer types. Further data collection may yield limited additional insight unless it focuses on specific deficiencies, particularly the lack of prospective cohorts from most nations of the world.

Presenting all the evidence together using uniform methods allows for systematic comparison of associations across cancer types. The nearly twentyfold variation in effect sizes across cancer types suggests that obesity may affect cancer risk through multiple mechanisms, to which different tissues and organs exhibit varying levels of susceptibility. For oesophageal adenocarcinoma, the hypothesised mechanisms are mechanical, with excess weight compressing the stomach and/or relaxing the gastroesophageal sphincter, thereby increasing the risk of gastro-oesophageal reflux^27^. For cancers of the kidney, gallbladder, liver, and pancreatic cancers, the proposed mechanisms are metabolic, i.e. hypertension, gallstones, non-alcoholic fatty liver disease, and diabetes, respectively. For female reproductive cancers such as breast (postmenopausal) and endometrial cancer, obesity increases concentrations of circulating oestrogens through aromatisation of testosterone and androstenedione in adipose tissue^28–32^. For other cancers, associations may be mediated by systemic inflammation, hyperinsulinemia or other indirect downstream effects. Differences in the magnitude of BMI-cancer associations likely also arise in part from other strong, type-specific risk factors—for example, the dominant role of *H. pylori* in non-cardia gastric cancer^33^, and the high prevalence of chronic hepatitis in some settings, which could attenuate BMI-liver cancer associations. In the coming years, the application of -omics technologies, such as proteomics, to epidemiologic research is expected to enhance scientific understanding of the biological mechanisms linked adiposity and cancer^34^.

Our analysis also provided important insights into the relationship between BMI and smoking-related cancers. Since the 1970s, epidemiologic studies have reported that a higher BMI is associated with a lower risk of several smoking-related cancers^35–37^, particularly lung cancer^38,39^, but the interpretation of these associations remains contentious. Smoking is the dominant risk factor for these malignancies—current smokers have a 10-fold higher risk of lung cancer compared to never-smokers^40,41^, for example—and is generally associated with a lower BMI^4243^. Whether statistical adjustment can fully account for the magnitude and complexity of this confounding remains uncertain^44^. Consequently, studies reach divergent conclusions; some suggest the inverse associations are due to causal effects of adiposity^45–48^, while others attribute them to residual confounding by smoking^44,49–54^.

To minimise residual confounding by smoking, we restricted analyses of lung, bladder, oesophageal SQ, and head and neck cancers to never-smokers. In this group, higher BMI was associated with *increased* risks of bladder and head and neck cancers, a lower risk of lung cancer, and a markedly lower risk of oesophageal SQ. These findings suggest that residual confounding may explain earlier inverse associations for bladder and head and neck cancers, may substantially contribute to those observed for lung cancer, but is unlikely to account for the strong inverse relationship observed for oesophageal SQ. For lung cancer, an important consideration is that histological subtypes and somatic mutation profiles differ between never-smokers and current smokers^55–58^, suggesting that it may represent a biologically distinct disease. Consequently, associations between BMI and lung cancer risk may also vary by smoking status^59,60^. In that light, our MR analyses provide some reassurance as they found similar results even in analyses not restricted to never-smokers. A previous analysis using overlapping data likewise found no association between genetically predicted BMI and overall lung cancer risk, although it did report heterogeneity across histological subtypes^14^.

One complex and long-standing finding is the inverse association between BMI and the risk of premenopausal breast cancer. Although consistent evidence for this relationship began to emerge in the 1990s^61^, it has continued to perplex researchers. However, recent evidence suggests that the distinction between pre- and post-menopausal breast cancer is not the salient distinction *per se.* Instead, the timing of BMI measurement appears to be critical, with higher BMI in childhood associated with a reduced risk of *both* pre- and postmenopausal breast cancer^62^. This may help explain why genetically-predicted BMI was associated with lower risk of *both* pre- and postmenopausal breast cancer in our study: genetically predicted BMI more closely reflects childhood body size through to adolescence than adult BMI, whereas observational associations are likely influenced by later life weight gain^63–65^.

### 3.2. Evaluation of regional heterogeneity

It is important to recognise that scientific disciplines differ in how they approach heterogeneity—both in its assigned importance and in how they interpret its implications. In epidemiology, examining heterogeneity is considered fundamental. It can reveal potential cofactors underlying aetiologic relationships^66^, and strengthen internal validity by demonstrating that findings are robust across varying confounding structures^8^. Regional analyses are particularly informative as they permit exploration of aetiologic relationships across a full spectrum of human lifestyles and environments.

In our analysis, we observed substantial regional heterogeneity in BMI-cancer associations for female reproductive cancers (breast and ovarian cancer), and gallbladder cancer. We also noted some evidence of heterogeneity for gastric cardia, thyroid, and bladder cancer, although differences were less pronounced. From a public health perspective, the large heterogeneity for breast cancer is particularly important. Current estimates of obesity-attributable breast cancer cases in East Asia are based on RRs from Western populations^67^ and our results suggest that these estimates may understate the true number of cases due to obesity by a factor of two or more, emphasising the need to use region-specific estimates in future studies.

Regional variation in reproductive cancer associations may stem from differences in background risk factors. For instance, postmenopausal hormone therapy is not widely used in East Asia^68–70^ and BMI is more strongly associated with breast cancer risk among non-hormone-users^71^. Additionally, postmenopausal oestrogen levels appear to be lower in East Asian women^72–74^, suggesting that high BMI could contribute more to a women’s overall oestrogenic exposure in this population. For ovarian cancer, regional differences may also reflect variation in hormone therapy and oral contraceptive use, although interpretation is more challenging given the disease’s marked heterogeneity in histological subtypes^75–77^. For gallbladder cancer, the weaker association with BMI in East Asia may reflect differences in gallbladder comorbidities. In Western populations, obesity is linked to cholesterol-rich gallstones, while in East Asia, brown pigment stones—more common—are associated with chronic infections and biliary stasis^78,79^. Regional differences may also reflect variation in the prevalence of strong risk factors (e.g., hepatitis B/C virus or *H. pylori*), or in the distribution of tumour subtypes, which sometimes have distinct etiological pathways. Alternatively, differences in relative risks across regions could arise from variation in surveillance practices or confounding structures. For instance, if confounding is less pronounced in East Asian populations, stronger associations might more closely approximate the true effect of obesity on cancer risk, suggesting that earlier studies underestimated these relationships. In addition, regional variation in the pattern of weight gain, particularly differences between early-life and later-life BMI and their respective associations with risk, may further contribute to heterogeneity.

Another potential explanation for the observed variation in associations among East Asians is differences in body fat distribution. Studies suggest that, for a given BMI, individuals of East Asian ancestry have higher levels of body fat and larger waist circumference compared to those of European ancestry^80,81^. However, this is unlikely to account for the pattern of heterogeneity observed, as we found both stronger *and* weaker associations in East Asians depending on the cancer type, rather than a consistent trend. It has also been noted that average BMI is lower in East Asian populations compared to Western countries^82^, which could yield heterogeneous findings if the BMI-cancer associations were non-linear (e.g. U- or J-shaped). However, unlike for mortality, the relationships between BMI and cancer are typically approximately linear^20,83–85^, therefore relative risk increases per unit of BMI would still be expected to remain consistent across populations.

### 3.3. Waist circumference vs. BMI and cancer risk

BMI is often critiqued for its inability to differentiate fat mass from lean muscle, and for not accounting for adiposity types, particularly visceral adipose tissue, an especially metabolically harmful fat depot stored in the abdominal cavity^86,87^. As a result, waist circumference has been proposed as a more informative alternative metric for investigating associations with cancer risk. However, findings across studies have been mixed; while some have reported stronger associations for waist circumference^21,38,88–90^, others have found little difference between the two measures^16,20,53,91,92^. In our analysis, we similarly observed minimal differences in associations with cancer risk, suggesting that substituting BMI for waist circumference, or *vice versa*, would yield broadly comparable conclusions. This likely reflects the high correlation between BMI and waist circumference at the population level (r ≈ 0.9), as well as evidence that both measures are more strongly associated with subcutaneous adipose tissue (r = 0.88 for BMI and 0.85 for waist circumference) than with visceral adipose tissue (0.78 and 0.82, respectively), and are moderately correlated with lean mass (0.75 and 0.58, respectively)^93^. These complexities make it challenging to disentangle their independent effects and highlight the limited specificity of waist circumference as a proxy for visceral adipose tissue. Nonetheless, we observed some modest differences between the two measures, particularly for smoking-related cancers, which may relate to smokers generally having lower BMI but higher waist circumference, greater body fat, and reduced muscle mass^38,94^. Possibly, smoking may be less of a confounder for waist circumference.

Imaging data suggests that fat depots have weaker correlations with BMI and waist circumference^91^. However, existing imaging studies are limited in statistical power. The UK Biobank’s planned collection of DXA and MRI scans for 100,000 participants^95^ may enhance the ability to assess fat depot-specific cancer risks and clarify the role of adipose tissue distribution in cancer development.

### 3.4. Strengths, limitations, and conclusions

This systematic review and meta-analysis has several key strengths. First, the large sample size enabled robust evaluation across a broad range of cancers, including several not previously assessed in WCRF reports or for which prior evidence was limited. We conducted detailed analyses of smoking-related cancers specifically among never-smokers, helping to minimise residual confounding. Most included studies were of high quality, with long follow-up and appropriate adjustment for confounders, reducing the likelihood of bias. To strengthen causal inference, we also triangulated findings from observational studies with evidence from MR^8^. The availability of new data from East Asia also allowed us to assess the geographic distribution of prospective cohorts and quantify regional differences in risk estimates. We systematically compared risk estimates for waist circumference and BMI to evaluate their relative associations with cancer risk and incorporated emerging data from imaging-based measures of adiposity. Finally, to support transparency and future updates, we have made the formatted data publicly available (**Supplementary Data**).

An important limitation to our study is that, although we comprehensively reviewed the world’s literature, many world regions were not represented, due to lack of prospective cohort cancer studies. These include Africa, South and Central America, Eastern Europe, South and Central Asia, the Caribbean, and the Pacific Islands. Additionally, aside from large administrative healthcare databases, most prospective cohorts are drawn from populations that are healthier and wealthier than the underlying population and often lack adequate representation of minority groups. These gaps raise concerns about the generalisability of our findings to individuals from lower socioeconomic backgrounds and underrepresented populations. Our analysis focused on the 25 most common cancers; however, many rarer cancers and specific subtypes are also likely associated with BMI^96^. We did not evaluate heterogeneity by sex, though existing evidence suggests that BMI increases the risk of most cancers in both sexes, with slightly stronger associations in men for colorectal cancer^17,53,96,97^. Given the stability of many risk estimates and the availability of large prospective datasets, future research should prioritise understudied populations, less common cancers and subtypes, while ensuring robust adjustment for confounding. Given the large volume of cancer–BMI literature, we used more restrictive search terms than are conventionally applied, which may have reduced sensitivity. We also limited inclusion to studies with ≥50,000 participants in East Asian, Northern/Western Europe and North America, thereby excluding some smaller cohorts.

However, given the substantial numbers available for each cancer, omission of these studies would have minimal to no effect on risk estimates.

In summary, BMI is positively associated with 19 common cancers and inversely associated with three. Notably, we identified positive associations for leukaemia, NHL, bladder, and glioma cancers, as well as inverse associations for lung and oesophageal SQ, which were not previously reported in WCRF or IARC reviews. MR studies generally supported causal relationships in the same directions. We also observed regional differences in associations, with larger magnitudes observed for breast and ovarian cancers in East Asia and weaker associations for gallbladder cancer. However, many populations are entirely unrepresented in obesity-cancer risk research. BMI and waist circumference showed similar associations with cancer. Collectively, these findings highlight the substantial and growing impact of obesity on cancer risk and the urgent need to address this modifiable risk factor worldwide.

## 4. Methods

This systematic review adhered to the standard criteria for reporting meta-analyses as outlined in the PRISMA guidelines^98^. Institutional review board approval was not required, as this study did not involve human participant research.

### 4.1. BMI and cancer risk

Our meta-analysis includes prospective cohort studies that examined the association of BMI with incidence of the 25 most common cancers by global incidence. A systematic search was conducted using PubMed, EMBASE, and Scopus databases for articles published from the databases’ inception through to April 23^rd^, 2025. The search terms used are detailed in **Supplementary Table 5**. To verify completeness, we also searched reference lists of recent studies. Additional details on cancer selection and the search strategy are provided in the **Supplementary Methods**.

We utilised Covidence for the screening process. We first deduplicated articles from the different databases. We then screened titles and abstracts of articles for eligibility, excluded ineligible articles, reviewed the full text of the remainder, and performed final eligibility exclusions. Screening and review were conducted independently by two authors (ELW and SCM), and a third author (AGF) broke the ties.

Eligible studies met the following criteria: 1) prospective risk estimates of adult (≥18 years) BMI-cancer associations; 2) ≥50,000 participants (unless the cohort was from an underrepresented geographic region: i.e., outside North America, North/West/Central Europe, East, and Southeast Asia); 3) continuous risk estimates or ≥3 categories of BMI; 4) provided estimates for subtype-specific cancers when pertinent (i.e., aggressive prostate cancer, gastric [cardia and non-cardia], breast [pre- and post-menopausal], oesophageal [adenocarcinoma and SQ]); 5) risk estimates for never-smokers (only for head and neck, lung, oesophageal squamous, and bladder cancers); 6) models did not adjust for early-life BMI or other measure of adiposity; 7) non-duplicative population (when duplicate populations were identified, we used the risk estimate based on the greatest number of cases).

We extracted risk estimates and study information, including hazard ratios, odds ratios, RRs, and their corresponding 95% CIs for the association between BMI and cancer risk. We prioritised estimates derived from continuous modelling of BMI and those most comprehensively adjusted for confounders, provided no adjustment for other adiposity measures (e.g. waist circumference, BMI from an earlier age). Risk estimates were converted to the 5 kg/m^2^ scale to enable comparison across studies. We also extracted sample size (cases and analytic population), cohort, country, N studies, duration of follow-up, and model adjustments. Quality for each study and cancer type was assessed using a two-point grading system: i) adjustment for confounders relevant for that type of cancer, and ii) a minimum follow-up period of 5-years to minimise the risk of reverse causation (**Supplementary Table 6**).

To understand how the landscape of cancer epidemiology has evolved, we extracted data from the WCRF reports and compared the risk estimates, N countries, sample size to each cancer type in our analysis. We focused on comparing with the WCRF reports because, unlike narrative or umbrella reviews, they conduct *de novo* meta-analyses using primary data, making them more directly comparable in terms of methods and outputs. To examine geographic patterns in risk estimates, we grouped cohorts by country and region, summarising findings both overall and by cancer type. We also evaluated trends in study size over time across geographic regions using a bubble plot, with trends modelled using natural cubic splines with three degrees of freedom.

Summary risk estimates for the association between BMI and each cancer type were estimated using random effects meta-analysis, which weights studies based on within-study and between-study variability and produces a weighted average of individual study effect sizes. Between-study heterogeneity was assessed using I^2^, tau^2^ (estimated using the Paule-Mandel method^99^), and the IQR (25^th^ and 75^th^ percentile) of RRs for each cancer type. Publication bias was evaluated using Egger’s test and the trim-and-fill method^100^.

In sensitivity analyses, we examined the impact of excluding risk estimates from studies that: 1) did not adjust for smoking; 2) were based on large administrative healthcare databases; and 3) received the maximum quality assessment score (2) (**Supplementary Methods**).

We also meta-analysed studies which investigated the associations of BMI with cancer using MR approaches and compared risk estimates with the observational. Our systematic search was conducted in PubMed, EMBASE and Scopus, covering articles published from the databases’ inception through February 6, 2025. Eligible studies met the following criteria: 1) prospective risk estimates of BMI-cancer associations; 2) estimates for subtype-specific cancers (where needed); 3) risk estimate based on a linear estimate; and 4) not duplicative study population (in the outcome population only). Univariable MR risk estimates based on the primary analytic model were extracted, generally based on the inverse variance weighted method, standardised to the 5 kg/m^2^ scale for comparability across studies, and meta-analysed.

Heterogeneity between studies was assessed using I^2^. For smoking-related cancers, we extracted multivariable MR estimates adjusted for genetically predicted smoking, where available, to account for the effect of smoking on risk estimates.

### 4.2. Assessment of certainty

We applied a tailored certainty assessment adapted from GRADE^101^, emphasising domains most relevant to observational epidemiology: magnitude of effect, confounding or reverse causation, consistency, publication bias, and concordance with MR evidence. Certainty was classified as high when all criteria were met: a large effect (RR > abs[10%]), robustness to bias (associations remained consistent when restricted to studies with the highest bias score), consistency across studies (25^th^ and 75^th^ percentiles of study-specific RRs aligned directionally with the summary estimate), robustness to publication bias (trim-and-fill analyses yielded estimates consistent with the summary), and MR findings directionally consistent with the summary estimate. Moderate certainty was assigned when the effect size was >5% and all other criteria were met, while low certainty was assigned to associations that were nominally significant (p < 0.05) but did not satisfy the thresholds for high or moderate certainty.

### 4.3. Evaluation of regional effects

To investigate heterogeneity in the associations between BMI and cancer by region, we used Cochran’s Q test, focusing on the following geographic regions: Australia, East Asia, South Asia, the Middle East, Europe, and North America. Heterogeneity in the risk estimates by BMI and waist circumference were assessed using the Wald statistic.

### 4.4. Comparison of BMI and waist circumference

To systematically compare how BMI and waist circumference relate to cancer risk, we selected from our meta-analysis only studies that met the following criteria: 1) reported results for both BMI and waist circumference using the same models and populations – to eliminate random and systematic variability that could arise from using different studies for each exposure; 2) investigated ≥5 cancer types – to minimise publication bias, as broader analyses are less prone to selective reporting (e.g. reporting only cancers where one measure “outperforms” the other)^102,103^; 3) did not mutually adjust for waist circumference or *vice versa* – mutual adjustment can distort associations between body composition measures and cancer risk^104^; 4) reported results per SD, or in continuous units convertible to SD – to ensure comparable estimates on the same scale.

These criteria ensured that comparisons between BMI and waist circumference were as methodologically consistent and unbiased as possible. We also included risk estimates for smoking-related cancers without restriction to never-smokers. This is because smokers generally have greater abdominal fat relative to their BMI^38,94,105^, and that differences between BMI and waist circumference in relation to disease outcomes are often most pronounced among current smokers^38,106,107^.

### 4.5. Adiposity imaging and cancer

We also evaluated associations between imaging-based adiposity and the risk of the top 25 cancer types. Our literature search was conducted in PubMed, EMBASE, and Scopus through to February 6, 2025.

Eligible studies met the following criteria: 1) prospective risk estimates of adiposity imaging and cancer; 2) estimates for subtype-specific cancers, where relevant; 3) risk estimates for never-smokers, where relevant; 4) risk estimates were based on participants who underwent imaging (studies using predictive models of adiposity based on a subset or an independent population were excluded); 5) no adjustment for a correlated adiposity measure; and 6) non-duplicative study populations (see **Supplementary Methods** for details).

## Data Availability

All data synthesised in this meta-analysis are provided in the Supplementary Data files.

## Code availability

The statistical code created for this analysis can be obtained at: https://github.com/elliewatts90/BMI_cancer_meta_analysis

## Ethics declarations Competing interests

The authors declare no competing interests.

## Data Availability

All data produced in the present work are contained the Data Supplement

## Data Availability

All data produced in the present work are contained the Data Supplement

## Acknowledgements

EW and SM are supported by the Intramural Research Program of the National Institutes of Health (NIH). NC is supported by the NIH grant U01CA284209. This research was supported in part by the Intramural Research Program of the National Institutes of Health (NIH). The contributions of the NIH author(s) are considered Works of the United States Government. The findings and conclusions presented in this paper are those of the author(s) and do not necessarily reflect the views of the NIH or the U.S. Department of Health and Human Services.

