## Supplementary Methods, Figures, and Tables for "Adiposity and cancer: systematic review and meta-analysis"

### Supplementary materials

#### Supplementary Tables

### Supplementary Methods

#### 1. Cancer selection

We selected cancer types *a priori* based on the 25 most common cancers by global incidence (https://www.wcrf.org/cancer-trends/worldwide-cancer-data/). Among these, we grouped cancers of the lip, oral cavity, larynx, nasopharynx, oropharynx, and hypopharynx as head and neck cancers per common convention. We subdivided cancers known to have etiologically distinct subtypes ("subtype-specific cancers") as follows: gastric (cardia, non-cardia), breast (pre- and post-menopausal), oesophageal (adenocarcinoma and squamous cell carcinoma), and central nervous system (CNS; glioma and meningioma).

We conducted special analyses for two groups of cancers, smoking-related cancers and prostate cancer, to address biases of particular importance for these cancers. For the four cancers most strongly related to smoking (head and neck, lung, oesophageal squamous cell carcinoma (SQ) and bladder), per Freedman et al.[1], we restricted analyses to never smokers to eliminate or reduce residual confounding by smoking. For prostate cancer, we restricted analyses to aggressive cancers to minimize screening bias, which can occur when healthier men are more likely to be screened and have indolent prostate cancers detected[2]. Aggressive prostate cancer was defined as any of stage 3-4 on the American Joint Committee on Cancer 1992 classification, advanced cancer, advanced or metastatic cancer; metastatic cancer; stage C or D on the Whitmore/Jewett scale; fatal cancer (prostate cancer-specific mortality); high stage or grade; Gleason grade $\geq$ 7). This definition was based on previous WCRF reviews[3].

Ultimately, the following 25 cancer types were investigated: bladder (never-smokers only), breast cancer (pre and postmenopausal), cervix, CNS (glioma and meningioma), colorectal, endometrial, gallbladder, gastric (non-cardia and cardia), head and neck (never-smokers only), kidney, leukaemia, liver, lung (never smokers only), melanoma, multiple myeloma, non-Hodgkin lymphoma (NHL), oesophageal (SQ never-smokers only and adenocarcinoma), ovarian, pancreas, prostate (aggressive), and thyroid.

#### 2. Search Strategy

We conducted a systematic search to identify relevant studies using the PubMed, EMBASE, and Scopus databases. The search covered articles published from the databases' inception through April 23rd, 2025. For each cancer type, studies examining the relationship between BMI and cancer risk were identified using specific search terms, which are detailed in **Supplementary Table 4**.

To ensure our literature search was complete, we searched reference lists of Larrson et al.[4], which included a narrative review of studies published from 2018-2021, and the most recent article for each cancer type. Additionally, we reviewed the WCRF reports and included relevant articles not identified by our initial search strategy. Our methodology adhered to the standard criteria for reporting meta-analyses as outlined in the Preferred Reporting Items for Systematic reviews and

Meta-Analyses (PRISMA) guidelines[5]. Our analysis was not pre-registered because it is a revision of an invited meta-analysis. As a result, it was not possible to prospectively register the protocol[6].

#### 3. Inclusion and exclusion criteria

Our inclusion criteria were for original large-scale prospective analyses utilising individual-level data that explored associations between adult (≥18 years) BMI and the risk of the 25 most common cancers. We did not impose any language restrictions in our search.

Studies were excluded based on the following criteria:

1) Non-cancer incidence outcome (e.g., survival/prognosis/wrong outcome) or non-prospective design.
2) N<50,000 participants in the study cohort or pooled study (unless outside North America, North/West/Central Europe, or East/Southeast Asia).
3) Risk estimates not convertible to continuous estimates, such as where only two BMI categories were reported (e.g., risk estimates comparing obese and non-obese participants only).
4) No risk estimates available for subtypes for subtype-specific cancers: breast cancer (pre and postmenopausal), CNS (glioma and meningioma), gastric (non-cardia and cardia), oesophageal (SQ and adenocarcinoma). In East and Southeast Asia >90% of oesophageal cancers are SQ and >80% of gastric cancers are non-cardia[7], therefore for cohorts in this region, cancers were categorised into respective subtypes if not otherwise stated.
5) No risk estimates for never-smokers for smoking-related cancers: head and neck, lung, oesophageal SQ and bladder.
6) Risk estimates were based on models which adjusted for other measures of adiposity (such as waist circumference) or early-life body size only (due to collinearity of variables with adult BMI).
7) Duplicate study population. Where one cohort had multiple publications identified, the publication with the greatest number of cases was included.

The number of studies excluded based on each exclusion criterion is detailed in **Supplementary Figure 1.** For studies identified in the literature search that reported results for multiple cancers, BMI-cancer associations were extracted for each reported cancer type and screened using the process outlined above.

#### 3. Selection process

We utilized Covidence web-based software (https://www.covidence.org/) for the screening process. After removing duplicates, titles and abstracts were screened against established eligibility criteria. Articles that met these criteria in the initial screening were then subjected to a full-text review, where the eligibility criteria were applied once again. Screening and review were conducted independently by two authors (ELW and SCM), in the case of discrepancies AGF acted as the tie-break.

Eligible articles were extracted into Excel and categorized by cohort and cancer type. For cohorts with multiple publications on the association between BMI and a specific cancer type, the analysis with the largest number of cases was selected by ELW.

#### 4. Data extraction

Data was extracted by ELW and AGF independently. For each study, risk estimates (hazard ratios [HRs], risk ratios [RRs] or odds ratios [ORs]) and their 95% confidence intervals (CIs) for the association of BMI with incident cancer risk were extracted, preferentially selecting estimates based on a continuous modelling of BMI. If continuous risk estimates were unavailable and BMI had at least three categories, categorical estimates were extracted. The estimates with the most comprehensive adjustments for confounders that did not also adjust for other measures of adiposity (such as waist circumference), or BMI from an earlier age were extracted.

Where estimates originated from pooled analyses arising from many different cohorts, study-specific risk estimates were extracted whenever possible. However, 15 articles provided only pooled estimates, precluding us from extracting study-specific estimates. For these consortia, we contacted the corresponding authors, who provided us with study-specific estimates for three articles[8–10]. For the remaining articles, we used pooled estimates, provided that the number of cases in the pooled study exceeded those reported in the individual study publications.

When multiple analyses published from the same cohort for each cancer type (e.g., Korean National Health Insurance Service [KNHIS] database), we extracted risk estimates from the study with the most cases. Where studies reported risk estimates for specific cancer subtypes (e.g., colon and rectal cancer) but not for the broader cancer group (e.g., colorectal cancer), we extracted and meta-analysed associations for each subtype to produce a single pooled estimate. This approach was adopted to maximise the use of available data and to avoid inflating heterogeneity in the primary meta-analysis.

For haematological cancers, which have been recently reclassified[11], we adhered to ICD-10 conventions where possible to ensure consistency across studies and to allow the inclusion of earlier research. The classifications used were as follows: NHL (C82-C85), multiple myeloma (C90), leukaemia (C91-C95).

The following information was extracted for each study: citation details (first author and publication year), study design (country, geographic region, N cohorts, follow-up duration [median preferred], large administrative health database study [yes; no]), outcome (cancer type and subtype, if applicable, N cases, N analytic set and the scale reported in the model), and adjustment covariates.

For studies included in our BMI-cancer analysis that investigated ≥5 cancer types and reported risk estimates for both BMI and waist circumference on the same scale or provided continuous risk estimates that could be converted to standard deviations (SDs), we extracted both sets of risk estimates. We also reviewed studies flagged by our search strategy but excluded from the main BMI-cancer analysis due to overlapping study populations, as exclusion based on missing waist circumference data occasionally resulted in smaller analytic cohorts and complementary findings[12]. In this analysis, we included risk estimates for smoking-related cancers regardless of smoking status, given evidence that smoking may differentially influence overall adiposity and fat distribution[13].

#### 5. Risk harmonisation

Risk estimates were converted to the 5 $kg/m^2$ scale to enable comparison across studies. Where estimates were reported in a different continuous scale (e.g., per 1 standard deviation [SD]), HRs and 95% CIs were rescaled to HR per 5 $kg/m^2$ using the formula: HR per 5 $kg/m^2 = {HR_x}^{5/x}$, where $x$ is equal to the originally reported units. The same method was used for the upper and lower CIs. For HRs reported as categorical estimates, we converted associations to a continuous scale using methods described in detail elsewhere[14]. This process involved estimating the midpoints of each BMI category (when not explicitly stated in the original article) and extracting sample sizes for each category and the continuous dose-response association was then estimated using restricted maximum likelihood via the dosresmeta R package[15].

For our investigation comparing BMI and waist circumference measures, continuous risk estimates were converted to SDs to allow comparison. One study also applied regression dilution corrections for BMI (regression dilution ratio [RDR]= 0.93) and waist circumference (0.81), which would amplify differences between the two measures[12]. To ensure comparability on the same scale, we removed this correction by applying the inverse transformation to the reported RRs and 95% CIs using the formula: $RR_{uncorrected} = (RR_{corrected})^{RDR}$.

All results are available in a harmonised excel file in the **Supplementary Data**.

#### 6. Statistical methods

<u>Meta-analysis</u>

Summary risk estimates for the association of BMI with each cancer type were estimated using random effects meta-analysis, which weights studies based on within-study and between-study variability and calculated a weighted average of individual study effect size. Heterogeneity across studies was assessed using $I^2$, $tau^2$, and the 25th and 75th percentile (interquartile range) of RRs for each cancer type. $I^2$ estimates the percent of total variation due to true differences in effect sizes between studies; an $I^2$ close to 100% indicates that chance plays little role in observed differences. However, $I^2$ is highly sensitive to sample size, when studies are large, even small differences can yield large $I^2$ values[16]. $Tau^2$ more directly quantifies differences in effect sizes, it is the between-study variance of the estimated underlying distribution of true effect sizes. A value close to 0 suggests that the true effect size varies little between populations. This metric was calculated using the Paule-Mandel estimator[17]. Finally, we present the interquartile range as a more intuitive metric, showing the typical spread of estimates.

<u>Heterogeneity by region</u>

We assessed heterogeneity in the associations of BMI and each cancer type by region. Cohorts were clustered into geographic regions: Oceania (Australia), North America (USA and Canada), Europe (Austria, Denmark, Finland, France, Germany, Greece, Italy, Netherlands, Norway, Lithuania, Spain, Sweden, UK), East Asia (Japan, South Korea, China, Taiwan, Singapore), South Asia (India), Middle East (Iran and Israel). Although Singapore is geographically located in Southeast Asia, the only study identified was the Singapore Chinese Health Study, which had a relatively small study population (N=51,000) and was specifically recruited from the Chinese population of Singapore. Therefore, Singapore was grouped with East Asia. Heterogeneity in the associations by geographic region was quantified using the Q-statistic.

<u>Heterogeneity in the associations of BMI and waist circumference with cancer risk</u>

The associations of BMI and waist circumference were based on the same population; therefore, heterogeneity was tested using the Wald statistic:

$$Wald\ statistic = \frac{\beta_1 - \beta_2}{\sqrt{Var(\beta_1) + Var(\beta_2) - 2Cov(\beta_1, \beta_2)}}$$

Where:

$\beta_1$ = log RR per 1 standard deviation increase in BMI

$\beta_2$= log RR per 1 standard deviation increase in waist circumference

$Cov = r \times \sqrt{Var(\beta_1) \times Var(\beta_2)}$

$r$= correlation coefficient of BMI and waist circumference ($r$= 0.87)[18]

<u>Assessment of study bias</u>

We assessed study bias using a simplified version of the Berrington de González et al. framework originally developed to review ionizing radiation and cancer risk[19]. Briefly, this framework considers four major sources of epidemiological bias and evaluates their potential relevance to study findings, including the likely direction and magnitude of any bias. In our assessment, substantial selection bias, information bias (referred to as “dose error”), and outcome ascertainment bias were considered unlikely, as our analysis was restricted to prospective cohort studies. Confounding was deemed a more serious concern, along with the possibility of reverse causation, whereby preclinical cancer might lead to weight loss, inducing a spurious inverse association between BMI and cancer risk.

To evaluate these concerns, we applied a two-point grading system to each study, evaluating:

i) adjustment for major confounders, and
ii) whether follow-up exceeded 5-years, which should minimize reverse causation.

To develop our lists of cancer-specific confounders, we considered both the direction and, importantly the magnitude of confounding needed to explain BMI-cancer associations, as recommended by Berrington de González et al. We specifically adopted the approach of Ding and VanderWeele[20], which enables estimation of the magnitude of confounder-disease associations needed to account for exposure-disease associations. For our calculations, we assumed a RR of 1.2 for obesity-cancer associations (vs a reference group of normal weight). This is comparable to a RR of ~1.1 per 5 $kg/m^2$, assuming a 10-unit BMI difference between individuals with obesity and those with normal weight). We also assumed a RR of 1.5 for obesity-confounder associations, a strong but plausible magnitude of effect. Under these assumptions, a confounder would need an RR of 2 or greater with cancer to fully account for the observed association, and we therefore considered any cancer risk factor with a reported RR $\geq 2$ as a potential major confounder.

To ensure a streamlined, high-quality, and systematic approach, we identified candidate confounders by reviewing type-specific chapters in the Schottenfeld and Fraumeni *Cancer Epidemiology and Prevention 4$^{th}$ edition*[21], a widely used text offering standardised expert assessments of cancer risk factors. For assessment of sex as a confounder a recent UK Biobank pan-cancer analysis was used[22]. For those risk factors with a RR $\geq 2$ (**Supplementary Table 5**), we excluded likely mediators (to avoid overadjustment), rare exposures (e.g. liver flukes, which are uncommon in the populations we studied), and exposures not consistently associated with BMI (e.g. ionizing radiation). Occupational exposures were also excluded due to low prevalence. Remaining risk factors were classified as important potential confounders. We also included age a key confounder for all cancers and required that studies either adjusted for age or used age as the underlying time scale in Cox models for assessment of bias[23]. Studies that adjusted for all of these key confounders (or for which no important confounders were identified) received 1 point; otherwise, they received 0 points.

We also assessed whether studies had sufficient follow-up time to mitigate the possibility of reverse causation. Studies with at least five years of follow-up were awarded 1 point, while those with shorter follow-up received 0 points. If information on confounder adjustment or follow-up duration was not reported, no grade was assigned for that criterion.

Assessment of publication bias

We used Egger's test to assess publication bias and applied the trim-and-fill method to re-estimate the effect size by accounting for potentially missing studies. The trim-and-fill method identifies and removes outlying studies to recalculate the overall effect size. Hypothetical studies are then imputed to restore symmetry to the funnel plot, mirroring the distribution of observed studies, and an adjusted effect size is subsequently calculated[24].

Sensitivity analyses

In sensitivity analyses, we restricted the meta-analysis to studies that adjusted for smoking, a factor independently associated with both BMI and many cancers[1]. Some individual study risk estimates were derived entirely from national health databases rather than prospective cohorts, which tend to have more limited control for residual confounding. For example, the Clinical Practice Research Datalink (CPRD) has approximately 50% missing smoking data[25]. Additionally, these databases are often not linked to gold-standard cancer registry data, increasing the possibility of misclassification biases[26,27]. These databases typically comprise large populations that can influence summary estimates. Therefore, sensitivity analyses were performed excluding these health databases. We also examined changes in risk estimates after restricting analyses to studies with a low risk of bias (quality assessment score = 2).

Inclusion details for each study and sensitivity analysis are provided in the **Supplementary Data**.

#### 7. Mendelian randomisation meta-analysis

In secondary analyses, we also investigated associations of BMI with cancer using Mendelian randomisation (MR) approaches. MR leverages genetic variants as instrumental variables to assess potential causal relationships, reducing the risk of confounding and reverse causation that can affect observational studies[29].

<u>Search strategy</u>

For each of the cancer types included in our BMI-cancer meta-analysis, we conducted a systematic search for MR studies. The literature search was conducted in PubMed, EMBASE, and Scopus, covering articles published from the databases' inception through February 6, 2025.

The search strategy included the following terms:

PubMed: cancer*[tiab] NOT "non-cancer*" [tiab] AND ("Mendelian randomization"[tiab] OR "Mendelian randomization"[ot]) AND ("body mass index"[ti] OR bmi[ti] OR ((obesity[ti] OR adipos*[ti] OR body fat*[ti] OR body weight[ti] OR body mass[ti] OR body composition[ti] OR lifestyle[ti] OR anthropom*[ti]) AND (bmi[tiab] OR bmi[ot] OR "body mass index"[tiab:~0] OR "body mass index"[ot]))) NOT (patient*[ti] OR prognosis[ti] OR surg*[ti] OR complication*[ti] OR surviv*[ti] OR screen*[ti] OR infection*[ti] OR virus[ti] )

EMBASE: 'cancer*':ti,ab,kw AND 'Mendelian randomization':ti,ab,kw AND ('body mass index':ti OR 'bmi':ti OR (('obesity':ti OR 'adipos*':ti OR 'body fat*':ti OR 'body weight':ti OR 'body mass':ti OR 'body composition':ti OR 'lifestyle':ti OR 'anthropom*':ti) AND ('bmi':ti,ab,kw OR 'body mass index':ti,ab,kw))) NOT ('patient*':ti OR 'prognosis':ti OR 'surg*':ti OR 'complication*':ti OR 'surviv*':ti OR 'screen*':ti OR 'infection*':ti OR 'virus':ti) AND 'article'/it

Scopus: TITLE-ABS-KEY(cancer*) AND NOT TITLE-ABS-KEY("non-cancer") AND TITLE-ABS-KEY("Mendelian randomization") AND (TITLE ("body mass index" OR {bmi}) OR (TITLE("obesity" OR "adipos*" OR "body fat*" OR "body weight" OR "body mass" OR "body composition" OR "lifestyle" OR "anthropom*") AND TITLE-ABS-KEY({bmi} OR {body mass index}))) AND NOT TITLE(patient* OR "prognosis" OR surg* OR complication* OR surviv* OR screen* OR infection* OR "virus") AND (LIMIT-TO(DOCTYPE,"ar")

MR studies not captured by these searches were identified using reference review.

Exclusion criteria

The exclusion criteria were based on the BMI-cancer analysis, with the following criteria applied:

1) Non-cancer incidence outcome, not top 25 cancer, or no BMI RRs.
2) No risk estimates available for subtypes for subtype-specific cancers.
3) Non-linear ORs.
4) Duplicative study population in the outcome population.

Articles underwent title and abstract screening by ELW, with those meeting the inclusion criteria proceeding to full-text review. Data was extracted by ELW, given prior experience in this methodology.

Data extraction and meta-analysis

The following information was extracted for each study: citation details (first author and publication year), genetic instrument (study, N participants, ancestry, PMID of original GWAS), outcome (study, cancer type, N cases, N controls, ancestry, PMID of original GWAS), and results (OR, 95% CI, model [e.g., inverse-variance weighted random effects], originally reported units). Univariable MR risk estimates were extracted, with preference given to those based on the inverse variance weighted method, converted to the 5 $kg/m^2$ scale for comparability across studies, and meta-analysed using random effects models (as described in **Section 6**). Heterogeneity between studies was assessed using $I^2$.

For smoking-related cancers, when studies provided multivariable MR analyses adjusted for genetically predicted smoking, we preferentially extracted these estimates (lung and head and neck cancers), else univariable MR estimates were extracted (bladder and oesophageal SQ).

The number of included studies for each cancer was low (maximum five) and primarily limited to European ancestry populations. Therefore, we did not assess publication bias or heterogeneity by ancestry. Additionally, as this analysis is secondary, we did not include a study bias evaluation.

#### 8. Assessment of certainty

Standard tools such as GRADE are designed for intervention studies and automatically rate observational evidence as "low certainty"[30]. For exposures like BMI, where randomised controlled trials are not feasible, this could undervalue robust prospective data. We therefore applied a tailored certainty assessment that emphasises domains most relevant to observational epidemiology, adapted from the GRADE framework: large effect size, risk of bias from confounding or reverse causation, consistency, risk of publication bias, and concordance with complementary MR evidence. GRADE also considers three additional domains—precision, directness, and dose-response. However, because we restricted inclusion to studies with analytic populations of ≥50,000, we did not apply an additional minimum case-count criterion. Moreover, all included studies directly addressed the exposure–outcome question of interest, so we did not consider indirectness to be a serious limitation We also did not assess dose–response separately, as our primary analyses already modelled BMI continuously per 5 kg/m$^2$. Our certainty criteria were as follows:

| Certainty level | Criteria |
|---|---|
| High | Large effect size: RR > abs(10%)<br>Risk of bias: Association remains following exclusion of studies with bias score < 2<br>Consistency:<br>- For positive associations (RR > 1): 25$^{th}$ percentile of study-specific RRs > 1<br>- For inverse associations (RR < 1): 75$^{th}$ percentile of study-specific RRs < 1<br>Publication bias: association remains following fill-and-trim analysis<br>Concordance with complementary MR evidence: MR studies are directionally consistent* |
| Moderate | RR > abs(5%) but all other criteria from "High" are met |
| Low | Association nominally significant (p < 0.05) but did not meet the criteria for "High" or "Moderate" certainty |
| Likely null | Limited evidence of an association (p ≥ 0.05) |

*Cancer types supported by strong observational data were not downgraded if MR analyses were unavailable. For postmenopausal breast cancer, we did not require MR findings to be directionally consistent, given the well-established protective association of early-life BMI with breast cancer risk[31].

#### 9. Imaging studies

In secondary analyses, we also assessed the evidence for associations between imaging-based body composition measures and cancer risk.

Search strategy

Our literature search was conducted in PubMed, EMBASE, and Scopus, covering articles published from the databases' inception through February 6, 2025.

The search strategy included the following terms:

PubMed: cancer*[tiab] AND ((adiposity) OR (body fat) OR (body composition)) AND ((DXA scan) OR (DEXA scan) OR (DEXA) OR (DXA) OR (CT scan) OR (MRI)) AND (RR[tiab] OR RR[ot] OR "relative risk"[tiab] OR "relative risk"[ot] OR "hazard ratio"[tiab] OR "hazard ratio"[ot] OR ("HR"[tiab] NOT "heart rate"[tiab])) NOT (child*[ti] OR patient*[ti] OR prognosis[ti] OR surg*[ti] OR complication*[ti] OR surviv*[ti] OR screen*[ti] OR infection*[ti] OR virus[ti])

EMBASE: 'cancer*':ti,ab,kw AND ('adiposity':ti,ab,kw OR 'body fat':ti,ab,kw OR 'body composition':ti,ab,kw) AND ('DXA scan':ti,ab,kw OR 'DEXA scan':ti,ab,kw OR 'DEXA':ti,ab,kw OR 'DXA':ti,ab,kw OR 'CT scan':ti,ab,kw OR 'MRI':ti,ab,kw) AND ('RR':ti,ab,kw OR ‘relative risk’:ti,ab,kw OR 'hazard ratio':ti,ab,kw OR ('HR':ti,ab NOT 'heart rate':ti,ab)) NOT ('child*':ti OR 'patient*':ti OR 'prognosis':ti OR 'surg*':ti OR 'complication*':ti OR 'surviv*':ti OR 'screen*':ti OR 'infection*':ti OR 'virus':ti) AND 'article'/it

Scopus: TITLE-ABS-KEY(cancer*) AND TITLE-ABS-KEY("adipos*" OR "body fat*" OR "body composition") AND TITLE-ABS-KEY(“DXA scan” OR “DEXA scan” OR “DEXA” OR “DXA” OR “CT scan” OR “MRI”) AND (TITLE-ABS-KEY( “RR” OR “relative risk” OR “hazard ratio”) OR TITLE-ABS("HR" AND NOT “heart rate”)) AND NOT TITLE(child* OR patient* OR "prognosis" OR surg* OR complication* OR surviv* OR screen* OR infection* OR "virus") AND (LIMIT-TO(DOCTYPE,"ar")

The exclusion criteria were aligned with those used in the BMI-cancer analyses, with the following criteria applied:

1) Non-cancer incidence outcome or non-prospective design.
2) No risk estimates available for subtypes for subtype-specific cancers.
3) No risk estimates for never-smokers for smoking-related cancer.
4) Indirect measure of adiposity imaging – e.g., estimated adiposity derived from a subset of participants with imaging data rather than requiring direct imaging for all participants.
5) Adjusted for a correlated measure of adiposity.
6) Duplicate study population.

We did not exclude studies based on the size and did not require risk estimates to be continuous or >2 categories.

Articles first underwent title and abstract screening, with those meeting the inclusion criteria proceeding to full-text review. This process was conducted independently by ELW and SCM, with AGF serving as adjudicator in cases of disagreement.

Data were extracted by ELW following the same rubric used for the BMI-cancer analysis. Given the lack of overlapping data across cancer types, a meta-analysis was not conducted.

#### 10. Software and packages

Data were analysed in R (v.4.3.1) using the ‘meta’ (v7.0.0)[32], and dosresmeta (v4.3.1)[15] R packages, a two-tailed $p<0.05$ was considered statistically significant.

### Supplementary Tables

##### Supplementary Table 1: Bias assessment score for each cancer type

| Cancer type | Key confounders adjusted for, mean (SD) | Long follow-up duration, mean (SD) | Total score*, mean (SD) |
|---|---|---|---|
| Endometrial | 1.00 (0.00) | 0.65 (0.49) | 1.65 (0.49) |
| Oesophageal (adeno) | 0.58 (0.51) | 1.00 (0.00) | 1.58 (0.51) |
| Kidney | 0.81 (0.40) | 0.92 (0.27) | 1.73 (0.45) |
| Gallbladder | 1.00 (0.00) | 1.00 (0.00) | 2.00 (0.00) |
| Gastric (cardia) | 1.00 (0.00) | 1.00 (0.00) | 2.00 (0.00) |
| Liver | 1.00 (0.00) | 0.90 (0.31) | 1.90 (0.31) |
| Breast (postmenopausal) | 1.00 (0.00) | 0.88 (0.33) | 1.88 (0.33) |
| Thyroid | 1.00 (0.00) | 0.93 (0.25) | 1.93 (0.25) |
| Meningioma | 1.00 (0.00) | 0.88 (0.35) | 1.88 (0.35) |
| Colorectal | 1.00 (0.00) | 0.94 (0.24) | 1.94 (0.24) |
| Multiple myeloma | 1.00 (0.00) | 1.00 (0.00) | 2.00 (0.00) |
| Leukaemia | 1.00 (0.00) | 1.00 (0.00) | 2.00 (0.00) |
| Head and neck (never-smokers) | 1.00 (0.00) | 0.30 (0.47) | 1.30 (0.47) |
| Pancreas | 0.89 (0.32) | 0.97 (0.17) | 1.86 (0.36) |
| NHL | 1.00 (0.00) | 1.00 (0.00) | 2.00 (0.00) |
| Ovarian | 1.00 (0.00) | 0.96 (0.20) | 1.96 (0.20) |
| Bladder (never-smokers) | 1.00 (0.00) | 1.00 (0.00) | 2.00 (0.00) |
| Cervix | 0.92 (0.29) | 0.92 (0.29) | 1.83 (0.58) |
| Prostate (aggressive) | 1.00 (0.00) | 0.84 (0.37) | 1.84 (0.37) |
| Glioma | 1.00 (0.00) | 1.00 (0.00) | 2.00 (0.00) |
| Melanoma | 1.00 (0.00) | 1.00 (0.00) | 2.00 (0.00) |
| Gastric (non-cardia) | 0.94 (0.24) | 1.00 (0.00) | 1.94 (0.24) |
| Lung (never-smokers) | 0.94 (0.24) | 0.94 (0.24) | 1.89 (0.32) |
| Breast (premenopausal) | 1.00 (0.00) | 0.88 (0.33) | 1.88 (0.33) |
| Oesophageal (SQ, never-smokers) | 1.00 (0.00) | 1.00 (0.00) | 2.00 (0.00) |

*Bias graded between 0 (high risk of bias) and 2 (low risk of bias) based on adjustment for relevant confounders (yes = 1, no = 0) (see **Supplementary Table 5**) and length of follow-up (>5 years = 1, < 5 years = 1). Scores for individual studies are available in the **Supplementary Data**.

**Supplementary Table 2: Sensitivity analyses by smoking adjustment, cohort type, and study bias**

| Cancer type | Analysis type | N cases | N cohorts | RR per 5 kg/m² (95 CI %) | P-value | Δ RR main analysis |
|---|---|---|---|---|---|---|
| Endometrial | Main analysis | 60,478 | 56 | 1.58 (1.51, 1.67) | < 0.001 | - |
| | Smoking-adjusted only | 30,405 | 22 | 1.65 (1.57, 1.74) | < 0.001 | 0.07 |
| | Traditional cohorts only | 18,812 | 25 | 1.60 (1.51, 1.71) | < 0.001 | 0.02 |
| | Health cohorts only | 41,666 | 31 | 1.53 (1.43, 1.64) | < 0.001 | -0.05 |
| | Low bias studies only | 52,318 | 45 | 1.54 (1.45, 1.64) | < 0.001 | -0.04 |
| Oesophageal (adeno) | Main analysis | 6,837 | 46 | 1.47 (1.39, 1.56) | < 0.001 | - |
| | Smoking-adjusted only | 3,313 | 16 | 1.46 (1.39, 1.52) | < 0.001 | -0.01 |
| | Traditional cohorts only | 3,159 | 16 | 1.46 (1.33, 1.61) | < 0.001 | -0.01 |
| | Health cohorts only | 3,678 | 30 | 1.48 (1.37, 1.61) | < 0.001 | 0.01 |
| | Low bias studies only | 3,313 | 16 | 1.46 (1.39, 1.52) | < 0.001 | -0.01 |
| Kidney | Main analysis | 61,233 | 52 | 1.30 (1.26, 1.33) | < 0.001 | - |
| | Smoking-adjusted only | 34,449 | 22 | 1.29 (1.25, 1.33) | < 0.001 | -0.01 |
| | Traditional cohorts only | 8,447 | 21 | 1.31 (1.27, 1.34) | < 0.001 | 0.01 |
| | Health cohorts only | 52,786 | 31 | 1.28 (1.20, 1.37) | < 0.001 | -0.02 |
| | Low bias studies only | 33,902 | 20 | 1.29 (1.26, 1.34) | < 0.001 | -0.01 |
| Gallbladder | Main analysis | 11,028 | 69 | 1.27 (1.23, 1.32) | < 0.001 | - |
| | Smoking-adjusted only | 6,998 | 40 | 1.23 (1.18, 1.29) | < 0.001 | -0.04 |
| | Traditional cohorts only | 2,594 | 39 | 1.24 (1.18, 1.31) | < 0.001 | -0.03 |
| | Health cohorts only | 8,434 | 30 | 1.29 (1.22, 1.37) | < 0.001 | 0.02 |
| | Low bias studies only | 11,028 | 69 | 1.27 (1.23, 1.32) | < 0.001 | 0 |
| Gastric (cardia) | Main analysis | 6,153 | 49 | 1.23 (1.11, 1.36) | < 0.001 | - |
| | Smoking-adjusted only | 2,775 | 19 | 1.17 (1.02, 1.34) | 0.025 | -0.06 |
| | Traditional cohorts only | 2,601 | 19 | 1.26 (1.11, 1.43) | < 0.001 | 0.03 |
| | Health cohorts only | 3,552 | 30 | 1.17 (0.98, 1.40) | 0.078 | -0.06 |
| | Low bias studies only | 6,153 | 49 | 1.23 (1.11, 1.36) | < 0.001 | 0 |
| Liver | Main analysis | 74,353 | 54 | 1.20 (1.12, 1.28) | < 0.001 | - |
| | Smoking-adjusted only | 67,175 | 23 | 1.19 (1.10, 1.29) | < 0.001 | -0.01 |
| | Traditional cohorts only | 12,025 | 23 | 1.21 (1.11, 1.32) | < 0.001 | 0.01 |
| | Health cohorts only | 62,328 | 31 | 1.18 (1.07, 1.29) | < 0.001 | -0.02 |
| | Low bias studies only | 73,879 | 51 | 1.22 (1.14, 1.31) | < 0.001 | 0.02 |
| Breast (postmenopausal) | Main analysis | 206,378 | 75 | 1.15 (1.12, 1.17) | < 0.001 | - |
| | Smoking-adjusted only | 149,435 | 35 | 1.15 (1.12, 1.19) | < 0.001 | 0 |
| | Traditional cohorts only | 91,010 | 46 | 1.15 (1.13, 1.18) | < 0.001 | 0 |
| | Health cohorts only | 115,368 | 29 | 1.10 (1.03, 1.19) | 0.008 | -0.05 |
| | Low bias studies only | 202,229 | 70 | 1.15 (1.12, 1.17) | < 0.001 | 0 |
| Thyroid | Main analysis | 31,249 | 70 | 1.12 (1.07, 1.17) | < 0.001 | - |
| | Smoking-adjusted only | 21,124 | 41 | 1.12 (1.07, 1.18) | < 0.001 | 0 |
| | Traditional cohorts only | 14,187 | 38 | 1.12 (1.06, 1.18) | < 0.001 | 0 |
| | Health cohorts only | 17,062 | 32 | 1.12 (1.07, 1.17) | < 0.001 | 0 |
| | Low bias studies only | 29,622 | 67 | 1.12 (1.07, 1.17) | < 0.001 | 0 |
| Meningioma | Main analysis | 8,307 | 33 | 1.11 (1.05, 1.16) | < 0.001 | - |
| | Smoking-adjusted only | 787 | 3 | 1.14 (0.99, 1.30) | 0.059 | 0.03 |
| | Traditional cohorts only | 1,253 | 6 | 1.14 (1.05, 1.24) | 0.002 | 0.03 |
| | Health cohorts only | 7,054 | 27 | 1.08 (1.02, 1.14) | 0.005 | -0.03 |

| | | | | | | |
|---|---|---|---|---|---|---|
| | Low bias studies only | 7,961 | 32 | 1.12 (1.06, 1.18) | < 0.001 | 0.01 |
| Colorectal | Main analysis | 382,393 | 90 | 1.10 (1.08, 1.12) | < 0.001 | - |
| | Smoking-adjusted only | 240,280 | 58 | 1.10 (1.08, 1.13) | < 0.001 | 0 |
| | Traditional cohorts only | 68,717 | 58 | 1.11 (1.09, 1.14) | < 0.001 | 0.01 |
| | Health cohorts only | 313,676 | 32 | 1.07 (1.04, 1.09) | < 0.001 | -0.03 |
| | Low bias studies only | 380,633 | 88 | 1.10 (1.08, 1.12) | < 0.001 | 0 |
| Multiple myeloma | Main analysis | 32,630 | 45 | 1.10 (1.08, 1.11) | < 0.001 | - |
| | Smoking-adjusted only | 12,231 | 13 | 1.07 (1.05, 1.10) | < 0.001 | -0.03 |
| | Traditional cohorts only | 5,294 | 12 | 1.08 (1.05, 1.12) | < 0.001 | -0.02 |
| | Health cohorts only | 27,336 | 33 | 1.10 (1.07, 1.12) | < 0.001 | 0 |
| | Low bias studies only | 32,630 | 45 | 1.10 (1.08, 1.11) | < 0.001 | 0 |
| Leukaemia | Main analysis | 42,771 | 41 | 1.09 (1.05, 1.13) | < 0.001 | - |
| | Smoking-adjusted only | 17,604 | 11 | 1.11 (1.05, 1.17) | < 0.001 | 0.02 |
| | Traditional cohorts only | 5,264 | 9 | 1.09 (1.02, 1.17) | 0.013 | 0 |
| | Health cohorts only | 37,507 | 32 | 1.09 (1.06, 1.11) | < 0.001 | 0 |
| | Low bias studies only | 42,771 | 41 | 1.09 (1.05, 1.13) | < 0.001 | 0 |
| Head and neck (never-smokers) | Main analysis | 11,872 | 48 | 1.07 (1.02, 1.13) | 0.007 | - |
| | Traditional cohorts only | 1,108 | 19 | 1.13 (1.05, 1.23) | 0.001 | 0.06 |
| | Health cohorts only | 10,764 | 29 | 1.04 (0.98, 1.10) | 0.246 | -0.03 |
| | Low bias studies only | 11,076 | 32 | 1.04 (0.99, 1.10) | 0.098 | -0.03 |
| Pancreas | Main analysis | 64,271 | 68 | 1.07 (1.05, 1.09) | < 0.001 | - |
| | Smoking-adjusted only | 40,019 | 39 | 1.07 (1.04, 1.10) | < 0.001 | 0 |
| | Traditional cohorts only | 18,052 | 38 | 1.09 (1.06, 1.11) | < 0.001 | 0.02 |
| | Health cohorts only | 46,219 | 30 | 1.05 (1.01, 1.09) | 0.02 | -0.02 |
| | Low bias studies only | 39,835 | 38 | 1.07 (1.05, 1.10) | < 0.001 | 0 |
| NHL | Main analysis | 68,755 | 49 | 1.05 (1.01, 1.08) | 0.004 | - |
| | Smoking-adjusted only | 26,553 | 12 | 1.04 (1.02, 1.07) | < 0.001 | -0.01 |
| | Traditional cohorts only | 24,982 | 17 | 1.04 (1.00, 1.09) | 0.047 | -0.01 |
| | Health cohorts only | 43,773 | 32 | 1.05 (1.00, 1.09) | 0.038 | 0 |
| | Low bias studies only | 68,755 | 49 | 1.05 (1.01, 1.08) | 0.004 | 0 |
| Ovarian | Main analysis | 46,535 | 49 | 1.05 (1.02, 1.08) | < 0.001 | - |
| | Smoking-adjusted only | 20,522 | 16 | 1.08 (1.04, 1.12) | < 0.001 | 0.03 |
| | Traditional cohorts only | 9,267 | 18 | 1.05 (1.02, 1.09) | < 0.001 | 0 |
| | Health cohorts only | 37,268 | 31 | 1.05 (1.00, 1.10) | 0.073 | 0 |
| | Low bias studies only | 46,368 | 48 | 1.05 (1.03, 1.08) | < 0.001 | 0 |
| Bladder (never-smokers) | Main analysis | 22,221 | 34 | 1.04 (1.01, 1.07) | 0.012 | - |
| | Traditional cohorts only | 1,610 | 5 | 1.04 (0.98, 1.11) | 0.211 | 0 |
| | Health cohorts only | 20,611 | 29 | 1.04 (0.99, 1.09) | 0.09 | 0 |
| | Low bias studies only | 22,221 | 34 | 1.04 (1.01, 1.07) | 0.012 | 0 |
| Cervix | Main analysis | 22,979 | 37 | 1.04 (0.99, 1.10) | 0.125 | - |
| | Smoking-adjusted only | 10,409 | 8 | 1.06 (1.03, 1.09) | < 0.001 | 0.02 |
| | Traditional cohorts only | 1,624 | 6 | 1.03 (0.91, 1.16) | 0.659 | -0.01 |
| | Health cohorts only | 21,355 | 31 | 1.04 (0.99, 1.10) | 0.14 | 0 |
| | Low bias studies only | 22,489 | 36 | 1.04 (0.99, 1.09) | 0.132 | 0 |
| Prostate (aggressive) | Main analysis | 33,746 | 60 | 1.04 (1.00, 1.09) | 0.047 | - |
| | Smoking-adjusted only | 17,600 | 13 | 1.04 (0.99, 1.10) | 0.154 | 0 |
| | Traditional cohorts only | 19,964 | 33 | 1.04 (1.00, 1.08) | 0.077 | 0 |
| | Health cohorts only | 13,782 | 27 | 1.16 (0.85, 1.58) | 0.344 | 0.12 |
| | Low bias studies only | 29,953 | 57 | 1.05 (1.02, 1.09) | 0.002 | 0.01 |
| Glioma | Main analysis | 16,846 | 37 | 1.03 (1.01, 1.05) | < 0.001 | - |

| | | | | | | |
|---|---|---|---|---|---|---|
| | Smoking-adjusted only | 5,663 | 5 | 1.06 (1.02, 1.10) | 0.001 | 0.03 |
| | Traditional cohorts only | 2,420 | 9 | 1.02 (1.00, 1.04) | 0.045 | -0.01 |
| | Health cohorts only | 14,426 | 28 | 1.08 (0.96, 1.22) | 0.215 | 0.05 |
| | Low bias studies only | 16,846 | 37 | 1.03 (1.01, 1.05) | < 0.001 | 0 |
| Melanoma | Main analysis | 77,022 | 43 | 0.99 (0.95, 1.03) | 0.626 | - |
| | Smoking-adjusted only | 24,332 | 9 | 1.02 (0.97, 1.07) | 0.523 | 0.03 |
| | Traditional cohorts only | 17,921 | 13 | 0.99 (0.95, 1.03) | 0.662 | 0 |
| | Health cohorts only | 59,101 | 30 | 1.05 (0.86, 1.28) | 0.651 | 0.06 |
| | Low bias studies only | 77,022 | 43 | 0.99 (0.95, 1.03) | 0.626 | 0 |
| Gastric (non-cardia) | Main analysis | 66,866 | 54 | 0.98 (0.94, 1.03) | 0.437 | - |
| | Smoking-adjusted only | 58,683 | 24 | 0.98 (0.93, 1.04) | 0.544 | 0 |
| | Traditional cohorts only | 16,449 | 24 | 0.98 (0.93, 1.04) | 0.516 | 0 |
| | Health cohorts only | 50,417 | 30 | 0.99 (0.93, 1.05) | 0.743 | 0.01 |
| | Low bias studies only | 65,825 | 53 | 0.98 (0.93, 1.02) | 0.312 | 0 |
| Lung (never-smokers) | Main analysis | 12,873 | 53 | 0.92 (0.86, 0.99) | 0.017 | - |
| | Traditional cohorts only | 5,329 | 25 | 0.93 (0.87, 1.00) | 0.066 | 0.01 |
| | Health cohorts only | 7,544 | 28 | 0.89 (0.76, 1.06) | 0.19 | -0.03 |
| | Low bias studies only | 12,427 | 51 | 0.94 (0.89, 1.00) | 0.049 | 0.02 |
| Breast (premenopausal) | Main analysis | 123,338 | 77 | 0.92 (0.89, 0.95) | < 0.001 | - |
| | Smoking-adjusted only | 57,371 | 30 | 0.94 (0.90, 0.99) | 0.014 | 0.02 |
| | Traditional cohorts only | 39,475 | 47 | 0.92 (0.89, 0.96) | < 0.001 | 0 |
| | Health cohorts only | 83,863 | 30 | 0.91 (0.85, 0.98) | 0.014 | -0.01 |
| | Low bias studies only | 119,943 | 72 | 0.92 (0.89, 0.96) | < 0.001 | 0 |
| Oesophageal (SQ, never-smokers) | Main analysis | 988 | 38 | 0.72 (0.60, 0.86) | < 0.001 | - |
| | Traditional cohorts only | 874 | 12 | 0.75 (0.61, 0.93) | 0.008 | 0.03 |
| | Health cohorts only | 114 | 26 | 0.61 (0.44, 0.85) | 0.003 | -0.11 |
| | Low bias studies only | 988 | 38 | 0.72 (0.60, 0.86) | < 0.001 | 0 |

Risk estimates calculated using random effects meta-analysis, restricted to:

i) RRs which included some adjustment for smoking

ii) RRs based on traditional prospective cohort studies (i.e., not recruited directly from large regional or national healthcare databases)

iii) RRs based solely on large healthcare databases.

iv) RRs with a low risk of bias (bias grade = 2)

Categorisation for each study is available from the **Supplementary Data**.

*Aggressive prostate cancer was defined as any of stage 3-4 on the American Joint Committee on Cancer 1992 classification, advanced cancer, advanced or metastatic cancer; metastatic cancer; stage C or D on the Whitmore/Jewett scale; fatal cancer (prostate cancer-specific mortality); high stage or grade; Gleason grade ≥7).

Abbreviations: CI=confidence interval, NHL=non-Hodgkin's Lymphoma, RR=risk ratio, SQ=squamous cell carcinoma.

#### Supplementary Table 3: Certainty assessment from meta-analysis of BMI and cancer risk

| Certainty | Positive association | Negative association |
|---|---|---|
| High | Endometrial<br>Oesophageal (adenocarcinoma)<br>Kidney<br>Gallbladder<br>Gastric (cardia)<br>Liver<br>Breast (postmenopausal)<br>Thyroid<br>Meningioma<br>Colorectal<br>Multiple myeloma | |
| Moderate | Leukaemia<br>Pancreas<br>Ovarian<br>NHL | Breast (pre-menopausal)<br>Lung (never-smokers)<br>Oesophageal (SQ, never-smokers) |
| Low | Head and neck (never-smokers)<br>Prostate (aggressive*)<br>Bladder (never-smokers)<br>Glioma | |
| Likely null | Melanoma<br>Cervix<br>Gastric (non-cardia) | |

Criteria to assess certainty are outlined in the **Supplementary Methods**.

*Aggressive prostate cancer was defined as any of stage 3-4 on the American Joint Committee on Cancer 1992 classification, advanced cancer, advanced or metastatic cancer; metastatic cancer; stage C or D on the Whitmore/Jewett scale; fatal cancer (prostate cancer-specific mortality); high stage or grade; Gleason grade ≥7).

Head and neck cancer was downgraded to low certainty due to limited consistency across studies ($25^{th}$ percentile RR per 5 $kg/m^2$ = 0.94), evidence of bias from confounding or reverse causation (association attenuated after excluding studies with bias score < 2; RR = 1.04, 95% CI 0.99–1.10), and lack of support from Mendelian randomisation (RR = 0.93, 95% CI 0.77–1.12). Oesophageal SQ was downgraded due to a lack of precision from MR studies (0.99, 0.38-2.55).

Abbreviations: CI=confidence interval, NHL=non-Hodgkin's Lymphoma, RR=risk ratio, SQ=squamous cell carcinoma.

#### Supplementary Table 4: Adiposity imaging studies identified by search strategy and exclusions

| Cancer | Cohort | Country | Technology | N total | N cases | Measure | Units | RR (95% CI) | Follow-up time | Adjustments | Bias assessment* | PMID |
|---|---|---|---|---|---|---|---|---|---|---|---|---|
| Breast (post-menopausal) | Women's Health Initiative | USA | DXA | 10,931 | 639 | Whole body fat mass | 1 SD | 1.21 (1.12, 1.30) | 15.0 yrs (med) | Age at enrolment, education, race/ethnicity, family history of breast cancer, age at menarche, age at first full-term birth, parity, age at menopause, oral contraceptive use, hormone therapy use, physical activity, alcohol intake, smoking, and study component | 2 | 31875358 |
| | | | | | | Whole body fat percent | 1 SD | 1.16 (1.07, 1.26) | | | | |
| | | | | | | Trunk fat mass | 1 SD | 1.21 (1.12, 1.31) | | | | |
| | | | | | | Fat mass of right leg | 1 SD | 1.16 (1.08, 1.25) | | | | |
| | | | | | | Fat mass of left leg | 1 SD | 1.16 (1.07, 1.25) | | | | |
| | | | | | | Fat mass index ($kg/m^2$) | 1 SD | 1.18 (1.09, 1.27) | | | | |
| | | | | | | Trunk fat mass index ($kg/m^2$) | 1 SD | 1.19 (1.10, 1.28) | | | | |
| Colorectal | Women's Health Initiative | USA | DXA | 11,124 | 169 | Whole-body fat mass | Q4 vs Q1 | 1.39 (0.86–2.23) | 12.9 yrs (med) | Age at enrolment, education, ethnicity, family history of colorectal cancer, hormone therapy, physical activity, pack-years of smoking, alcohol intake, history of diabetes, intake of energy, and randomization status | 2 | 23546610 |
| | | | | | | Whole body fat percent | Q4 vs Q1 | 1.07 (0.68–1.69) | | | | |
| | | | | | | Trunk fat mass | Q4 vs Q1 | 1.31 (0.82–2.10) | | | | |
| | | | | | | Fat mass of right leg | Q4 vs Q1 | 1.04 (0.66–1.65) | | | | |
| | | | | | | Fat mass of left leg | Q4 vs Q1 | 1.00 (0.64–1.57) | | | | |
| | | | | | | Trunk fat mass: average of R and L leg fat mass | Q4 vs Q1 | 1.09 (0.70–1.70) | | | | |
| Pancreas | UK Biobank | UK | MRI | 42,599 | 52 | IPFD | 1 quintile increase | 1.55 (1.26, 1.92) | 4.6 yrs (med) | None, quintiles for IPFD and FP based on sex- and age-specific 95th percentile normal upper limit of IPFD | 0 | 38587286 |
| | | | | | | FP | Present (yes, no) | 2.73 (1.55, 4.80) | | | | |
| Prostate (high-grade) | AGES-Reykjavik | Iceland | CT | 1,832 | 43 | Abdominal VAT | 1 SD | 0.98 (0.72, 1.33) | 9 yrs (mean) | Age at study entry, family history of prostate cancer, smoking status, education, physical activity, and physician visit over past 12 months, height | 2 | 31179538 |
| | | | | | | Abdominal SAT | 1 SD | 1.02 (0.76, 1.38) | | | | |
| | | | | | | Thigh, intermuscular | 1 SD | 0.92 (0.66, 1.27) | | | | |
| | | | | | | Thigh, subcutaneous | 1 SD | 1.14 (0.86, 1.50) | | | | |
| Prostate (advanced) | | | | | 41 | Abdominal VAT | 1 SD | 1.31 (1.00, 1.72) | | | | |
| | | | | | | Abdominal SAT | 1 SD | 1.22 (0.91, 1.63) | | | | |
| | | | | | | Thigh, intermuscular | 1 SD | 1.02 (0.75, 1.40) | | | | |
| | | | | | | Thigh, subcutaneous | 1 SD | 1.25 (0.95, 1.64) | | | | |
| Prostate (fatal) | | | | | 31 | Abdominal VAT | 1 SD | 1.24 (0.89, 1.73) | | | | |
| | | | | | | Abdominal SAT | 1 SD | 1.26 (0.89, 1.78) | | | | |
| | | | | | | Thigh, intermuscular | 1 SD | 1.27 (0.91, 1.78) | | | | |
| | | | | | | Thigh, subcutaneous | 1 SD | 1.37 (1.00, 1.88) | | | | |

*Bias graded between 0 (high risk of bias) and 2 (low risk of bias) based on length of follow-up (>5 years = 1 point) and adjustment for relevant confounders (see **Supplementary Table 5**).

Abbreviations: AGES-Reykjavik=Age, Gene/Environment Susceptibility-Reykjavik, CT=computer tomography, DXA=dual energy X-ray absorptiometry, FP= Fatty change of the pancreas, IPFD=Intrapancreatic fat deposition, MRI=Magnetic resonance imaging, SAT=subcutaneous adipose tissue, SD=standard deviation, VAT=visceral adipose tissue

#### Supplementary Table 5: Literature search terms

| Cancer type | PubMed search terms | EMBASE search terms | Scopus search terms |
|---|---|---|---|
| BODY OF SEARCH | AND ("body mass index"[ti] OR bmi[ti] OR ((obesity[ti] OR adipos*[ti] OR body fat*[ti] OR body weight[ti] OR body mass[ti] OR body composition[ti] OR lifestyle[ti] OR anthropom*[ti]) AND (bmi[tiab] OR bmi[ot] OR "body mass index"[tiab:~0] OR "body mass index"[ot]))) AND (cohort*[tiab] OR cohort*[ot] OR pool*[tiab] OR million[tiab] OR risk*[ti] OR incidence[ti] OR associat*[ti] ) NOT (child*[ti] OR patient*[ti] OR prognosis[ti] OR surg*[ti] OR complication*[ti] OR surviv*[ti] OR screen*[ti] OR infection*[ti] OR virus[ti] OR odds[tiab] OR "ORs"[tiab] ) NOT ("bmi-1"[ti] OR genome[ti] OR genetic[ti] OR gene[ti] OR Mendelian random*[ti] OR polymorphism*[ti] OR pathway*[ti] OR expression[ti] OR DNA[ti] OR RNA[ti] OR microRNA[ti] OR mRNA[ti] OR "prediction model"[ti]) AND ("journal article"[pt] OR "systematic review"[pt] OR "meta-analysis"[pt]) NOT review[pt] | AND ('body mass index':ti OR 'bmi':ti OR (('obesity':ti OR 'adipos*':ti OR 'body fat*':ti OR 'body weight':ti OR 'body mass':ti OR 'body composition':ti OR 'lifestyle':ti OR 'anthropom*':ti) AND ('bmi':ti,ab,kw OR 'body mass index':ti,ab,kw))) AND ('cohort*':ti,ab,kw OR 'pool*':ti,ab OR 'million':ti,ab OR 'risk*':ti OR 'incidence':ti OR 'associat*':ti) NOT ('child*':ti OR 'patient*':ti OR 'prognosis':ti OR 'surg*':ti OR 'complication*':ti OR 'surviv*':ti OR 'screen*':ti OR 'infection*':ti OR 'virus':ti OR 'odds':ti,ab OR 'ORs':ti,ab) NOT ('bmi-1':ti OR 'genome':ti OR 'genetic':ti OR 'gene':ti OR 'mendelian random*':ti OR 'polymorphism*':ti OR 'pathway*':ti OR 'expression':ti OR 'DNA':ti OR 'RNA*':ti OR 'microRNA':ti OR 'mRNA':ti OR 'prediction model':ti) AND 'article'/it NOT 'review':it | AND (TITLE ("body mass index" OR {bmi} ) OR (TITLE("obesity" OR "adipos*" OR "body fat*" OR "body weight" OR "body mass" OR "body composition" OR "lifestyle" OR "anthropom*") AND TITLE-ABS-KEY({bmi} OR {body mass index}))) AND (TITLE-ABS-KEY(cohort*) OR TITLE-ABS(pool* OR {million}) OR TITLE(risk* OR {incidence} OR associat*)) AND NOT (TITLE(child* OR patient* OR "prognosis" OR surg* OR complication* OR surviv* OR screen* OR infection* OR "virus") OR TITLE-ABS("ORs" OR {odds})) AND NOT TITLE("BMI-1" OR "genome" OR "genetic" OR "gene" OR "mendelian random*" OR polymorphism* OR pathway* OR "expression" OR "DNA" OR "RNA*" OR "microRNA" OR "mRNA" OR "prediction model") AND (LIMIT-TO(DOCTYPE,"ar") |
| **Cancer-specific search terms. Add database-specific *body of search* to each (see below)** | | | |

| Cancer type | PubMed search terms | EMBASE search terms | Scopus search terms |
|---|---|---|---|
| Stomach / gastric: cardia, non-cardia OR Kidney / renal OR Liver / hepatic OR Lung / pulmonary OR Bladder OR Thyroid OR Ovarian / ovary OR Cervical / cervix OR Endometrial / endometrium | "stomach neoplasms"[MeSH] OR "stomach cancer*"[tiab] OR "gastric cancer*"[tiab] OR "gastric cardia"[tiab] OR "gastric non cardia"[tiab] OR "gastric noncardia"[tiab] OR "stomach cancer*"[ot] OR "gastric cancer*"[ot] OR "gastric cardia"[ot] OR "gastric non cardia"[ot] OR "gastric noncardia"[ot] OR "kidney neoplasms"[MeSH:noexp] OR "carcinoma, renal cell"[MeSH] OR "kidney cancer*"[tiab] OR "renal cancer*"[tiab] OR "renal carcinoma*"[tiab] OR "kidney cancer*"[ot] OR "renal cancer*"[ot] OR "renal carcinoma*"[ot] OR "liver neoplasms"[MeSH:noexp] OR "carcinoma, hepatocellular"[MeSH] OR "liver cancer*"[tiab] OR hepatocarcinoma*[tiab] OR hepatoma*[tiab] OR "liver cell carcinoma*"[tiab] OR "liver cancer*"[ot] OR hepatocarcinoma*[ot] OR hepatoma*[ot] OR "liver cell carcinoma*"[ot] OR "lung neoplasms"[MeSH] OR "lung cancer*"[tiab] OR "pulmonary cancer*"[tiab] OR "lung cancer*"[ot] OR "pulmonary cancer*"[ot] OR "urinary bladder neoplasms"[MeSH] OR "bladder cancer*"[tiab] OR "bladder cancer*"[ot] OR "thyroid neoplasms"[MeSH:noexp] OR "Thyroid Cancer, Papillary"[MeSH] OR "thyroid cancer*"[tiab] OR "thyroid papillary cancer"[tiab] OR "thyroid cancer*"[ot] OR "thyroid papillary cancer"[ot] OR "ovarian neoplasms"[MeSH:noexp] OR "ovarian cancer*"[tiab] OR "ovary cancer*"[tiab] OR "ovarian cancer*"[ot] OR "ovary cancer*"[ot] OR "uterine cervical neoplasms"[MeSH] OR "cervical cancer*"[tiab] OR "cervix cancer*"[tiab] OR "cervical cancer*"[ot] OR "cervix cancer*"[ot] OR "endometrial neoplasms"[MeSH] OR "endometr* cancer*"[tiab] OR "endometr* cancer*"[ot] | 'stomach cancer'/exp OR 'stomach cancer*':ti,ab,kw OR 'gastric cancer*':ti,ab,kw OR 'gastric cardia':ti,ab,kw OR 'gastric non cardia':ti,ab,kw OR 'gastric noncardia':ti,ab,kw OR 'kidney cancer'/de OR 'renal cell carcinoma'/exp OR 'kidney cancer*':ti,ab,kw OR 'renal cancer*':ti,ab,kw OR 'renal carcinoma*':ti,ab,kw OR 'liver cancer'/de OR 'liver cell carcinoma'/exp OR 'liver cancer*':ti,ab,kw OR 'hepatocarcinoma*':ti,ab,kw OR 'hepatoma*':ti,ab,kw OR 'liver cell carcinoma*':ti,ab,kw OR 'lung cancer'/exp OR 'lung cancer*':ti,ab,kw OR 'pulmonary cancer*':ti,ab,kw OR 'bladder cancer'/exp OR 'bladder cancer*':ti,ab,kw OR 'thyroid cancer'/de OR 'thyroid papillary carcinoma'/exp OR 'thyroid cancer*':ti,ab,kw OR 'thyroid papillary cancer':ti,ab,kw OR 'ovary cancer'/de OR 'ovarian cancer*':ti,ab,kw OR 'ovary cancer*':ti,ab,kw OR 'uterine cervix cancer'/exp OR 'cervical cancer*':ti,ab,kw OR 'cervix cancer*':ti,ab,kw OR 'endometrium cancer'/exp OR 'endometr* cancer*':ti,ab,kw | TITLE-ABS-KEY ("stomach cancer*" OR "gastric cancer*" OR "gastric cardia" OR "gastric non cardia" OR "gastric noncardia" OR "kidney cancer*" OR "renal cancer*" OR "renal carcinoma*" OR "liver cancer*" OR "hepatocarcinoma*" OR "hepatoma*" OR "liver cell carcinoma*" OR "lung cancer*" OR "pulmonary cancer*" OR "bladder cancer*" OR "thyroid cancer*" OR "thyroid papillary cancer*" OR "ovarian cancer*" OR "ovary cancer*" OR "cervical cancer*" OR "cervix cancer*" OR "endometr* cancer*") |

| Cancer type | PubMed search terms | EMBASE search terms | Scopus search terms |
|---|---|---|---|
| Melanoma OR Multiple myeloma OR Non-Hodgkin lymphoma OR Leukaemia / Leukemia OR meningioma, glioma | "melanoma"[MeSH:noexp] OR melanoma*[tiab] OR melanoma*[ot] OR "multiple myeloma"[MeSH] OR "multiple myeloma*"[tiab] OR "multiple myeloma*"[ot] OR "lymphoma, non hodgkin"[MeSH] OR "non-Hodgkin* lymphoma*"[tiab] OR "nonHodgkin* lymphoma*"[tiab] OR "non-Hodgkin* lymphoma*"[ot] OR "nonHodgkin* lymphoma*"[ot] OR "leukemia"[MeSH] OR "leukemia*"[tiab] OR "leukaemia*"[tiab] OR "leukemia*"[ot] OR "leukaemia*"[ot] OR "meningioma"[MeSH] OR "glioma"[MeSH] OR "meningioma*"[tiab] OR "glioma*"[tiab] OR "meningioma*"[ot] OR "glioma*"[ot] | 'melanoma'/de OR 'melanoma*':ti,ab,kw OR 'multiple myeloma'/exp OR 'multiple myeloma*':ti,ab,kw OR 'non-Hodgkin lymphoma'/exp OR 'non-hodgkin* lymphoma*':ti,ab,kw OR 'nonHodgkin* lymphoma*':ti,ab,kw OR 'leukemia'/exp OR 'leukemia*':ti,ab,kw OR 'leukaemia*':ti,ab,kw OR 'meningioma'/exp OR 'glioma'/exp OR 'meningioma*':ti,ab,kw OR 'glioma*':ti,ab,kw | TITLE-ABS-KEY (melanoma* OR "multiple myeloma*" OR "non-Hodgkin* lymphoma*" OR "nonHodgkin* lymphoma*" OR "leukemia*" OR "leukaemia*" OR "meningioma*" OR "glioma*")…AND body of search… |
| Gallbladder / part of biliary tract | ("gallbladder neoplasms"[MeSH] OR "gallbladder cancer"[tiab:~1] OR "gall bladder cancer"[tiab:~1] OR "biliary tract cancer"[tiab:~1] OR "gallbladder cancers"[tiab:~1] OR "gall bladder cancers"[tiab:~1] OR "biliary tract cancers"[tiab:~1] OR "gallbladder cancer*"[ot] OR "gall bladder cancer*"[ot] OR "biliary tract cancer*"[ot] ) NOT ("gallbladder stone*"[ti]) | ('gallbladder cancer'/exp OR 'gallbladder cancer*':ti,ab,kw OR 'gall bladder cancer*':ti,ab,kw OR 'biliary tract cancer*':ti,ab,kw) NOT ('gallbladder stone*':ti) | TITLE-ABS-KEY (" gallbladder cancer*" OR "gall bladder cancer*" OR "biliary tract cancer*") AND NOT TITLE("gallbladder stone*") |
| Pancreatic / pancreas | ("pancreatic neoplasms"[MeSH:noexp] OR "carcinoma, pancreatic ductal"[MeSH] OR "pancrea* cancer*"[tiab] OR "pancrea* carcinoma*"[tiab] OR "pancrea* adenocarcinoma*"[tiab] OR "pancrea* cancer*"[ot] OR "pancrea* carcinoma*"[ot] OR "pancrea* adenocarcinoma*"[ot]) NOT (pancreatectomy [ti]) | ('pancreas cancer'/de OR 'pancreas adenocarcinoma'/exp OR 'pancrea* cancer*':ti,ab,kw OR 'pancrea* carcinoma*':ti,ab,kw OR 'pancrea* adenocarcinoma*':ti,ab,kw) NOT ('pancreatectomy':ti) | TITLE-ABS-KEY ("pacrea* cancer*" OR "pancrea* carcinoma*" OR "pancrea* adenocarcinoma*" OR "pancrea* ductal") AND NOT TITLE("pancreatectomy") |
| Colorectal / colorectum | ("colorectal neoplasms"[MeSH:noexp] OR "rectal neoplasms"[MeSH] OR "colorect* cancer*"[tiab] OR "colon cancer*"[tiab] OR "rectal cancer*"[tiab] OR "rectum cancer*"[tiab] OR "colorect* cancer*"[ot] OR "colon cancer*"[ot] OR "rectal cancer*"[ot] OR "rectum cancer*"[ot]) NOT ("adenoma*" [ti] OR "polyp*"[ti]) | ('colorectal cancer'/de OR 'rectum cancer'/exp OR 'colorect* cancer*':ti,ab,kw OR 'colon cancer*':ti,ab,kw OR 'rectal cancer*':ti,ab,kw OR 'rectum cancer*':ti,ab,kw) NOT ('adenoma*':ti OR 'polyp*':ti) | TITLE-ABS-KEY ("colorect* cancer*" OR "colon cancer*" OR "rectal cancer*" OR "rectum cancer*") AND NOT TITLE("adenoma*" OR "polyp*") |

| Cancer type | PubMed search terms | EMBASE search terms | Scopus search terms |
|---|---|---|---|
| Oesophagus / Esophagus: adenocarcinoma, squamous cell | "esophageal neoplasms"[MeSH] OR "esophageal squamous cell carcinoma"[MeSH] OR "esophag* cancer*" [tiab] OR "oesophag* cancer*"[tiab] OR "esophag* adenocarcinoma*" [tiab] OR "oesophag* adenocarcinoma*"[tiab] OR "esophag* squamous cell carcinoma*" [tiab] OR "oesophag* squamous cell carcinoma*"[tiab] OR "esophag* cancer*" [ot] OR "oesophag* cancer*"[ot] OR "esophag* adenocarcinoma*"[ot] OR "oesophag* adenocarcinoma*"[ot] OR "esophag* squamous cell carcinoma*"[ot] OR "oesophag* squamous cell carcinoma*"[ot] | esophagus cancer'/exp OR 'esophageal squamous cell carcinoma'/exp OR 'esophag* cancer*':ti,ab,kw OR 'oesophag* cancer*':ti,ab,kw OR 'esophag* adenocarcinoma*':ti,ab,kw OR 'oesophag* adenocarcinoma*':ti,ab,kw OR 'esophag* squamous cell carcinoma*':ti,ab,kw OR 'oesophag* squamous cell carcinoma*':ti,ab,kw | TITLE-ABS-KEY ("esophag* cancer*" OR "oesophag* cancer*" OR "esophag* adenocarcinoma*" OR "oesophag* adenocarcinoma*" OR "esophag* squamous cell carcinoma*" OR "oesophag* squamous cell carcinoma*" ) |
| Head and neck | "Nose Neoplasms"[Mesh] OR "Mouth Neoplasms"[Mesh] OR "Laryngeal Neoplasms"[Mesh] OR "Pharyngeal Neoplasms"[Mesh] OR "nose cancer*"[tiab] OR "nasal cancer*"[tiab] OR "mouth cancer*"[tiab] OR "oral cancer*"[tiab] OR "laryn* cancer*"[tiab] OR "pharyn* cancer*"[tiab] OR "nose cancer*"[ot] OR "nasal cancer*"[ot] OR "mouth cancer*"[ot] OR "oral cancer*"[ot] OR "laryn* cancer*"[ot] OR "pharyn* cancer*"[ot] OR "Head and Neck Neoplasms"[Mesh:noexp] OR "Head Neck cancer"[tiab:~1] OR "Head Neck cancers"[tiab:~1] | nose cancer'/exp OR 'mouth cancer'/exp OR 'larynx cancer'/exp OR 'pharynx cancer'/exp OR 'nose cancer*':ti,ab,kw OR 'nasal cancer*':ti,ab,kw OR 'mouth cancer*':ti,ab,kw OR 'oral cancer*':ti,ab,kw OR 'laryn* cancer*':ti,ab,kw OR 'pharyn* cancer*':ti,ab,kw OR 'head neck cancer*':ti,ab | TITLE-ABS-KEY ("nose cancer*" OR "nasal cancer*" OR "mouth cancer*" OR "oral cancer*" OR "laryn* cancer*" OR "pharyn* cancer*") OR TITLE-ABS(head W/2 neck W/2 cancer*) |
| Breast: pre-, post-menopausal | ("breast neoplasms"[MeSH:noexp] OR "breast cancer*"[tiab] OR "breast cancer*"[ot]) AND ("premenopaus*" OR "postmenopau*" OR "pre-menopaus*" OR "post-menopaus*" OR "menopaus*") | ('breast cancer'/de OR 'breast cancer*':ti,ab,kw) AND ('premenopaus*' OR 'postmenopau*' OR 'pre-menopaus*' OR 'post-menopaus*' OR 'menopaus*') | TITLE-ABS-KEY ("breast cancer*") AND ("premenopaus*" OR "postmenopau*" OR "pre-menopaus*" OR "post-menopaus*" OR "menopaus*") |
| Prostatic / Prostate: aggressive | ("prostatic neoplasms"[MeSH] OR "prostat* cancer*"[tiab] OR "prostat* cancer*"[ot]) AND ("aggressive*" OR "lethal" OR "fatal" OR "death" OR "mortality" OR "advanced" OR "metastatic") | ('prostate cancer'/exp OR 'prostat* cancer*':ti,ab,kw) AND ('aggressive*' OR 'lethal' OR 'fatal' OR 'death' OR 'mortality' OR 'advanced' OR 'metastatic') | TITLE-ABS-KEY ("prostat* cancer*") AND ("aggressive*" OR "lethal" OR "fatal" OR "death" OR "mortality" OR "advanced" OR "metastatic") |

**Supplementary Table 6: Criteria for assessing key confounders for each cancer type**

| Cancer type | Risk factors with strong associations | Reason for exclusion | Confounders for bias check |
|---|---|---|---|
| Endometrial | Age | | Age |
| | Endogenous oestrogens | Mediator | |
| | Oestrogen-only therapy | Uncommon among women with intact uterus | |
| Oesophageal (adenocarcinoma) | Age | | Age, sex, smoking, race/ethnicity* |
| | Sex | | |
| | Smoking | | |
| | Race/ethnicity | | |
| | GERD | Mediator | |
| Kidney | Age | | Age, smoking |
| | Smoking | | |
| | Hypertension | Mediator | |
| Gallbladder | Age | | Age |
| | Gallstones | Mediator | |
| | Liver flukes | Low prevalence in regions examined, incl. in Asia | |
| Gastric (cardia) | Age | | Age, sex |
| | Sex | | |
| | GERD | Mediator | |
| | Barrett's oesophagus | Mediator | |
| Liver cancer | Age | | Age, race/ethnicity* |
| | Race/ethnicity | | |
| | Hepatitis C virus, Hepatitis B virus | Not associated with BMI | |
| | Alcohol | Not associated with BMI | |
| Breast (postmenopausal) | Age | | Age |
| | Endogenous oestrogens | Mediator | |
| | Benign breast disease and other precursors | Mediator | |
| | Nulliparity / older age at menopause | Not strongly associated with BMI | |
| | Older age at first live birth | Confounding in opposite direction - older age at first birth is associated with lower BMI[33] | |
| Thyroid | Age | | Age, sex |
| | Sex | | |
| | Goiter | Mediator | |
| | Genetic susceptibilities (e.g., acromegaly) | Low prevalence | |
| | Ionizing radiation | Not associated with BMI | |
| Meningioma | Age | | Age |
| | Central nervous system exposure to radiation therapy | Not associated with BMI | |
| Colorectal | Age | | Age |

| | | | |
|---|---|---|---|
| | Inflammatory bowel disease | Not strongly associated with BMI and possible mediator | |
| Multiple myeloma | Age<br>Race / ethnicity<br>MGUS and other precursors | <br><br>Mediator | Age, race/ethnicity* |
| Leukaemia | Age<br>Ionizing radiation<br>Severe immune dysregulation | <br>Not associated with BMI<br>Low prevalence | Age |
| Head and neck | Age<br>Sex<br>Smoking<br>Smokeless tobacco<br>Alcohol<br>Human papillomavirus | <br><br><br>Low prevalence and no clear association with BMI<br>Not associated with BMI<br>Not associated with BMI | Age, sex, smoking |
| Pancreas | Age<br>Smoking<br>Chronic pancreatitis<br>Diabetes<br>ABO blood type | <br><br>Mediator<br>Mediator<br>Weak evidence for an association with BMI | Age, smoking |
| NHL | Age<br>Epstein-Barr virus<br>Human immunodeficiency virus | Age<br>Practically ubiquitous—and no strong evidence for associations with BMI[34]<br>Lower than 1% prevalence in each of N. America, Europe, and Asia | Age |
| Ovarian | Age | | Age |
| Bladder | Age<br>Sex<br>Smoking<br>*Schistosoma haematobium*<br>Occupational exposures | <br><br><br>Lower than 1% prevalence in each of N. America, Europe, and Asia<br>Low prevalence | Age, sex, smoking |
| Cervix | Age<br>Human papillomavirus | <br>Not associated with BMI[35] | Age |
| Prostate (aggressive) | Age | | Age |
| Glioma | Age<br>Central nervous system exposure to radiation therapy | <br>No association with BMI | Age |
| Melanoma | Age<br>Race/ethnicity<br>Sun exposure | <br><br>No consistent association with BMI | Age. race/ethnicity* |
| Gastric non-cardia | Age<br>*H. pylori*<br>Gastritis | <br>Not associated with BMI[36,37]<br>Mediator | Age |
| Lung | Age<br>Smoking | | Age, smoking |

| | | | |
|---|---|---|---|
| | Occupational exposures (e.g. asbestos) | Low prevalence | |
| | Radon | Not strongly associated with BMI[38] | |
| | Indoor air pollution | Low prevalence (common in rural China, but included Chinese studies are urban) | |
| Breast (premenopausal) | Age<br>Endogenous oestrogens | Mediator | Age |
| Oesophageal squamous cell carcinoma | Age<br>Smoking<br>Race/ethnicity | | Age, smoking, race/ethnicity* |
| | High dose ionizing radiation (>35 Gy) | Low prevalence | |
| | Alcohol | No strong association with BMI | |
| | Betel nut chewing | Low prevalence in regions of interest | |
| | Exceptionally hot beverages | Low prevalence in regions of interest | |

*Race/ethnicity criterion applied only to cohorts with >80% racial diversity. All non-US cohorts where race was a potentially key confounder were >90% racially homogenous.

Abbreviations: BMI=body mass index, GERD=gastro-oesophageal reflux disease, MGUS=monoclonal gammopathy of undetermined significance, NHL=non-Hodgkin's Lymphoma, RR=risk ratio

### Supplementary Figures

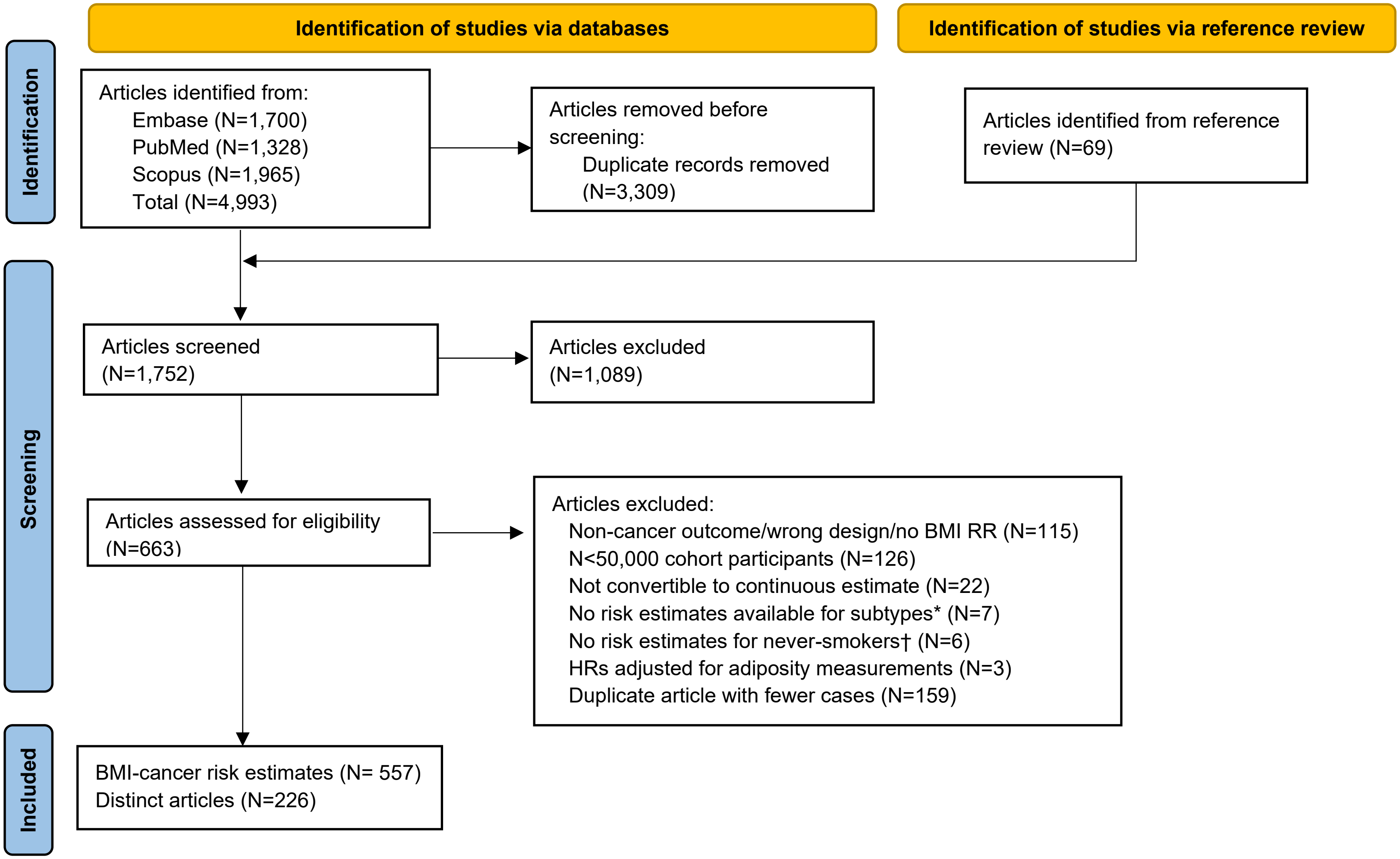

**Supplementary Figure 1: Studies identified by search strategy in observational studies**

Numbers reflect the number of distinct articles. Some cancer-specific risk estimates may be excluded if they did not meet the inclusion criteria for individual cancer types (e.g., an article may contribute risk estimates for one cancer type and be included in the total, but its estimates for another cancer may be excluded if superseded by a study based on the same cohort with more cases).

*Subtypes: breast cancer (pre and postmenopausal), CNS (glioma and meningioma), gastric (non-cardia and cardia), oesophageal (SQ and adenocarcinoma)

† For smoking-related cancers only (lung, head and neck, bladder, oesophageal SQ)

Abbreviations: CNS=central nervous system, HR=hazard ratio, PMID=PubMed ID, SQ=squamous cell carcinoma

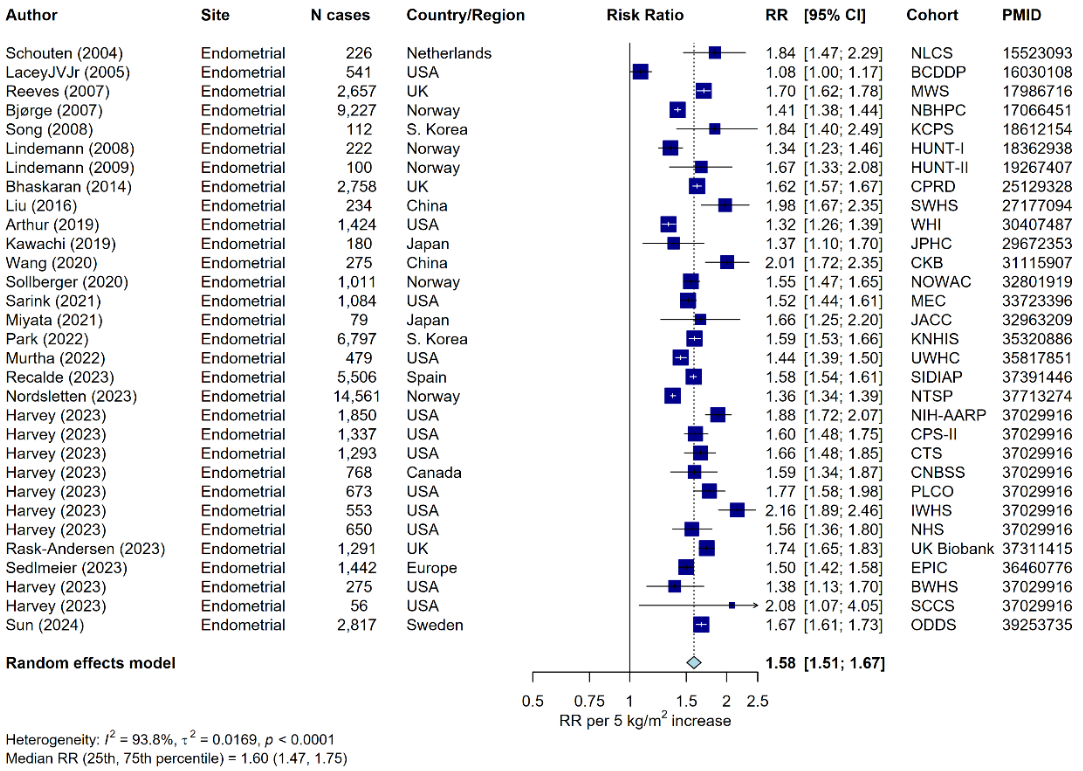

#### Supplementary Figure 2: Meta-analysis of prospective studies on the risk of endometrial cancer in relation to BMI

Study-specific RRs are represented by squares (with their 95% CIs as lines). RRs were combined using weighted averages of the log RRs in the separate studies, with weights assigned based on within and between study variance, yielding a pooled risk estimate and its 95% CI (diamond). Heterogeneity across studies was assessed using $I^2$, $tau^2$, and the $25^{th}$ and $75^{th}$ percentile of the RRs. Further details of model adjustments, follow-up time, analytic population for each study are available from **Supplementary Data**.

Abbreviations: BMI=body mass index, CI=confidence interval, PMID=PubMed ID, RR=risk ratio

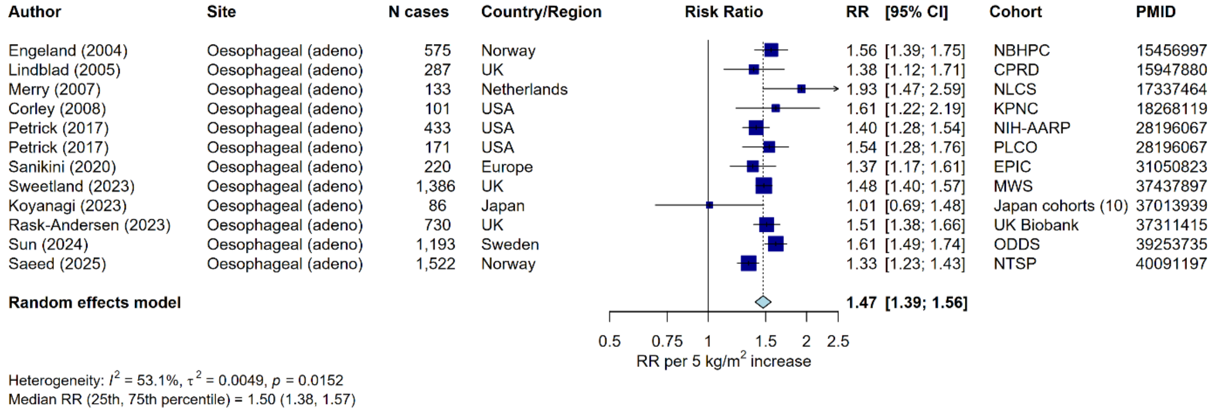

#### Supplementary Figure 3: Meta-analysis of prospective studies on the risk of oesophageal adenocarcinoma cancer in relation to BMI

Study-specific RRs are represented by squares (with their 95% CIs as lines). RRs were combined using weighted averages of the log RRs in the separate studies, with weights assigned based on within and between study variance, yielding a pooled risk estimate and its 95% CI (diamond). Heterogeneity across studies was assessed using $I^2$, $tau^2$, and the 25th and 75th percentile of the RRs. Further details of model adjustments, follow-up time, analytic population for each study are available from **Supplementary Data**.

Abbreviations: BMI=body mass index, CI=confidence interval, PMID=PubMed ID, RR=risk ratio

| Author | Site | N cases | Country/Region | RR | [95% CI] | Cohort | PMID |
|---|---|---|---|---|---|---|---|
| Bjørge (2004) | Renal cell carcinoma | 6,453 | Norway | 1.28 | [1.23; 1.32] | NBHPC | 15583369 |
| vanDijk (2004) | Renal cell carcinoma | 275 | Netherlands | 1.40 | [1.10; 1.76] | NLCS | 15583368 |
| Reeves (2007) | Kidney | 723 | UK | 1.24 | [1.13; 1.36] | MWS | 17986716 |
| Setiawan (2007) | Renal cell carcinoma | 347 | USA | 1.34 | [1.18; 1.54] | MEC | 17656615 |
| Adams (2008) | Renal cell carcinoma | 1,366 | USA | 1.37 | [1.29; 1.47] | NIH-AARP | 18544571 |
| Sawada (2010) | Kidney | 139 | Japan | 1.17 | [0.88; 1.56] | JPHC | 20470974 |
| Andreotti (2010) | Kidney | 106 | USA | 1.05 | [0.81; 1.37] | AgHealth | 20730623 |
| Macleod (2013) | Renal cell carcinoma | 238 | USA | 1.18 | [0.99; 1.40] | VITAL | 23665301 |
| Bhaskaran (2014) | Kidney | 1,906 | UK | 1.25 | [1.19; 1.31] | CPRD | 25129328 |
| Sanfilippo (2014) | Kidney | 407 | USA | 1.22 | [1.11; 1.33] | WHI | 24637660 |
| Guo (2014) | Kidney | 69 | China | 1.28 | [0.91; 1.80] | Kailun Cohort | 24831616 |
| Song (2014) | Kidney | 195 | Finland | 1.34 | [1.10; 1.63] | FINRISK | 24997743 |
| Liu (2016) | Renal cell carcinoma | 131 | China | 1.26 | [0.97; 1.63] | SWHS | 27177094 |
| daSilva (2018) | Kidney | 211 | Norway | 1.33 | [1.15; 1.54] | NOWAC | 30202086 |
| Nam (2019) | Kidney | 18,036 | S. Korea | 1.34 | [1.31; 1.37] | KNHIS | 31231120 |
| Everatt (2019) | Kidney | 79 | Lithuania | 1.40 | [1.05; 1.93] | KRIS, MIHDPS | 31951685 |
| Murtha (2022) | Kidney | 478 | USA | 1.22 | [1.16; 1.29] | UWHC | 35817851 |
| Graff (2022) | Renal cell carcinoma | 394 | USA | 1.41 | [1.25; 1.59] | NHS | 35715363 |
| Graff (2022) | Renal cell carcinoma | 110 | USA | 1.45 | [1.19; 1.75] | NHS-II | 35715363 |
| Graff (2022) | Renal cell carcinoma | 291 | USA | 1.28 | [1.05; 1.56] | HPFS | 35715363 |
| Recalde (2023) | Kidney | 7,065 | Spain | 1.15 | [1.12; 1.19] | SIDIAP | 37391446 |
| Nordsletten (2023) | Kidney | 13,133 | Norway | 1.25 | [1.22; 1.28] | NTSP | 37713274 |
| Rask-Andersen (2023) | Kidney | 1,462 | UK | 1.37 | [1.29; 1.45] | UK Biobank | 37311415 |
| Sedlmeier (2023) | Kidney | 899 | Europe | 1.31 | [1.21; 1.42] | EPIC | 36460776 |
| Deng (2023) | Renal cell carcinoma | 527 | USA | 1.37 | [1.26; 1.49] | PLCO | 37624040 |
| Sun (2024) | Renal cell carcinoma | 6,193 | Sweden | 1.45 | [1.40; 1.50] | ODDS | 39253735 |
| **Random effects model** | | | | **1.30** | **[1.26; 1.33]** | | |

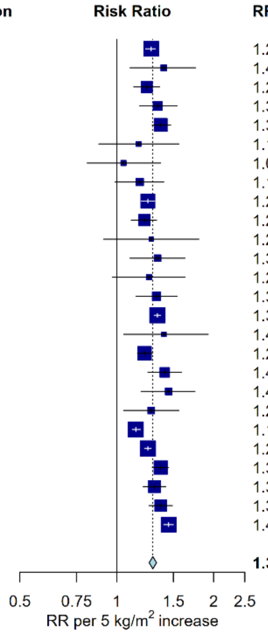

Heterogeneity: $I^2$ = 83.0%, $\tau^2$ = 0.0021, $p$ < 0.0001
Median RR (25th, 75th percentile) = 1.29 (1.24, 1.37)

#### Supplementary Figure 4: Meta-analysis of prospective studies on the risk of kidney cancer in relation to BMI

Study-specific RRs are represented by squares (with their 95% CIs as lines). RRs were combined using weighted averages of the log RRs in the separate studies, with weights assigned based on within and between study variance, yielding a pooled risk estimate and its 95% CI (diamond). Heterogeneity across studies was assessed using $I^2$, $tau^2$, and the 25th and 75th percentile of the RRs. Further details of model adjustments, follow-up time, analytic population for each study are available from **Supplementary Data**.

Abbreviations: BMI=body mass index, CI=confidence interval, PMID=PubMed ID, RR=risk ratio

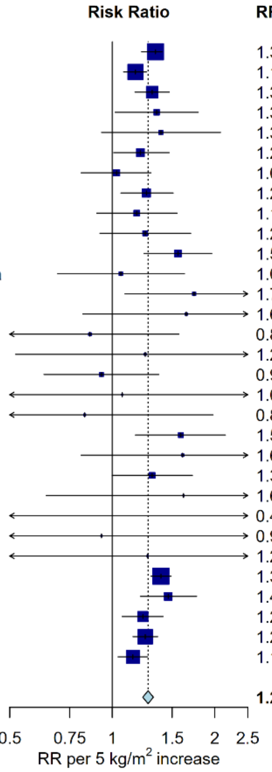

| Author | Site | N cases | Country/Region | RR | [95% CI] | Cohort | PMID |
|---|---|---|---|---|---|---|---|
| Engeland (2005) | Gallbladder | 1,715 | Norway | 1.34 | [1.22; 1.40] | NBHPC | 16132807 |
| Sun (2008) | Gallbladder | 4,400 | S. Korea | 1.17 | [1.08; 1.26] | KNHIS | 18651571 |
| Bhaskaran (2014) | Gallbladder | 303 | UK | 1.31 | [1.17; 1.47] | CPRD | 25129328 |
| Song (2014) | Gallbladder | 77 | Finland | 1.35 | [1.02; 1.79] | FINRISK | 24997743 |
| Campbell (2017) | Gallbladder | 21 | USA | 1.39 | [0.93; 2.08] | CTS | 28314823 |
| Jackson (2019) | Gallbladder | 210 | USA | 1.21 | [1.01; 1.47] | NIH-AARP | 31113819 |
| Jackson (2019) | Gallbladder | 109 | USA | 1.03 | [0.81; 1.30] | MEC | 31113819 |
| Jackson (2019) | Gallbladder | 87 | USA | 1.26 | [1.06; 1.51] | WHI | 31113819 |
| Jackson (2019) | Gallbladder | 69 | USA | 1.18 | [0.90; 1.55] | CPS-II | 31113819 |
| Jackson (2019) | Gallbladder | 45 | USA | 1.25 | [0.92; 1.70] | PLCO | 31113819 |
| Jackson (2019) | Gallbladder | 52 | USA | 1.56 | [1.24; 1.96] | NHS | 31113819 |
| Jackson (2019) | Gallbladder | 15 | USA and Canada | 1.06 | [0.69; 1.63] | AHS-II | 31113819 |
| Jackson (2019) | Gallbladder | 16 | USA | 1.74 | [1.09; 2.78] | VITAL | 31113819 |
| Jackson (2019) | Gallbladder | 18 | USA | 1.65 | [0.82; 3.31] | AgHealth | 31113819 |
| Jackson (2019) | Gallbladder | 29 | Singapore | 0.86 | [0.47; 1.57] | SCHS | 31113819 |
| Jackson (2019) | Gallbladder | 11 | USA | 1.25 | [0.52; 3.00] | HPFS | 31113819 |
| Jackson (2019) | Gallbladder | 140 | Japan | 0.93 | [0.63; 1.37] | RERF | 31113819 |
| Jackson (2019) | Gallbladder | 4 | USA | 1.07 | [0.36; 3.17] | SISTER | 31113819 |
| Jackson (2019) | Gallbladder | 8 | USA | 0.83 | [0.35; 1.98] | BCDDP | 31113819 |
| Jackson (2019) | Gallbladder | 35 | Australia | 1.59 | [1.17; 2.15] | MCCS | 31113819 |
| Jackson (2019) | Gallbladder | 10 | USA | 1.61 | [0.81; 3.22] | WHS | 31113819 |
| Jackson (2019) | Gallbladder | 69 | USA | 1.31 | [1.00; 1.72] | IWHS | 31113819 |
| Jackson (2019) | Gallbladder | 17 | Finland | 1.62 | [0.64; 4.12] | ATBC | 31113819 |
| Jackson (2019) | Gallbladder | 7 | USA | 0.48 | [0.04; 5.13] | PHS | 31113819 |
| Jackson (2019) | Gallbladder | 7 | Taiwan | 0.93 | [0.30; 2.87] | REVEAL | 31113819 |
| Jackson (2019) | Gallbladder | 6 | USA | 1.27 | [0.26; 6.15] | NYUWHS | 31113819 |
| Nordsletten (2023) | Gallbladder | 1,099 | Norway | 1.39 | [1.30; 1.49] | NTSP | 37713274 |
| Rask-Andersen (2023) | Gallbladder | 124 | UK | 1.46 | [1.21; 1.77] | UK Biobank | 37311415 |
| Sedlmeier (2023) | Gallbladder | 299 | Europe | 1.23 | [1.07; 1.41] | EPIC | 36460776 |
| Sun (2024) | Gallbladder | 917 | Sweden | 1.25 | [1.15; 1.36] | ODDS | 39253735 |
| Oze (2024) | Gallbladder | 1,109 | Asia | 1.15 | [1.04; 1.27] | ACC | 37966009 |
| **Random effects model** | | | | **1.27** | **[1.23; 1.32]** | | |

Heterogeneity: $I^2$ = 22.2%, $\tau^2$ = 0.0012, $p$ = 0.1363
Median RR (25th, 75th percentile) = 1.25 (1.11, 1.39)

#### Supplementary Figure 5: Meta-analysis of prospective studies on the risk of gallbladder cancer in relation to BMI

Study-specific RRs are represented by squares (with their 95% CIs as lines). RRs were combined using weighted averages of the log RRs in the separate studies, with weights assigned based on within and between study variance, yielding a pooled risk estimate and its 95% CI (diamond). Heterogeneity across studies was assessed using $I^2$, $tau^2$, and the $25^{th}$ and $75^{th}$ percentile of the RRs. Further details of model adjustments, follow-up time, analytic population for each study are available from **Supplementary Data**.

Abbreviations: BMI=body mass index, CI=confidence interval, PMID=PubMed ID, RR=risk ratio

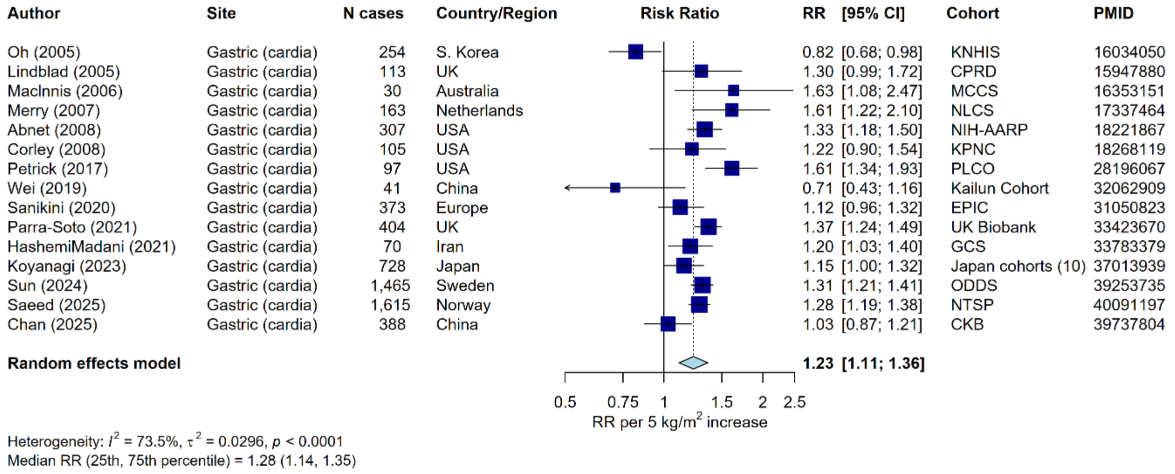

#### Supplementary Figure 6: Meta-analysis of prospective studies on the risk of gastric cancer (cardia) in relation to BMI

Study-specific RRs are represented by squares (with their 95% CIs as lines). RRs were combined using weighted averages of the log RRs in the separate studies, with weights assigned based on within and between study variance, yielding a pooled risk estimate and its 95% CI (diamond). Heterogeneity across studies was assessed using $I^2$, $tau^2$, and the $25^{th}$ and $75^{th}$ percentile of the RRs. Further details of model adjustments, follow-up time, analytic population for each study are available from **Supplementary Data**.

Abbreviations: BMI=body mass index, CI=confidence interval, PMID=PubMed ID, RR=risk ratio

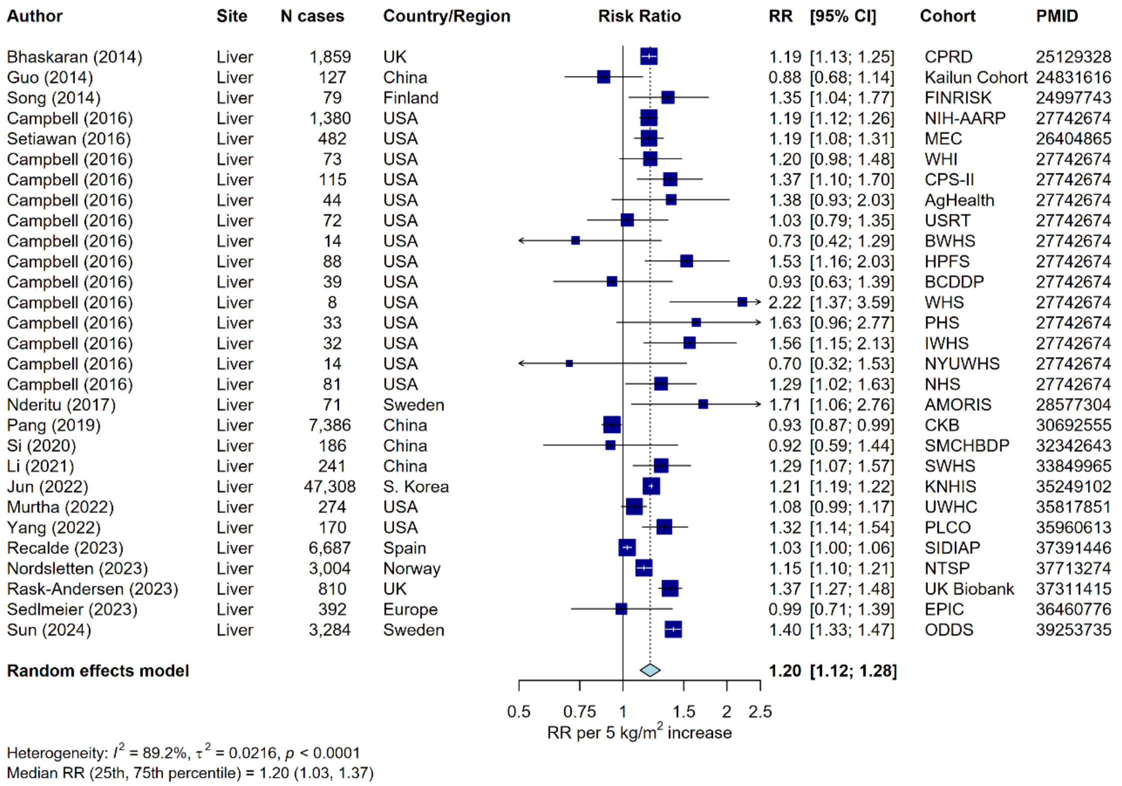

#### Supplementary Figure 7: Meta-analysis of prospective studies on the risk of liver cancer in relation to BMI

Study-specific RRs are represented by squares (with their 95% CIs as lines). RRs were combined using weighted averages of the log RRs in the separate studies, with weights assigned based on within and between study variance, yielding a pooled risk estimate and its 95% CI (diamond). Heterogeneity across studies was assessed using $I^2$, $tau^2$, and the $25^{th}$ and $75^{th}$ percentile of the RRs. Further details of model adjustments, follow-up time, analytic population for each study are available from **Supplementary Data**.

Abbreviations: BMI=body mass index, CI=confidence interval, PMID=PubMed ID, RR=risk ratio

| Author | Site | N cases | Country/Region | RR [95% CI] | Cohort | PMID |
|---|---|---|---|---|---|---|
| Vatten (1990) | Breast (postmenopausal) | 99 | Norway | 0.90 [0.68; 1.18] | NCS | 2307535 |
| van den Brandt (2000) | Breast (postmenopausal) | 420 | Netherlands | 0.99 [0.83; 1.17] | NLCS | 10997541 |
| van den Brandt (2000) | Breast (postmenopausal) | 242 | Canada | 1.14 [0.89; 1.47] | CNBSS | 10997541 |
| van den Brandt (2000) | Breast (postmenopausal) | 643 | USA | 1.14 [1.05; 1.24] | IWHS | 10997541 |
| van den Brandt (2000) | Breast (postmenopausal) | 358 | USA | 1.05 [0.94; 1.19] | NYSC | 10997541 |
| van den Brandt (2000) | Breast (postmenopausal) | 87 | USA | 1.23 [0.96; 1.58] | AHS-I | 10997541 |
| Feigelson (2004) | Breast (postmenopausal) | 1,934 | USA | 1.10 [1.05; 1.15] | CPS-II | 14973094 |
| Tehard (2006) | Breast (postmenopausal) | 1,037 | France | 1.15 [1.02; 1.30] | E3N | 16231021 |
| Reinier (2007) | Breast (postmenopausal) | 572 | USA | 1.38 [1.23; 1.53] | VMC | 17063272 |
| Lacey (2009) | Breast (postmenopausal) | 2,063 | USA | 1.06 [1.02; 1.10] | PLCO | 19292893 |
| Andreotti (2010) | Breast (postmenopausal) | 464 | USA | 1.16 [1.05; 1.34] | AgHealth | 20730623 |
| Sczaniecka (2012) | Breast (postmenopausal) | 741 | USA | 1.06 [0.98; 1.16] | VITAL | 23137008 |
| Bhaskaran (2014) | Breast (postmenopausal) | 28,409 | UK | 1.05 [1.03; 1.07] | CPRD | 25129328 |
| Guo (2014) | Breast (postmenopausal) | 57 | China | 1.32 [0.95; 1.83] | Kailun Cohort | 24831616 |
| Catsburg (2014) | Breast (postmenopausal) | 541 | Canada | 1.13 [1.00; 1.29] | CSDLH | 24781974 |
| White (2015) | Breast (postmenopausal) | 1,582 | USA | 1.10 [1.05; 1.16] | SISTER | 26193782 |
| Heo (2015) | Breast (postmenopausal) | 6,798 | USA | 1.10 [1.09; 1.13] | WHI | 25587642 |
| Chen (2016) | Breast (postmenopausal) | 6,023 | Taiwan | 1.21 [1.17; 1.25] | TBCSP | 26443343 |
| Freisling (2017) | Breast (postmenopausal) | 125 | Germany | 1.19 [0.99; 1.44] | ESTHER | 28441380 |
| Freisling (2017) | Breast (postmenopausal) | 22 | Greece | 1.15 [0.73; 1.82] | EPIC (Greece) | 28441380 |
| Freisling (2017) | Breast (postmenopausal) | 64 | Norway | 1.32 [0.96; 1.84] | Tromso | 28441380 |
| Cifu (2018) | Breast (postmenopausal) | 7,095 | USA | 1.08 [1.05; 1.11] | NIH-AARP | 30309689 |
| daSilva (2018) | Breast (postmenopausal) | 1,336 | Norway | 1.07 [1.00; 1.15] | NOWAC | 30202086 |
| Sandvei (2019) | Breast (postmenopausal) | 2,080 | Norway | 1.10 [1.10; 1.16] | NBCSP | 30083811 |
| Bissell (2020) | Breast (postmenopausal) | 14,357 | USA | 1.17 [1.16; 1.18] | BCSC | 32727722 |
| Renehan (2020) | Breast (postmenopausal) | 829 | UK | 1.24 [1.15; 1.34] | PROCAS | 32203222 |
| Wang (2020) | Breast (postmenopausal) | 1,202 | China | 1.29 [1.18; 1.40] | CKB | 31115907 |
| Houghton (2021) | Breast (postmenopausal) | 2,909 | USA | 1.19 [1.12; 1.26] | NHS, NHS-II | 33367714 |
| Yang (2021) | Breast (postmenopausal) | 789 | Australia | 1.16 [1.09; 1.24] | MCCS | 33268487 |
| Park (2021) | Breast (postmenopausal) | 39,485 | S. Korea | 1.23 [1.21; 1.25] | KNHIS | 33010023 |
| Yang (2022) | Breast (postmenopausal) | 15,506 | UK | 1.20 [1.18; 1.22] | MWS | 35255844 |
| Recalde (2023) | Breast (postmenopausal) | 28,178 | Spain | 1.08 [1.06; 1.09] | SIDIAP | 37391446 |
| Gram (2023) | Breast (postmenopausal) | 1,919 | Denmark | 1.10 [1.04; 1.15] | DCHS | 36750141 |
| Sedlmeier (2023) | Breast (postmenopausal) | 4,988 | Europe | 1.21 [1.12; 1.31] | EPIC (not Greece) | 36460776 |
| Wada (2024) | Breast (postmenopausal) | 1,685 | Japan | 1.33 [1.24; 1.42] | ACC (Japan) | 39543702 |
| Wada (2024) | Breast (postmenopausal) | 759 | China | 1.33 [1.21; 1.27] | SWHS | 39543702 |
| Wada (2024) | Breast (postmenopausal) | 173 | S. Korea | 1.05 [0.82; 1.34] | ACC (Korea) | 39543702 |
| Sun (2024) | Breast (postmenopausal) | 19,296 | Sweden | 1.06 [1.04; 1.08] | ODDS | 39253735 |
| Lu (2024) | Breast (postmenopausal) | 6,956 | UK | 1.12 [1.09; 1.15] | UK Biobank | 39150790 |
| Ogbenna (2025) | Breast (postmenopausal) | 4,555 | USA | 1.13 [1.09; 1.17] | MEC | 40112020 |
| **Random effects model** | | | | **1.15 [1.12; 1.17]** | | |

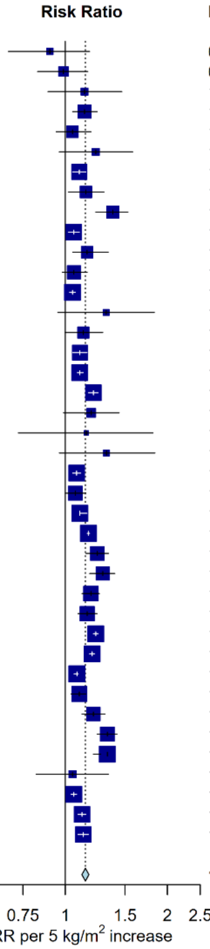

Heterogeneity: $I^2$ = 93.7%, $\tau^2$ = 0.0040, $p$ < 0.0001
Median RR (25th, 75th percentile) = 1.14 (1.08, 1.21)

#### Supplementary Figure 8: Meta-analysis of prospective studies on the risk of breast (postmenopausal) cancer in relation to BMI

Study-specific RRs are represented by squares (with their 95% CIs as lines). RRs were combined using weighted averages of the log RRs in the separate studies, with weights assigned based on within and between study variance, yielding a pooled risk estimate and its 95% CI (diamond). Heterogeneity across studies was assessed using $I^2$, tau$^2$, and the 25$^{th}$ and 75$^{th}$ percentile of the RRs. Further details of model adjustments, follow-up time, analytic population for each study are available from **Supplementary Data**.

Abbreviations: BMI=body mass index, CI=confidence interval, PMID=PubMed ID, RR=risk ratio

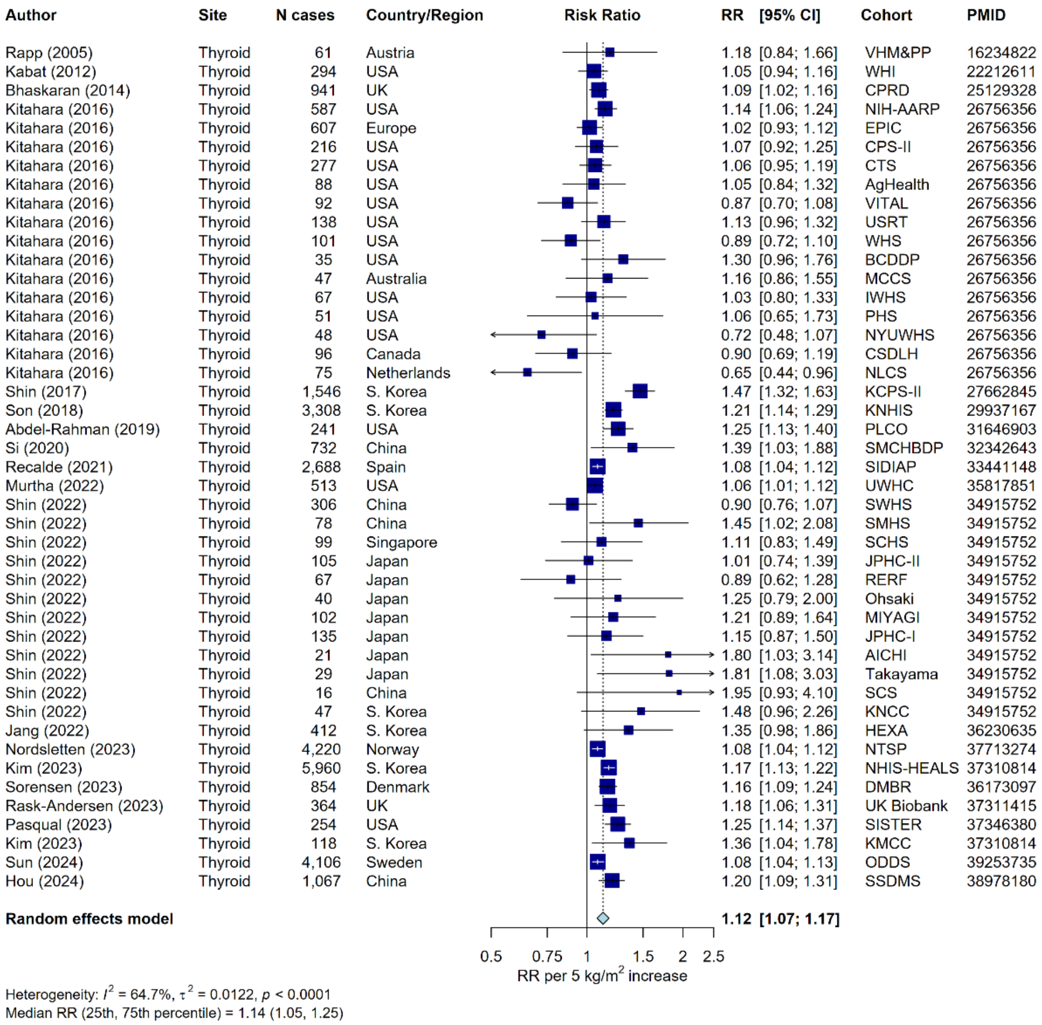

#### Supplementary Figure 9: Meta-analysis of prospective studies on the risk of thyroid cancer in relation to BMI

Study-specific RRs are represented by squares (with their 95% CIs as lines). RRs were combined using weighted averages of the log RRs in the separate studies, with weights assigned based on within and between study variance, yielding a pooled risk estimate and its 95% CI (diamond). Heterogeneity across studies was assessed using $I^2$, $tau^2$, and the 25$^{th}$ and 75$^{th}$ percentile of the RRs. Further details of model adjustments, follow-up time, analytic population for each study are available from **Supplementary Data**.

Abbreviations: BMI=body mass index, CI=confidence interval, PMID=PubMed ID, RR=risk ratio

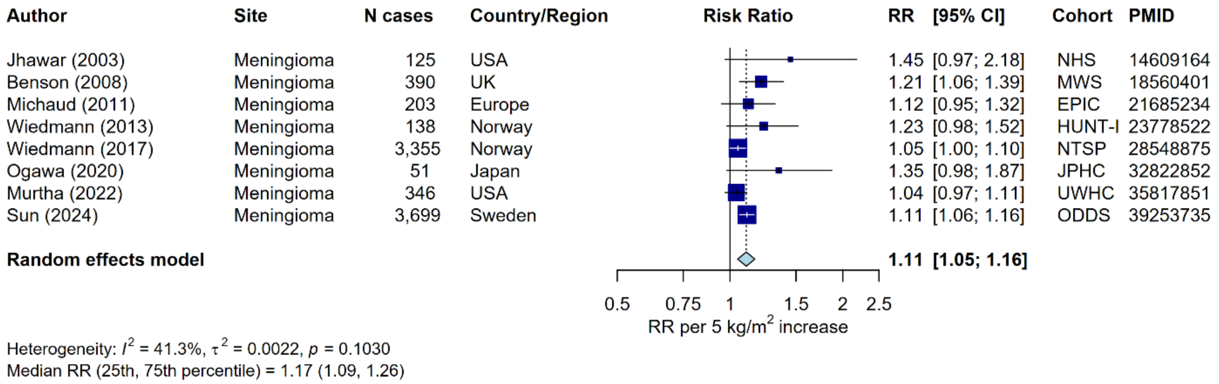

#### Supplementary Figure 10: Meta-analysis of prospective studies on the risk of meningioma in relation to BMI

Study-specific RRs are represented by squares (with their 95% CIs as lines). RRs were combined using weighted averages of the log RRs in the separate studies, with weights assigned based on within and between study variance, yielding a pooled risk estimate and its 95% CI (diamond). Heterogeneity across studies was assessed using $I^2$, $tau^2$, and the $25^{th}$ and $75^{th}$ percentile of the RRs. Further details of model adjustments, follow-up time, analytic population for each study are available from **Supplementary Data**.

Abbreviations: BMI=body mass index, CI=confidence interval, PMID=PubMed ID, RR=risk ratio

| Author | Site | N cases | Country/Region | RR [95% CI] | Cohort | PMID |
|---|---|---|---|---|---|---|
| Nilsen (2001) | Colorectal | 712 | Norway | 1.02 [0.90; 1.16] | HUNT-I | 11161410 |
| Terry (2002) | Colorectal | 527 | Canada | 1.01 [0.98; 1.04] | CNBSS | 12117878 |
| Engeland (2005) | Colorectal | 24,130 | Norway | 1.05 [1.04; 1.07] | NBHPC | 16132807 |
| Andreotti (2010) | Colorectal | 479 | USA | 1.18 [1.06; 1.32] | AgHealth | 20730623 |
| Hughes (2011) | Colorectal | 1,211 | Netherlands | 1.06 [0.96; 1.16] | NLCS | 21984660 |
| Renehan (2012) | Colorectal | 4,076 | USA | 1.10 [1.06; 1.14] | NIH-AARP | 23186750 |
| Bhaskaran (2014) | Colorectal | 19,588 | UK | 1.08 [1.06; 1.10] | CPRD | 25129328 |
| Song (2014) | Colorectal | 609 | Finland | 1.08 [0.97; 1.20] | FINRISK | 24997743 |
| Harding (2015) | Colorectal | 1,918 | Australia | 1.12 [1.06; 1.18] | ANZDCC | 25810218 |
| Heo (2015) | Colorectal | 1,904 | USA | 1.10 [1.05; 1.14] | WHI | 25587642 |
| Lu (2016) | Colorectal | 2,044 | Norway | 1.08 [1.01; 1.16] | CONOR | 26808598 |
| Buron Pust (2017) | Colorectal | 18,518 | UK | 1.08 [1.05; 1.10] | MWS | 27859268 |
| Hanyuda (2017) | Colorectal | 1,054 | USA | 1.08 [1.00; 1.15] | NHS | 31276608 |
| Hanyuda (2017) | Colorectal | 963 | USA | 1.33 [1.22; 1.47] | HPFS | 31276608 |
| Freisling (2017) | Colorectal | 111 | Germany | 1.24 [0.96; 1.64] | ESTHER | 28441380 |
| Freisling (2017) | Colorectal | 41 | UK | 1.92 [1.14; 3.24] | PRIME Belfast | 28441380 |
| Pang (2018) | Colorectal | 3,461 | China | 1.17 [1.11; 1.23] | CKB | 29872150 |
| Nunez (2018) | Colorectal | 1,204 | Australia | 1.14 [1.06; 1.24] | 45 and Up | 29510753 |
| Wei (2018) | Colorectal | 318 | China | 1.26 [0.98; 1.62] | Kailun Cohort | 29996293 |
| daSilva (2018) | Colorectal | 1,805 | Norway | 1.04 [0.98; 1.11] | NOWAC | 30202086 |
| Si (2020) | Colorectal | 219 | China | 1.08 [0.73; 1.60] | SMCHBDP | 32342643 |
| Kim (2021) | Colorectal | 120,579 | S. Korea | 1.08 [1.07; 1.09] | KNHIS | 32542773 |
| HashemiMadani (2021) | Colorectal | 136 | Iran | 1.10 [0.92; 1.32] | GCS | 33783379 |
| Maliniak (2021) | Colon | 964 | USA | 1.13 [1.03; 1.25] | CPS-II | 33185805 |
| Gepner (2021) | Colon | 328 | Israel | 1.26 [1.07; 1.47] | IIHD | 34639717 |
| Murtha (2022) | Colorectal | 556 | USA | 1.05 [1.00; 1.12] | UWHC | 35817851 |
| Recalde (2023) | Colorectal | 37,812 | Spain | 1.04 [1.03; 1.06] | SIDIAP | 37391446 |
| Nordsletten (2023) | Colorectal | 82,636 | Norway | 1.02 [1.01; 1.03] | NTSP | 37713274 |
| Sedlmeier (2023) | Colorectal | 5,225 | Europe | 1.14 [1.10; 1.18] | EPIC | 36460776 |
| Loomans-Kropp (2023) | Colorectal | 2,803 | USA | 1.16 [1.10; 1.22] | PLCO | 37163267 |
| Sun (2024) | Colorectal | 28,712 | Sweden | 1.13 [1.11; 1.15] | ODDS | 39253735 |
| Paragomi (2024) | Colorectal | 7,691 | Japan | 1.13 [1.08; 1.19] | ACC (Japan) | 39196559 |
| Paragomi (2024) | Colorectal | 3,774 | China | 1.10 [1.04; 1.17] | ACC (China) | 39196559 |
| Paragomi (2024) | Colorectal | 308 | S. Korea | 1.26 [1.00; 1.58] | ACC (Korea) | 39196559 |
| Safizadeh (2025) | Colorectal | 5,977 | UK | 1.10 [1.07; 1.14] | UK Biobank | 39562688 |
| **Random effects model** | | | | **1.10 [1.08; 1.12]** | | |

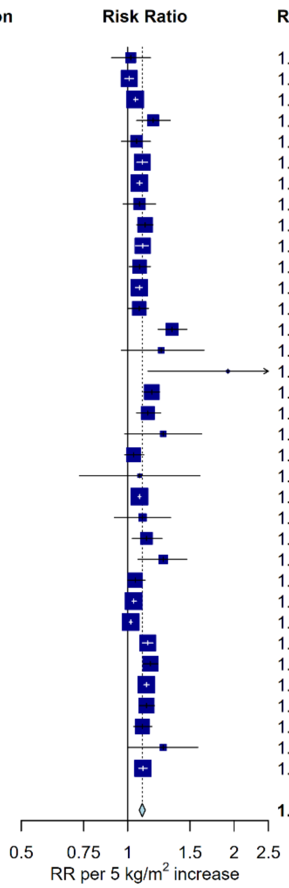

Heterogeneity: $I^2 = 87.2\%$, $\tau^2 = 0.0022$, $p < 0.0001$

Median RR (25th, 75th percentile) = 1.10 (1.08, 1.15)

#### Figure 11: Meta-analysis of prospective studies on the risk of colorectal cancer in relation to BMI

Study-specific RRs are represented by squares (with their 95% CIs as lines). RRs were combined using weighted averages of the log RRs in the separate studies, with weights assigned based on within and between study variance, yielding a pooled risk estimate and its 95% CI (diamond). Heterogeneity across studies was assessed using $I^2$, $tau^2$, and the 25th and 75th percentile of the RRs. Further details of model adjustments, follow-up time, analytic population for each study are available from **Supplementary Data**.

Abbreviations: BMI=body mass index, CI=confidence interval, PMID=PubMed ID, RR=risk ratio

| Author | Site | N cases | Country/Region | RR | [95% CI] | Cohort | PMID |
|---|---|---|---|---|---|---|---|
| Friedman (1994) | Multiple myeloma | 66 | USA | 1.09 | [0.89; 1.33] | KPNC | 7999970 |
| Oh (2005) | Multiple myeloma | 200 | S. Korea | 1.24 | [0.77; 2.02] | KNHIS | 16034050 |
| Engeland (2007) | Multiple myeloma + plasmacytoma | 6,115 | Norway | 1.12 | [1.09; 1.15] | NBHPC | 17041129 |
| Britton (2008) | Multiple myeloma | 268 | Europe | 1.09 | [0.91; 1.31] | EPIC | 18835833 |
| Pylypchuk (2009) | Multiple myeloma | 279 | Netherlands | 1.17 | [0.96; 1.40] | NLCS | 19478235 |
| Andreotti (2010) | Multiple myeloma | 68 | USA | 1.00 | [0.72; 1.40] | AgHealth | 20730623 |
| Murphy (2013) | Multiple myeloma | 1,503 | UK | 1.08 | [1.01; 1.15] | MWS | 23640394 |
| Hofmann (2013) | Multiple myeloma | 813 | USA | 1.08 | [1.00; 1.16] | NIH-AARP | 23543160 |
| Bhaskaran (2014) | Multiple myeloma | 2,969 | UK | 1.03 | [0.98; 1.09] | CPRD | 25129328 |
| Heo (2015) | Multiple myeloma | 282 | USA | 1.00 | [0.90; 1.12] | WHI | 25587642 |
| Parra-Soto (2021) | Multiple myeloma | 763 | UK | 1.10 | [1.02; 1.19] | UK Biobank | 33423670 |
| Bertrand (2022) | Multiple myeloma | 563 | USA | 1.13 | [1.02; 1.26] | CPS-II | 35840735 |
| Bertrand (2022) | Multiple myeloma | 330 | USA | 1.09 | [0.97; 1.23] | NHS | 35840735 |
| Bertrand (2022) | Multiple myeloma | 47 | USA | 1.33 | [1.06; 1.67] | NHS-II | 35840735 |
| Bertrand (2022) | Multiple myeloma | 196 | USA | 0.97 | [0.84; 1.13] | CTS | 35840735 |
| Bertrand (2022) | Multiple myeloma | 182 | USA | 1.12 | [0.89; 1.42] | HPFS | 35840735 |
| Bertrand (2022) | Multiple myeloma | 1,438 | USA | 1.09 | [1.02; 1.17] | KPSC | 35840735 |
| Recalde (2023) | Multiple myeloma | 3,490 | Spain | 1.08 | [1.05; 1.11] | SIDIAP | 37391446 |
| Nordsletten (2023) | Multiple myeloma | 9,338 | Norway | 1.11 | [1.08; 1.14] | NTSP | 37713274 |
| Sun (2024) | Multiple myeloma | 3,720 | Sweden | 1.14 | [1.08; 1.19] | ODDS | 39253735 |
| **Random effects model** | | | | **1.10** | **[1.08; 1.11]** | | |

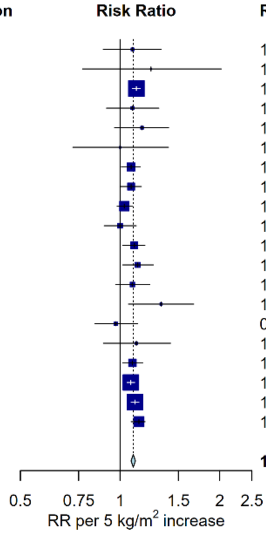

Heterogeneity: $I^2$ = 14.4%, $\tau^2$ = 0.0001, $p$ = 0.2749
Median RR (25th, 75th percentile) = 1.09 (1.08, 1.12)

#### Supplementary Figure 12: Meta-analysis of prospective studies on the risk of multiple myeloma in relation to BMI

Study-specific RRs are represented by squares (with their 95% CIs as lines). RRs were combined using weighted averages of the log RRs in the separate studies, with weights assigned based on within and between study variance, yielding a pooled risk estimate and its 95% CI (diamond). Heterogeneity across studies was assessed using $I^2$, $tau^2$, and the $25^{th}$ and $75^{th}$ percentile of the RRs. Further details of model adjustments, follow-up time, analytic population for each study are available from **Supplementary Data**.

Abbreviations: BMI=body mass index, CI=confidence interval, PMID=PubMed ID, RR=risk ratio

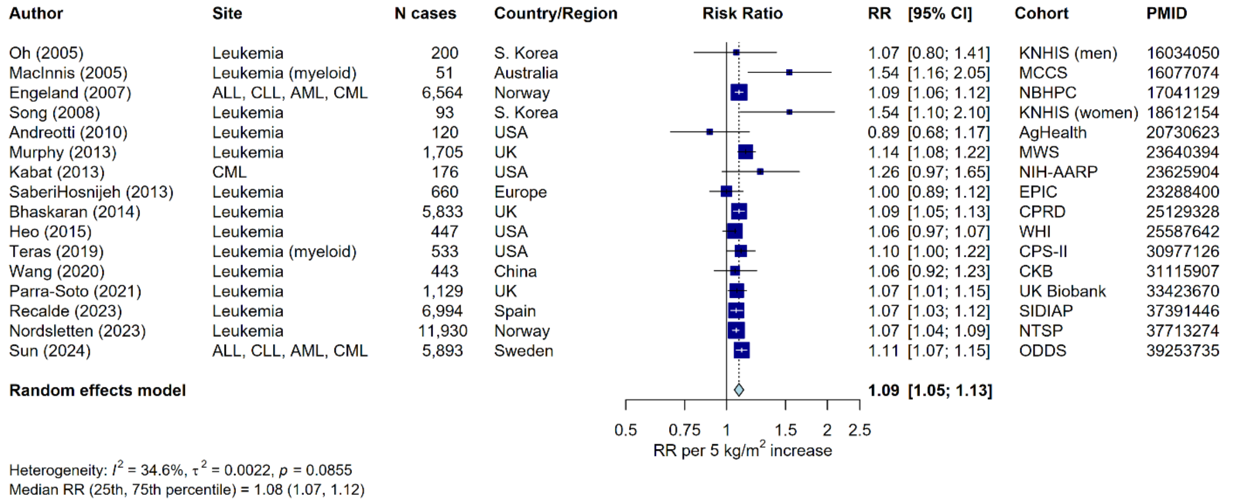

#### Supplementary Figure 13: Meta-analysis of prospective studies on the risk of leukaemia in relation to BMI

Study-specific RRs are represented by squares (with their 95% CIs as lines). RRs were combined using weighted averages of the log RRs in the separate studies, with weights assigned based on within and between study variance, yielding a pooled risk estimate and its 95% CI (diamond). Heterogeneity across studies was assessed using $I^2$, $tau^2$, and the $25^{th}$ and $75^{th}$ percentile of the RRs. Further details of model adjustments, follow-up time, analytic population for each study are available from **Supplementary Data**.

Abbreviations: ALL=acute lymphocytic leukaemia, AML=acute myeloid leukaemia, BMI=body mass index, CI=confidence interval, CLL=chronic lymphocytic leukaemia, CML=chronic myeloid leukaemia, PMID=PubMed ID, RR=risk ratio

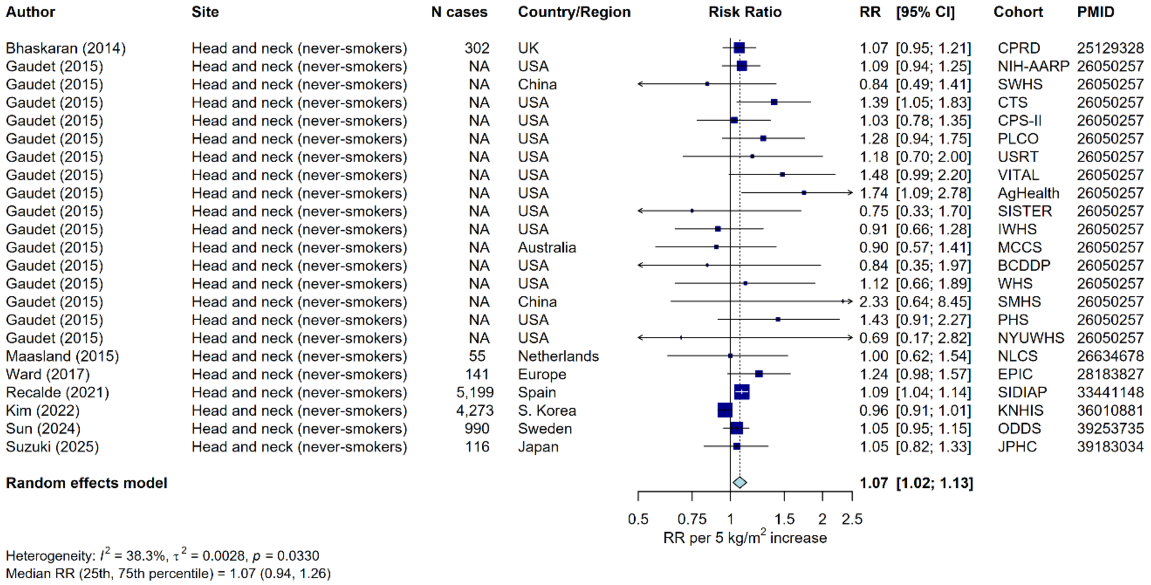

#### Supplementary Figure 14: Meta-analysis of prospective studies on the risk of head and neck cancer (restricted to never-smokers) in relation to BMI

Study-specific RRs are represented by squares (with their 95% CIs as lines). RRs were combined using weighted averages of the log RRs in the separate studies, with weights assigned based on within and between study variance, yielding a pooled risk estimate and its 95% CI (diamond). Heterogeneity across studies was assessed using $I^2$, $tau^2$, and the $25^{th}$ and $75^{th}$ percentile of the RRs. Further details of model adjustments, follow-up time, analytic population for each study are available from **Supplementary Data**. Cases of head and neck cancers for never-smokers were not available in individual studies (N cases total=796).

Abbreviations: BMI=body mass index, CI=confidence interval, PMID=PubMed ID, RR=risk ratio

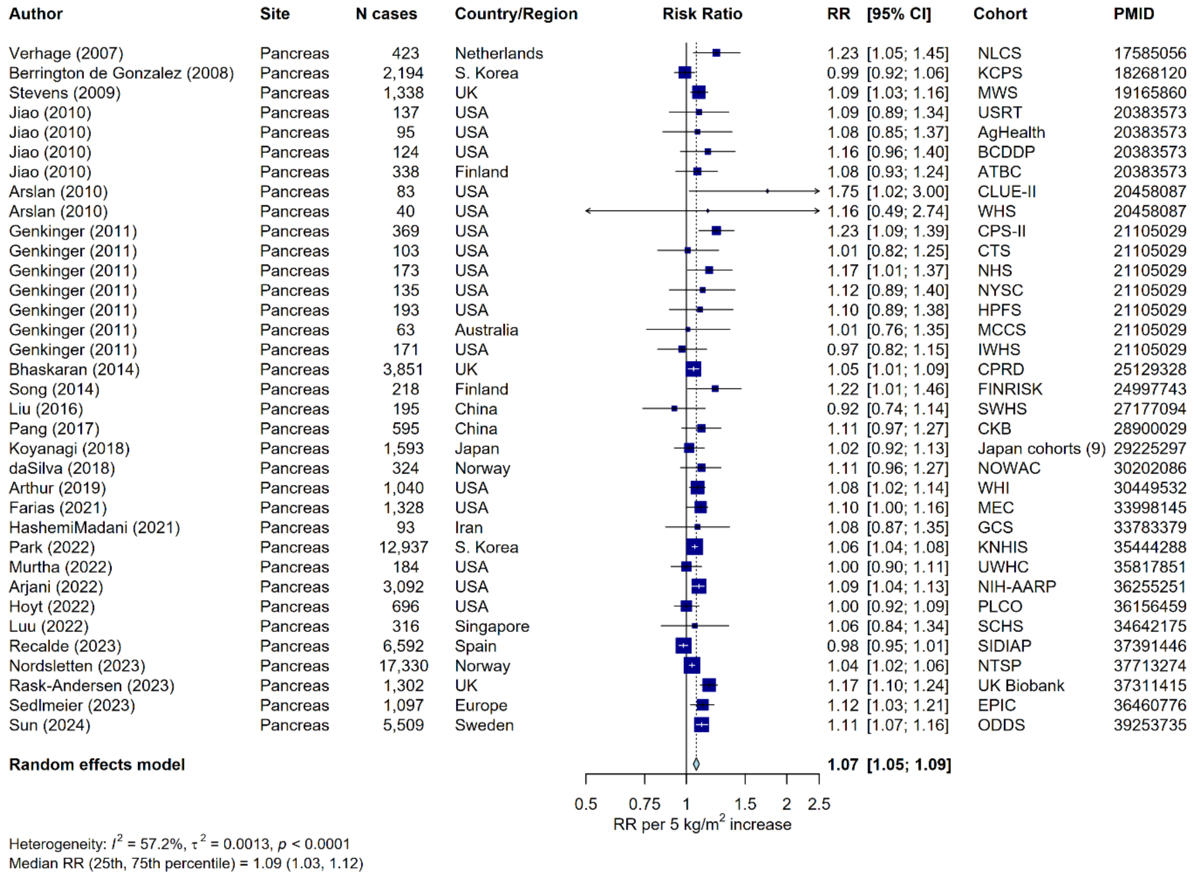

#### Supplementary Figure 15: Meta-analysis of prospective studies on the risk of pancreatic cancer in relation to BMI

Study-specific RRs are represented by squares (with their 95% CIs as lines). RRs were combined using weighted averages of the log RRs in the separate studies, with weights assigned based on within and between study variance, yielding a pooled risk estimate and its 95% CI (diamond). Heterogeneity across studies was assessed using $I^2$, $tau^2$, and the 25$^{th}$ and 75$^{th}$ percentile of the RRs. Further details of model adjustments, follow-up time, analytic population for each study are available from **Supplementary Data**.

Abbreviations: BMI=body mass index, CI=confidence interval, PMID=PubMed ID, RR=risk ratio

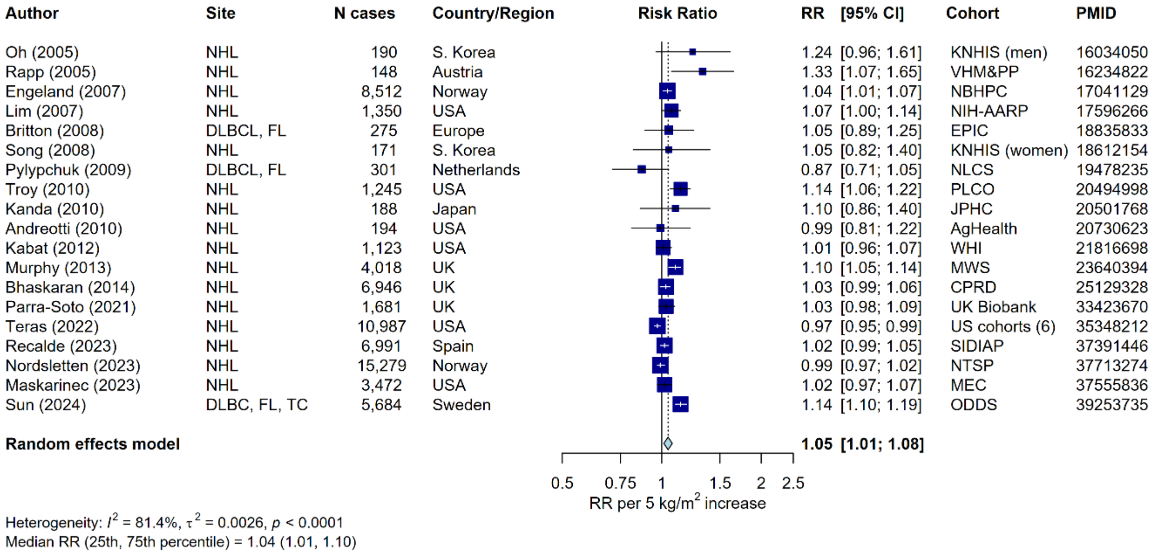

#### Supplementary Figure 16: Meta-analysis of prospective studies on the risk of NHL in relation to BMI

Study-specific RRs are represented by squares (with their 95% CIs as lines). RRs were combined using weighted averages of the log RRs in the separate studies, with weights assigned based on within and between study variance, yielding a pooled risk estimate and its 95% CI (diamond). Heterogeneity across studies was assessed using $I^2$, $tau^2$, and the $25^{th}$ and $75^{th}$ percentile of the RRs. Further details of model adjustments, follow-up time, analytic population for each study are available from **Supplementary Data**.

Abbreviations: BMI=body mass index, CI=confidence interval, DLBCL=diffuse large B-cell lymphoma, FL=follicular lymphoma, NHL=non-Hodgkin lymphoma, PMID=PubMed ID, RR=risk ratio, TC=T-cell lymphoma

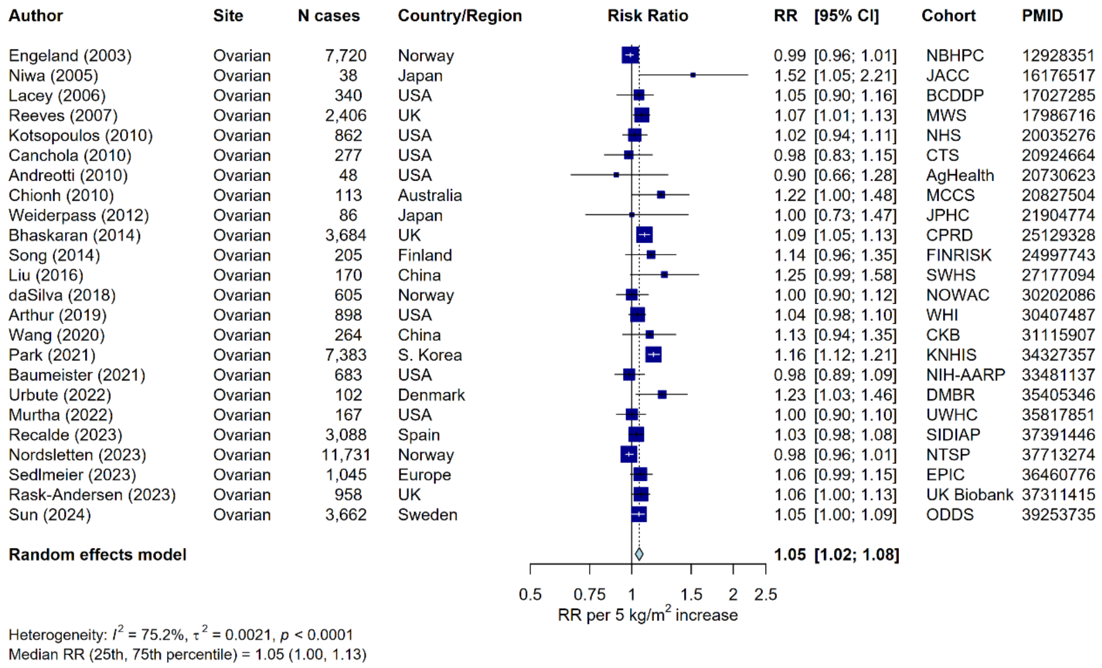

#### Supplementary Figure 17: Meta-analysis of prospective studies on the risk of ovarian cancer in relation to BMI

Study-specific RRs are represented by squares (with their 95% CIs as lines). RRs were combined using weighted averages of the log RRs in the separate studies, with weights assigned based on within and between study variance, yielding a pooled risk estimate and its 95% CI (diamond). Heterogeneity across studies was assessed using $I^2$, $tau^2$, and the $25^{th}$ and $75^{th}$ percentile of the RRs. Further details of model adjustments, follow-up time, analytic population for each study are available from **Supplementary Data**.

Abbreviations: BMI=body mass index, CI=confidence interval, PMID=PubMed ID, RR=risk ratio

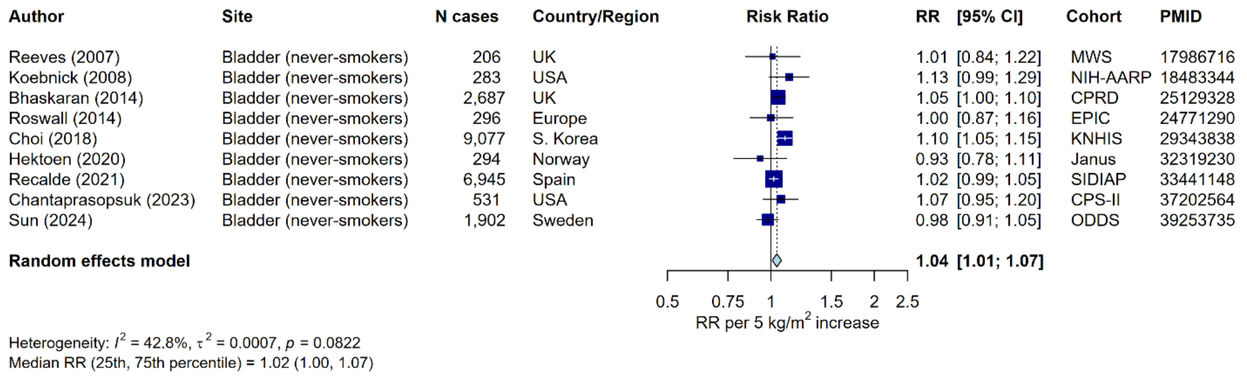

**Supplementary Figure 18: Meta-analysis of prospective studies on the risk of bladder cancer (restricted to never-smokers) in relation to BMI**

Study-specific RRs are represented by squares (with their 95% CIs as lines). RRs were combined using weighted averages of the log RRs in the separate studies, with weights assigned based on within and between study variance, yielding a pooled risk estimate and its 95% CI (diamond). Heterogeneity across studies was assessed using $I^2$, $tau^2$, and the $25^{th}$ and $75^{th}$ percentile of the RRs. Further details of model adjustments, follow-up time, analytic population for each study are available from **Supplementary Data**.

Abbreviations: BMI=body mass index, CI=confidence interval, PMID=PubMed ID, RR=risk ratio

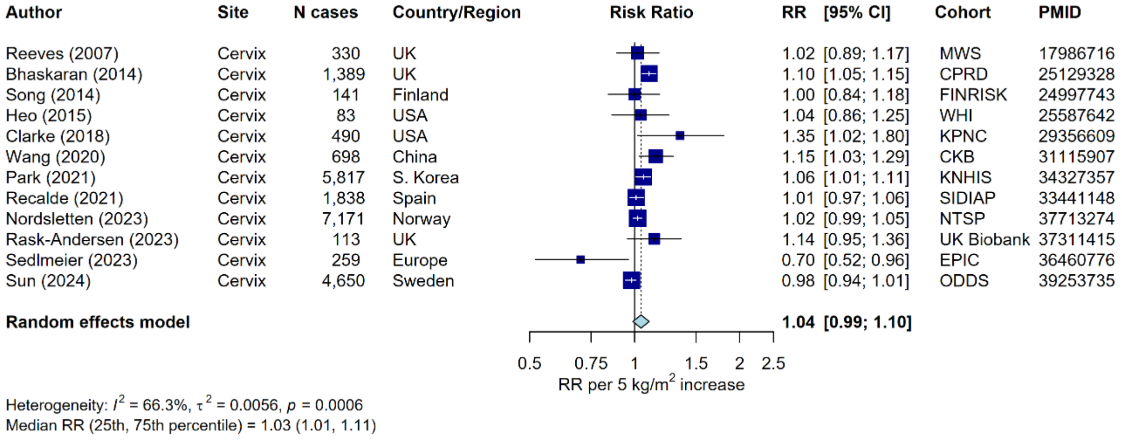

#### Supplementary Figure 19: Meta-analysis of prospective studies on the risk of cervical cancer in relation to BMI

Study-specific RRs are represented by squares (with their 95% CIs as lines). RRs were combined using weighted averages of the log RRs in the separate studies, with weights assigned based on within and between study variance, yielding a pooled risk estimate and its 95% CI (diamond). Heterogeneity across studies was assessed using $I^2$, $tau^2$, and the 25$^{th}$ and 75$^{th}$ percentile of the RRs. Further details of model adjustments, follow-up time, analytic population for each study are available from **Supplementary Data**.

Abbreviations: BMI=body mass index, CI=confidence interval, PMID=PubMed ID, RR=risk ratio

| Author | Site | N cases | Country/Region | Risk Ratio | RR [95% CI] | Cohort | PMID |
|---|---|---|---|---|---|---|---|
| Rodriguez (2001) | Prostate (death) | 3,622 | USA | | 1.10 [1.05; 1.16] | CPS-II | 11319175 |
| Rodriguez (2001) | Prostate (death) | 1,590 | USA | | 1.07 [0.99; 1.16] | CPS-I | 11319175 |
| Littman (2007) | Prostate (aggressive) | 373 | USA | | 1.07 [0.91; 1.26] | VITAL | 17395597 |
| Martin (2009) | Prostate (advanced) | 136 | Norway | | 0.97 [0.75; 1.26] | HUNT-II | 19277881 |
| Bassett (2012) | Prostate (aggressive) | 410 | Australia | | 1.27 [1.08; 1.49] | MCCS | 22213024 |
| Fowke (2015) | Prostate (death) | 33 | India | | 1.13 [0.63; 2.05] | ACC | 26243736 |
| Fowke (2015) | Prostate (death) | 601 | East Asia | | 1.04 [0.92; 1.18] | ACC | 26243736 |
| Kelly (2017) | Prostate (aggressive) | 3,078 | USA | | 1.02 [0.97; 1.07] | PLCO | 27754927 |
| Perez-Cornago (2017) | Prostate (advanced) | 1,384 | Europe | | 1.05 [0.96; 1.15] | EPIC | 28701188 |
| Hendriks (2018) | Prostate (aggressive) | 170 | Netherlands | | 0.93 [0.75; 1.16] | ZODIAC | 29371278 |
| Lee (2019) | Prostate (death) | 289 | S. Korea | | 1.39 [1.11; 1.73] | KNHIS | 30282445 |
| Kelly (2019) | Prostate (aggressive) | 2,185 | USA | | 0.99 [0.94; 1.05] | NIH-AARP | 30376043 |
| Genkinger (2020) | Prostate (death) | 450 | Netherlands | | 1.15 [0.94; 1.41] | NLCS | 31912782 |
| Genkinger (2020) | Prostate (death) | 518 | USA | | 1.09 [0.95; 1.26] | HPFS | 31912782 |
| Genkinger (2020) | Prostate (death) | 269 | Finland | | 1.07 [0.91; 1.26] | ATBC | 31912782 |
| Sattayapiwat (2021) | Prostate (advanced) | 1,236 | USA | | 1.00 [0.92; 1.08] | MEC | 33751036 |
| Perez-Cornago (2022) | Prostate (death) | 659 | UK | | 1.07 [0.97; 1.17] | UK Biobank | 35509091 |
| Sun (2024) | Prostate (aggressive) | 13,493 | Sweden | | 1.01 [0.98; 1.04] | ODDS | 39253735 |
| Pagadala (2024) | Prostate (death) | 3,250 | USA | | 0.86 [0.82; 0.95] | MVP | 38865417 |
| **Random effects model** | | | | | **1.04 [1.00; 1.09]** | | |

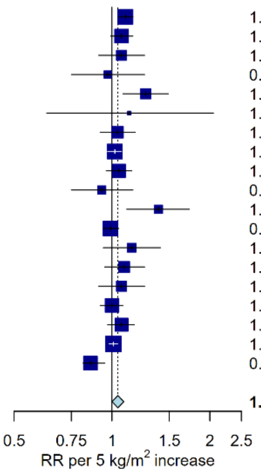

Heterogeneity: $I^2 = 64.6\%$, $\tau^2 = 0.0047$, $p < 0.0001$
Median RR (25th, 75th percentile) = 1.07 (1.00, 1.10)

#### Supplementary Figure 20: Meta-analysis of prospective studies on the risk of prostate cancer (aggressive) in relation to BMI

Aggressive prostate cancer was defined as any of stage 3-4 on the American Joint Committee on Cancer 1992 classification, advanced cancer, advanced or metastatic cancer; metastatic cancer; stage C or D on the Whitmore/Jewett scale; fatal cancer (prostate cancer-specific mortality); high stage or grade; Gleason grade ≥ 7). Study-specific RRs are represented by squares (with their 95% CIs as lines). RRs were combined using weighted averages of the log RRs in the separate studies, with weights assigned based on within and between study variance, yielding a pooled risk estimate and its 95% CI (diamond). Heterogeneity across studies was assessed using $I^2$, $tau^2$, and the 25th and 75th percentile of the RRs. Further details of model adjustments, follow-up time, analytic population for each study are available from **Supplementary Data**.

Abbreviations: BMI=body mass index, CI=confidence interval, PMID=PubMed ID, RR=risk ratio

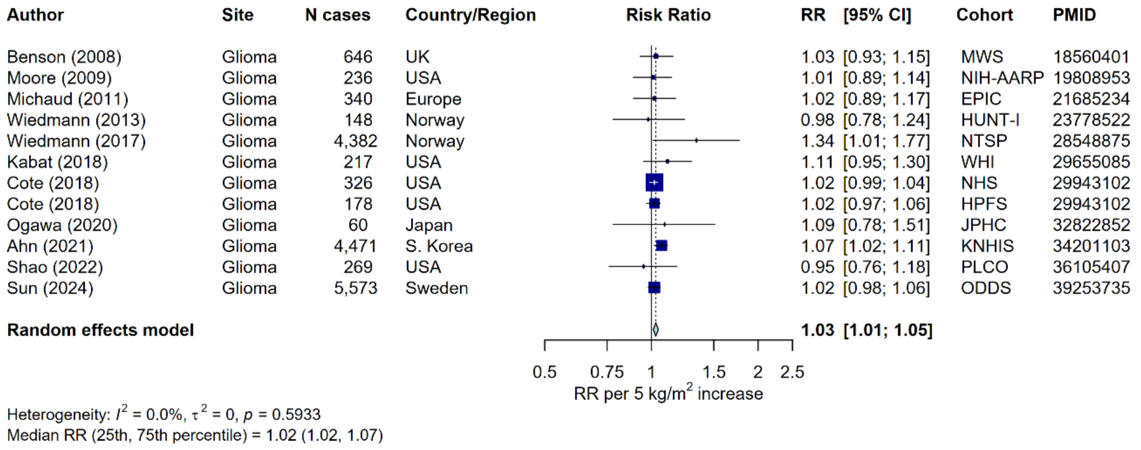

#### Supplementary Figure 21: Meta-analysis of prospective studies on the risk of glioma in relation to BMI

Study-specific RRs are represented by squares (with their 95% CIs as lines). RRs were combined using weighted averages of the log RRs in the separate studies, with weights assigned based on within and between study variance, yielding a pooled risk estimate and its 95% CI (diamond). Heterogeneity across studies was assessed using $I^2$, tau$^2$, and the 25$^{th}$ and 75$^{th}$ percentile of the RRs. Further details of model adjustments, follow-up time, analytic population for each study are available from **Supplementary Data**.

Abbreviations: BMI=body mass index, CI=confidence interval, PMID=PubMed ID, RR=risk ratio

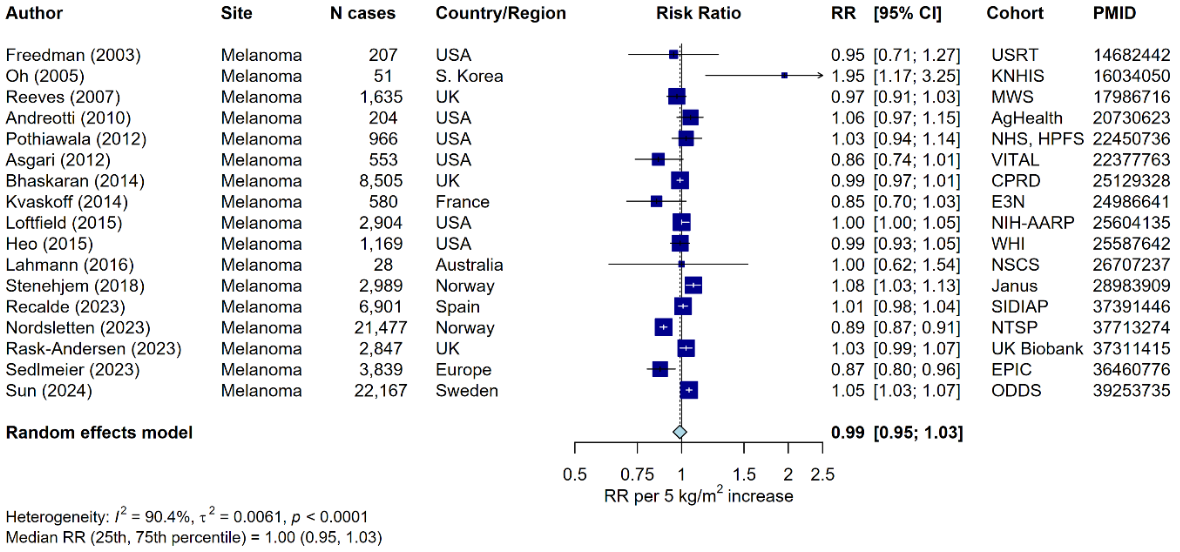

#### Supplementary Figure 22: Meta-analysis of prospective studies on the risk of melanoma in relation to BMI

Study-specific RRs are represented by squares (with their 95% CIs as lines). RRs were combined using weighted averages of the log RRs in the separate studies, with weights assigned based on within and between study variance, yielding a pooled risk estimate and its 95% CI (diamond). Heterogeneity across studies was assessed using $I^2$, $tau^2$, and the 25th and 75th percentile of the RRs. Further details of model adjustments, follow-up time, analytic population for each study are available from **Supplementary Data**.

Abbreviations: BMI=body mass index, CI=confidence interval, PMID=PubMed ID, RR=risk ratio

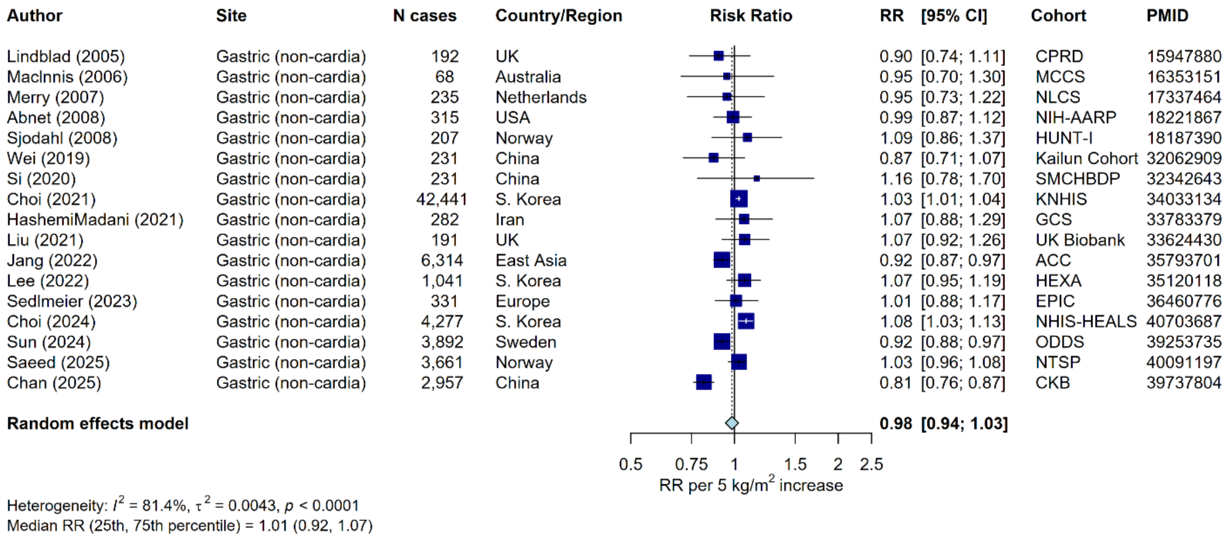

#### Supplementary Figure 23: Meta-analysis of prospective studies on the risk of gastric cancer (non-cardia) in relation to BMI

Study-specific RRs are represented by squares (with their 95% CIs as lines). RRs were combined using weighted averages of the log RRs in the separate studies, with weights assigned based on within and between study variance, yielding a pooled risk estimate and its 95% CI (diamond). Heterogeneity across studies was assessed using $I^2$, $tau^2$, and the 25$^{th}$ and 75$^{th}$ percentile of the RRs. Further details of model adjustments, follow-up time, analytic population for each study are available from **Supplementary Data**.

Abbreviations: BMI=body mass index, CI=confidence interval, PMID=PubMed ID, RR=risk ratio

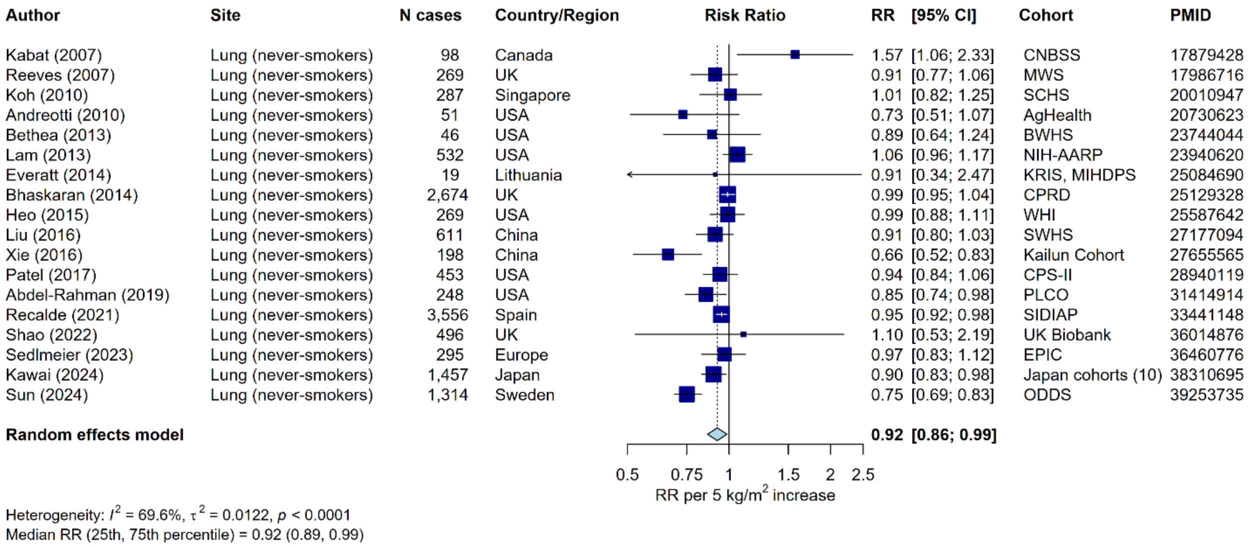

#### Supplementary Figure 24: Meta-analysis of prospective studies on the risk of lung cancer (restricted to never-smokers) in relation to BMI

Study-specific RRs are represented by squares (with their 95% CIs as lines). RRs were combined using weighted averages of the log RRs in the separate studies, with weights assigned based on within and between study variance, yielding a pooled risk estimate and its 95% CI (diamond). Heterogeneity across studies was assessed using $I^2$, $tau^2$, and the 25th and 75th percentile of the RRs. Further details of model adjustments, follow-up time, analytic population for each study are available from **Supplementary Data**.

Abbreviations: BMI=body mass index, CI=confidence interval, PMID=PubMed ID, RR=risk ratio

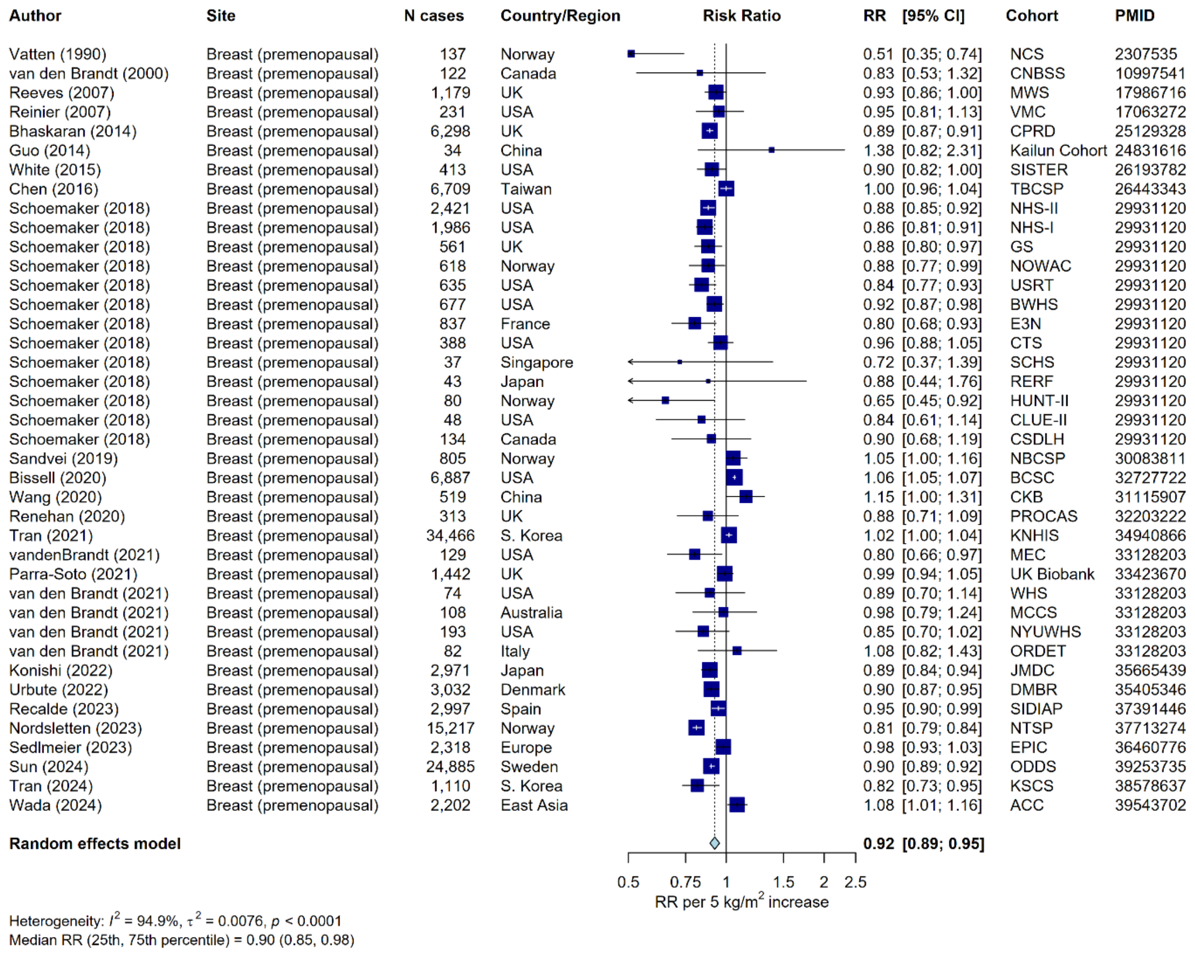

#### Supplementary Figure 25: Meta-analysis of prospective studies on the risk of breast cancer (premenopausal) in relation to BMI

Study-specific RRs are represented by squares (with their 95% CIs as lines). RRs were combined using weighted averages of the log RRs in the separate studies, with weights assigned based on within and between study variance, yielding a pooled risk estimate and its 95% CI (diamond). Heterogeneity across studies was assessed using $I^2$, $tau^2$, and the 25th and 75th percentile of the RRs. Further details of model adjustments, follow-up time, analytic population for each study are available from **Supplementary Data**.

Abbreviations: BMI=body mass index, CI=confidence interval, PMID=PubMed ID, RR=risk ratio

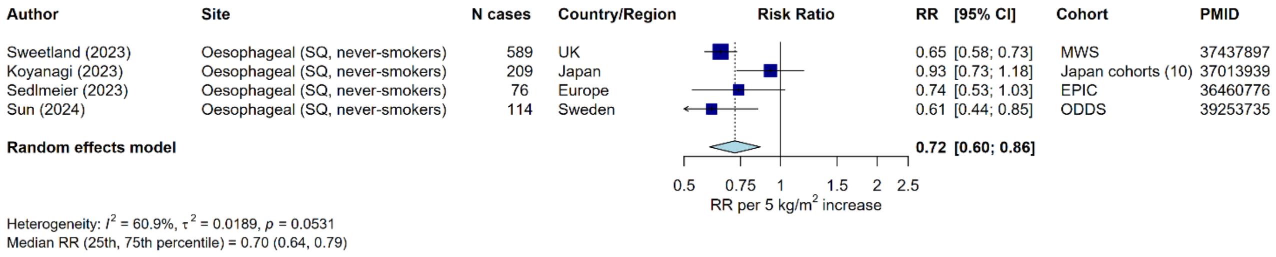

#### Supplementary Figure 26: Meta-analysis of prospective studies on the risk of oesophageal (SQ, restricted to never-smokers) in relation to BMI

Study-specific RRs are represented by squares (with their 95% CIs as lines). RRs were combined using weighted averages of the log RRs in the separate studies, with weights assigned based on within and between study variance, yielding a pooled risk estimate and its 95% CI (diamond). Heterogeneity across studies was assessed using $I^2$, $tau^2$, and the $25^{th}$ and $75^{th}$ percentile of the RRs. Further details of model adjustments, follow-up time, analytic population for each study are available from **Supplementary Data**.

Abbreviations: BMI=body mass index, CI=confidence interval, PMID=PubMed ID, RR=risk ratio, SQ=squamous cell carcinoma.

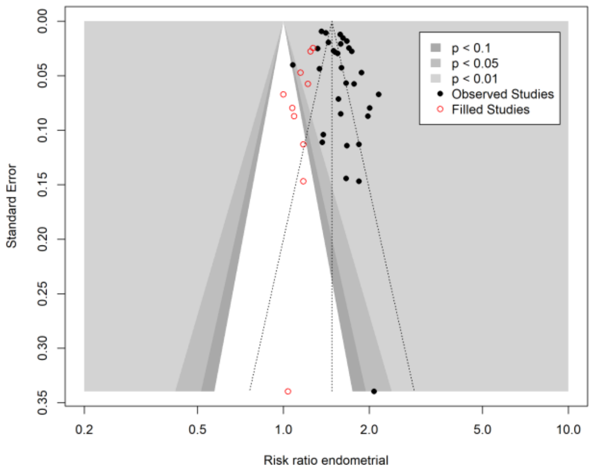

N filled studies: 10
Trim-and-fill RR (95% CI) = 1.48 (1.4 to 1.57)
Meta-analysis RR (95% CI) = 1.58 (1.51 to 1.67)
Difference in RRs = -0.11
Egger's test: Intercept (95% CI) = 2.24 (0.14 to 4.34), p = 0.045

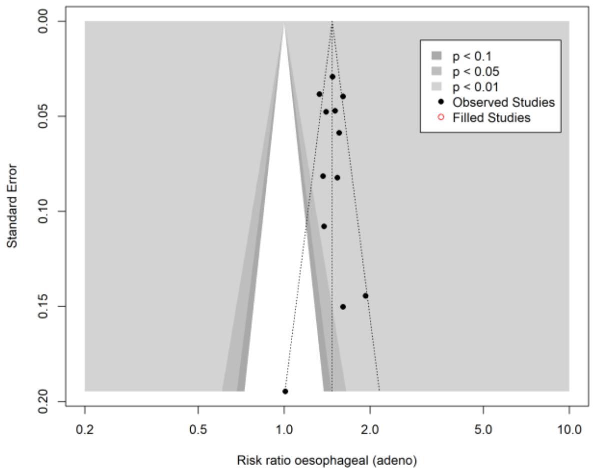

N filled studies: 0
Trim-and-fill RR (95% CI) = 1.47 (1.39 to 1.56)
Meta-analysis RR (95% CI) = 1.47 (1.39 to 1.56)
Difference in OR = N/A
Egger's test: Intercept (95% CI) = -0.01 (-1.85 to 1.82), p = 0.989

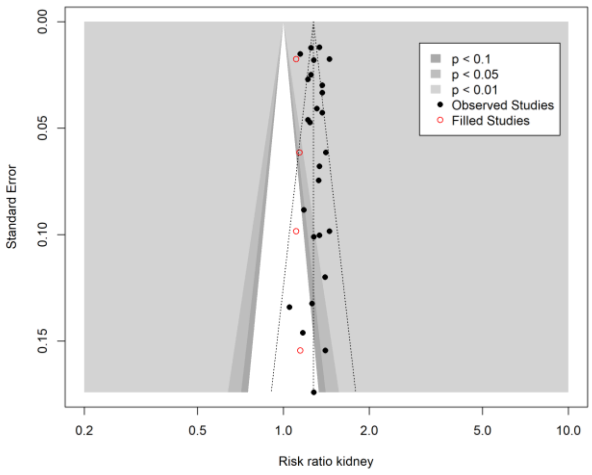

N filled studies: 4
Trim-and-fill RR (95% CI) = 1.27 (1.24 to 1.31)
Meta-analysis RR (95% CI) = 1.3 (1.26 to 1.33)
Difference in RRs = -0.02
Egger's test: Intercept (95% CI) = 0.26 (-1.21 to 1.72), p = 0.732

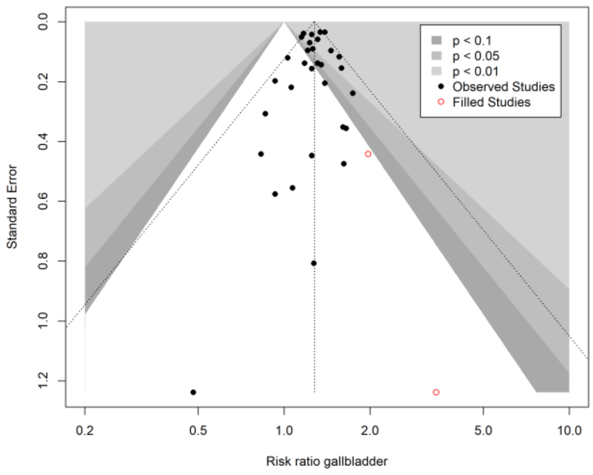

N filled studies: 2
Trim-and-fill RR (95% CI) = 1.28 (1.23 to 1.32)
Meta-analysis RR (95% CI) = 1.27 (1.23 to 1.32)
Difference in RRs = 0.00
Egger's test: Intercept (95% CI) = -0.18 (-0.78 to 0.43), p = 0.571

#### Supplementary Figure 27: Trim-and-fill for publication bias and Egger's intercept test

RRs were combined using random effects meta-analysis.
Abbreviations: CI=confidence interval, RR= risk ratio

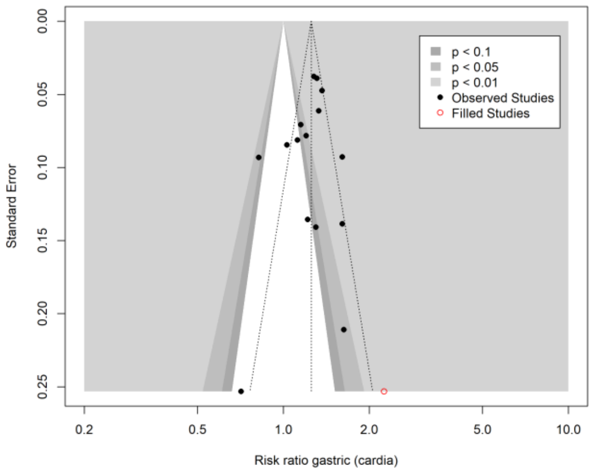

N filled studies: 1
Trim-and-fill RR (95% CI) = 1.25 (1.12 to 1.4)
Meta-analysis RR (95% CI) = 1.23 (1.11 to 1.36)
Difference in RRs = 0.02
Egger's test: Intercept (95% CI) = -0.99 (-3.12 to 1.14), p = 0.378

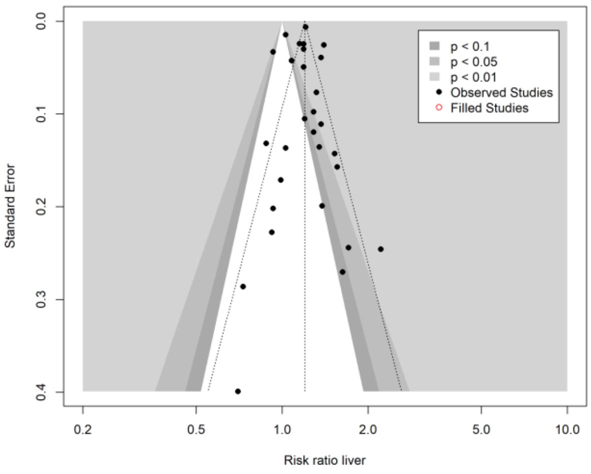

N filled studies: 0
Trim-and-fill RR (95% CI) = 1.2 (1.12 to 1.28)
Meta-analysis RR (95% CI) = 1.2 (1.12 to 1.28)
Difference in OR = N/A
Egger's test: Intercept (95% CI) = -0.02 (-1.39 to 1.35), p = 0.979

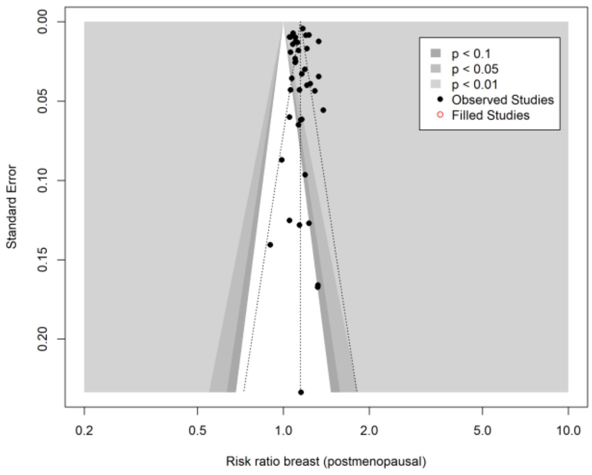

N filled studies: 0
Trim-and-fill RR (95% CI) = 1.15 (1.12 to 1.17)
Meta-analysis RR (95% CI) = 1.15 (1.12 to 1.17)
Difference in OR = N/A
Egger's test: Intercept (95% CI) = -0.14 (-1.92 to 1.65), p = 0.883

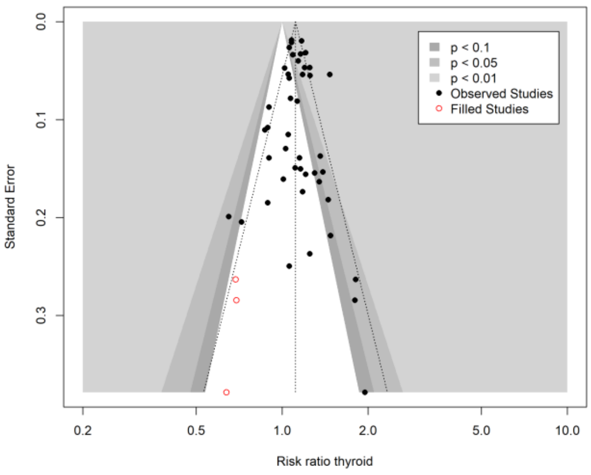

N filled studies: 3
Trim-and-fill RR (95% CI) = 1.11 (1.06 to 1.17)
Meta-analysis RR (95% CI) = 1.12 (1.07 to 1.17)
Difference in RRs = -0.01
Egger's test: Intercept (95% CI) = 0.23 (-0.51 to 0.97), p = 0.538

**Supplementary Figure 27 (continued) Trim-and-fill for publication bias and Egger's intercept test**

RRs were combined using random effects meta-analysis.
Abbreviations: CI=confidence interval, RR= risk ratio

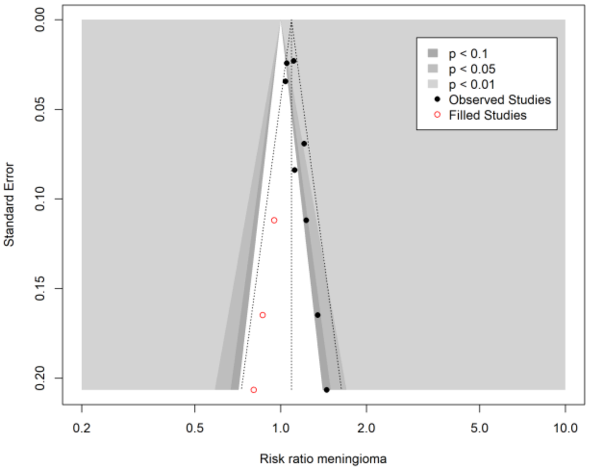

N filled studies: 3
Trim-and-fill RR (95% CI) = 1.09 (1.02 to 1.17)
Meta-analysis RR (95% CI) = 1.11 (1.05 to 1.16)
Difference in RRs = -0.01
Egger's test: Intercept (95% CI) = 1.46 (0.29 to 2.63), p = 0.05

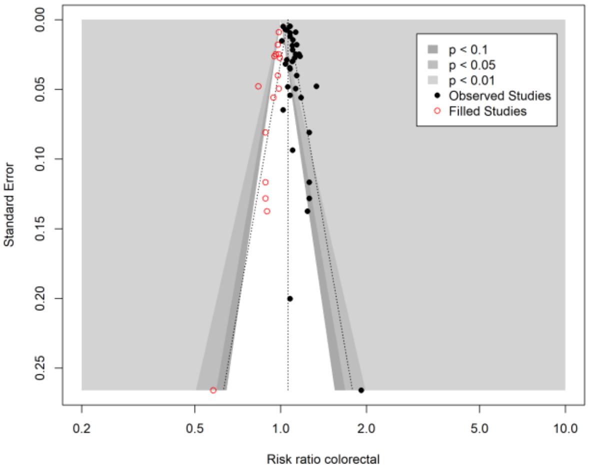

N filled studies: 15
Trim-and-fill RR (95% CI) = 1.06 (1.03 to 1.09)
Meta-analysis RR (95% CI) = 1.1 (1.08 to 1.12)
Difference in RRs = -0.04
Egger's test: Intercept (95% CI) = 1.8 (0.64 to 2.97), p = 0.005

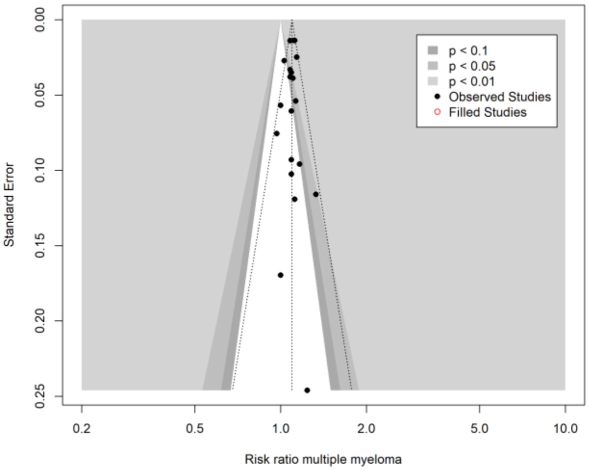

N filled studies: 0
Trim-and-fill RR (95% CI) = 1.1 (1.08 to 1.11)
Meta-analysis RR (95% CI) = 1.1 (1.08 to 1.11)
Difference in OR = N/A
Egger's test: Intercept (95% CI) = -0.17 (-0.93 to 0.6), p = 0.673

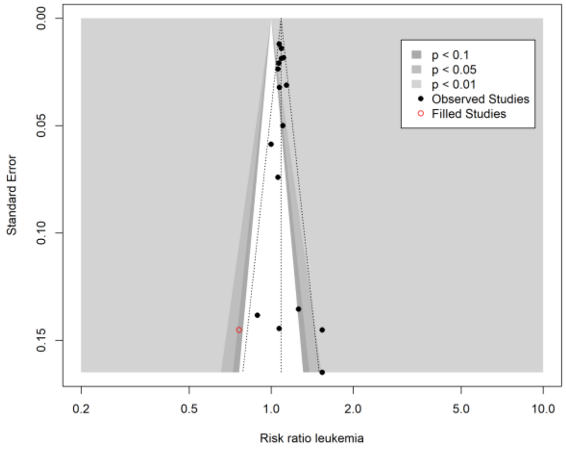

N filled studies: 1
Trim-and-fill RR (95% CI) = 1.09 (1.03 to 1.14)
Meta-analysis RR (95% CI) = 1.09 (1.05 to 1.13)
Difference in RRs = -0.00
Egger's test: Intercept (95% CI) = 0.57 (-0.41 to 1.55), p = 0.274

**Supplementary Figure 27 (continued) Trim-and-fill for publication bias and Egger's intercept test**

RRs were combined using random effects meta-analysis.
Abbreviations: CI=confidence interval, RR= risk ratio

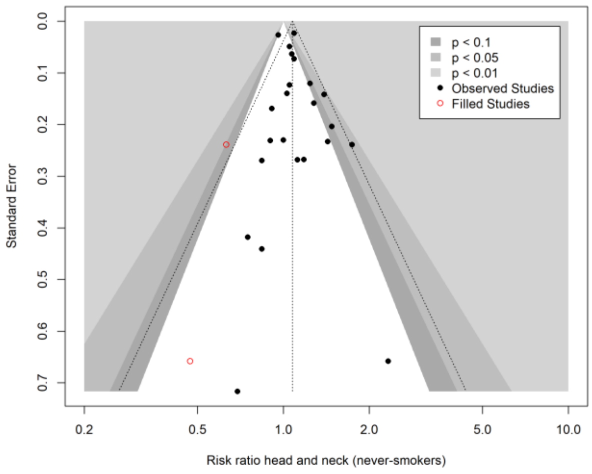

N filled studies: 2
Trim-and-fill RR (95% CI) = 1.08 (1.01 to 1.15)
Meta-analysis RR (95% CI) = 1.07 (1.02 to 1.13)
Difference in RRs = 0.00
Egger's test: Intercept (95% CI) = 0.47 (-0.21 to 1.15), p = 0.187

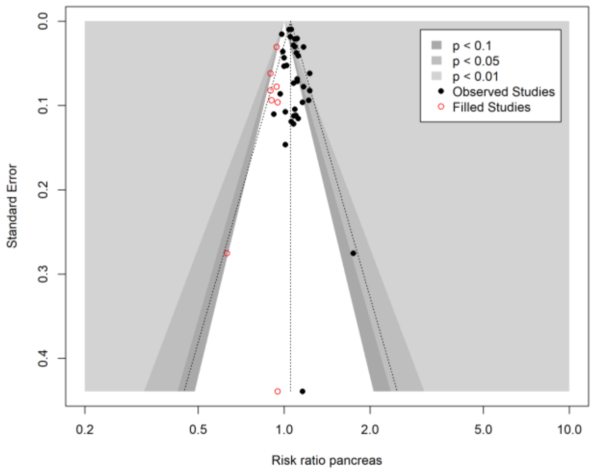

N filled studies: 8
Trim-and-fill RR (95% CI) = 1.05 (1.03 to 1.08)
Meta-analysis RR (95% CI) = 1.07 (1.05 to 1.09)
Difference in RRs = -0.02
Egger's test: Intercept (95% CI) = 0.67 (-0.03 to 1.36), p = 0.07

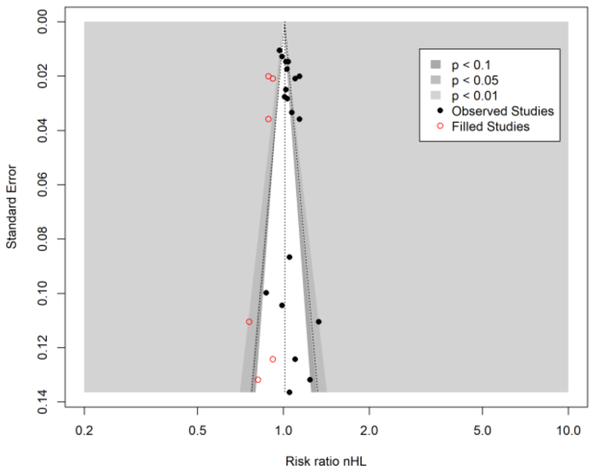

N filled studies: 6
Trim-and-fill RR (95% CI) = 1.01 (0.97 to 1.05)
Meta-analysis RR (95% CI) = 1.05 (1.01 to 1.08)
Difference in RRs = -0.04
Egger's test: Intercept (95% CI) = 1.54 (-0.14 to 3.22), p = 0.089

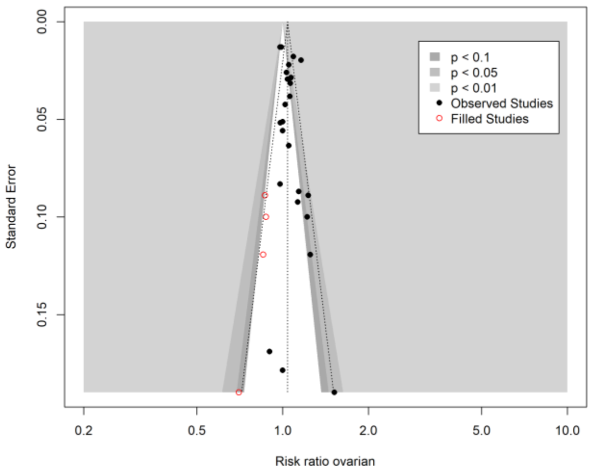

N filled studies: 4
Trim-and-fill RR (95% CI) = 1.04 (1.01 to 1.08)
Meta-analysis RR (95% CI) = 1.05 (1.02 to 1.08)
Difference in RRs = -0.01
Egger's test: Intercept (95% CI) = 1.06 (-0.21 to 2.33), p = 0.117

**Supplementary Figure 27 (continued) Trim-and-fill for publication bias and Egger's intercept test**

RRs were combined using random effects meta-analysis.
Abbreviations: CI=confidence interval, RR= risk ratio

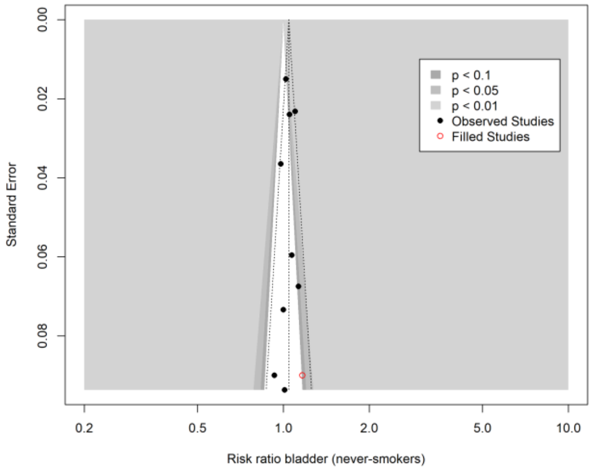

N filled studies: 1
Trim-and-fill RR (95% CI) = 1.04 (1.01 to 1.08)
Meta-analysis RR (95% CI) = 1.04 (1.01 to 1.07)
Difference in RRs = 0.00
Egger's test: Intercept (95% CI) = -0.1 (-1.76 to 1.56), p = 0.908

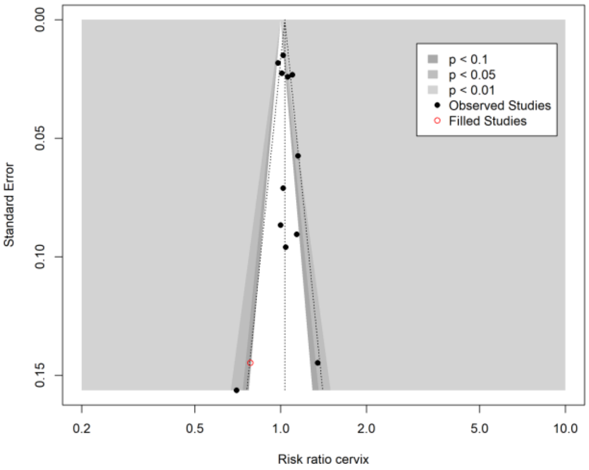

N filled studies: 1
Trim-and-fill RR (95% CI) = 1.03 (0.97 to 1.1)
Meta-analysis RR (95% CI) = 1.04 (0.99 to 1.1)
Difference in RRs = -0.01
Egger's test: Intercept (95% CI) = 0.55 (-1.13 to 2.23), p = 0.537

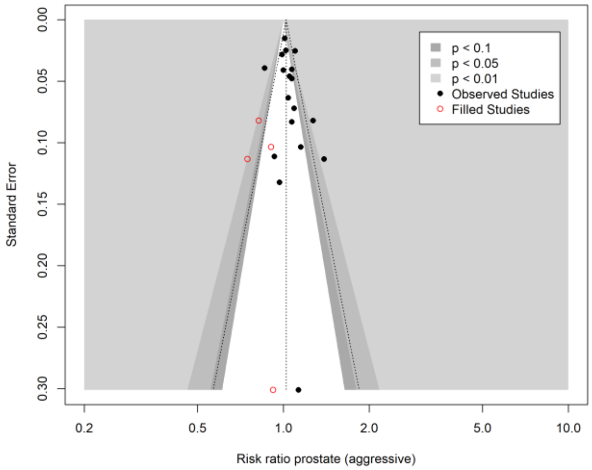

N filled studies: 4
Trim-and-fill RR (95% CI) = 1.02 (0.97 to 1.07)
Meta-analysis RR (95% CI) = 1.04 (1 to 1.09)
Difference in RRs = -0.02
Egger's test: Intercept (95% CI) = 0.82 (-0.47 to 2.12), p = 0.23

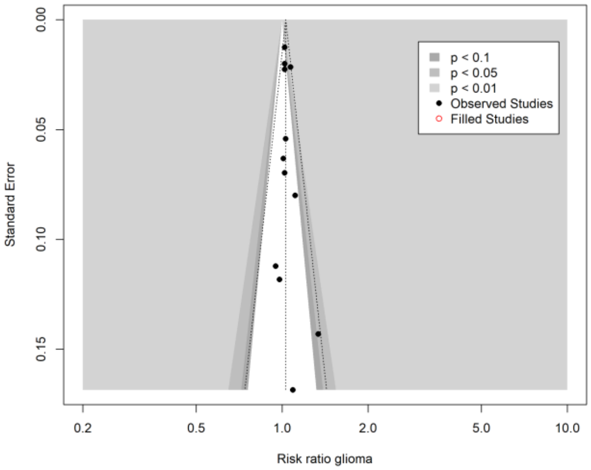

N filled studies: 0
Trim-and-fill RR (95% CI) = 1.03 (1.01 to 1.05)
Meta-analysis RR (95% CI) = 1.03 (1.01 to 1.05)
Difference in OR = N/A
Egger's test: Intercept (95% CI) = 0.37 (-0.43 to 1.17), p = 0.39

**Supplementary Figure 27 (continued) Trim-and-fill for publication bias and Egger's intercept test**

RRs were combined using random effects meta-analysis.
Abbreviations: CI=confidence interval, RR= risk ratio

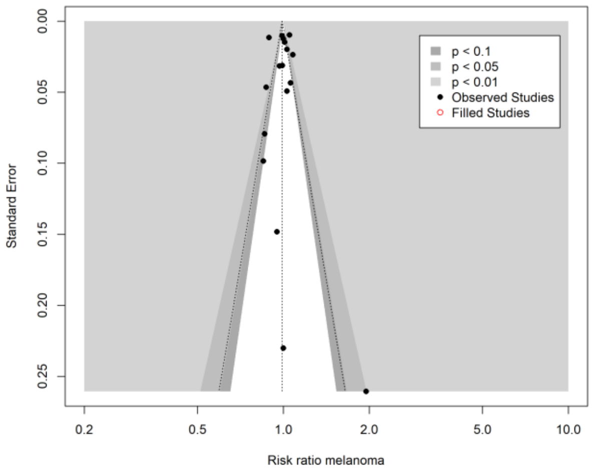

N filled studies: 0
Trim-and-fill RR (95% CI) = 0.99 (0.95 to 1.03)
Meta-analysis RR (95% CI) = 0.99 (0.95 to 1.03)
Difference in OR = N/A
Egger's test: Intercept (95% CI) = 0.02 (-2.5 to 2.54), p = 0.988

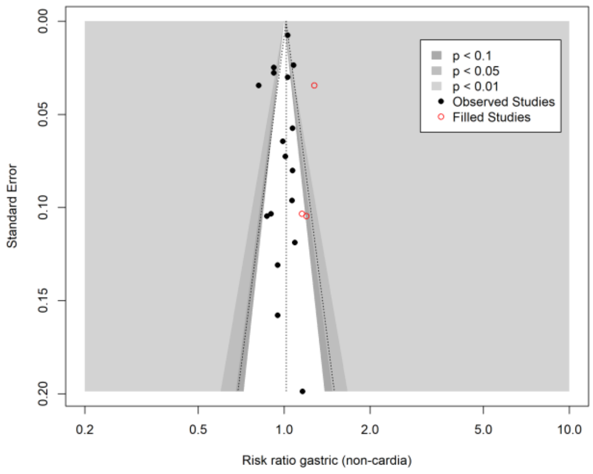

N filled studies: 3
Trim-and-fill RR (95% CI) = 1.02 (0.96 to 1.07)
Meta-analysis RR (95% CI) = 0.98 (0.94 to 1.03)
Difference in RRs = 0.03
Egger's test: Intercept (95% CI) = -0.82 (-2.26 to 0.61), p = 0.28

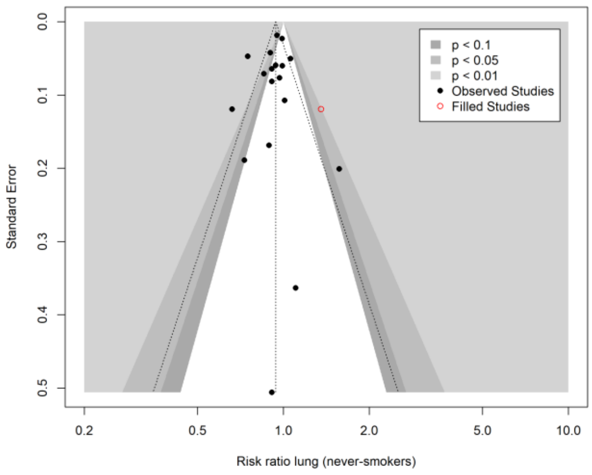

N filled studies: 1
Trim-and-fill RR (95% CI) = 0.94 (0.87 to 1.01)
Meta-analysis RR (95% CI) = 0.92 (0.86 to 0.99)
Difference in RRs = 0.02
Egger's test: Intercept (95% CI) = -0.48 (-1.82 to 0.86), p = 0.492

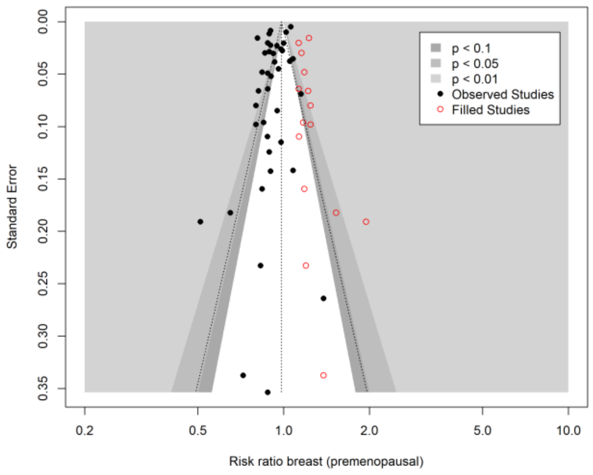

N filled studies: 15
Trim-and-fill RR (95% CI) = 0.98 (0.94 to 1.03)
Meta-analysis RR (95% CI) = 0.92 (0.89 to 0.95)
Difference in RRs = 0.06
Egger's test: Intercept (95% CI) = -2.19 (-3.85 to -0.53), p = 0.014

**Supplementary Figure 27 (continued) Trim-and-fill for publication bias and Egger's intercept test**

RRs were combined using random effects meta-analysis.
Abbreviations: CI=confidence interval, RR= risk ratio

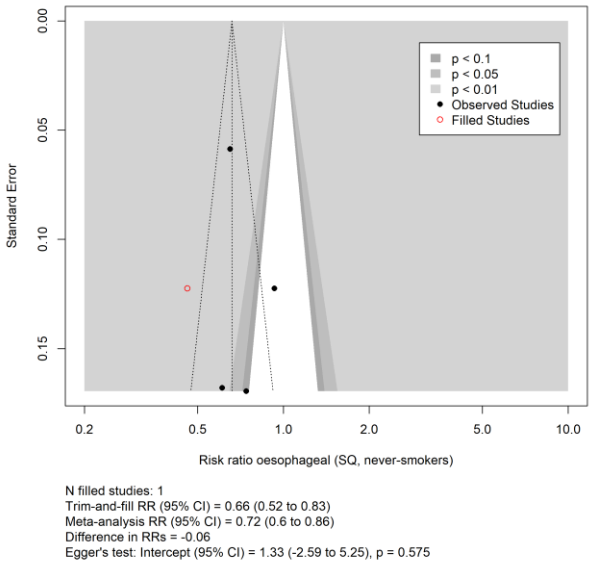

**Supplementary Figure 27 (continued) Trim-and-fill for publication bias and Egger's intercept test**

RRs were combined using random effects meta-analysis.
Abbreviations: CI=confidence interval, RR= risk ratio

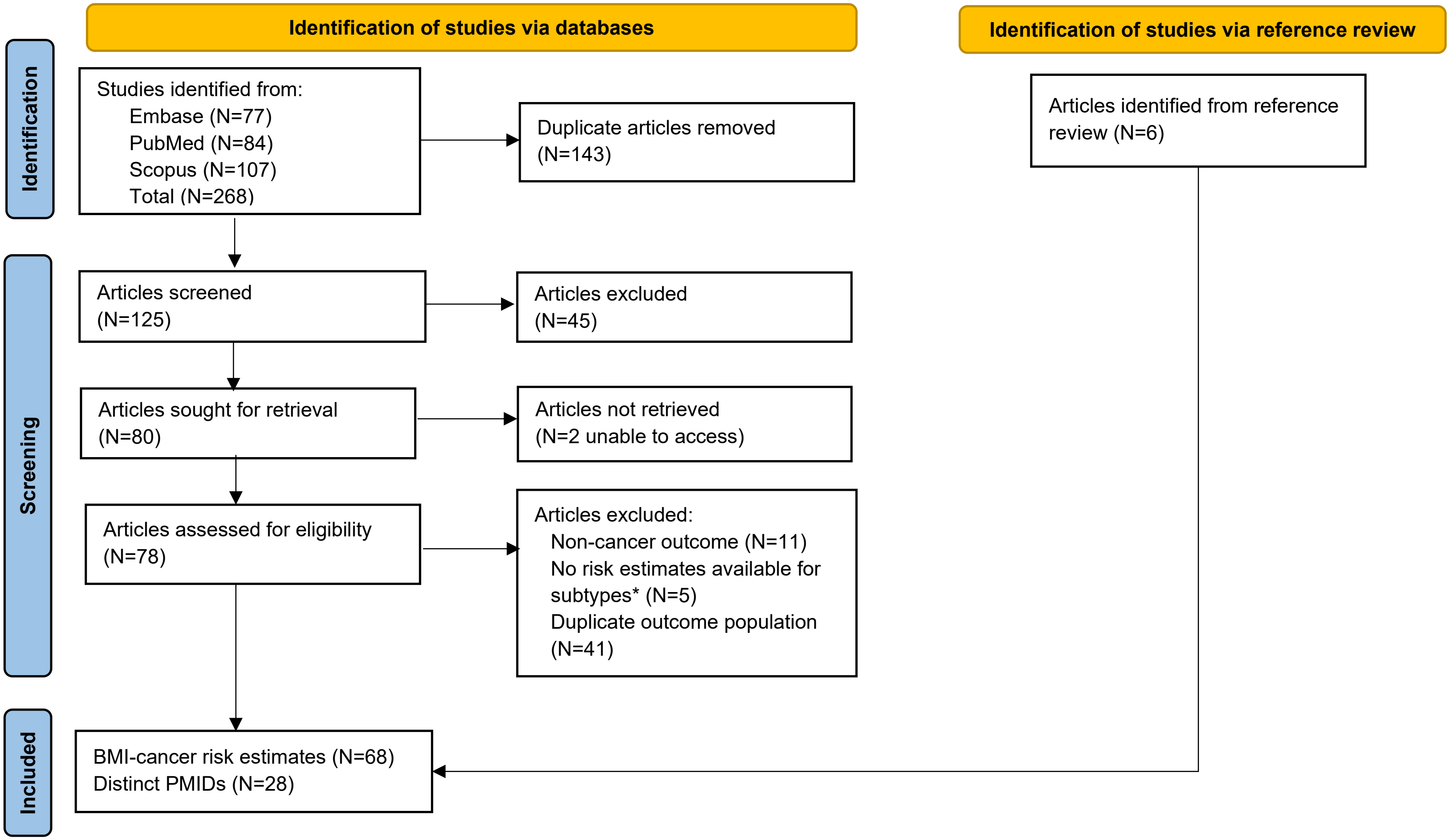

**Supplementary Figure 28: Flow diagram of search strategy and study selection for Mendelian randomisation studies**

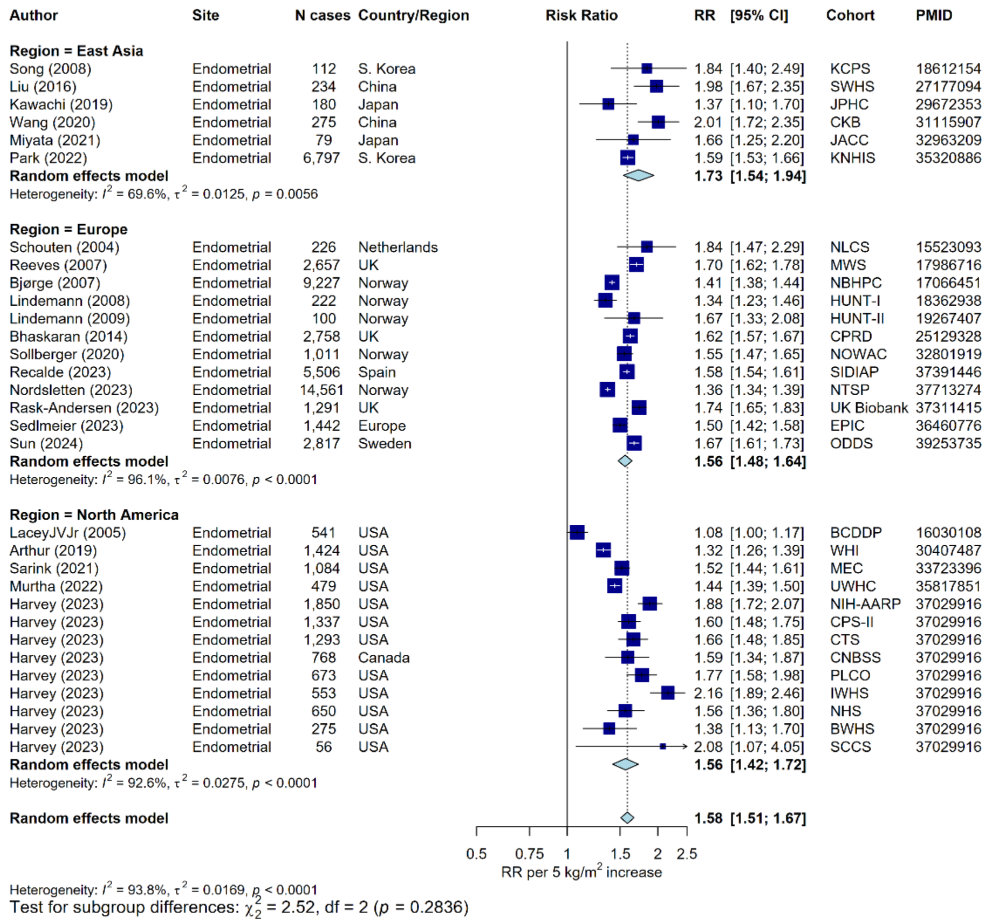

#### Supplementary Figure 29: Meta-analysis of prospective studies on the risk of endometrial cancer in relation to BMI, by region

Study-specific RRs are represented by squares (with their 95% CIs as lines). RRs were combined using random effects meta-analysis (represented by a diamond). Heterogeneity across studies was assessed using $I^2$ and $tau^2$, within region and across all regions. Heterogeneity in the pooled estimates by region was assessed using the Q-statistic. Further details of model adjustments, follow-up time, analytic population for each study are available from **Supplementary Data**.

Abbreviations: BMI=body mass index, CI=confidence interval, PMID=PubMed ID, RR=risk ratio

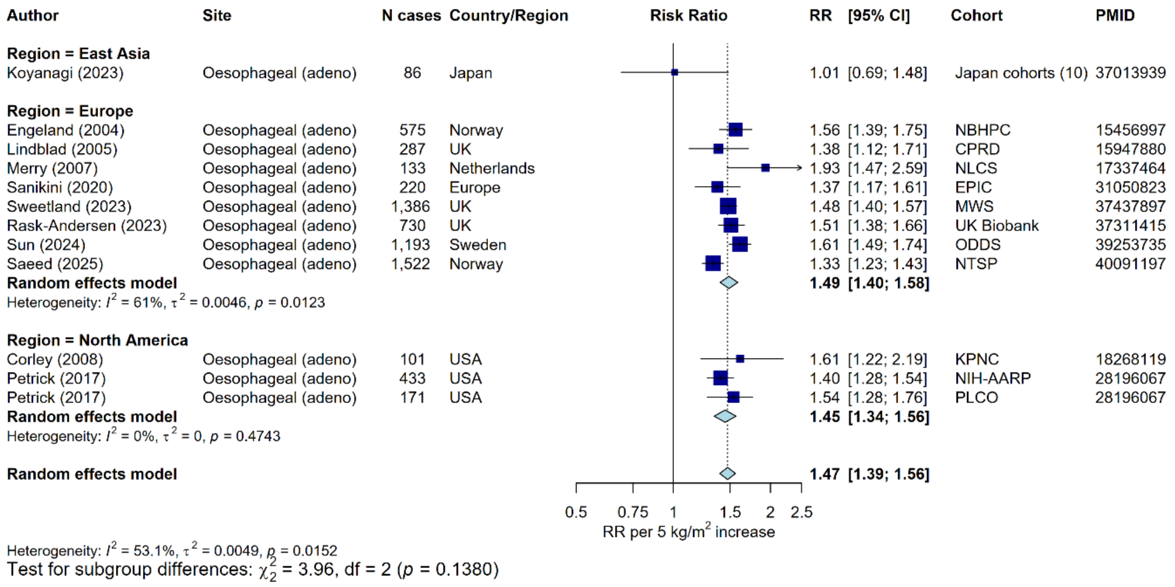

#### Supplementary Figure 30: Meta-analysis of prospective studies on the risk of oesophageal adenocarcinoma cancer in relation to BMI, by region

Study-specific RRs are represented by squares (with their 95% CIs as lines). RRs were combined using random effects meta-analysis (represented by a diamond). Heterogeneity across studies was assessed using $I^2$ and tau$^2$, within region and across all regions. Heterogeneity in the pooled estimates by region was assessed using the Q-statistic. Further details of model adjustments, follow-up time, analytic population for each study are available from **Supplementary Data**.

Abbreviations: BMI=body mass index, CI=confidence interval, PMID=PubMed ID, RR=risk ratio

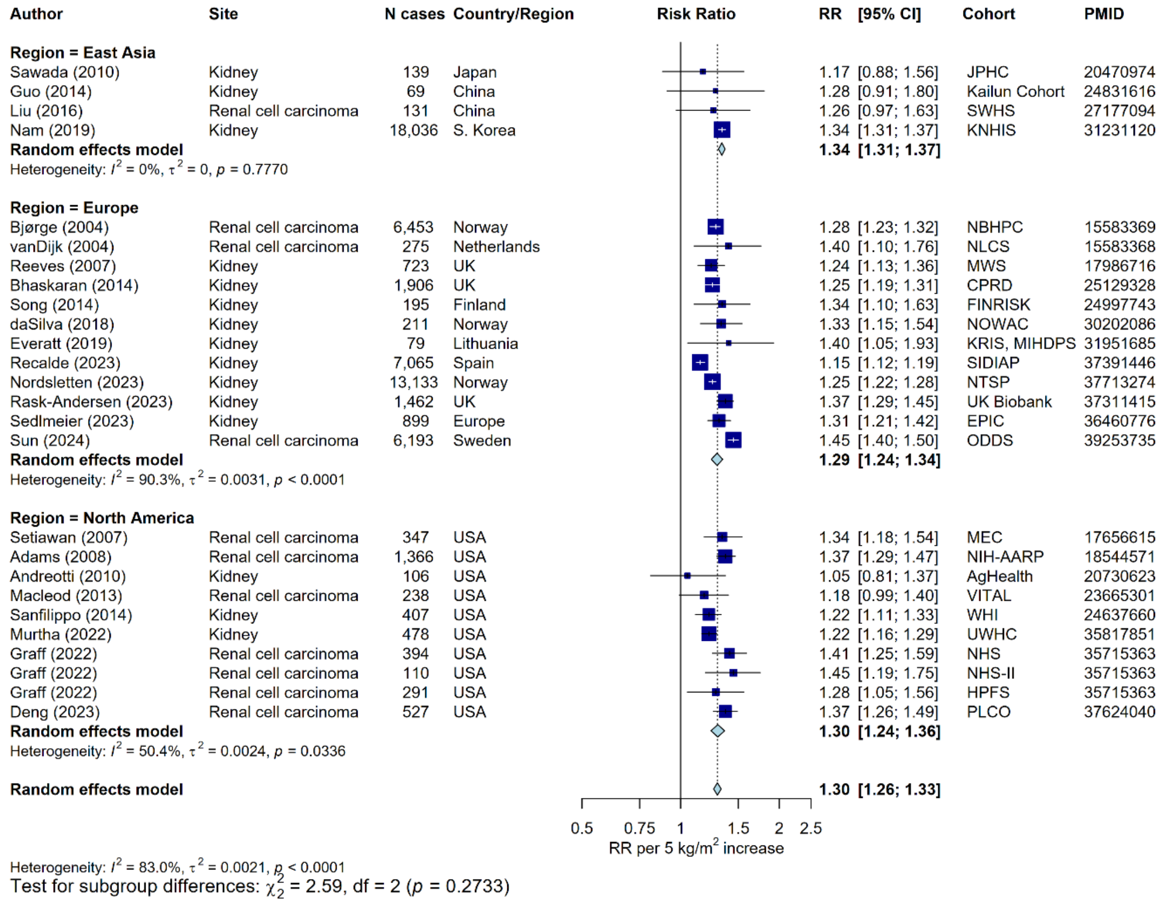

#### Supplementary Figure 31: Meta-analysis of prospective studies on the risk of kidney cancer in relation to BMI, by region

Study-specific RRs are represented by squares (with their 95% CIs as lines). RRs were combined using random effects meta-analysis (represented by a diamond). Heterogeneity across studies was assessed using $I^2$ and $tau^2$, within region and across all regions. Heterogeneity in the pooled estimates by region was assessed using the Q-statistic. Further details of model adjustments, follow-up time, analytic population for each study are available from **Supplementary Data**.

Abbreviations: BMI=body mass index, CI=confidence interval, PMID=PubMed ID, RR=risk ratio

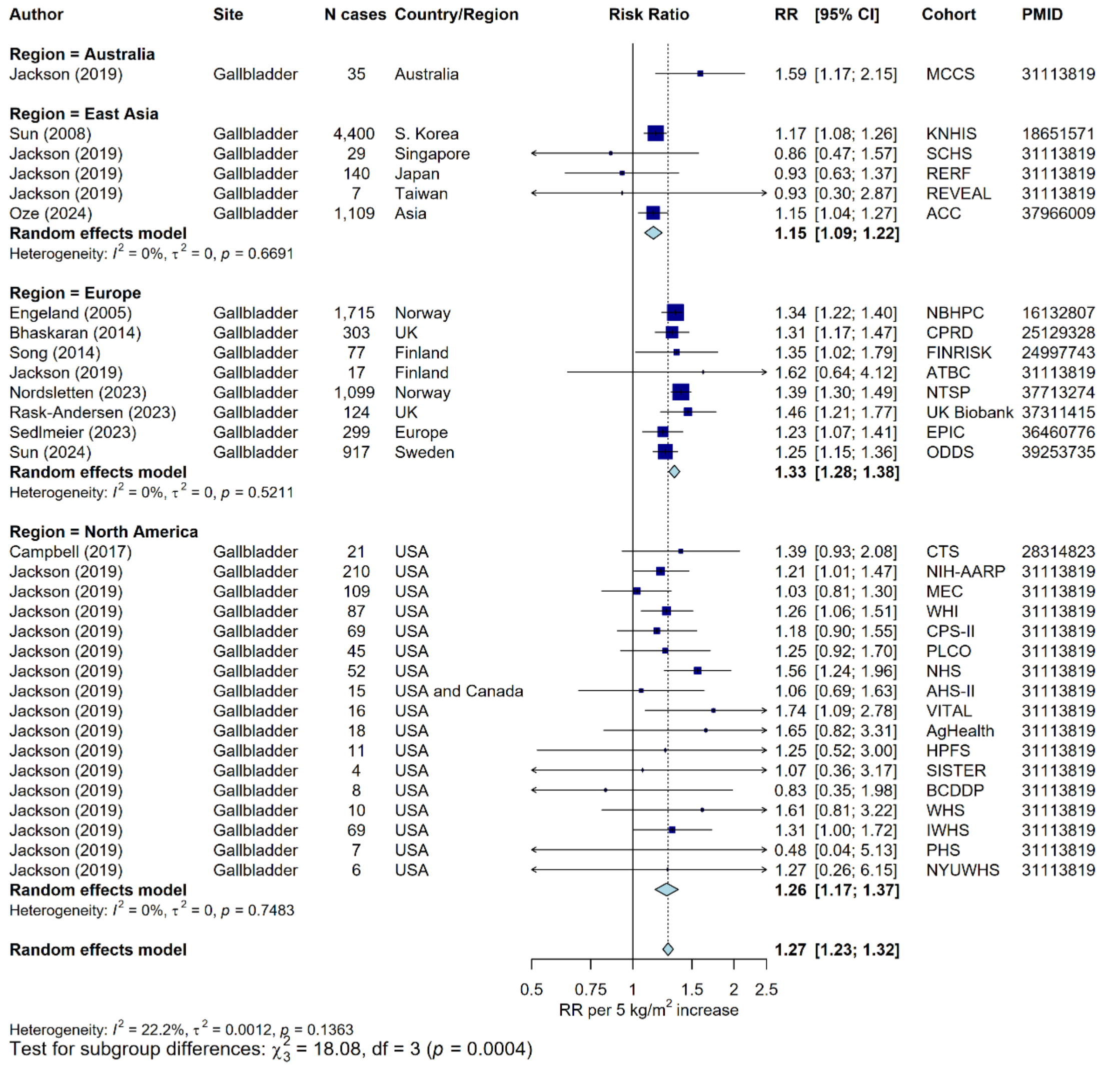

#### Supplementary Figure 32: Meta-analysis of prospective studies on the risk of gallbladder cancer in relation to BMI, by region

Study-specific RRs are represented by squares (with their 95% CIs as lines). RRs were combined using random effects meta-analysis (represented by a diamond). Heterogeneity across studies was assessed using $I^2$ and $tau^2$, within region and across all regions. Heterogeneity in the pooled estimates by region was assessed using the Q-statistic. Further details of model adjustments, follow-up time, analytic population for each study are available from **Supplementary Data**.

Abbreviations: BMI=body mass index, CI=confidence interval, PMID=PubMed ID, RR=risk ratio

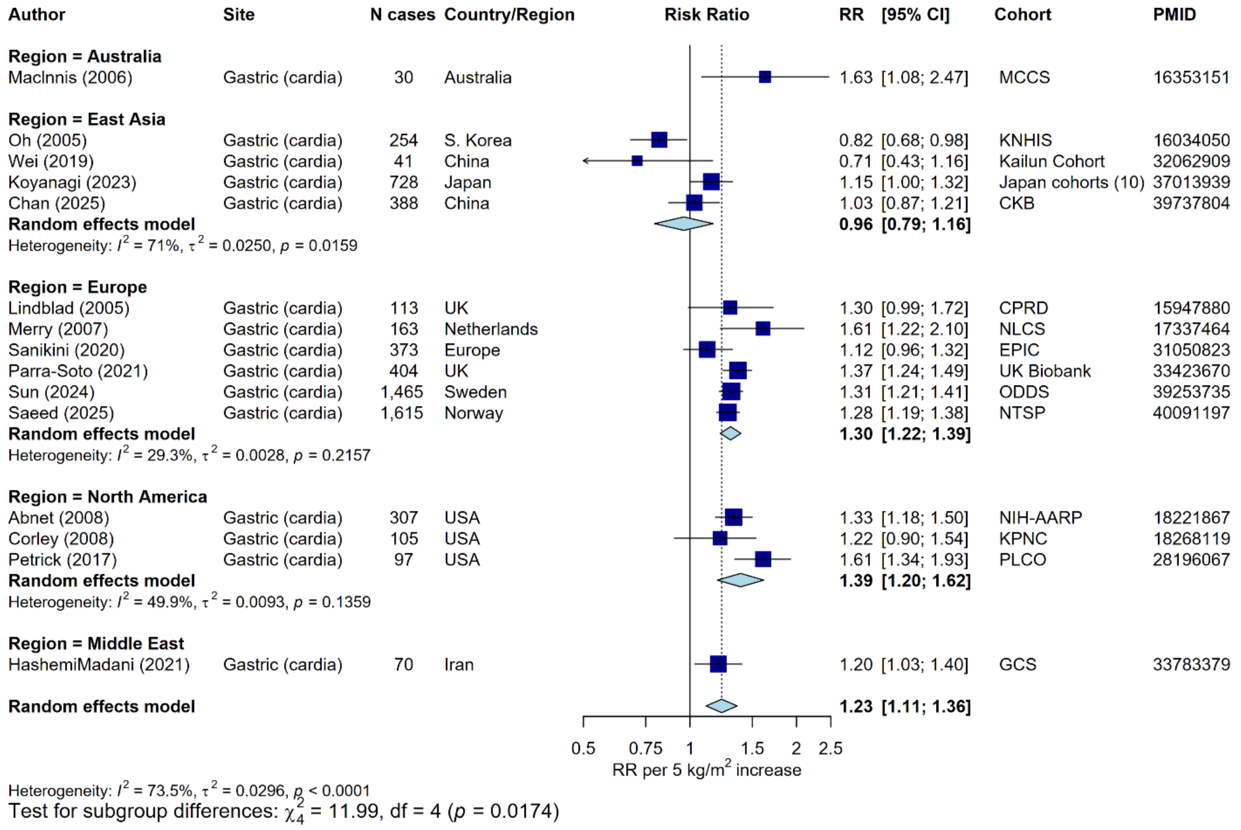

#### Supplementary Figure 33: Meta-analysis of prospective studies on the risk of gastric cancer (cardia) in relation to BMI, by region

Study-specific RRs are represented by squares (with their 95% CIs as lines). RRs were combined using random effects meta-analysis (represented by a diamond). Heterogeneity across studies was assessed using $I^2$ and $tau^2$, within region and across all regions. Heterogeneity in the pooled estimates by region was assessed using the Q-statistic. Further details of model adjustments, follow-up time, analytic population for each study are available from **Supplementary Data**.

Abbreviations: BMI=body mass index, CI=confidence interval, PMID=PubMed ID, RR=risk ratio

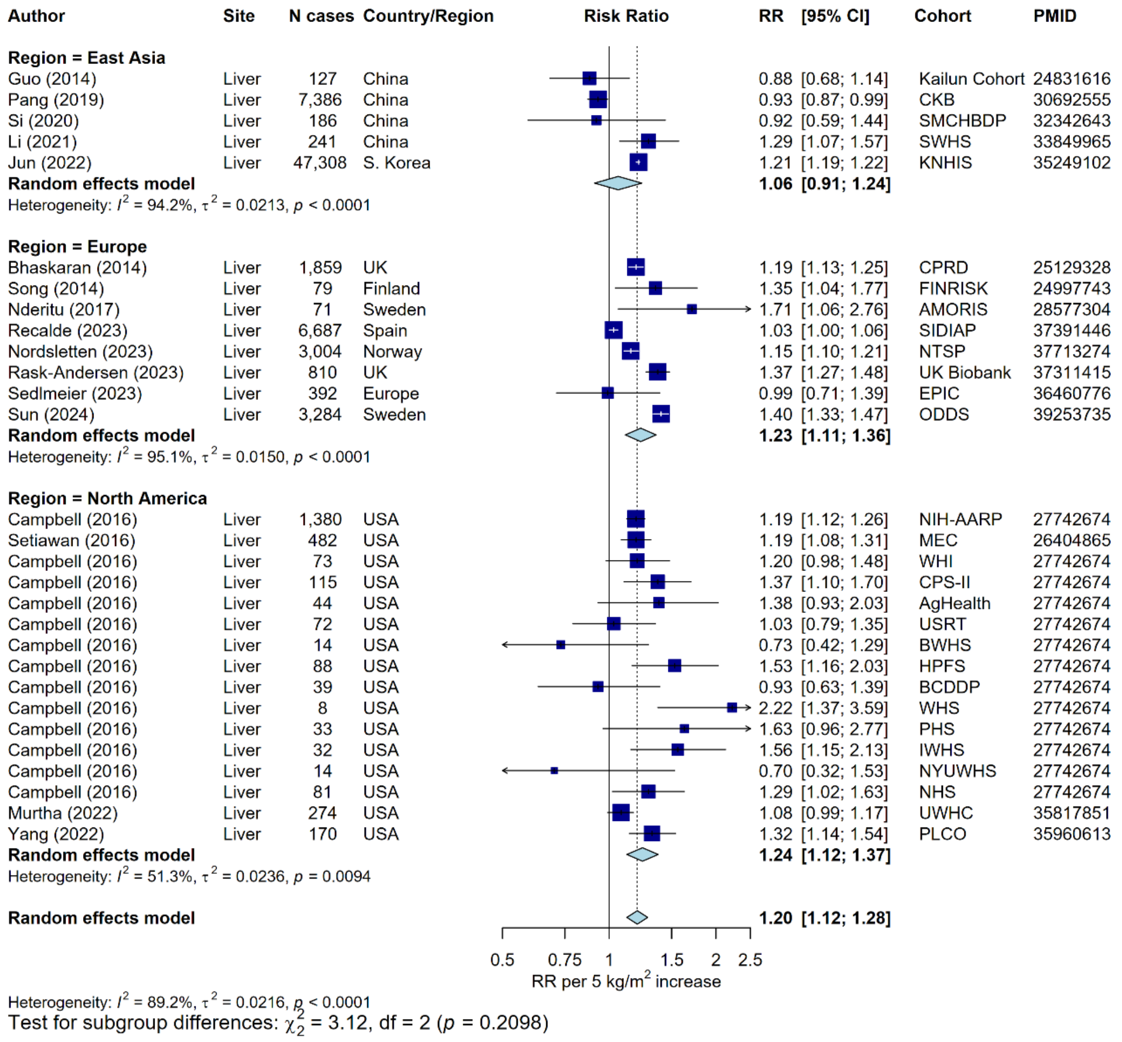

#### Supplementary Figure 34: Meta-analysis of prospective studies on the risk of liver cancer in relation to BMI, by region

Study-specific RRs are represented by squares (with their 95% CIs as lines). RRs were combined using random effects meta-analysis (represented by a diamond). Heterogeneity across studies was assessed using $I^2$ and $tau^2$, within region and across all regions. Heterogeneity in the pooled estimates by region was assessed using the Q-statistic. Further details of model adjustments, follow-up time, analytic population for each study are available from **Supplementary Data**.

Abbreviations: BMI=body mass index, CI=confidence interval, PMID=PubMed ID, RR=risk ratio

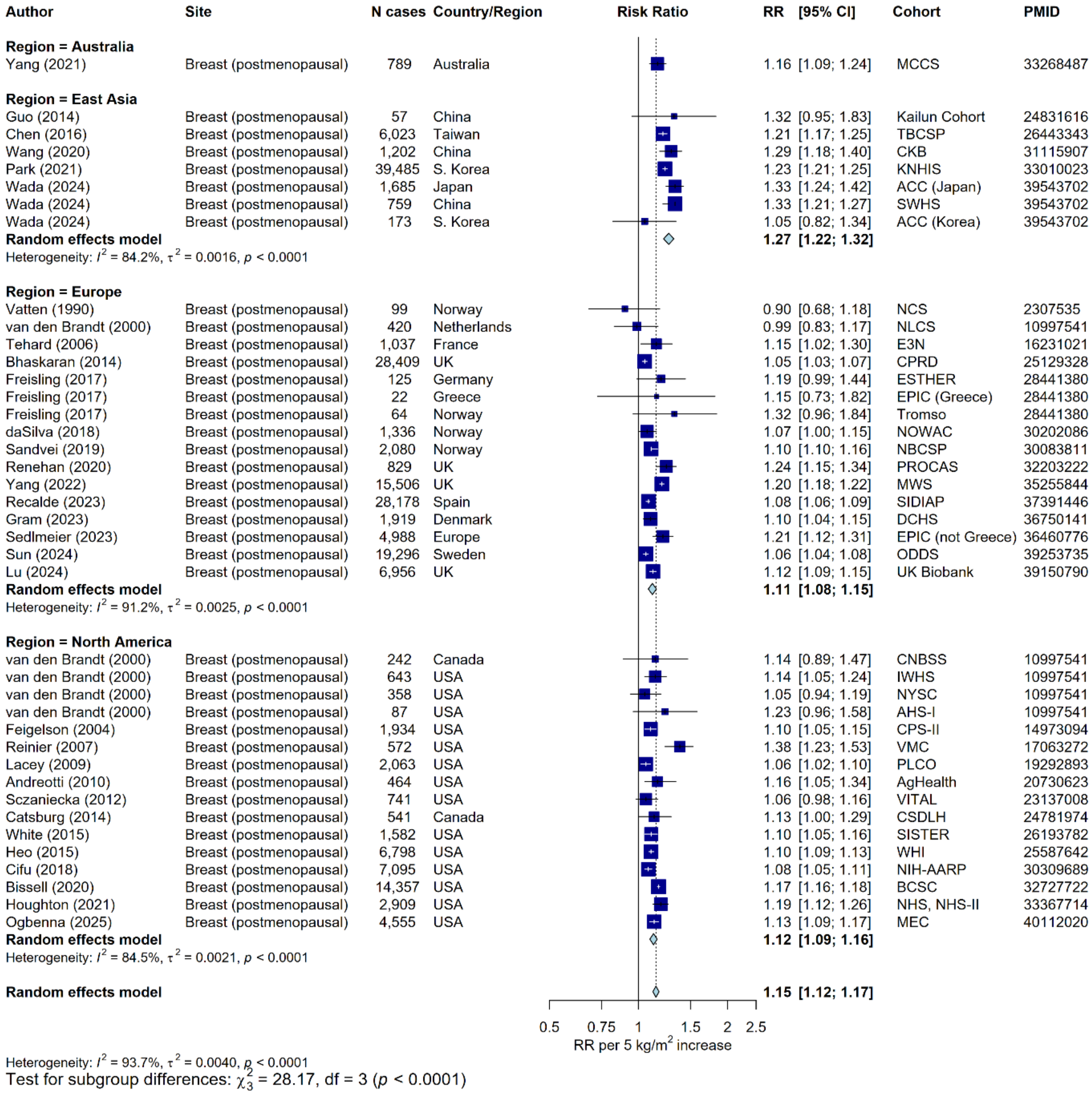

### Figure 35: Meta-analysis of prospective studies on the risk of breast cancer (postmenopausal) in relation to BMI, by region

Study-specific RRs are represented by squares (with their 95% CIs as lines). RRs were combined using random effects meta-analysis (represented by a diamond). Heterogeneity across studies was assessed using $I^2$ and $tau^2$, within region and across all regions. Heterogeneity in the pooled estimates by region was assessed using the Q-statistic. Further details of model adjustments, follow-up time, analytic population for each study are available from **Supplementary Data**. Pooled estimates spanning multiple regions where region-specific RRs were not available were excluded[10].
Abbreviations: BMI=body mass index, CI=confidence interval, PMID=PubMed ID, RR=risk ratio

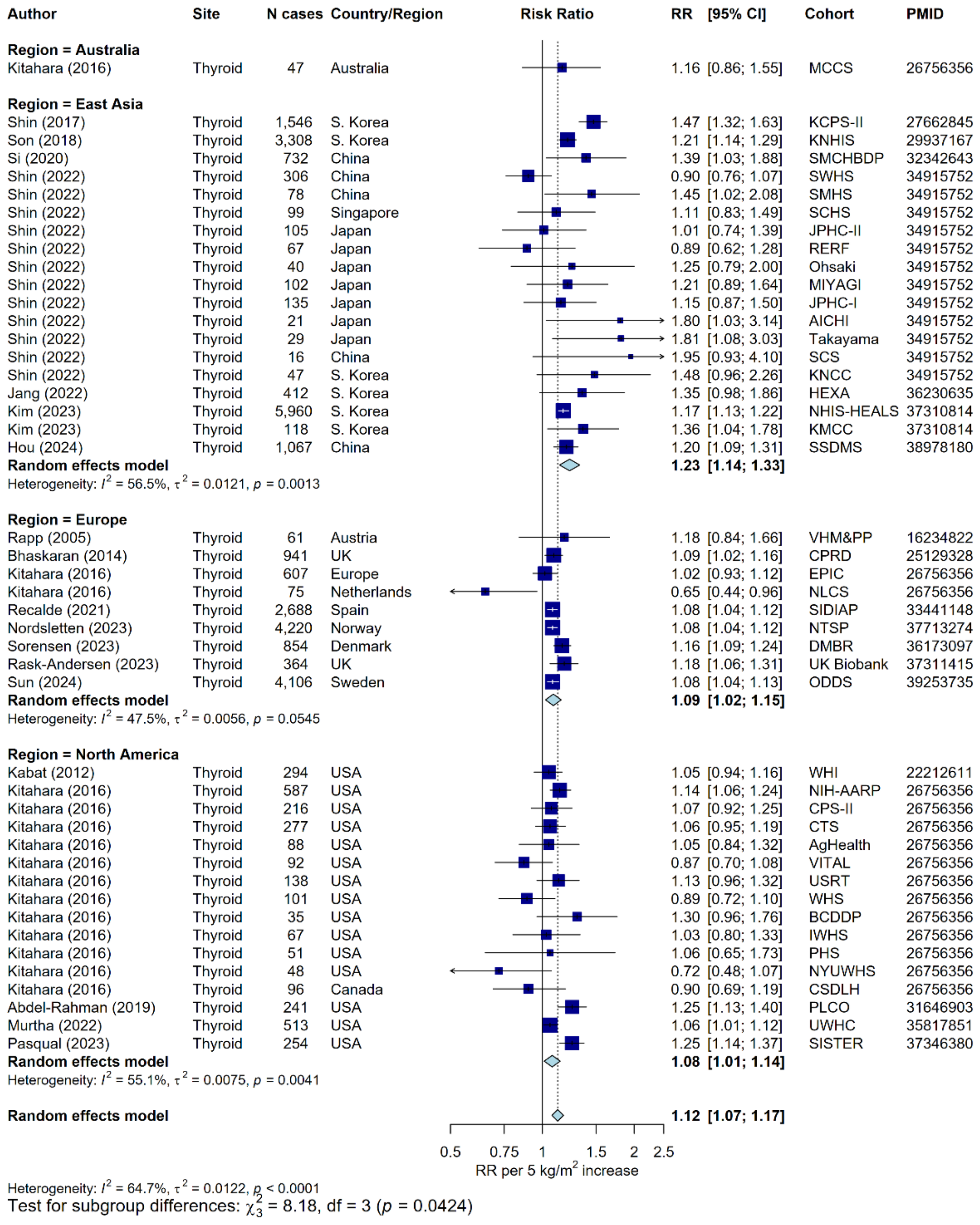

#### Supplementary Figure 36: Meta-analysis of prospective studies on the risk of thyroid cancer in relation to BMI, by region

Study-specific RRs are represented by squares (with their 95% CIs as lines). RRs were combined using random effects meta-analysis (represented by a diamond). Heterogeneity across studies was assessed using $I^2$ and tau$^2$, within region and across all regions. Heterogeneity in the pooled estimates by region was assessed using the Q-statistic. Further details of model adjustments, follow-up time, analytic population for each study are available from **Supplementary Data**.

Abbreviations: BMI=body mass index, CI=confidence interval, PMID=PubMed ID, RR=risk ratio

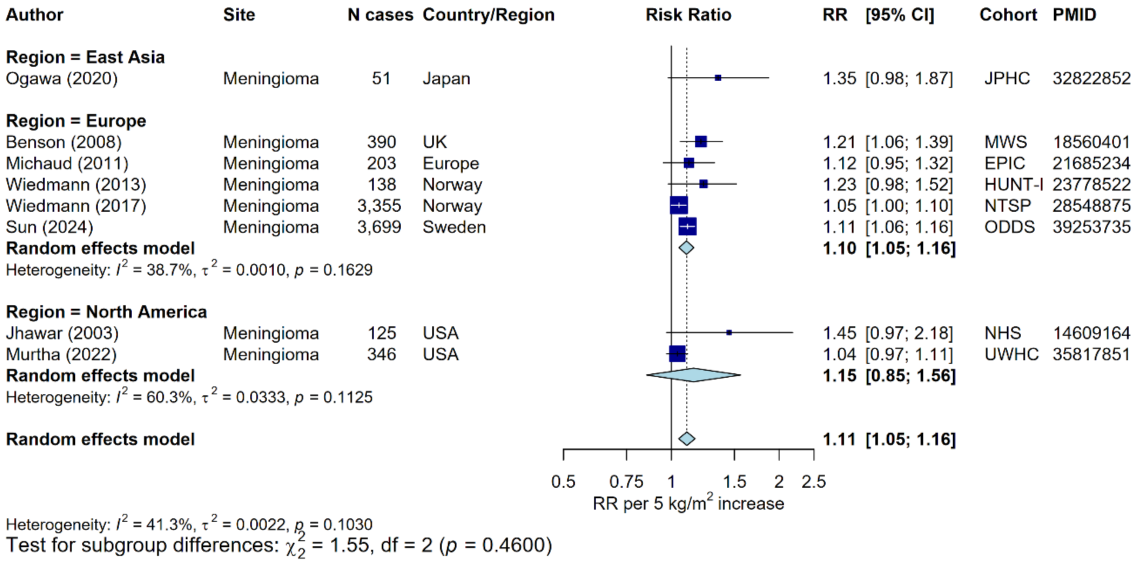

#### Supplementary Figure 37: Meta-analysis of prospective studies on the risk of meningioma cancer in relation to BMI, by region

Study-specific RRs are represented by squares (with their 95% CIs as lines). RRs were combined using random effects meta-analysis (represented by a diamond). Heterogeneity across studies was assessed using $I^2$ and $tau^2$, within region and across all regions. Heterogeneity in the pooled estimates by region was assessed using the Q-statistic. Further details of model adjustments, follow-up time, analytic population for each study are available from **Supplementary Data**.

Abbreviations: BMI=body mass index, CI=confidence interval, PMID=PubMed ID, RR=risk ratio

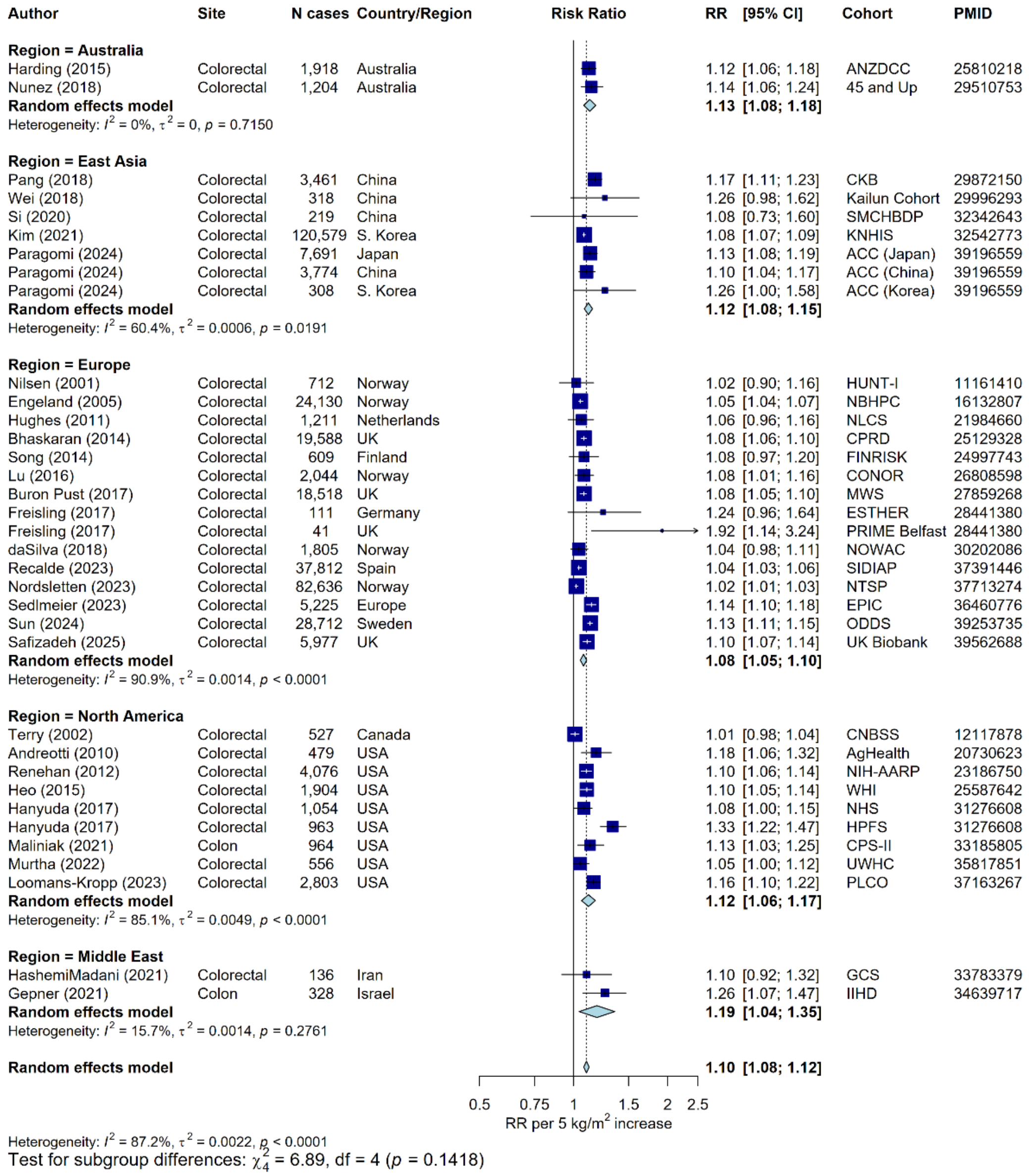

#### Supplementary Figure 38: Meta-analysis of prospective studies on the risk of colorectal cancer in relation to BMI, by region

Study-specific RRs are represented by squares (with their 95% CIs as lines). RRs were combined using random effects meta-analysis (represented by a diamond). Heterogeneity across studies was assessed using $I^2$ and tau$^2$, within region and across all regions. Heterogeneity in the pooled estimates by region was assessed using the Q-statistic. Further details of model adjustments, follow-up time, analytic population for each study are available from **Supplementary Data**.

Abbreviations: BMI=body mass index, CI=confidence interval, PMID=PubMed ID, RR=risk ratio

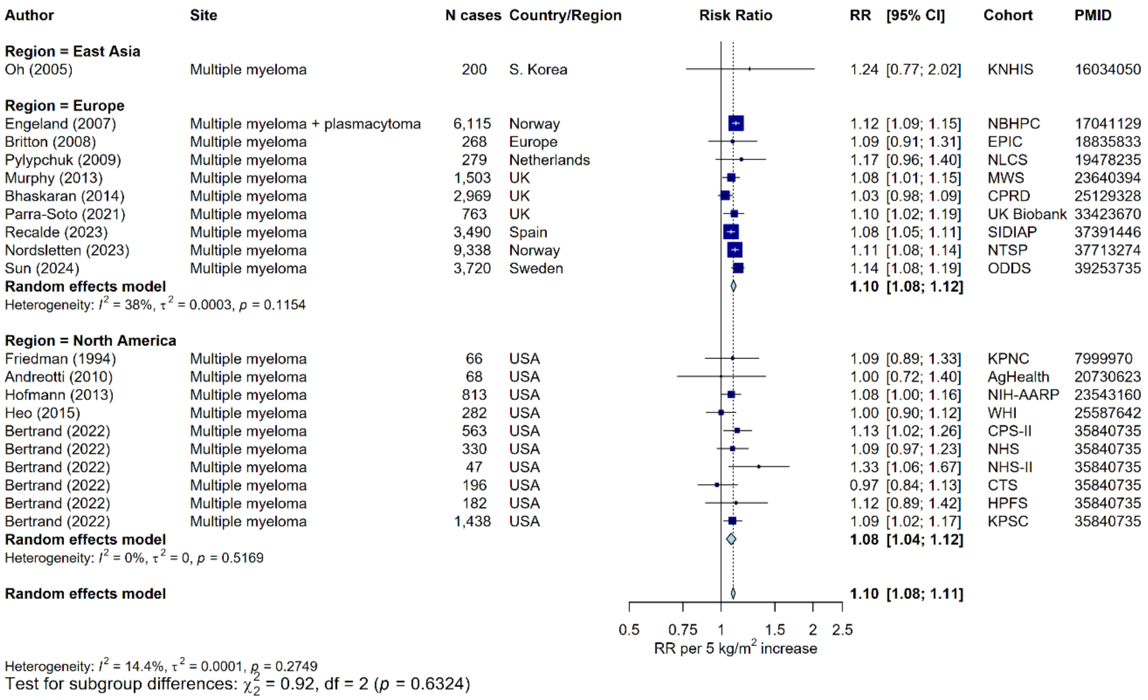

#### Supplementary Figure 39: Meta-analysis of prospective studies on the risk of multiple myeloma in relation to BMI, by region

Study-specific RRs are represented by squares (with their 95% CIs as lines). RRs were combined using random effects meta-analysis (represented by a diamond). Heterogeneity across studies was assessed using $I^2$ and tau$^2$, within region and across all regions. Heterogeneity in the pooled estimates by region was assessed using the Q-statistic. Further details of model adjustments, follow-up time, analytic population for each study are available from **Supplementary Data**.

Abbreviations: BMI=body mass index, CI=confidence interval, PMID=PubMed ID, RR=risk ratio

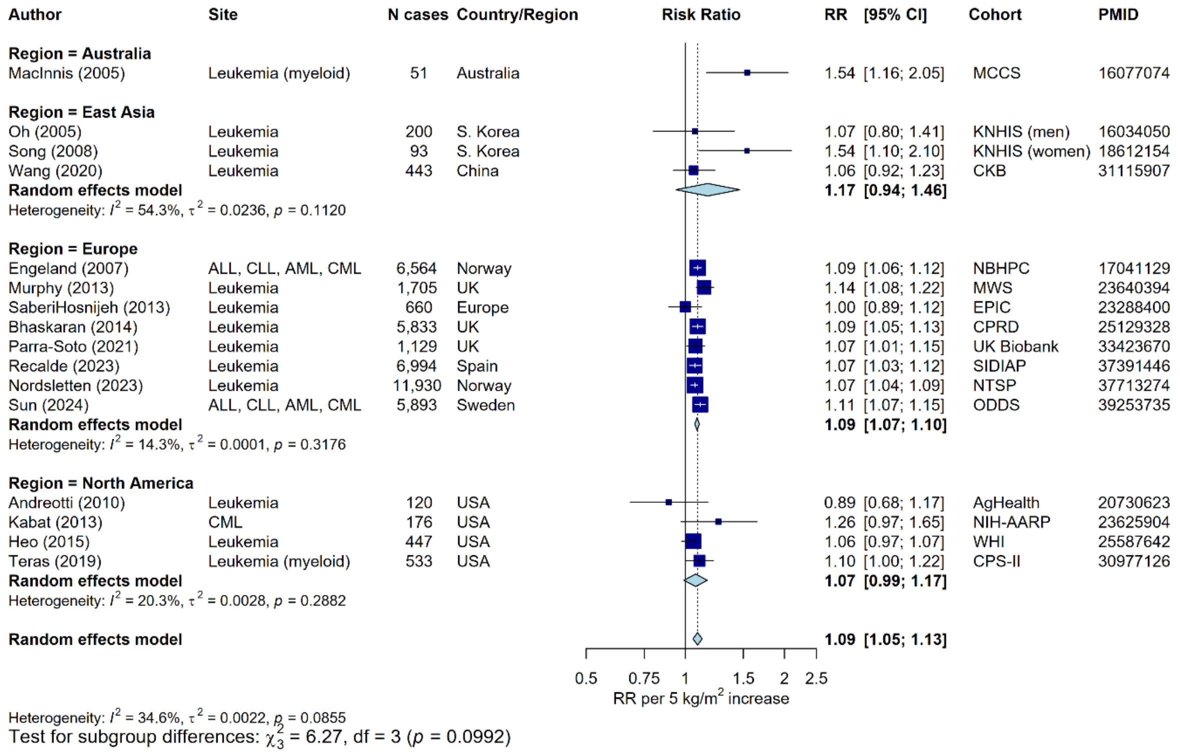

#### Supplementary Figure 40: Meta-analysis of prospective studies on the risk of leukaemia in relation to BMI, by region

Study-specific RRs are represented by squares (with their 95% CIs as lines). RRs were combined using random effects meta-analysis (represented by a diamond). Heterogeneity across studies was assessed using $I^2$ and $tau^2$, within region and across all regions. Heterogeneity in the pooled estimates by region was assessed using the Q-statistic. Further details of model adjustments, follow-up time, analytic population for each study are available from **Supplementary Data**.

Abbreviations: ALL=acute lymphocytic leukaemia, AML=acute myeloid leukaemia, BMI=body mass index, CI=confidence interval, CLL=chronic lymphocytic leukaemia, CML=chronic myeloid leukaemia, PMID=PubMed ID, RR=risk ratio

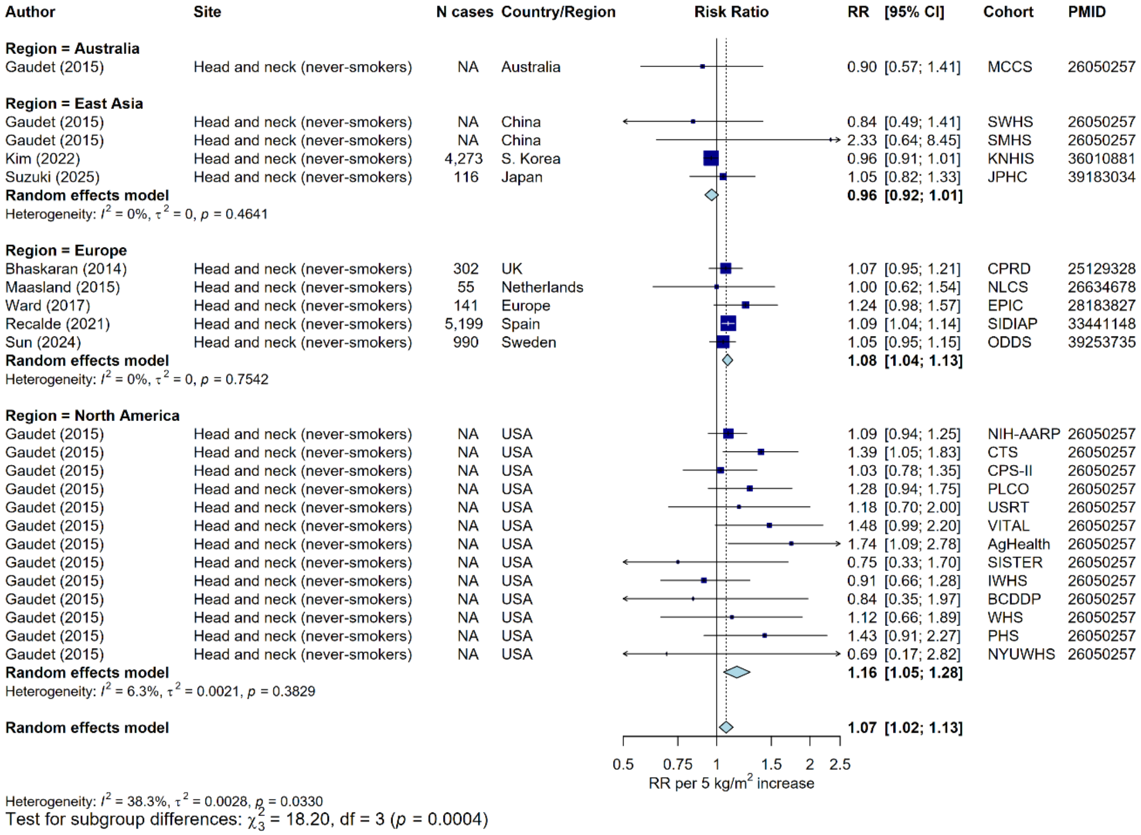

#### Supplementary Figure 41: Meta-analysis of prospective studies on the risk of head and neck cancer (restricted to never-smokers) in relation to BMI, by region

Study-specific RRs are represented by squares (with their 95% CIs as lines). RRs were combined using random effects meta-analysis (represented by a diamond). Heterogeneity across studies was assessed using $I^2$ and $tau^2$, within region and across all regions. Heterogeneity in the pooled estimates by region was assessed using the Q-statistic. Further details of model adjustments, follow-up time, analytic population for each study are available from **Supplementary Data**. Cases numbers for never-smokers for each individual study were not specified in Gaudet et al., 2015 (total cases=796).

Abbreviations: BMI=body mass index, CI=confidence interval, PMID=PubMed ID, RR=risk ratio

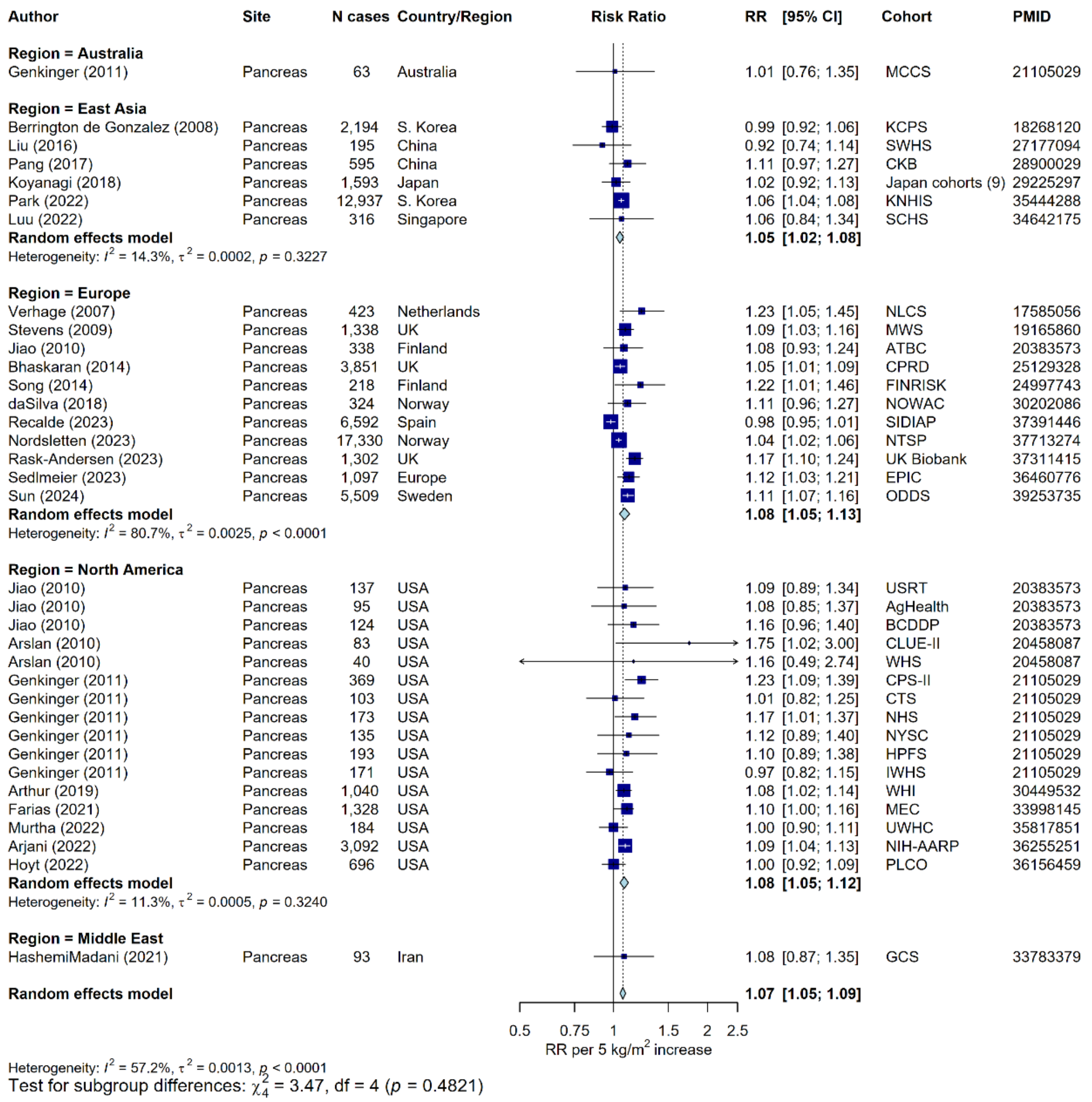

#### Supplementary Figure 42: Meta-analysis of prospective studies on the risk of pancreatic cancer in relation to BMI, by region

Study-specific RRs are represented by squares (with their 95% CIs as lines). RRs were combined using random effects meta-analysis (represented by a diamond). Heterogeneity across studies was assessed using $I^2$ and tau$^2$, within region and across all regions. Heterogeneity in the pooled estimates by region was assessed using the Q-statistic. Further details of model adjustments, follow-up time, analytic population for each study are available from **Supplementary Data**.

Abbreviations: BMI=body mass index, CI=confidence interval, PMID=PubMed ID, RR=risk ratio

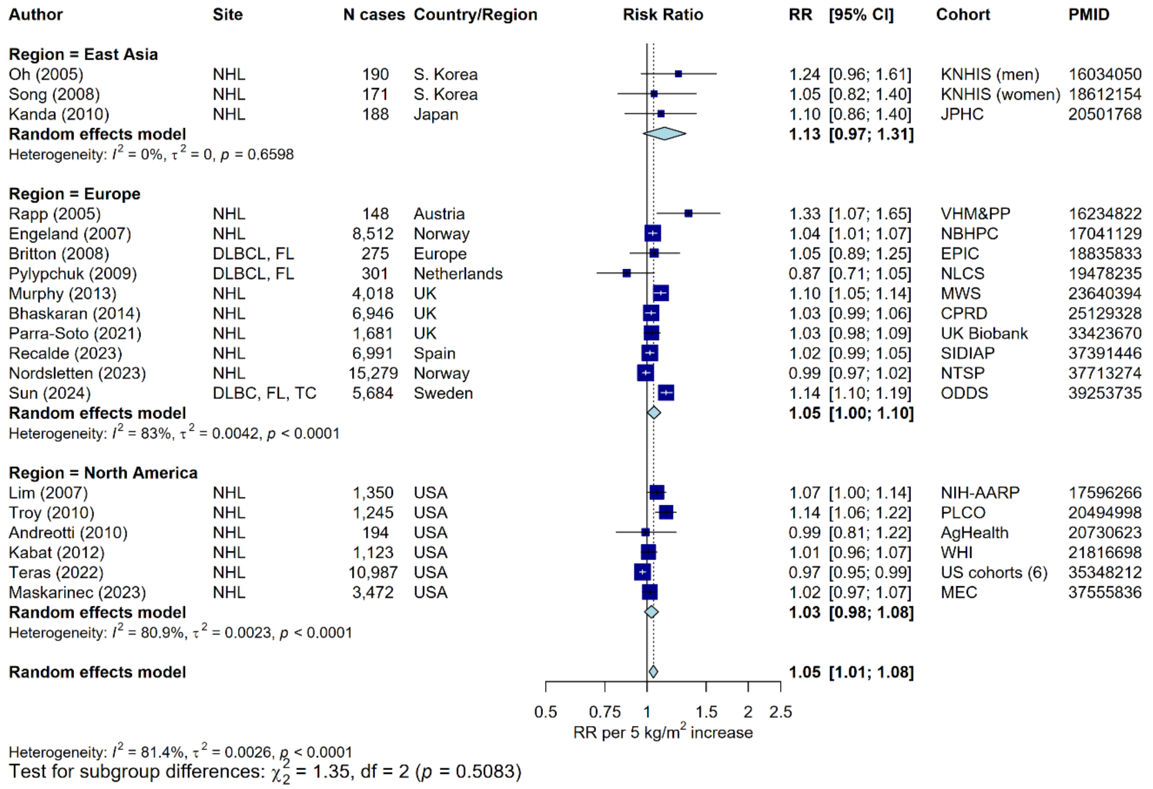

#### Supplementary Figure 43: Meta-analysis of prospective studies on the risk of NHL in relation to BMI, by region

Study-specific RRs are represented by squares (with their 95% CIs as lines). RRs were combined using random effects meta-analysis (represented by a diamond). Heterogeneity across studies was assessed using $I^2$ and tau$^2$, within region and across all regions. Heterogeneity in the pooled estimates by region was assessed using the Q-statistic. Further details of model adjustments, follow-up time, analytic population for each study are available from **Supplementary Data**.

Abbreviations: BMI=body mass index, CI=confidence interval, DLBCL=diffuse large B-cell lymphoma, FL=follicular lymphoma, NHL=non-Hodgkin lymphoma, PMID=PubMed ID, RR=risk ratio, TC=T-cell lymphoma

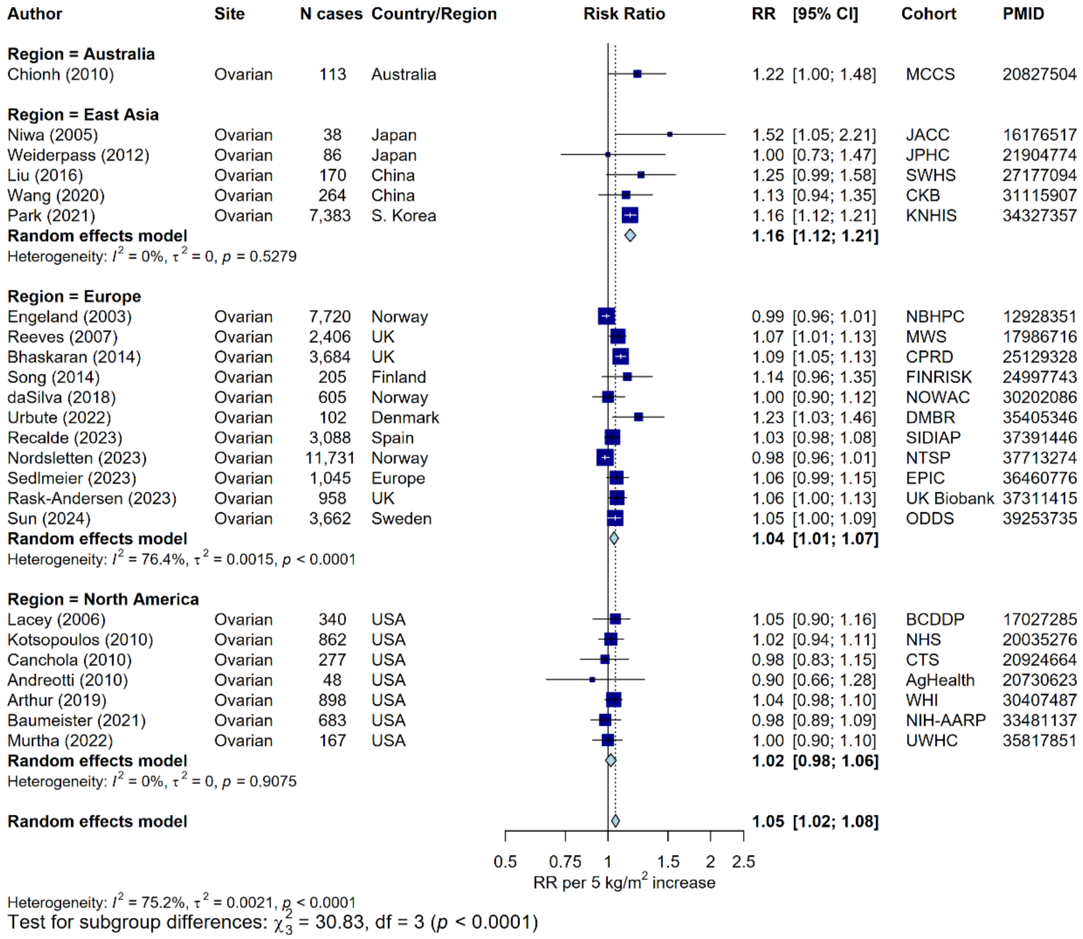

#### Supplementary Figure 44: Meta-analysis of prospective studies on the risk of ovarian cancer in relation to BMI, by region

Study-specific RRs are represented by squares (with their 95% CIs as lines). RRs were combined using random effects meta-analysis (represented by a diamond). Heterogeneity across studies was assessed using $I^2$ and tau$^2$, within region and across all regions. Heterogeneity in the pooled estimates by region was assessed using the Q-statistic. Further details of model adjustments, follow-up time, analytic population for each study are available from **Supplementary Data**.

Abbreviations: BMI=body mass index, CI=confidence interval, PMID=PubMed ID, RR=risk ratio

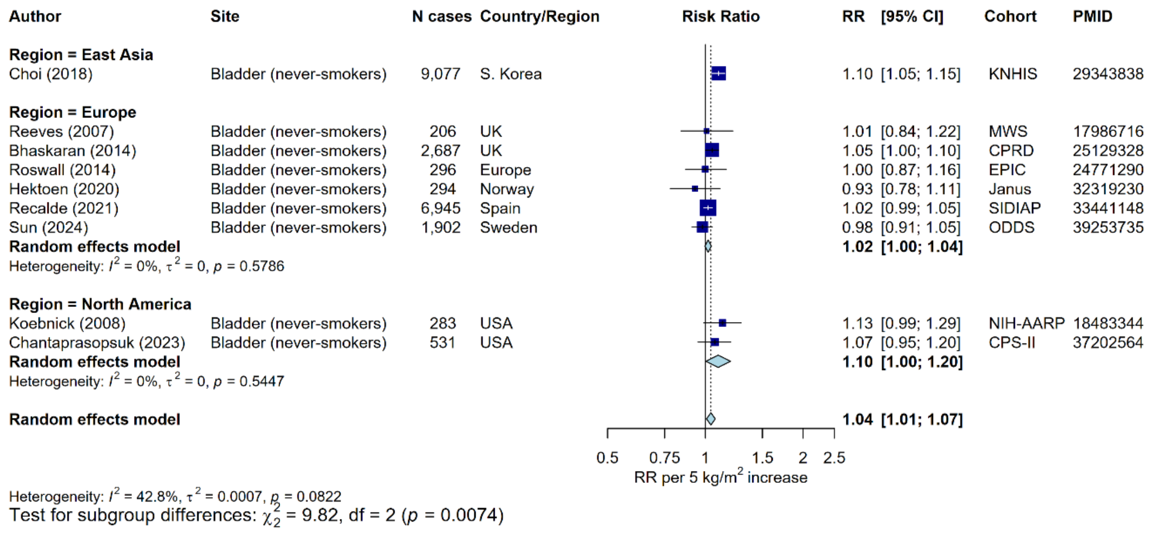

#### Supplementary Figure 45: Meta-analysis of prospective studies on the risk of bladder cancer (restricted to never-smokers) in relation to BMI, by region

Study-specific RRs are represented by squares (with their 95% CIs as lines). RRs were combined using random effects meta-analysis (represented by a diamond). Heterogeneity across studies was assessed using $I^2$ and $tau^2$, within region and across all regions. Heterogeneity in the pooled estimates by region was assessed using the Q-statistic. Further details of model adjustments, follow-up time, analytic population for each study are available from **Supplementary Data**.

Abbreviations: BMI=body mass index, CI=confidence interval, PMID=PubMed ID, RR=risk ratio

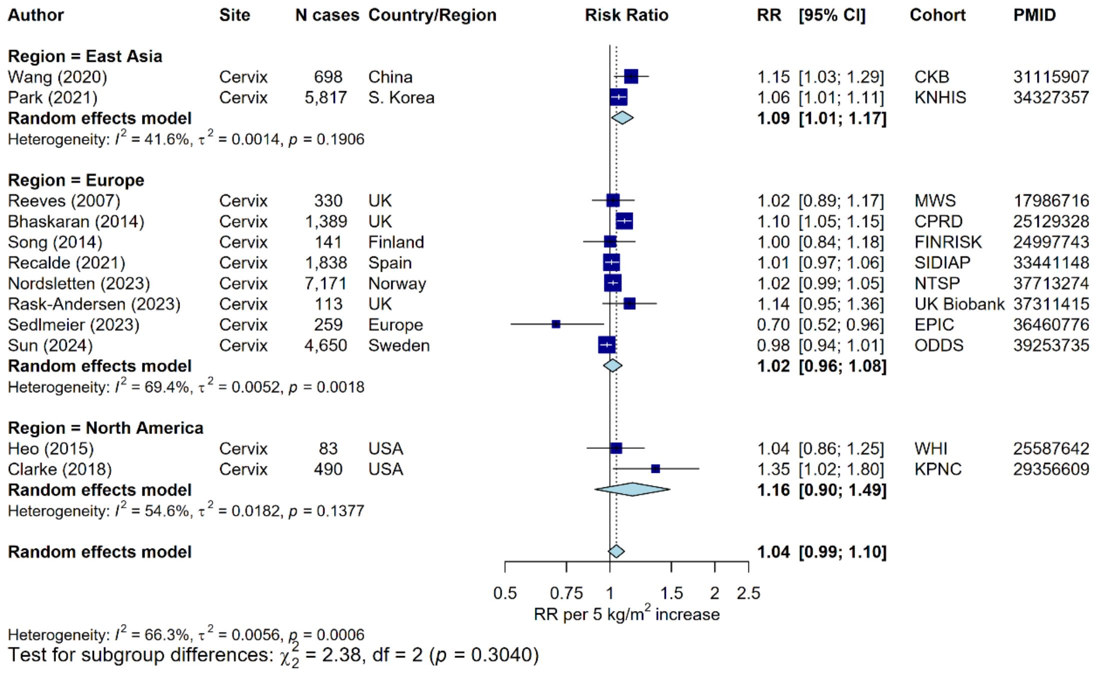

#### Supplementary Figure 46: Meta-analysis of prospective studies on the risk of cervical cancer in relation to BMI, by region

Study-specific RRs are represented by squares (with their 95% CIs as lines). RRs were combined using random effects meta-analysis (represented by a diamond). Heterogeneity across studies was assessed using $I^2$ and $tau^2$, within region and across all regions. Heterogeneity in the pooled estimates by region was assessed using the Q-statistic. Further details of model adjustments, follow-up time, analytic population for each study are available from **Supplementary Data**.

Abbreviations: BMI=body mass index, CI=confidence interval, PMID=PubMed ID, RR=risk ratio

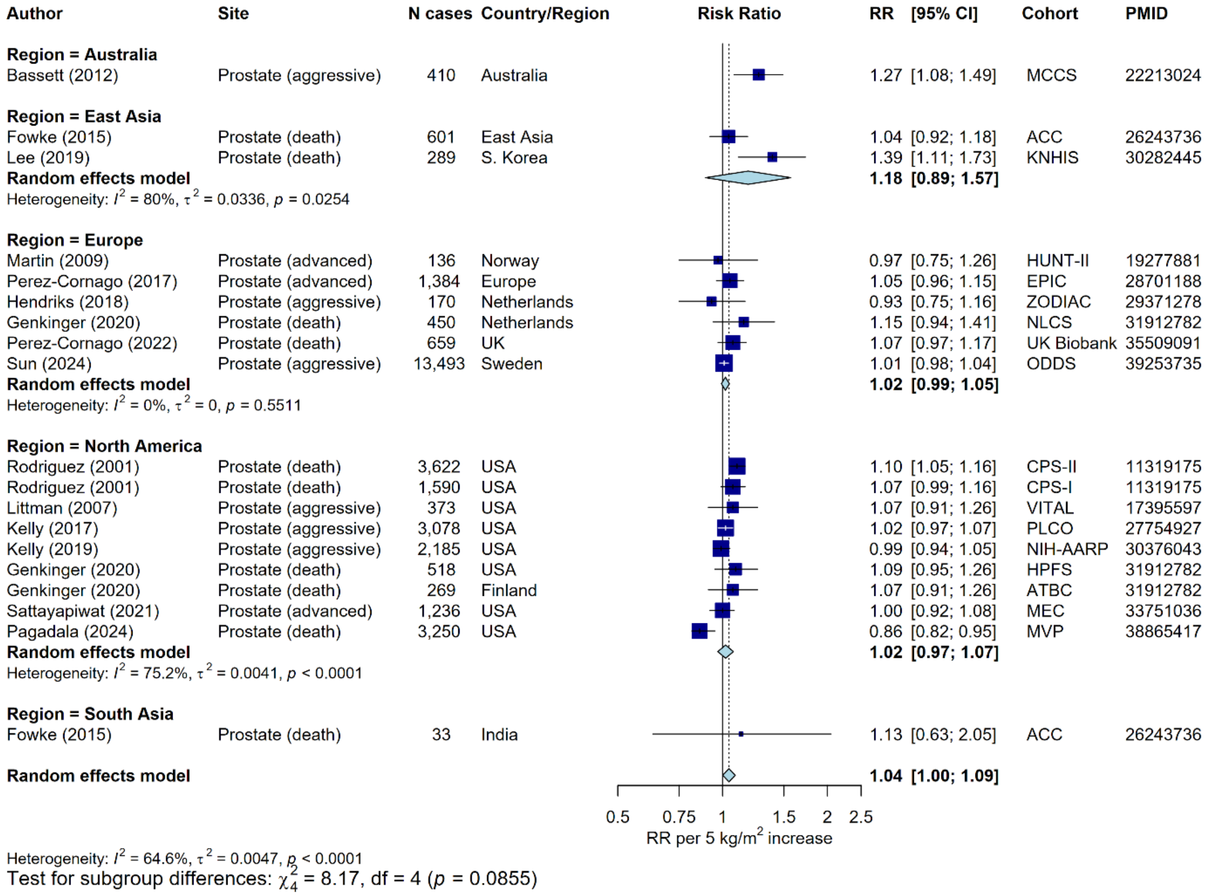

#### Supplementary Figure 47: Meta-analysis of prospective studies on the risk of aggressive prostate cancer in relation to BMI, by region

Aggressive prostate cancer was defined as any of stage 3-4 on the American Joint Committee on Cancer 1992 classification, advanced cancer, advanced or metastatic cancer; metastatic cancer; stage C or D on the Whitmore/Jewett scale; fatal cancer (prostate cancer-specific mortality); high stage or grade; Gleason grade ≥ 7). Study-specific RRs are represented by squares (with their 95% CIs as lines). RRs were combined using random effects meta-analysis (represented by a diamond). Heterogeneity across studies was assessed using $I^2$ and tau$^2$, within region and across all regions. Heterogeneity in the pooled estimates by region was assessed using the Q-statistic. Further details of model adjustments, follow-up time, analytic population for each study are available from **Supplementary Data**.

Abbreviations: BMI=body mass index, CI=confidence interval, PMID=PubMed ID, RR=risk ratio

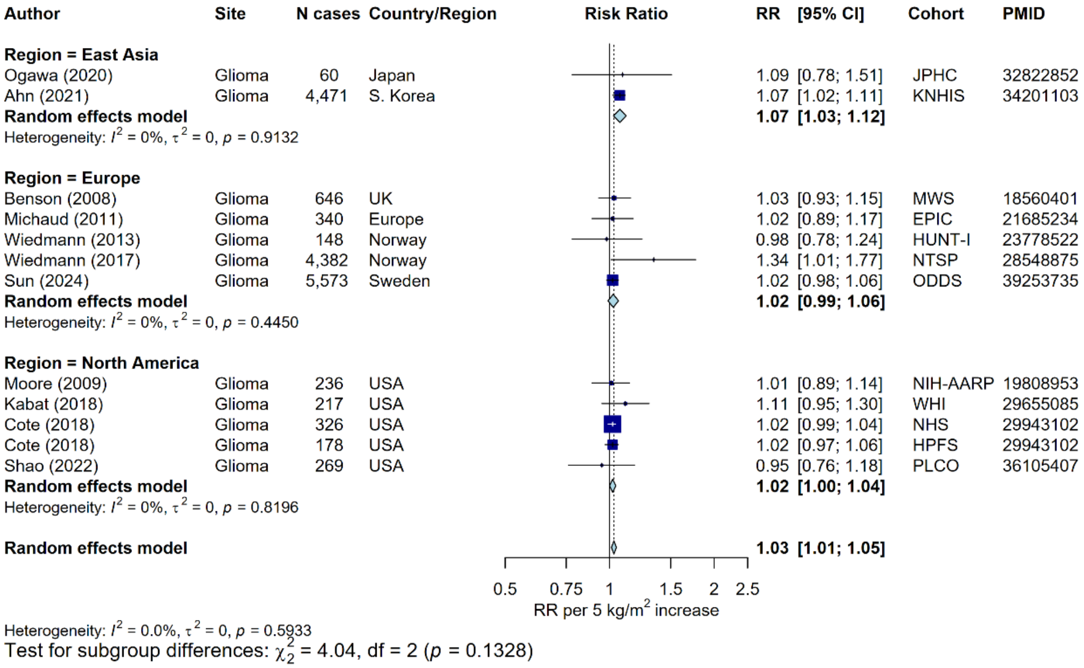

#### Supplementary Figure 48: Meta-analysis of prospective studies on the risk of glioma in relation to BMI, by region

Study-specific RRs are represented by squares (with their 95% CIs as lines). RRs were combined using random effects meta-analysis (represented by a diamond). Heterogeneity across studies was assessed using $I^2$ and $tau^2$, within region and across all regions. Heterogeneity in the pooled estimates by region was assessed using the Q-statistic. Further details of model adjustments, follow-up time, analytic population for each study are available from **Supplementary Data**.

Abbreviations: BMI=body mass index, CI=confidence interval, PMID=PubMed ID, RR=risk ratio

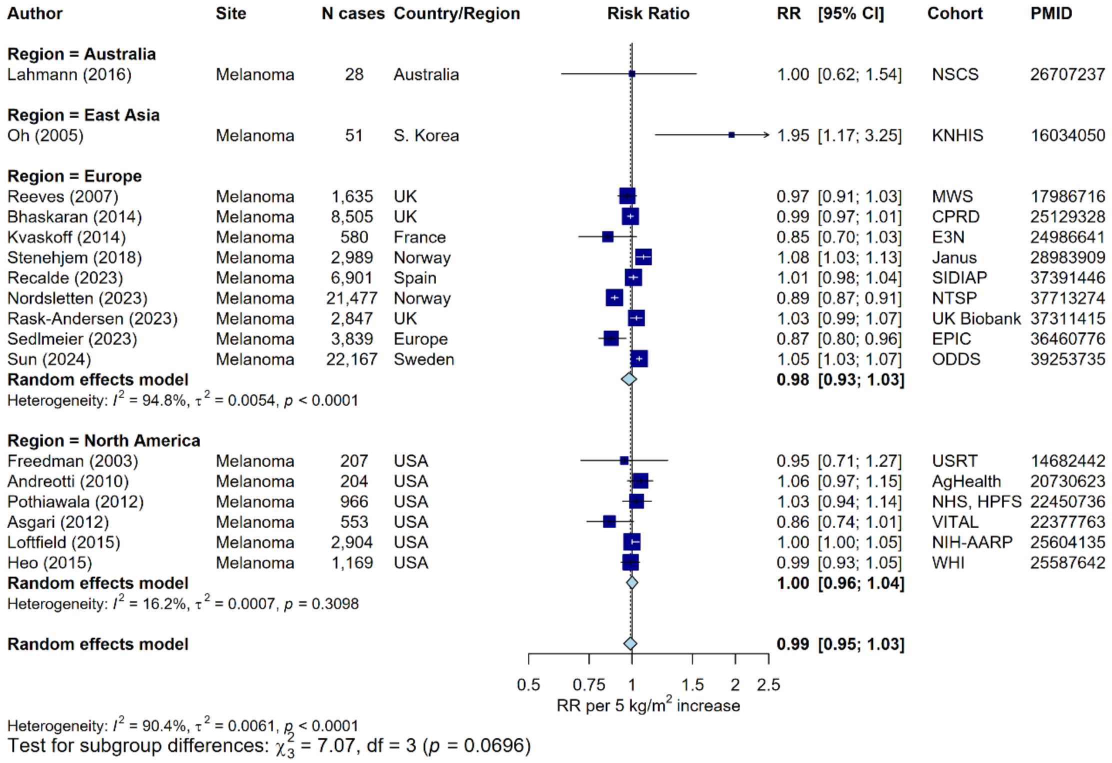

#### Supplementary Figure 49: Meta-analysis of prospective studies on the risk of melanoma in relation to BMI, by region

Study-specific RRs are represented by squares (with their 95% CIs as lines). RRs were combined using random effects meta-analysis (represented by a diamond). Heterogeneity across studies was assessed using $I^2$ and $tau^2$, within region and across all regions. Heterogeneity in the pooled estimates by region was assessed using the Q-statistic. Further details of model adjustments, follow-up time, analytic population for each study are available from **Supplementary Data**.

Abbreviations: BMI=body mass index, CI=confidence interval, PMID=PubMed ID, RR=risk ratio

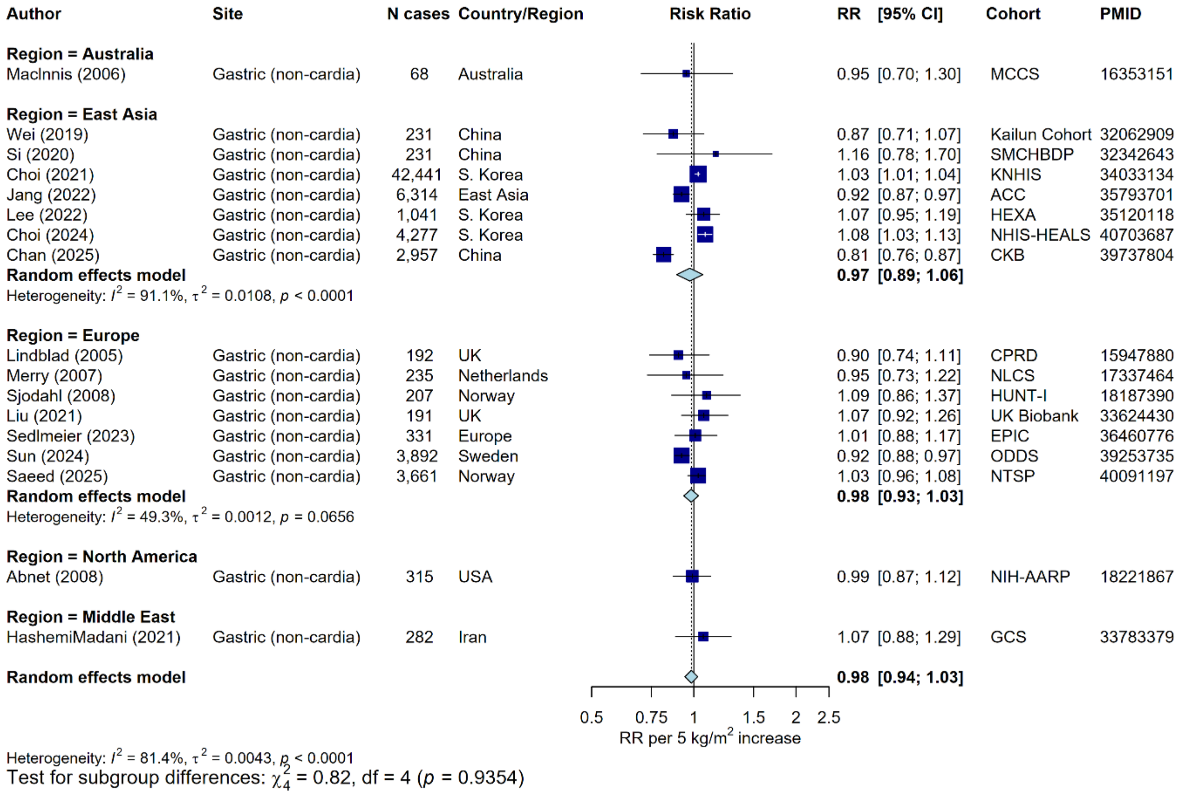

#### Supplementary Figure 50: Meta-analysis of prospective studies on the risk of gastric cancer (non-cardia) in relation to BMI, by region

Study-specific RRs are represented by squares (with their 95% CIs as lines). RRs were combined using random effects meta-analysis (represented by a diamond). Heterogeneity across studies was assessed using $I^2$ and $tau^2$, within region and across all regions. Heterogeneity in the pooled estimates by region was assessed using the Q-statistic. Further details of model adjustments, follow-up time, analytic population for each study are available from **Supplementary Data**.

Abbreviations: BMI=body mass index, CI=confidence interval, PMID=PubMed ID, RR=risk ratio

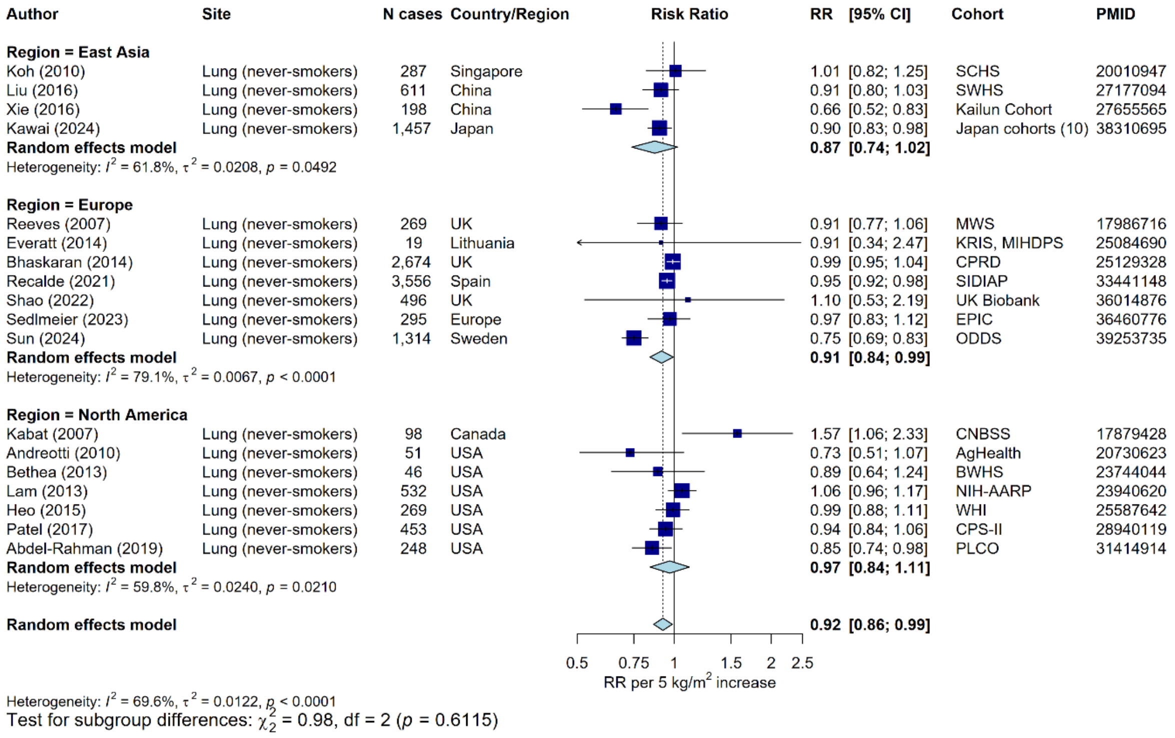

#### Supplementary Figure 51: Meta-analysis of prospective studies on the risk of lung cancer (restricted to never-smokers) in relation to BMI, by region

Study-specific RRs are represented by squares (with their 95% CIs as lines). RRs were combined using random effects meta-analysis (represented by a diamond). Heterogeneity across studies was assessed using $I^2$ and $tau^2$, within region and across all regions. Heterogeneity in the pooled estimates by region was assessed using the Q-statistic. Further details of model adjustments, follow-up time, analytic population for each study are available from **Supplementary Data**.

Abbreviations: BMI=body mass index, CI=confidence interval, PMID=PubMed ID, RR=risk ratio

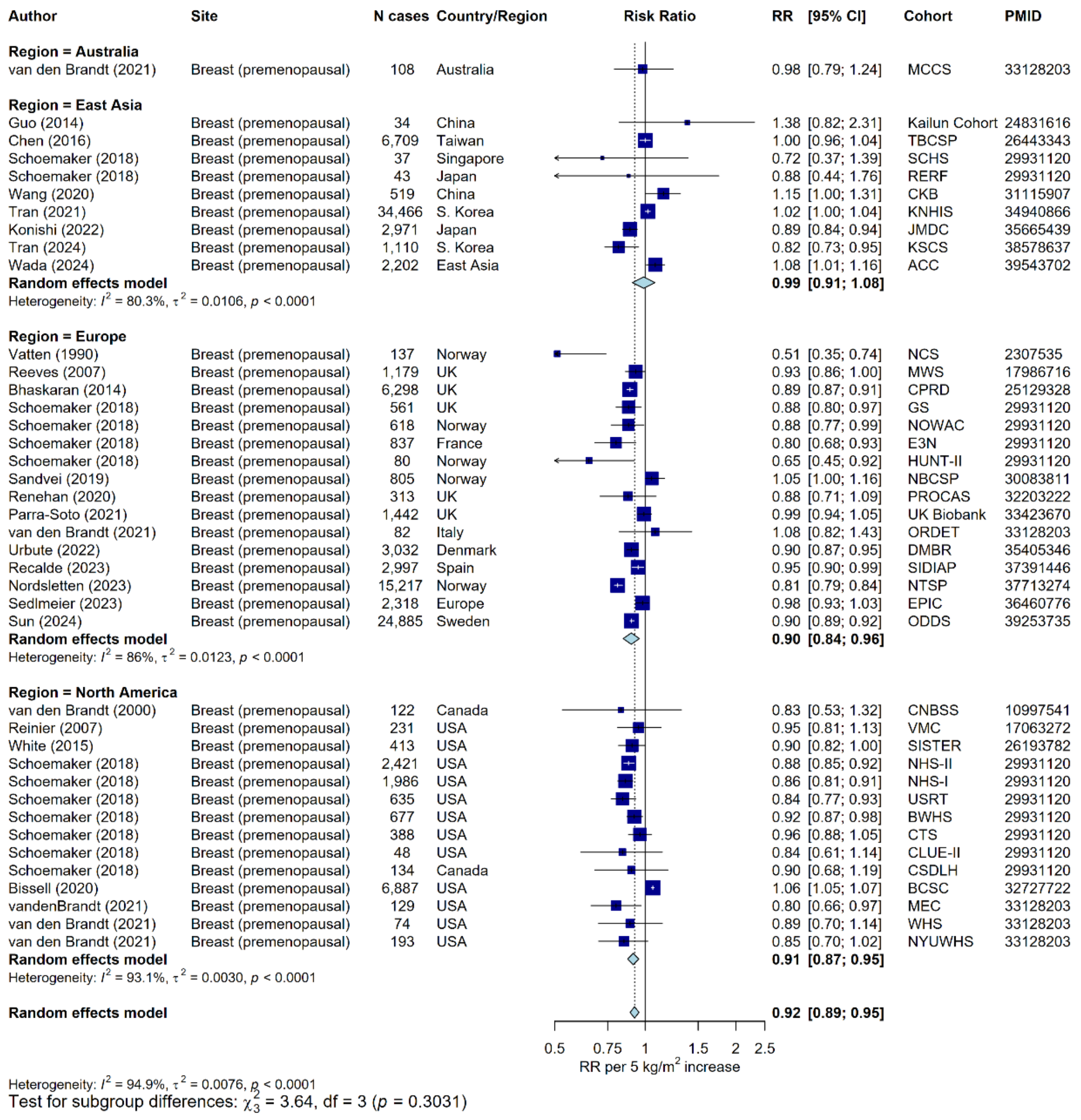

#### Supplementary Figure 52: Meta-analysis of prospective studies on the risk of breast cancer (premenopausal) in relation to BMI, by region

Study-specific RRs are represented by squares (with their 95% CIs as lines). RRs were combined using random effects meta-analysis (represented by a diamond). Heterogeneity across studies was assessed using $I^2$ and $tau^2$, within region and across all regions. Heterogeneity in the pooled estimates by region was assessed using the Q-statistic. Further details of model adjustments, follow-up time, analytic population for each study are available from **Supplementary Data**.

Abbreviations: BMI=body mass index, CI=confidence interval, PMID=PubMed ID, RR=risk ratio

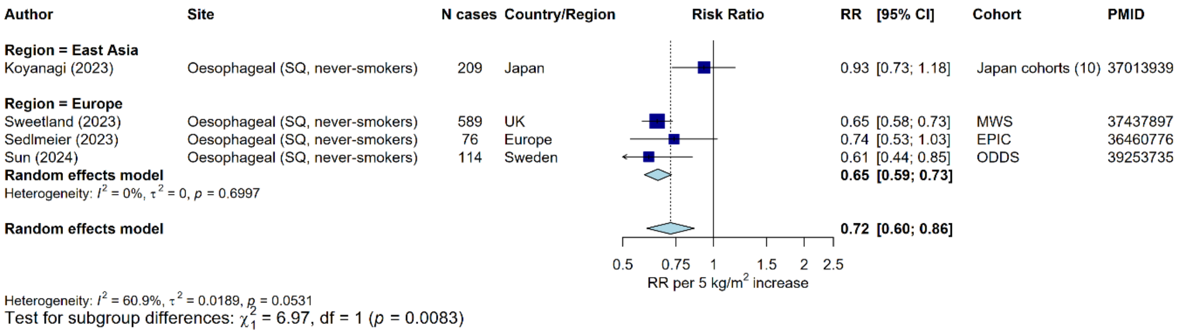

#### Supplementary Figure 53: Meta-analysis of prospective studies on the risk of oesophageal SQ in relation to BMI, by region

Study-specific RRs are represented by squares (with their 95% CIs as lines). RRs were combined using random effects meta-analysis (represented by a diamond). Heterogeneity across studies was assessed using $I^2$ and tau$^2$, within region and across all regions. Heterogeneity in the pooled estimates by region was assessed using the Q-statistic. Further details of model adjustments, follow-up time, analytic population for each study are available from **Supplementary Data**.

Abbreviations: BMI=body mass index, CI=confidence interval, PMID=PubMed ID, RR=risk ratio, SQ=squamous cell carcinoma.

| Author | Site | Adiposity measure | N cases | Country | RR | [95% CI] | Cohort | PMID |
|---|---|---|---|---|---|---|---|---|
| **Exposure = BMI** | | | | | | | | |
| Heo (2015) | Endometrial | BMI | 1,115 | USA | 1.45 | [1.38; 1.52] | WHI | 25587642 |
| Liu (2016) | Endometrial | BMI | 234 | China | 1.55 | [1.39; 1.73] | SWHS | 27177094 |
| Wang (2020) | Endometrial | BMI | 275 | China | 1.60 | [1.44; 1.78] | CKB | 31115907 |
| Recalde (2021) | Endometrial | BMI | 761 | Spain | 1.60 | [1.50; 1.71] | SIDIAP | 33441148 |
| Rask-Andersen (2023) | Endometrial | BMI | 1,291 | UK | 1.78 | [1.68; 1.88] | UK Biobank | 37311415 |
| Sun (2025) | Endometrial | BMI | 889 | Sweden | 1.52 | [1.44; 1.60] | ODDS | 40156135 |
| **Random effects model** | | | | | **1.58** | **[1.49; 1.67]** | | |
| Heterogeneity: $I^2$ = 83.9%, $\tau^2$ = 0.0039, $p$ < 0.0001 | | | | | | | | |
| **Exposure = WC** | | | | | | | | |
| Heo (2015) | Endometrial | WC | 1,115 | USA | 1.42 | [1.35; 1.50] | WHI | 25587642 |
| Liu (2016) | Endometrial | WC | 234 | China | 1.55 | [1.37; 1.76] | SWHS | 27177094 |
| Wang (2020) | Endometrial | WC | 275 | China | 1.67 | [1.48; 1.89] | CKB | 31115907 |
| Recalde (2021) | Endometrial | WC | 761 | Spain | 1.52 | [1.37; 1.69] | SIDIAP | 33441148 |
| Rask-Andersen (2023) | Endometrial | WC | 1,297 | UK | 1.74 | [1.64; 1.84] | UK Biobank | 37311415 |
| Sun (2025) | Endometrial | WC | 889 | Sweden | 1.50 | [1.41; 1.60] | ODDS | 40156135 |
| **Random effects model** | | | | | **1.56** | **[1.46; 1.66]** | | |
| Heterogeneity: $I^2$ = 82.6%, $\tau^2$ = 0.0043, $p$ < 0.0001 | | | | | | | | |

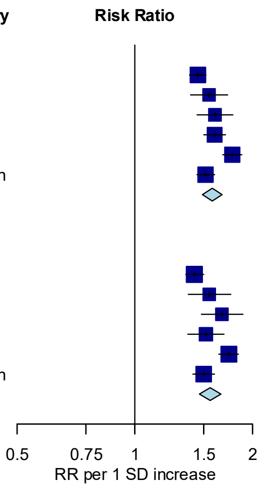

P-value for heterogeneity = 0.44

#### Supplementary Figure 54: Associations of BMI, waist circumference and endometrial cancer risk, per 1 SD increase

Study-specific RRs are represented by squares (with their 95% CIs as lines). RRs were combined using random effects meta-analysis (represented by a diamond). Heterogeneity across studies was assessed using $I^2$ and $tau^2$, within adiposity measure. Heterogeneity in the pooled estimates by adiposity measurement was assessed using the Wald statistic.

Abbreviations: BMI=body mass index, CI=confidence interval, PMID=PubMed ID, RR=risk ratio, SD=standard deviation, WC=waist circumference.

| Author | Site | Adiposity measure | N cases | Country | Risk Ratio | RR | [95% CI] | Cohort | PMID |
|---|---|---|---|---|---|---|---|---|---|
| **Exposure = BMI** | | | | | | | | | |
| Rask-Andersen (2023) | Oesophageal (adeno) | BMI | 730 | UK | | 1.44 | [1.34; 1.55] | UK Biobank | 37311415 |
| Sun (2025) | Oesophageal (adeno) | BMI | 189 | Sweden | | 1.18 | [1.04; 1.35] | ODDS | 40156135 |
| **Random effects model** | | | | | | **1.31** | **[1.08; 1.60]** | | |
| Heterogeneity: $I^2$ = 85.7%, $\tau^2$ = 0.0176, $p$ = 0.0082 | | | | | | | | | |
| **Exposure = WC** | | | | | | | | | |
| Rask-Andersen (2023) | Oesophageal (adeno) | WC | 732 | UK | | 1.47 | [1.37; 1.58] | UK Biobank | 37311415 |
| Sun (2025) | Oesophageal (adeno) | WC | 189 | Sweden | | 1.23 | [1.07; 1.41] | ODDS | 40156135 |
| **Random effects model** | | | | | | **1.36** | **[1.14; 1.62]** | | |
| Heterogeneity: $I^2$ = 80.5%, $\tau^2$ = 0.0129, $p$ = 0.0234 | | | | | | | | | |

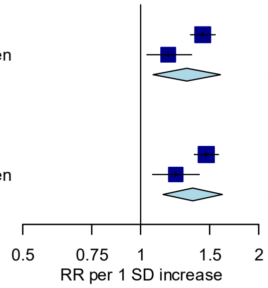

P-value for heterogeneity = 0.48

#### Supplementary Figure 55: Associations of BMI, waist circumference and oesophageal adenocarcinoma risk, per 1 SD increase

Study-specific RRs are represented by squares (with their 95% CIs as lines). RRs were combined using random effects meta-analysis (represented by a diamond). Heterogeneity across studies was assessed using $I^2$ and $tau^2$, within adiposity measure. Heterogeneity in the pooled estimates by adiposity measurement was assessed using the Wald statistic.

Abbreviations: BMI=body mass index, CI=confidence interval, PMID=PubMed ID, RR=risk ratio, SD=standard deviation, WC=waist circumference.

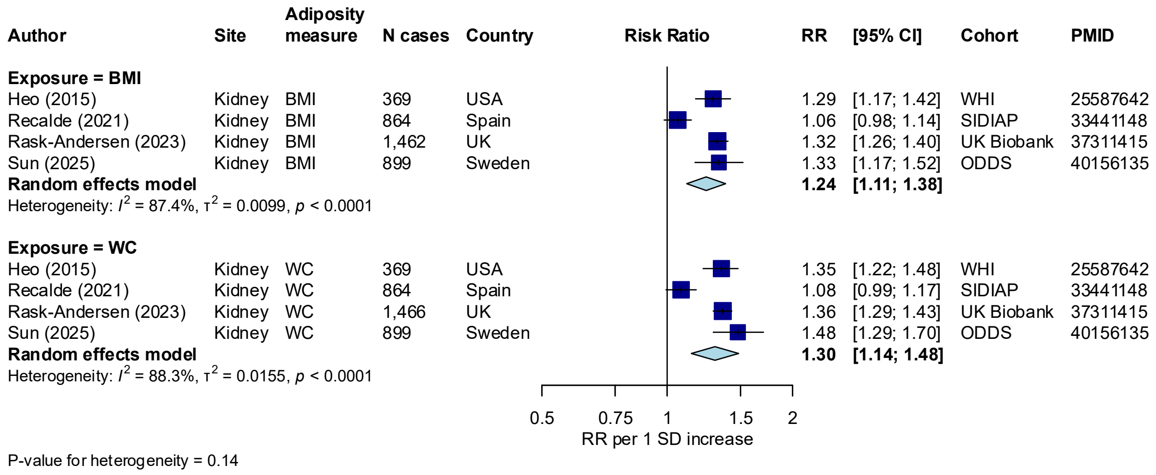

#### Supplementary Figure 56: Associations of BMI, waist circumference and kidney cancer risk, per 1 SD increase

Study-specific RRs are represented by squares (with their 95% CIs as lines). RRs were combined using random effects meta-analysis (represented by a diamond). Heterogeneity across studies was assessed using $I^2$ and $tau^2$, within adiposity measure. Heterogeneity in the pooled estimates by adiposity measurement was assessed using the Wald statistic.

Abbreviations: BMI=Body mass index, CI=confidence interval, PMID=PubMed ID, RR=risk ratio, SD=standard deviation, WC=waist circumference.

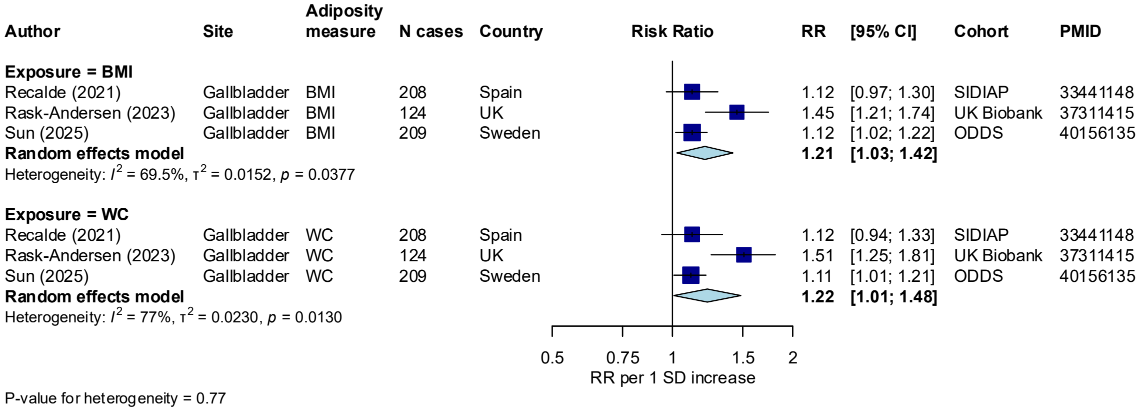

#### Supplementary Figure 57: Associations of BMI, waist circumference and gallbladder cancer risk, per 1 SD increase

Study-specific RRs are represented by squares (with their 95% CIs as lines). RRs were combined using random effects meta-analysis (represented by a diamond). Heterogeneity across studies was assessed using $I^2$ and $tau^2$, within adiposity measure. Heterogeneity in the pooled estimates by adiposity measurement was assessed using the Wald statistic.

Abbreviations: BMI=Body mass index, CI=confidence interval, PMID=PubMed ID, RR=risk ratio, SD=standard deviation, WC=waist circumference.

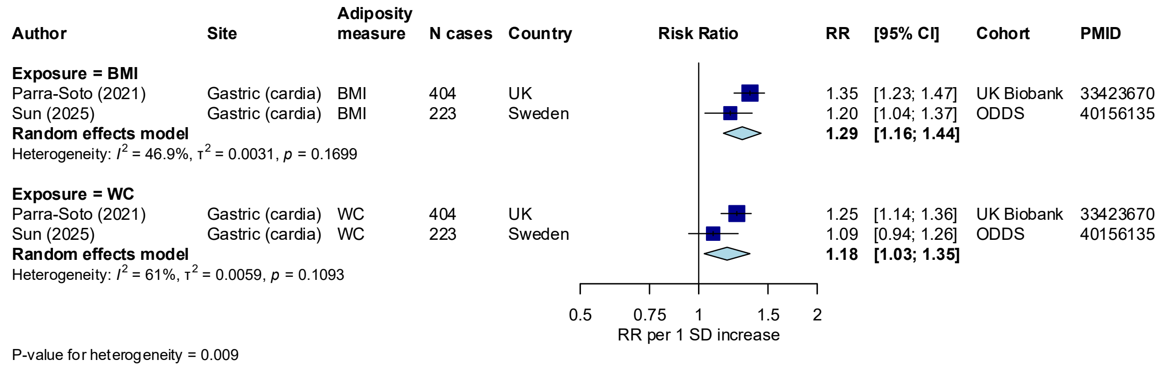

#### Supplementary Figure 58: Associations of BMI, waist circumference and gastric cancer (cardia) risk, per 1 SD increase

Study-specific RRs are represented by squares (with their 95% CIs as lines). RRs were combined using random effects meta-analysis (represented by a diamond). Heterogeneity across studies was assessed using $I^2$ and $tau^2$, within adiposity measure. Heterogeneity in the pooled estimates by adiposity measurement was assessed using the Wald statistic.

Abbreviations: BMI=Body mass index, CI=confidence interval, PMID=PubMed ID, RR=risk ratio, SD=standard deviation, WC=waist circumference.

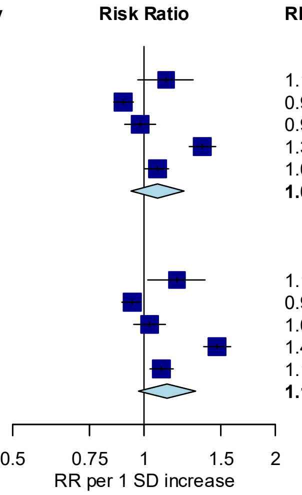

| Author | Site | Adiposity measure | N cases | Country | RR | [95% CI] | Cohort | PMID |
|---|---|---|---|---|---|---|---|---|
| **Exposure = BMI** | | | | | | | | |
| Liu (2016) | Liver | BMI | 165 | China | 1.12 | [0.97; 1.30] | SWHS | 27177094 |
| Wang (2020) | Liver | BMI | 1,861 | China | 0.90 | [0.85; 0.95] | CKB | 31115907 |
| Recalde (2021) | Liver | BMI | 813 | Spain | 0.98 | [0.90; 1.06] | SIDIAP | 33441148 |
| Rask-Andersen (2023) | Liver | BMI | 810 | UK | 1.36 | [1.27; 1.46] | UK Biobank | 37311415 |
| Sun (2025) | Liver | BMI | 458 | Sweden | 1.07 | [1.00; 1.14] | ODDS | 40156135 |
| **Random effects model** | | | | | **1.07** | **[0.93; 1.24]** | | |
| Heterogeneity: $I^2 = 95.7\%$, $\tau^2 = 0.0236$, $p < 0.0001$ | | | | | | | | |
| **Exposure = WC** | | | | | | | | |
| Liu (2016) | Liver | WC | 165 | China | 1.19 | [1.02; 1.38] | SWHS | 27177094 |
| Wang (2020) | Liver | WC | 1,861 | China | 0.94 | [0.89; 0.98] | CKB | 31115907 |
| Recalde (2021) | Liver | WC | 813 | Spain | 1.03 | [0.95; 1.12] | SIDIAP | 33441148 |
| Rask-Andersen (2023) | Liver | WC | 812 | UK | 1.47 | [1.37; 1.58] | UK Biobank | 37311415 |
| Sun (2025) | Liver | WC | 458 | Sweden | 1.10 | [1.03; 1.17] | ODDS | 40156135 |
| **Random effects model** | | | | | **1.13** | **[0.97; 1.31]** | | |
| Heterogeneity: $I^2 = 96.3\%$, $\tau^2 = 0.0276$, $p < 0.0001$ | | | | | | | | |

P-value for heterogeneity = 0.19

**Supplementary Figure 59: Associations of BMI, waist circumference and liver cancer risk, per 1 SD increase**

Study-specific RRs are represented by squares (with their 95% CIs as lines). RRs were combined using random effects meta-analysis (represented by a diamond). Heterogeneity across studies was assessed using $I^2$ and $tau^2$, within adiposity measure. Heterogeneity in the pooled estimates by adiposity measurement was assessed using the Wald statistic.

Abbreviations: BMI=Body mass index, CI=confidence interval, PMID=PubMed ID, RR=risk ratio, SD=standard deviation, WC=waist circumference.

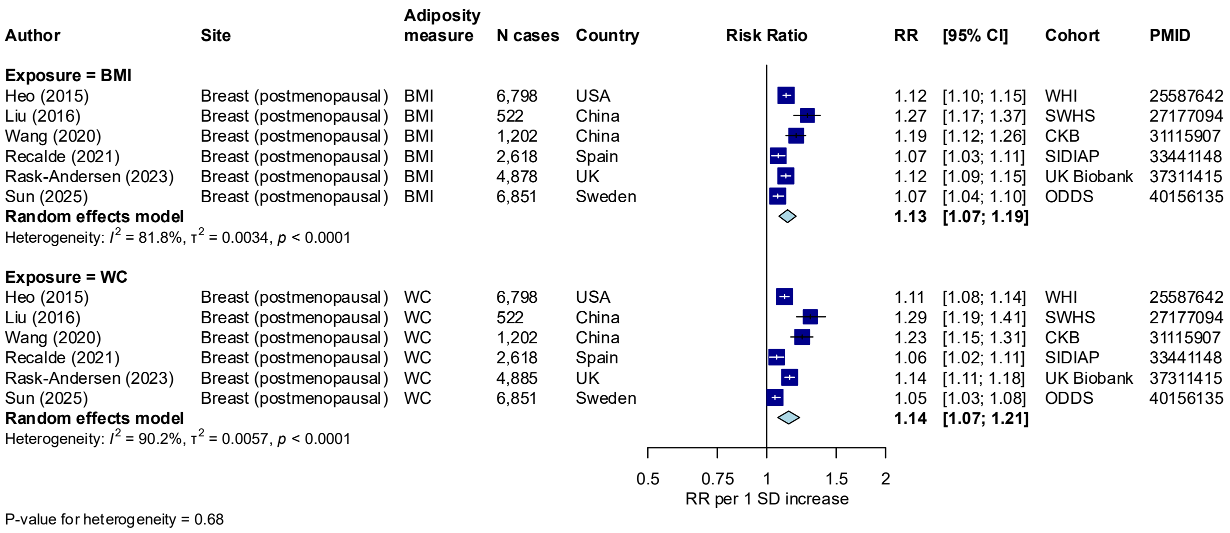

#### Supplementary Figure 60: Associations of BMI, waist circumference and breast (postmenopausal) cancer risk, per 1 SD increase

Study-specific RRs are represented by squares (with their 95% CIs as lines). RRs were combined using random effects meta-analysis (represented by a diamond). Heterogeneity across studies was assessed using $I^2$ and $tau^2$, within adiposity measure. Heterogeneity in the pooled estimates by adiposity measurement was assessed using the Wald statistic.

Abbreviations: BMI=Body mass index, CI=confidence interval, PMID=PubMed ID, RR=risk ratio, SD=standard deviation, WC=waist circumference.

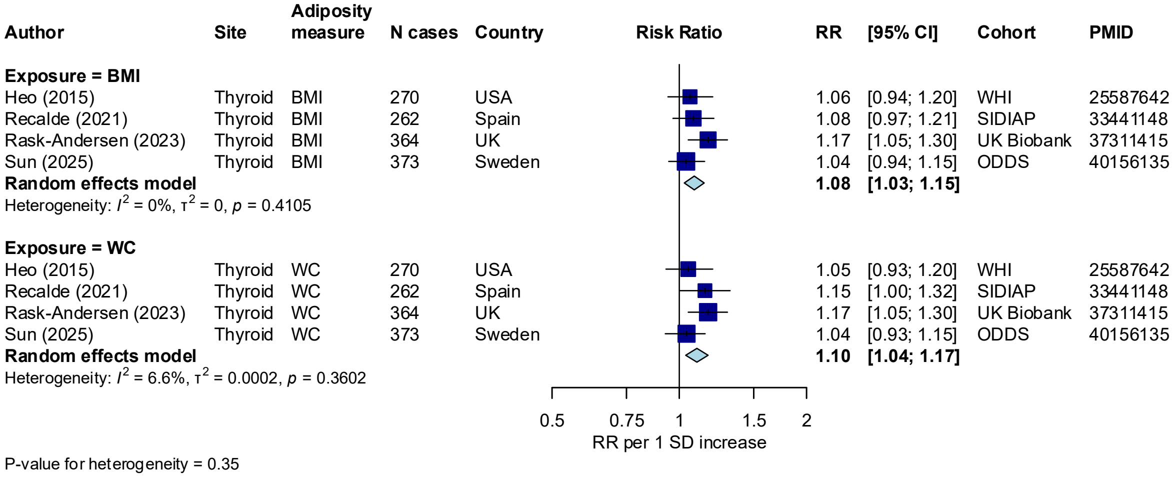

#### Supplementary Figure 61: Associations of BMI, waist circumference and thyroid cancer risk, per 1 SD increase

Study-specific RRs are represented by squares (with their 95% CIs as lines). RRs were combined using random effects meta-analysis (represented by a diamond). Heterogeneity across studies was assessed using $I^2$ and $tau^2$, within adiposity measure. Heterogeneity in the pooled estimates by adiposity measurement was assessed using the Wald statistic.

Abbreviations: BMI=Body mass index, CI=confidence interval, PMID=PubMed ID, RR=risk ratio, SD=standard deviation, WC=waist circumference.

| Author | Site | Adiposity measure | N cases | Country | Risk Ratio | RR | [95% CI] | Cohort | PMID |
|---|---|---|---|---|---|---|---|---|---|
| **Exposure = BMI** | | | | | | | | | |
| Sun (2025) | Meningioma | BMI | 532 | Sweden | | 1.11 | [1.01; 1.23] | ODDS | 40156135 |
| **Exposure = WC** | | | | | | | | | |
| Sun (2025) | Meningioma | WC | 532 | Sweden | | 1.13 | [1.01; 1.25] | ODDS | 40156135 |

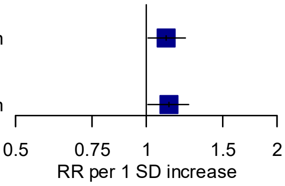

P-value for heterogeneity = 0.59

#### Supplementary Figure 62: Associations of BMI, waist circumference and meningioma risk, per 1 SD increase

Study-specific RRs are represented by squares (with their 95% CIs as lines). RRs were combined using random effects meta-analysis (represented by a diamond). Heterogeneity across studies was assessed using $I^2$ and $tau^2$, within adiposity measure. Heterogeneity in the pooled estimates by adiposity measurement was assessed using the Wald statistic.

Abbreviations: BMI=Body mass index, CI=confidence interval, PMID=PubMed ID, RR=risk ratio, SD=standard deviation, WC=waist circumference.

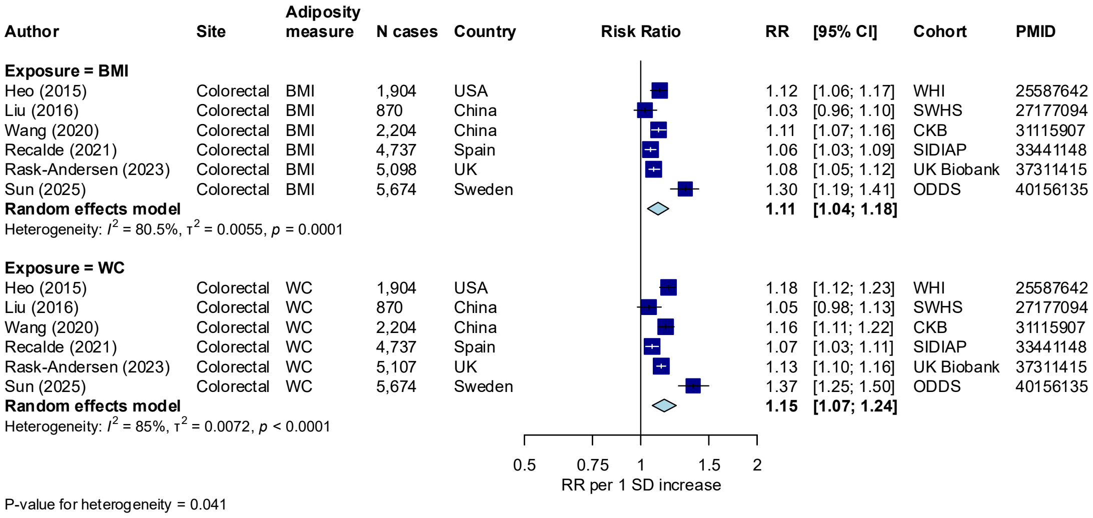

#### Supplementary Figure 63: Associations of BMI, waist circumference and colorectal cancer risk, per 1 SD increase

Study-specific RRs are represented by squares (with their 95% CIs as lines). RRs were combined using random effects meta-analysis (represented by a diamond). Heterogeneity across studies was assessed using $I^2$ and $tau^2$, within adiposity measure. Heterogeneity in the pooled estimates by adiposity measurement was assessed using the Wald statistic.

Abbreviations: BMI=Body mass index, CI=confidence interval, PMID=PubMed ID, RR=risk ratio, SD=standard deviation, WC=waist circumference.

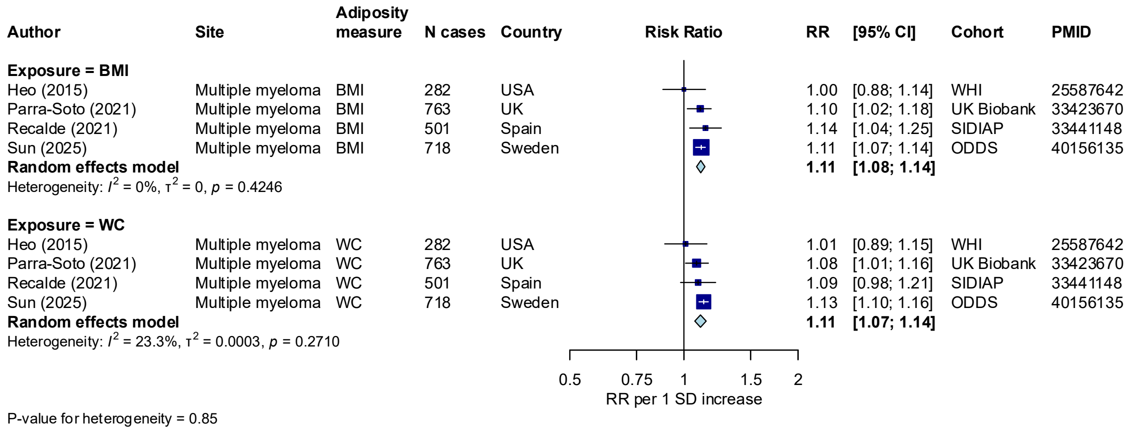

#### Supplementary Figure 64: Associations of BMI, waist circumference and multiple myeloma risk, per 1 SD increase

Study-specific RRs are represented by squares (with their 95% CIs as lines). RRs were combined using random effects meta-analysis (represented by a diamond). Heterogeneity across studies was assessed using $I^2$ and $tau^2$, within adiposity measure. Heterogeneity in the pooled estimates by adiposity measurement was assessed using the Wald statistic.

Abbreviations: BMI=Body mass index, CI=confidence interval, PMID=PubMed ID, RR=risk ratio, SD=standard deviation, WC=waist circumference.

| Author | Site | Adiposity measure | N cases | Country | RR | [95% CI] | Cohort | PMID |
|---|---|---|---|---|---|---|---|---|
| **Exposure = BMI** | | | | | | | | |
| Heo (2015) | Leukemia | BMI | 447 | USA | 1.07 | [0.97; 1.08] | WHI | 25587642 |
| Wang (2020) | Leukemia | BMI | 443 | China | 1.04 | [0.95; 1.15] | CKB | 31115907 |
| Parra-Soto (2021) | Leukemia | BMI | 1,129 | UK | 1.07 | [1.01; 1.14] | UK Biobank | 33423670 |
| Recalde (2021) | Leukemia | BMI | 969 | Spain | 1.04 | [0.97; 1.12] | SIDIAP | 33441148 |
| **Random effects model** | | | | | **1.06** | **[1.03; 1.10]** | | |
| Heterogeneity: $I^2 = 0\%$, $\tau^2 = 0$, $p = 0.8788$ | | | | | | | | |
| **Exposure = WC** | | | | | | | | |
| Heo (2015) | Leukemia | WC | 447 | USA | 1.06 | [0.96; 1.16] | WHI | 25587642 |
| Wang (2020) | Leukemia | WC | 443 | China | 1.03 | [0.93; 1.14] | CKB | 31115907 |
| Parra-Soto (2021) | Leukemia | WC | 1,129 | UK | 1.08 | [1.02; 1.15] | UK Biobank | 33423670 |
| Recalde (2021) | Leukemia | WC | 969 | Spain | 1.09 | [1.01; 1.18] | SIDIAP | 33441148 |
| **Random effects model** | | | | | **1.07** | **[1.03; 1.11]** | | |
| Heterogeneity: $I^2 = 0\%$, $\tau^2 = 0$, $p = 0.8314$ | | | | | | | | |

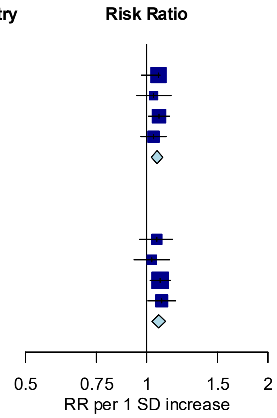

P-value for heterogeneity = 0.33

#### Supplementary Figure 65: Associations of BMI, waist circumference and leukaemia risk, per 1 SD increase

Study-specific RRs are represented by squares (with their 95% CIs as lines). RRs were combined using random effects meta-analysis (represented by a diamond). Heterogeneity across studies was assessed using $I^2$ and $tau^2$, within adiposity measure. Heterogeneity in the pooled estimates by adiposity measurement was assessed using the Wald statistic.

Abbreviations: BMI=Body mass index, CI=confidence interval, PMID=PubMed ID, RR=risk ratio, SD=standard deviation, WC=waist circumference.

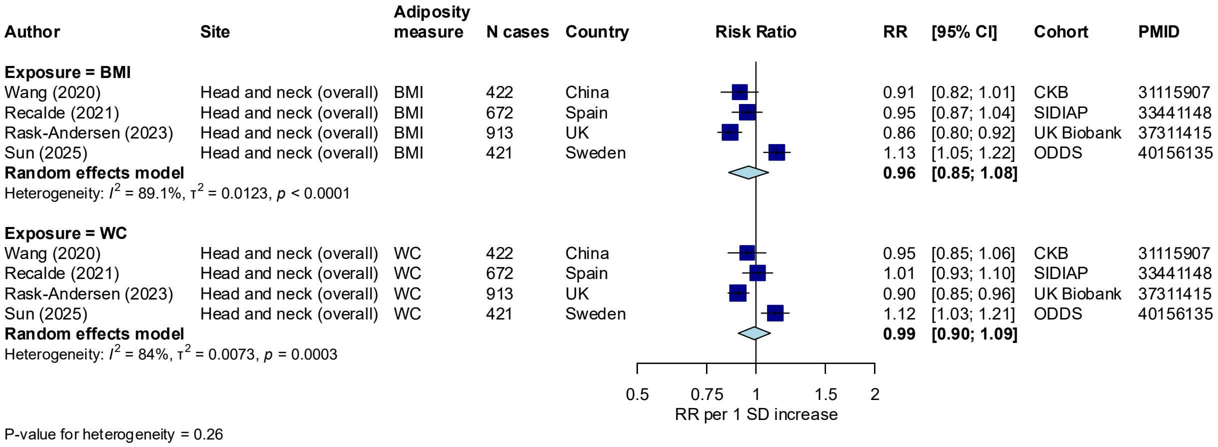

#### Supplementary Figure 66: Associations of BMI, waist circumference and head and neck cancer risk, per 1 SD increase

Study-specific RRs are represented by squares (with their 95% CIs as lines). RRs were combined using random effects meta-analysis (represented by a diamond). Heterogeneity across studies was assessed using $I^2$ and $tau^2$, within adiposity measure. Heterogeneity in the pooled estimates by adiposity measurement was assessed using the Wald statistic.

Abbreviations: BMI=Body mass index, CI=confidence interval, PMID=PubMed ID, RR=risk ratio, SD=standard deviation, WC=waist circumference.

| Author | Site | Adiposity measure | N cases | Country | RR | [95% CI] | Cohort | PMID |
|---|---|---|---|---|---|---|---|---|
| **Exposure = BMI** | | | | | | | | |
| Heo (2015) | Pancreas | BMI | 459 | USA | 1.03 | [0.93; 1.14] | WHI | 25587642 |
| Wang (2020) | Pancreas | BMI | 522 | China | 1.05 | [0.96; 1.14] | CKB | 31115907 |
| Recalde (2021) | Pancreas | BMI | 641 | Spain | 0.99 | [0.91; 1.08] | SIDIAP | 33441148 |
| Rask-Andersen (2023) | Pancreas | BMI | 1,302 | UK | 1.16 | [1.10; 1.23] | UK Biobank | 37311415 |
| Sun (2025) | Pancreas | BMI | 1,069 | Sweden | 1.24 | [1.16; 1.32] | ODDS | 40156135 |
| **Random effects model** | | | | | **1.10** | **[1.01; 1.19]** | | |
| Heterogeneity: $I^2 = 82.6\%$, $\tau^2 = 0.0071$, $p = 0.0001$ | | | | | | | | |
| **Exposure = WC** | | | | | | | | |
| Heo (2015) | Pancreas | WC | 459 | USA | 1.04 | [0.94; 1.15] | WHI | 25587642 |
| Wang (2020) | Pancreas | WC | 522 | China | 1.04 | [0.95; 1.14] | CKB | 31115907 |
| Recalde (2021) | Pancreas | WC | 641 | Spain | 1.05 | [0.96; 1.15] | SIDIAP | 33441148 |
| Rask-Andersen (2023) | Pancreas | WC | 1,306 | UK | 1.17 | [1.11; 1.24] | UK Biobank | 37311415 |
| Sun (2025) | Pancreas | WC | 1,069 | Sweden | 1.29 | [1.21; 1.38] | ODDS | 40156135 |
| **Random effects model** | | | | | **1.12** | **[1.03; 1.22]** | | |
| Heterogeneity: $I^2 = 83.9\%$, $\tau^2 = 0.0077$, $p < 0.0001$ | | | | | | | | |

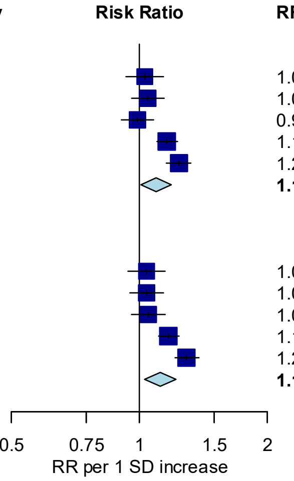

P-value for heterogeneity = 0.30

#### Supplementary Figure 67: Associations of BMI, waist circumference and pancreatic cancer risk, per 1 SD increase

Study-specific RRs are represented by squares (with their 95% CIs as lines). RRs were combined using random effects meta-analysis (represented by a diamond). Heterogeneity across studies was assessed using $I^2$ and $tau^2$, within adiposity measure. Heterogeneity in the pooled estimates by adiposity measurement was assessed using the Wald statistic.

Abbreviations: BMI=Body mass index, CI=confidence interval, PMID=PubMed ID, RR=risk ratio, SD=standard deviation, WC=waist circumference.

| Author | Site | Adiposity measure | N cases | Country | RR | [95% CI] | Cohort | PMID |
|---|---|---|---|---|---|---|---|---|
| **Exposure = BMI** | | | | | | | | |
| Heo (2015) | NHL | BMI | 888 | USA | 1.04 | [0.97; 1.12] | WHI | 25587642 |
| Parra-Soto (2021) | NHL | BMI | 1,681 | UK | 1.03 | [0.98; 1.09] | UK Biobank | 33423670 |
| Recalde (2021) | NHL | BMI | 863 | Spain | 1.04 | [0.97; 1.12] | SIDIAP | 33441148 |
| **Random effects model** | | | | | **1.04** | **[1.00; 1.07]** | | |
| Heterogeneity: $I^2$ = 0%, $\tau^2$ = 0, $p$ = 0.9676 | | | | | | | | |
| **Exposure = WC** | | | | | | | | |
| Heo (2015) | NHL | WC | 888 | USA | 1.07 | [0.99; 1.14] | WHI | 25587642 |
| Parra-Soto (2021) | NHL | WC | 1,681 | UK | 1.05 | [1.00; 1.11] | UK Biobank | 33423670 |
| Recalde (2021) | NHL | WC | 863 | Spain | 1.03 | [0.95; 1.11] | SIDIAP | 33441148 |
| **Random effects model** | | | | | **1.05** | **[1.01; 1.09]** | | |
| Heterogeneity: $I^2$ = 0%, $\tau^2$ = 0, $p$ = 0.7714 | | | | | | | | |

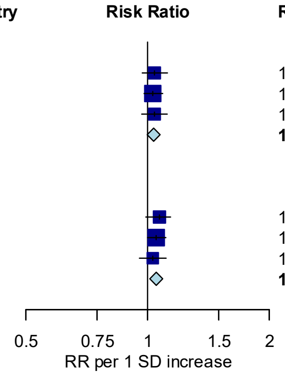

P-value for heterogeneity = 0.12

#### Supplementary Figure 68: Associations of BMI, waist circumference and NHL risk, per 1 SD increase

Study-specific RRs are represented by squares (with their 95% CIs as lines). RRs were combined using random effects meta-analysis (represented by a diamond). Heterogeneity across studies was assessed using $I^2$ and $tau^2$, within adiposity measure. Heterogeneity in the pooled estimates by adiposity measurement was assessed using the Wald statistic.

Abbreviations: BMI=Body mass index, CI=confidence interval, NHL=non-Hodgkin lymphoma, PMID=PubMed ID, RR=risk ratio, SD=standard deviation, WC=waist circumference.

| Author | Site | Adiposity measure | N cases | Country | RR | [95% CI] | Cohort | PMID |
|---|---|---|---|---|---|---|---|---|
| **Exposure = BMI** | | | | | | | | |
| Heo (2015) | Ovarian | BMI | 702 | USA | 1.04 | [0.95; 1.12] | WHI | 25587642 |
| Liu (2016) | Ovarian | BMI | 170 | China | 1.15 | [0.99; 1.34] | SWHS | 27177094 |
| Wang (2020) | Ovarian | BMI | 264 | China | 1.09 | [0.96; 1.22] | CKB | 31115907 |
| Recalde (2021) | Ovarian | BMI | 247 | Spain | 0.97 | [0.85; 1.11] | SIDIAP | 33441148 |
| Rask-Andersen (2023) | Ovarian | BMI | 958 | UK | 1.07 | [1.00; 1.14] | UK Biobank | 37311415 |
| Sun (2025) | Ovarian | BMI | 858 | Sweden | 1.03 | [0.96; 1.10] | ODDS | 40156135 |
| **Random effects model** | | | | | **1.05** | **[1.01; 1.09]** | | |
| Heterogeneity: $I^2 = 0\%$, $\tau^2 = 0$, $p = 0.5741$ | | | | | | | | |
| **Exposure = WC** | | | | | | | | |
| Heo (2015) | Ovarian | WC | 702 | USA | 1.03 | [0.95; 1.11] | WHI | 25587642 |
| Liu (2016) | Ovarian | WC | 170 | China | 1.16 | [0.99; 1.36] | SWHS | 27177094 |
| Wang (2020) | Ovarian | WC | 264 | China | 1.08 | [0.95; 1.24] | CKB | 31115907 |
| Recalde (2021) | Ovarian | WC | 247 | Spain | 0.99 | [0.87; 1.13] | SIDIAP | 33441148 |
| Rask-Andersen (2023) | Ovarian | WC | 960 | UK | 1.09 | [1.02; 1.16] | UK Biobank | 37311415 |
| Sun (2025) | Ovarian | WC | 858 | Sweden | 1.02 | [0.95; 1.10] | ODDS | 40156135 |
| **Random effects model** | | | | | **1.05** | **[1.01; 1.09]** | | |
| Heterogeneity: $I^2 = 0\%$, $\tau^2 = 0$, $p = 0.4525$ | | | | | | | | |

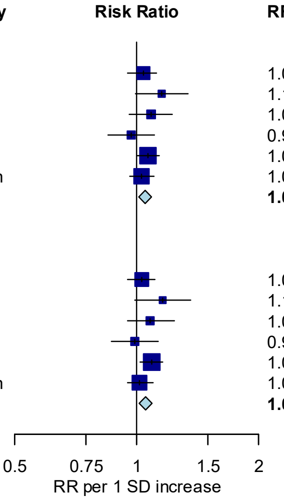

P-value for heterogeneity = 0.78

#### Supplementary Figure 69: Associations of BMI, waist circumference and ovarian cancer risk, per 1 SD increase

Study-specific RRs are represented by squares (with their 95% CIs as lines). RRs were combined using random effects meta-analysis (represented by a diamond). Heterogeneity across studies was assessed using $I^2$ and $tau^2$, within adiposity measure. Heterogeneity in the pooled estimates by adiposity measurement was assessed using the Wald statistic.

Abbreviations: BMI=Body mass index, CI=confidence interval, PMID=PubMed ID, RR=risk ratio, SD=standard deviation, WC=waist circumference.

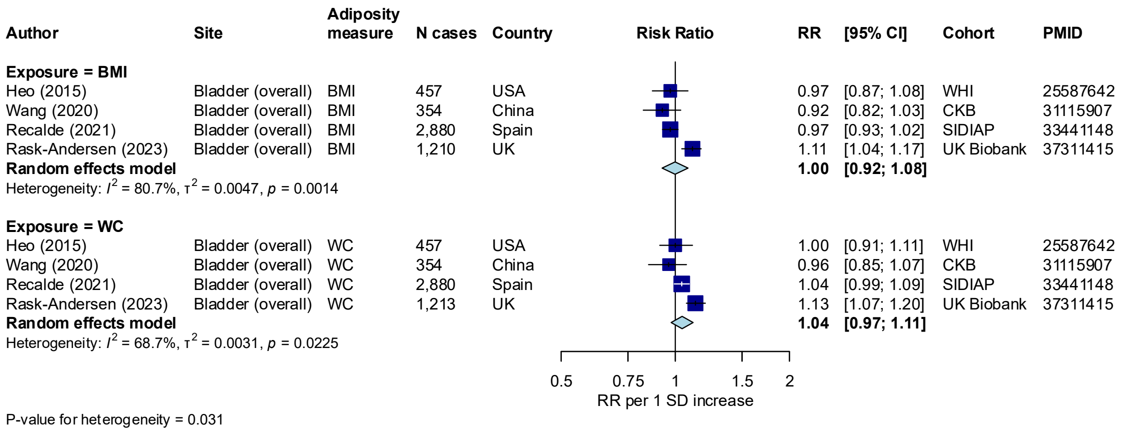

#### Supplementary Figure 70: Associations of BMI, waist circumference and bladder cancer risk, per 1 SD increase

Study-specific RRs are represented by squares (with their 95% CIs as lines). RRs were combined using random effects meta-analysis (represented by a diamond). Heterogeneity across studies was assessed using $I^2$ and $tau^2$, within adiposity measure. Heterogeneity in the pooled estimates by adiposity measurement was assessed using the Wald statistic.

Abbreviations: BMI=Body mass index, CI=confidence interval, PMID=PubMed ID, RR=risk ratio, SD=standard deviation, WC=waist circumference.

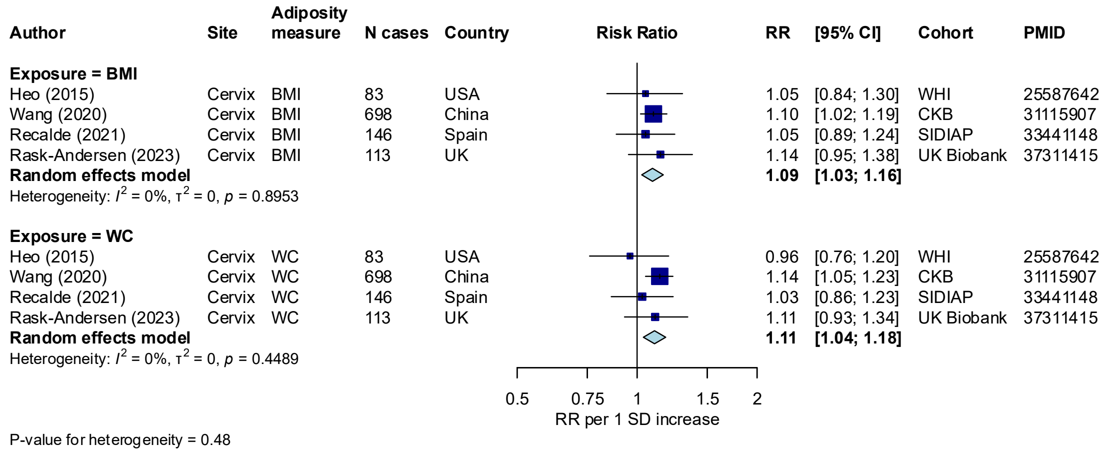

#### Supplementary Figure 71: Associations of BMI, waist circumference and cervical cancer risk, per 1 SD increase

Study-specific RRs are represented by squares (with their 95% CIs as lines). RRs were combined using random effects meta-analysis (represented by a diamond). Heterogeneity across studies was assessed using $I^2$ and $tau^2$, within adiposity measure. Heterogeneity in the pooled estimates by adiposity measurement was assessed using the Wald statistic.

Abbreviations: BMI=Body mass index, CI=confidence interval, PMID=PubMed ID, RR=risk ratio, SD=standard deviation, WC=waist circumference.

| Author | Site | Adiposity measure | N cases | Country | RR | [95% CI] | Cohort | PMID |
|---|---|---|---|---|---|---|---|---|
| **Exposure = BMI** | | | | | | | | |
| Wang (2020) | Prostate (overall) | BMI | 263 | China | 1.11 | [0.97; 1.26] | CKB | 31115907 |
| Recalde (2021) | Prostate (overall) | BMI | 4,094 | Spain | 0.96 | [0.92; 1.00] | SIDIAP | 33441148 |
| Rask-Andersen (2023) | Prostate (overall) | BMI | 9,348 | UK | 0.95 | [0.93; 0.97] | UK Biobank | 37311415 |
| **Random effects model** | | | | | **0.98** | **[0.91; 1.07]** | | |
| Heterogeneity: $I^2$ = 58%, $\tau^2$ = 0.0039, $p$ = 0.0924 | | | | | | | | |
| **Exposure = WC** | | | | | | | | |
| Wang (2020) | Prostate (overall) | WC | 263 | China | 1.13 | [0.99; 1.29] | CKB | 31115907 |
| Recalde (2021) | Prostate (overall) | WC | 4,094 | Spain | 0.98 | [0.95; 1.01] | SIDIAP | 33441148 |
| Rask-Andersen (2023) | Prostate (overall) | WC | 9,361 | UK | 0.96 | [0.95; 0.98] | UK Biobank | 37311415 |
| **Random effects model** | | | | | **1.00** | **[0.92; 1.09]** | | |
| Heterogeneity: $I^2$ = 64.9%, $\tau^2$ = 0.0045, $p$ = 0.0580 | | | | | | | | |

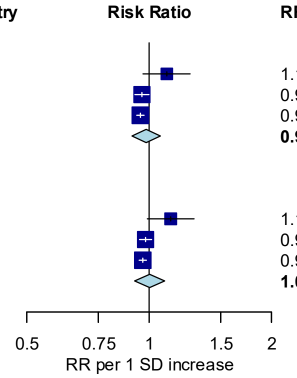

P-value for heterogeneity = 0.36

#### Supplementary Figure 72: Associations of BMI, waist circumference and prostate cancer risk, per 1 SD increase

Study-specific RRs are represented by squares (with their 95% CIs as lines). RRs were combined using random effects meta-analysis (represented by a diamond). Heterogeneity across studies was assessed using $I^2$ and $tau^2$, within adiposity measure. Heterogeneity in the pooled estimates by adiposity measurement was assessed using the Wald statistic.

Abbreviations: BMI=Body mass index, CI=confidence interval, PMID=PubMed ID, RR=risk ratio, SD=standard deviation, WC=waist circumference.

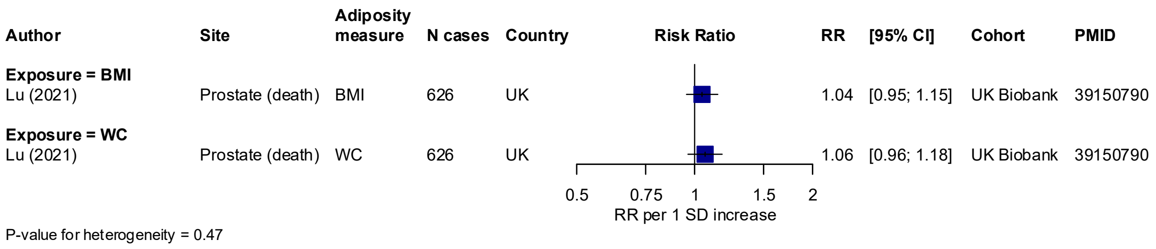

#### Supplementary Figure 73: Associations of BMI, waist circumference and fatal prostate cancer risk, per 1 SD increase

Study-specific RRs are represented by squares (with their 95% CIs as lines). RRs were combined using random effects meta-analysis (represented by a diamond). Heterogeneity across studies was assessed using $I^2$ and $tau^2$, within adiposity measure. Heterogeneity in the pooled estimates by adiposity measurement was assessed using the Wald statistic.

Abbreviations: BMI=Body mass index, CI=confidence interval, PMID=PubMed ID, RR=risk ratio, SD=standard deviation, WC=waist circumference.

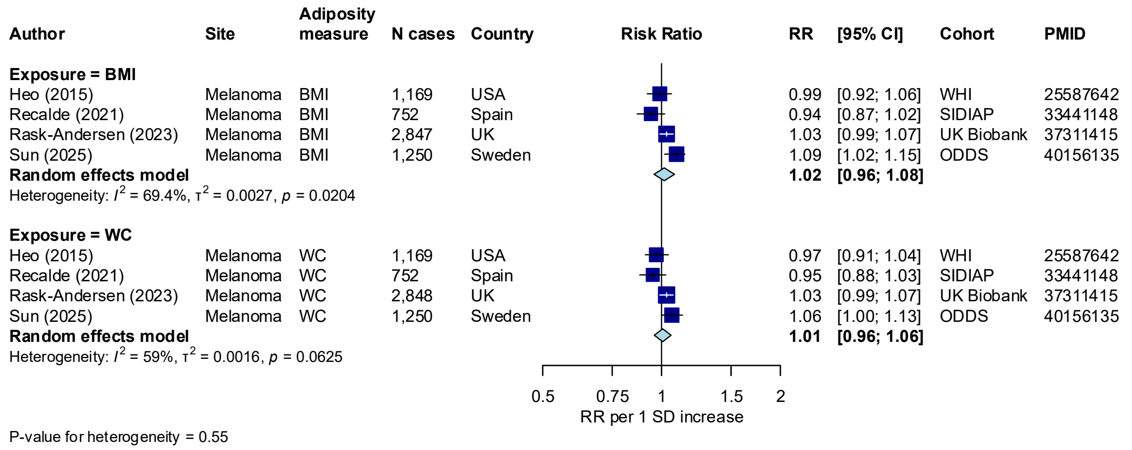

#### Supplementary Figure 74: Associations of BMI, waist circumference and melanoma risk, per 1 SD increase

Study-specific RRs are represented by squares (with their 95% CIs as lines). RRs were combined using random effects meta-analysis (represented by a diamond). Heterogeneity across studies was assessed using $I^2$ and $tau^2$, within adiposity measure. Heterogeneity in the pooled estimates by adiposity measurement was assessed using the Wald statistic.

Abbreviations: BMI=Body mass index, CI=confidence interval, PMID=PubMed ID, RR=risk ratio, SD=standard deviation, WC=waist circumference.

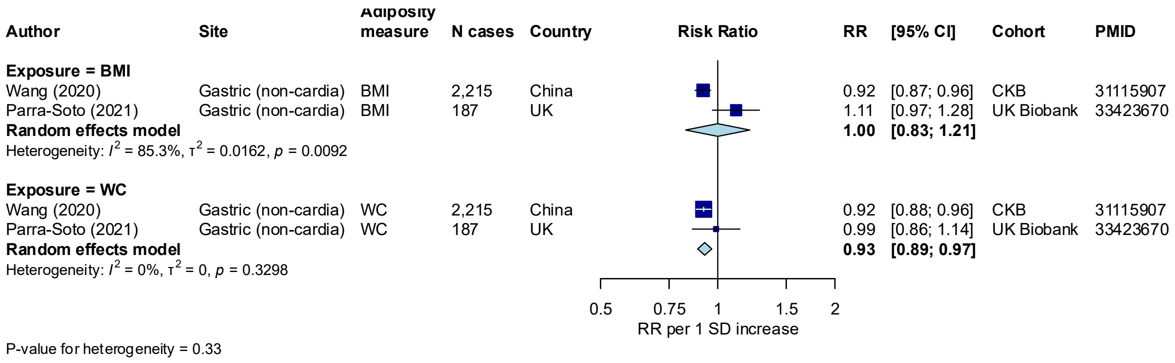

#### Supplementary Figure 75: Associations of BMI, waist circumference and gastric cancer (non-cardia) risk, per 1 SD increase

Study-specific RRs are represented by squares (with their 95% CIs as lines). RRs were combined using random effects meta-analysis (represented by a diamond). Heterogeneity across studies was assessed using $I^2$ and $tau^2$, within adiposity measure. Heterogeneity in the pooled estimates by adiposity measurement was assessed using the Wald statistic.

Abbreviations: BMI=Body mass index, CI=confidence interval, PMID=PubMed ID, RR=risk ratio, SD=standard deviation, WC=waist circumference.

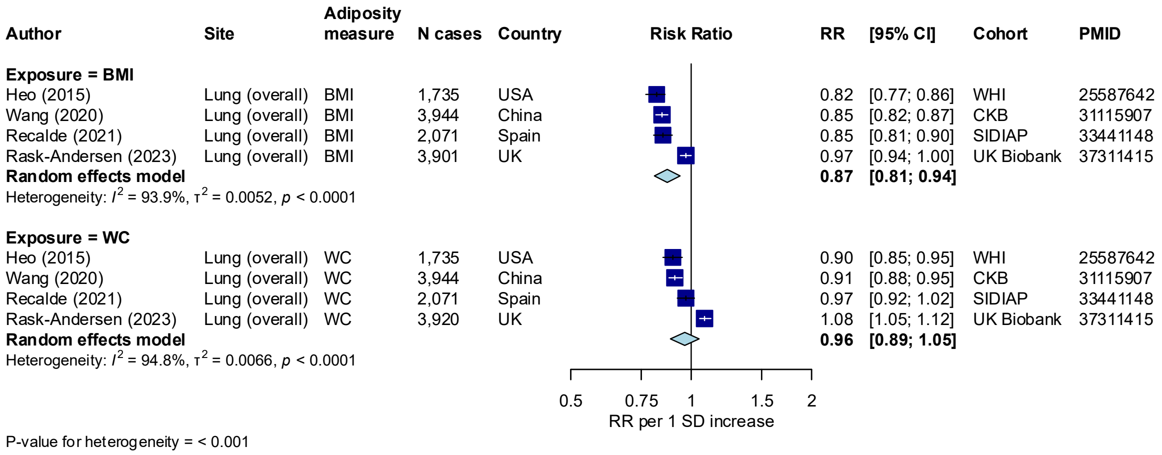

#### Supplementary Figure 76: Associations of BMI, waist circumference and lung cancer risk, per 1 SD increase

Study-specific RRs are represented by squares (with their 95% CIs as lines). RRs were combined using random effects meta-analysis (represented by a diamond). Heterogeneity across studies was assessed using $I^2$ and $tau^2$, within adiposity measure. Heterogeneity in the pooled estimates by adiposity measurement was assessed using the Wald statistic.

Abbreviations: BMI=Body mass index, CI=confidence interval, PMID=PubMed ID, RR=risk ratio, SD=standard deviation, WC=waist circumference.

| Author | Site | Adiposity measure | N cases | Country | RR | [95% CI] | Cohort | PMID |
|---|---|---|---|---|---|---|---|---|
| **Exposure = BMI** | | | | | | | | |
| Heo (2015) | Lung (never-smokers) | BMI | 269 | USA | 0.99 | [0.86; 1.13] | WHI | 25587642 |
| Lu (2021) | Lung (never-smokers) | BMI | 474 | UK | 0.95 | [0.86; 1.04] | UK Biobank | 39150790 |
| **Random effects model** | | | | | **0.96** | **[0.89; 1.04]** | | |
| Heterogeneity: $I^2 = 0\%$, $\tau^2 = 0$, $p = 0.6270$ | | | | | | | | |
| **Exposure = WC** | | | | | | | | |
| Heo (2015) | Lung (never-smokers) | WC | 269 | USA | 1.01 | [0.88; 1.15] | WHI | 25587642 |
| Lu (2021) | Lung (never-smokers) | WC | 474 | UK | 1.00 | [0.89; 1.11] | UK Biobank | 39150790 |
| **Random effects model** | | | | | **1.00** | **[0.92; 1.09]** | | |
| Heterogeneity: $I^2 = 0\%$, $\tau^2 = 0$, $p = 0.9105$ | | | | | | | | |

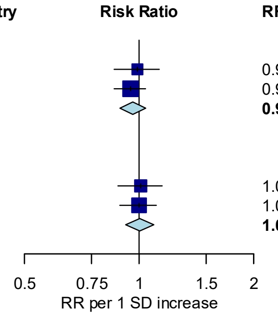

P-value for heterogeneity = 0.052

#### Supplementary Figure 77: Associations of BMI, waist circumference and lung cancer risk (restricted to never-smokers), per 1 SD increase

Study-specific RRs are represented by squares (with their 95% CIs as lines). RRs were combined using random effects meta-analysis (represented by a diamond). Heterogeneity across studies was assessed using $I^2$ and $tau^2$, within adiposity measure. Heterogeneity in the pooled estimates by adiposity measurement was assessed using the Wald statistic.

Abbreviations: BMI=Body mass index, CI=confidence interval, PMID=PubMed ID, RR=risk ratio, SD=standard deviation, WC=waist circumference.

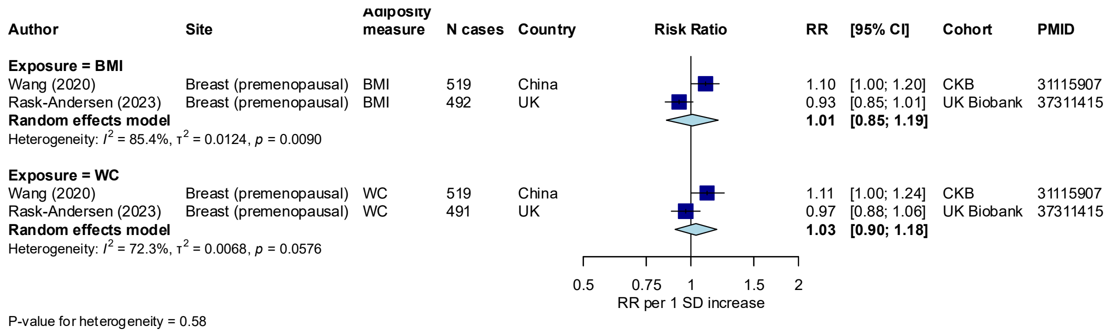

#### Supplementary Figure 78: Associations of BMI, waist circumference and breast (premenopausal) cancer risk, per 1 SD increase

Study-specific RRs are represented by squares (with their 95% CIs as lines). RRs were combined using random effects meta-analysis (represented by a diamond). Heterogeneity across studies was assessed using $I^2$ and $tau^2$, within adiposity measure. Heterogeneity in the pooled estimates by adiposity measurement was assessed using the Wald statistic.

Abbreviations: BMI=Body mass index, CI=confidence interval, PMID=PubMed ID, RR=risk ratio, SD=standard deviation, WC=waist circumference.

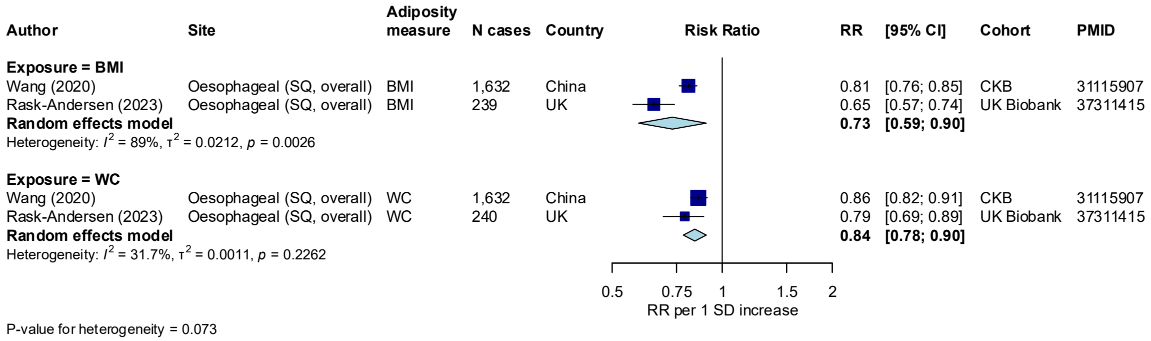

#### Supplementary Figure 79: Associations of BMI, waist circumference and oesophageal SQ cancer risk, per 1 SD increase

Study-specific RRs are represented by squares (with their 95% CIs as lines). RRs were combined using random effects meta-analysis (represented by a diamond). Heterogeneity across studies was assessed using $I^2$ and $tau^2$, within adiposity measure. Heterogeneity in the pooled estimates by adiposity measurement was assessed using the Wald statistic.

Abbreviations: BMI=Body mass index, CI=confidence interval, PMID=PubMed ID, RR=risk ratio, SD=standard deviation, SQ=squamous cell carcinoma, WC=waist circumference.

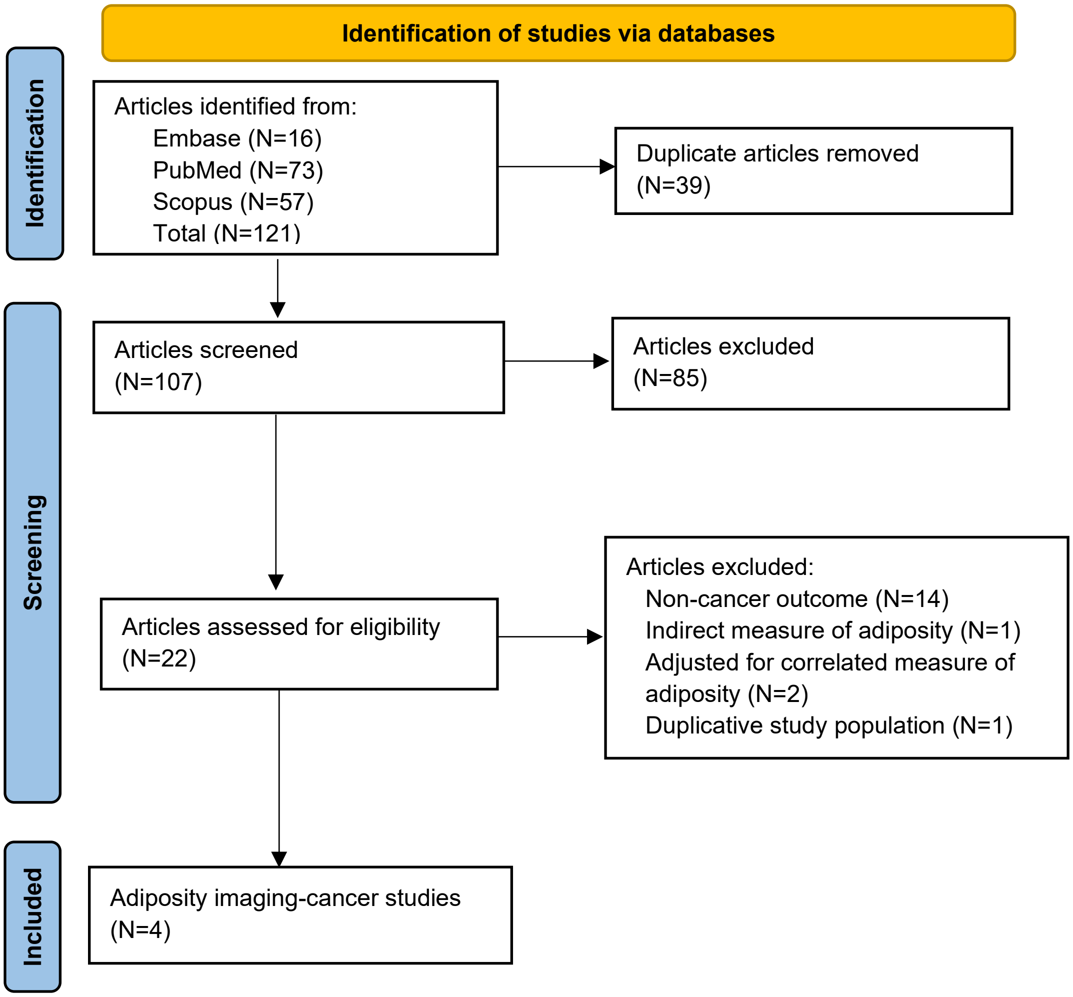

**Supplementary Figure 80: Flow diagram of search strategy and study selection for adiposity imaging studies**
